# Recommendations for surveillance of pulmonary dysfunction among childhood, adolescent, and young adult cancer survivors: a report from the International Late Effects of Childhood Cancer Guideline Harmonization Group

**DOI:** 10.1101/2023.08.28.23294741

**Authors:** Maria Otth, Rahel Kasteler, Renée L. Mulder, Jennifer Agrusa, Saro H. Armenian, Dana Barnea, Anne Bergeron, Neel S. Bhatt, Stephen J. Bourke, Louis S. Constine, Myrofora Goutaki, Daniel M. Green, Ulrike Hennewig, Veronique Houdouin, Melissa M. Hudson, Leontien Kremer, Philipp Latzin, Antony Ng, Kevin C. Oeffinger, Christina Schindera, Roderick Skinner, Grit Sommer, Saumini Srinivasan, Dennis C. Stokes, Birgitta Versluys, Nicolas Waespe, Daniel J. Weiner, Andrew C. Dietz, Claudia E. Kuehni

## Abstract

Childhood, adolescent, and young adult (CAYA) cancer survivors are at risk of pulmonary dysfunction. Current follow-up care guidelines are discordant. Therefore, the International Late Effects of Childhood Cancer Guideline Harmonization Group established and convened a panel of 33 experts to develop evidence-based surveillance guidelines. We critically reviewed available evidence regarding risk factors for pulmonary dysfunction, types of pulmonary function testing, and timings of surveillance, then we formulated our recommendations. We recommend that CAYA cancer survivors and healthcare providers are aware of reduced pulmonary function risks and pay vigilant attention to potential symptoms of pulmonary dysfunction, especially among survivors treated with allogeneic haematopoietic stem cell transplantation, thoracic radiotherapy, and thoracic surgery. Based on existing limited evidence, our panel currently recommends pulmonary function testing only for symptomatic survivors. Since scarce existing evidence informs our recommendation, we highlight the need for prospective collaborative studies to address pulmonary function knowledge gaps among CAYA cancer survivors.

## INTRODUCTION

Children, adolescents, and young adults (CAYA) diagnosed with cancer are at risk for pulmonary dysfunction and death from pulmonary conditions years to decades after completing treatment (1–5). Treatment modalities previously defined as lung-toxic include chemotherapeutic agents, such as busulfan, bleomycin, carmustine, and lomustine, and thoracic radiotherapy, thoracic surgery, and allogeneic haematopoietic stem cell transplantation (HSCT) (6, 7). Pathophysiological mechanisms of pulmonary damage include oxidative stress from lung-toxic chemotherapeutics, free radical formation during radiotherapy, and transplant-specific pulmonary complications, such as idiopathic pneumonia syndrome or bronchiolitis obliterans (7–9). Free radicals injure type II pneumocytes, resulting in decreased proliferative capacity, less surfactant production, and ultimately reduced lung compliance (10). Activations of an inflammatory cascade and changes in endothelial cells of surrounding vasculature result in leakage of proteins and inflammatory cells into alveoli (10). Such inflammation is commonly the final path of different pathophysiological mechanisms which can either resolve or progress to fibrotic changes in alveolar septa, causing restrictive impairments. Surgical removal of parts of the lung or chest wall as part of cancer therapy reduces lung volumes and causes restrictive changes.

Symptomatic pulmonary dysfunction presents with chronic cough or dyspnea, especially on exertion. With large pulmonary functional reserve, dyspnea may not be noticed until a substantial decline in pulmonary function has occurred. Pulmonary function testing (PFT) detects pulmonary dysfunction before symptoms arise. Commonly used PFT include spirometry, body plethysmography, and measurement of diffusion capacity for carbon-monoxide (DLCO). Spirometry and body plethysmography mainly assess changes in larger airways. Examinations detecting changes in lung periphery or inhomogeneous ventilation, such as washout tests, are used to answer research questions, yet remain unintroduced into routine clinical care (11). Among CAYA cancer survivors exposed to lung-toxic treatments, obstructive changes have been reported in up to 4%, restrictive disease 24%, and diffusion capacity impairment 49% (1, 2). Proportions are even higher among certain sub-groups of CAYA cancer survivors, such as following HSCT (12).

Since long-term CAYA cancer survivor numbers constantly increase from diagnostic, risk stratification, and treatment strategy advances, long-term CAYA cancer survivor surveillance is a high priority (13). Awareness of the risk of late effects from cancers or treatments led to the development of different long-term follow-up (LTFU) guidelines, such as those from the Children’s Oncology Group (COG), the Dutch Childhood Oncology Group (DCOG), and from the United Kingdom Children’s Cancer and Leukaemia Group (UKCCLG) (14–16). Although COG, DCOG, and UKCCLG guidelines recommend screening for pulmonary dysfunction, they are discordant regarding indication, timing of initiation, frequency, and screening method. The International Late Effects of Childhood Cancer Guideline Harmonization Group (IGHG) surveillance guidelines develop internationally harmonised and implementable recommendations based on evidence from existing literature and international expert consensus when evidence is unavailable (17). In this current IGHG initiative, we specifically define which CAYA cancer survivors likely benefit from screening for pulmonary dysfunction and when and how screening should be performed. We also further highlight limitations of the current evidence informing surveillance recommendations for pulmonary dysfunction and knowledge gaps to address in future research.

## METHODS

### Methodology of International Late Effects of Childhood Cancer Guideline Harmonization Group (IGHG) and formulating key questions

Information about methods used to formulate IGHG recommendations was published previously (18). For our current recommendation, we organised a guideline panel of 33 members—representing COG, DCOG, UKCCLG, the Pan-European Network for Care of Survivors after Childhood and Adolescent Cancer (PanCare)—and pulmonary health and late effects experts from various medical specialties: paediatric oncology and haematology; paediatric and adult pulmonology; radiation oncology; epidemiology; and guideline experts (Appendix A) (19–21).

IGHG’s approach to formulating recommendations involves answering five key questions: 1) "Who needs surveillance?" 2) "What surveillance modality should be used?" 3) "At what frequency should surveillance be performed?" 4) “When should surveillance be initiated?” and 5) "What should be done when abnormalities are found?" Based on our preliminary literature search and a resulting absence of data, we performed the systematic literature search for only question 1) “Who needs surveillance?” and used expert opinions from paediatric and adult pulmonologists for questions 2–5. We did not use guidelines for surveillance of other pulmonary diseases, such as idiopathic interstitial pneumonitis or chronic obstructive pulmonary disease, due to different underlying pathophysiological mechanisms. Notably, guidelines for other diseases start with symptomatic patients diagnosed with specific pulmonary diseases, which differs from our population of interest—asymptomatic CAYA cancer survivors at risk of developing pulmonary problems.

### Comparing existing guidelines and formulating clinical questions

First, we separately compared COG, DCOG, Scottish Intercollegiate Guidelines Network, and UKCCLG recommendations for each of the five key questions (15, 16, 19, 22). For the key question, “What should be done when abnormalities are found?” we also included vaccination and lifestyle factors. For the key question “Who needs surveillance?” we subsequently formulated 11 clinical questions and sub-questions to strengthen concordant recommendations and find consensus for discordant recommendations (Appendix B).

We used the PICO-framework to formulate clinical questions (23). Our *population* of interest included CAYA cancer survivors—defined by at least 50% of survivors diagnosed before age 30—who completed cancer treatment at least two years previously. We included potentially lung-toxic treatment modalities (selected chemotherapeutic agents, thoracic radiotherapy, thoracic surgery, and allogeneic HSCT) and tobacco exposure as *interventions*. We also included all chemotherapeutic agents mentioned in current LTFU guidelines as risk factors for pulmonary dysfunction (bleomycin, busulfan, and nitrosoureas [lomustine and carmustine]). Based on expert opinion, we additionally included treatment with cyclophosphamide, gemcitabine, and methotrexate. *Comparators* were considered during data extraction and differed between studies according to study design, such as non-exposed survivors, survivors exposed to lower chemotherapeutic doses, or community controls. Our *outcome* of interest was pulmonary dysfunction assessed by PFT. We focused on this single outcome because our preliminary literature search showed that other commonly reported clinical endpoints—in particular survivor-reported symptoms or clinician-reported diagnoses—had been assessed and reported heterogeneously and had the risk of subjectivity, such as different definitions of chronic cough or dyspnea (24–28). We categorised pulmonary dysfunction into four groups: obstruction (by FEV1, FEV1/FVC, MEF25–75%); restriction (by TLC, FVC); hyperinflation (by RV, RV/TLC); and diffusion capacity impairment (by DLCO). Although clinically relevant hyperinflation should be interpreted together with obstruction, we defined hyperinflation as a separate pulmonary outcome since it was defined as such in included studies.

### Systematic literature search on "Who needs surveillance?"

We conducted our first systematic literature search in accordance with the preferred reporting items for systematic reviews and meta-analyses (PRISMA) guidelines restricted to PubMed and Ovid in November 2016 with updates in June 2019; December 2020; June 2022; and April 2023 (29). We developed our search strategy based on 11 clinical questions and five concepts: cancer diagnosis, population of CAYA cancer survivors, potential lung-toxic treatment modalities, pulmonary outcomes, and late effects (Appendix C). The inclusion criteria were given through the PICO-framework. We also excluded studies with fewer than 20 participants and studies only assessing prevalence of pulmonary dysfunction without measuring effect sizes of associations between exposures and pulmonary dysfunction (Appendix D).

Guideline panel members screened titles, abstracts, and full-texts. Two authors independently screened each study. Coordinators (MO, RK) resolved discrepancies. We extracted data from each study and entered it into evidence summary tables (Appendix E); separately performed risk of bias assessments for each study (Appendix F); and completed overall quality assessments of available evidence for each clinical question, according to the GRADE criteria (Appendix G) (18, 30). Each eligible study could contribute to answering more than one clinical question. For our overall conclusion of evidence, we summarised findings by type of pulmonary dysfunction (Appendix H).

### Expert consensus on surveillance modality, start and frequency of screening, and procedures in case of pulmonary dysfunction

For questions about surveillance modality, start and frequency of screening, and procedures in cases of pulmonary dysfunction (key questions 2ℒ5), we held numerous meetings with paediatric and adult pulmonologists and guideline development experts. We formulated suggestions based on initial meetings, which we then discussed with guideline panel members until reaching consensus through an iterative approach with successive revisions and implementing suggestions from all panel members.

### Translating evidence into recommendations

We assessed evidence and information we gathered from our systematic literature search and expert consensus within the evidence-to-decision framework—it weighs the impact of screening by estimating benefits and harms, resources and costs, impact on health inequities, acceptability, and feasibility. Through panel discussions, we achieved consensus for final recommendations, which we subsequently discussed with additional experts, including oncologists and survivors (Appendix A).

## RESULTS

### Comparison with existing guidelines

Comparing existing guidelines revealed relevant discordance (Appendix I). Since the Scottish guideline omitted recommendations for pulmonary dysfunction, we excluded it (22). The remaining three guidelines considered CAYA cancer survivors at risk for pulmonary dysfunction after exposure to bleomycin, busulfan, nitrosoureas, thoracic radiotherapy, or thoracic surgery. Practical details revealed discordances, including cut-off doses for chemotherapeutic agents or thoracic radiotherapy; radiation volume; age at treatment; and additional risk factors, such as renal dysfunction and pulmonary infection. All three guidelines recommended PFT, yet did not specify tests. We found no concordance for screening frequency and starting screening with five and ten years after diagnosis in the Dutch guideline; two years after completion of treatment in the COG guideline; and end of treatment in the UK guideline. Guidelines agreed about referring CAYA cancer survivors to pulmonologists in cases of pulmonary dysfunction; alerting anaesthetists about previous bleomycin treatment; advising survivors not to smoke; and considering pneumococcal and influenza immunisation (Appendix I).

### General results from systematic literature search

Our systematic literature search identified 9284 studies. We assessed 704 full-texts for eligibility; 26 studies fulfilled inclusion criteria (Figure 1, Table 1) (1–3, 31–53). Reasons for excluding full-texts mainly included 1) assessing outcomes other than pulmonary function by PFT (n=186); 2) including non-CAYA cancer survivors (n=164); and 3) assessing outcomes fewer than two years after completing treatment (n=91). Most studies (n=12) included CAYA survivors of different cancer types; followed by studies on leukaemia (n=7), lymphoma (n=4), and brain tumors, neuroblastoma, and osteosarcoma with one study each. Appendix J contains key characteristics and our summary of evidence for each included study; Appendix K presents our evidence assessment summary and quality of data contributing to recommendations per clinical question. Quality of evidence was very low or low for most clinical questions (Table 2). We summarise primary reasons for downgrading the quality of evidence in Table 3 and provide more detail in Appendix K.

**Figure 1:**
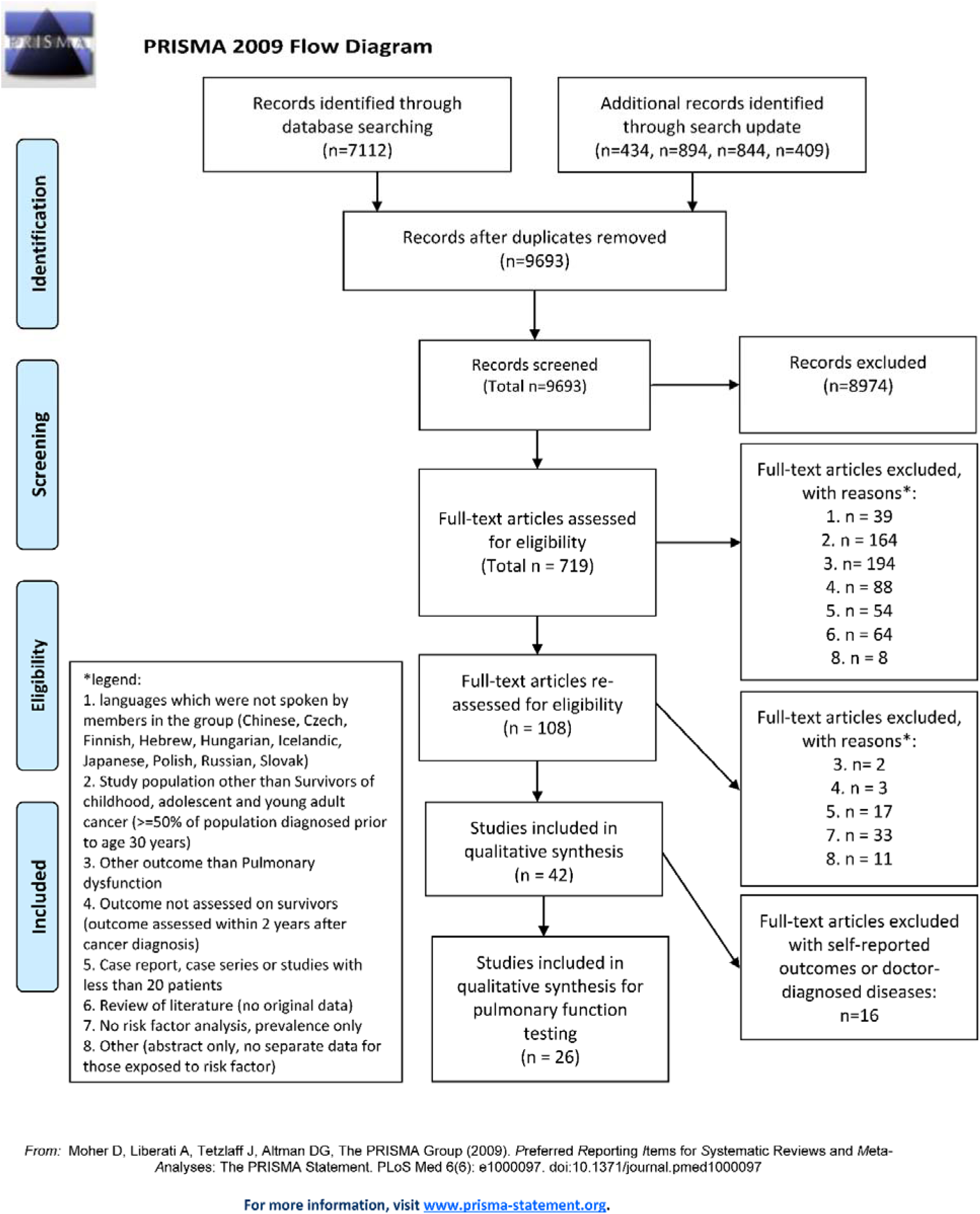
Flow diagram for selection of studies. (Studies could be included in more than one category)

**Table 1:**
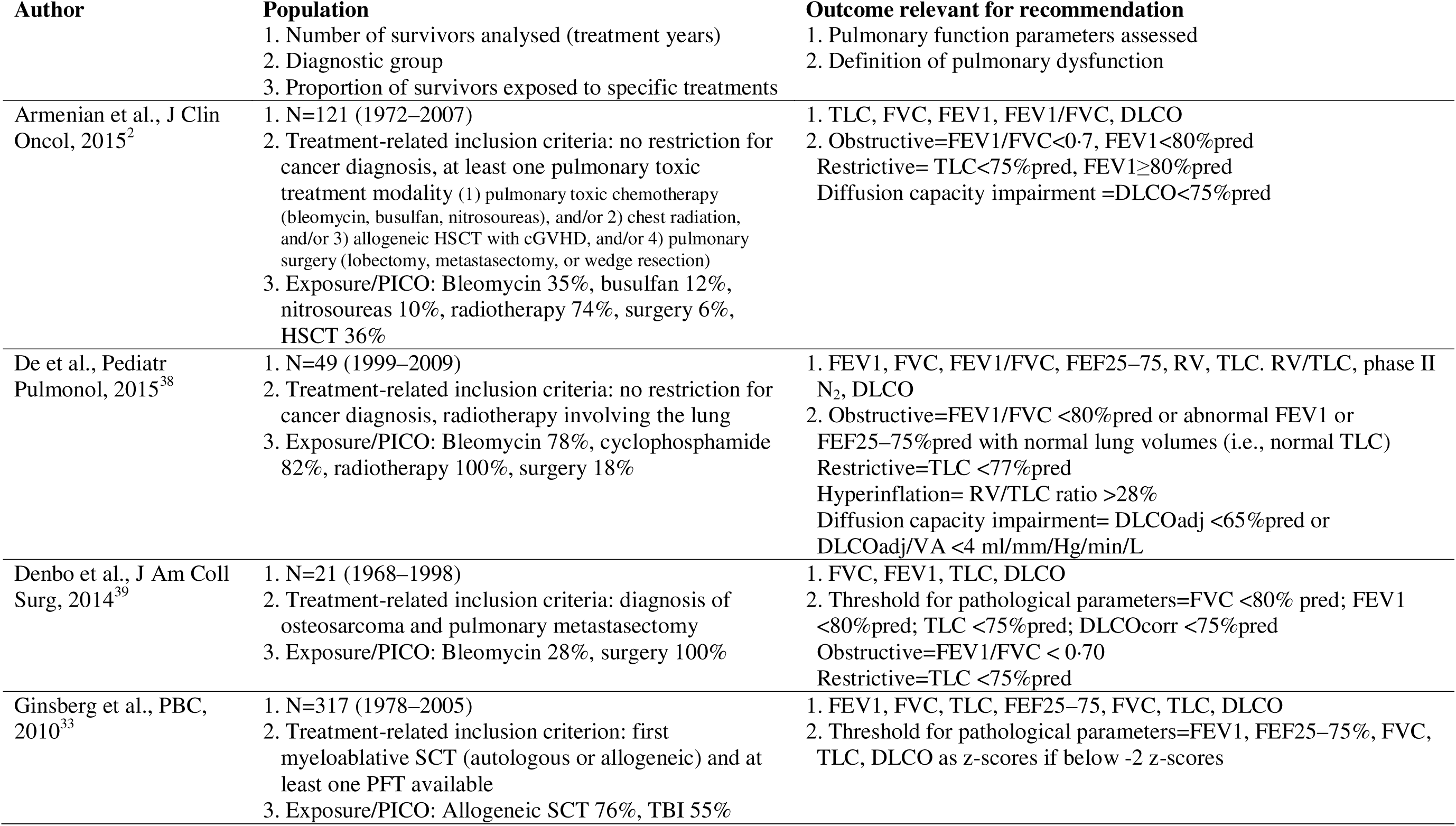
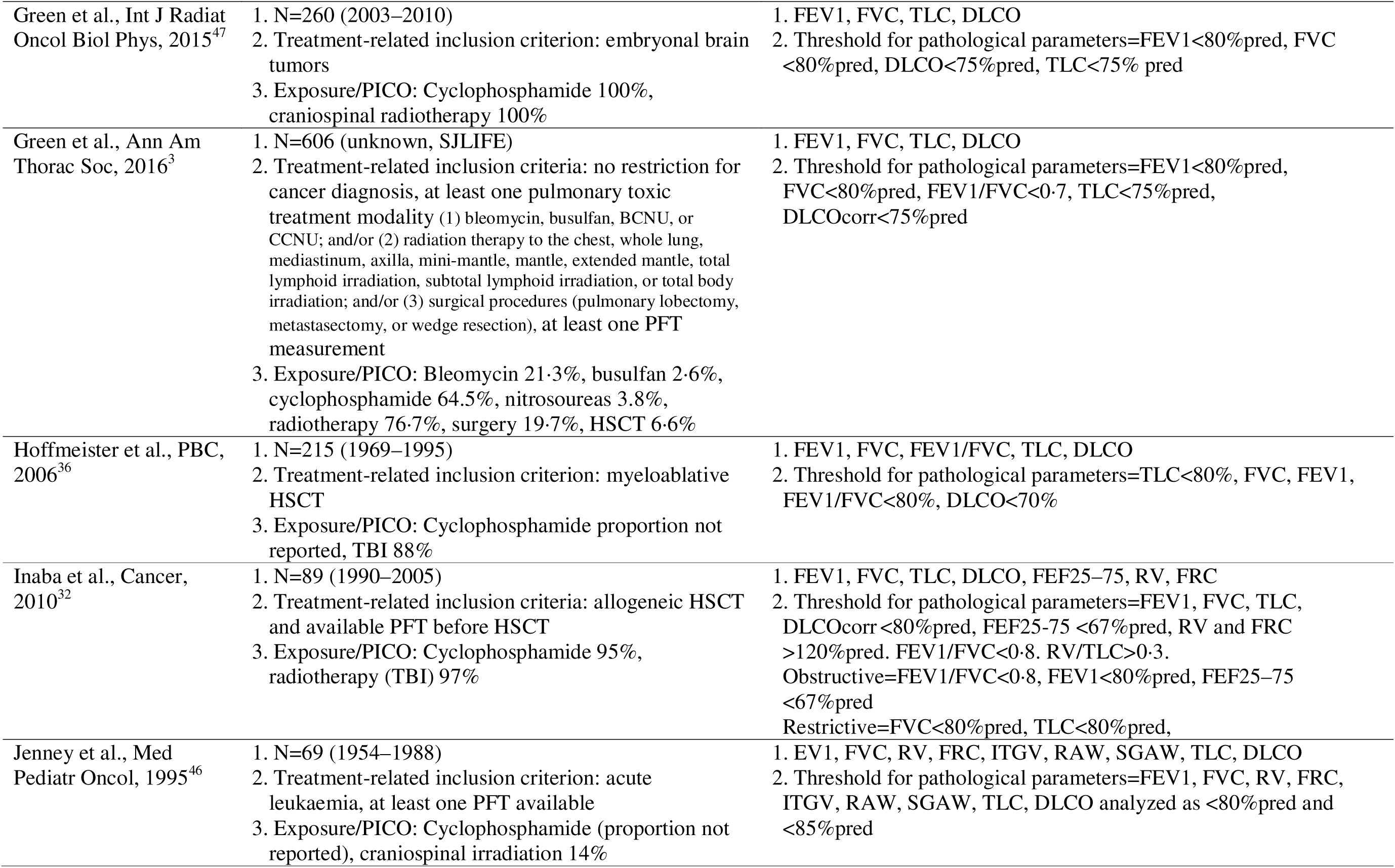
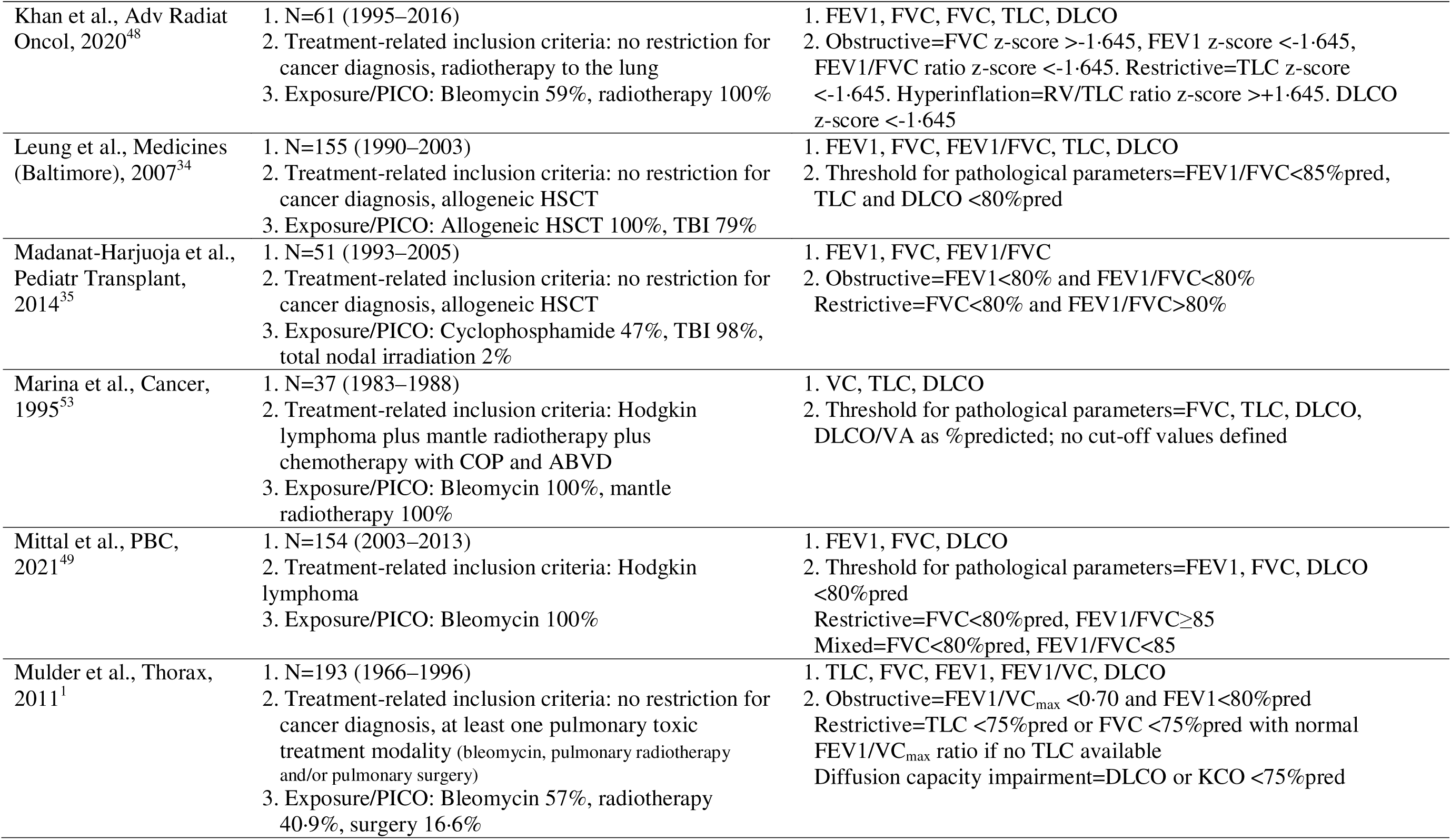
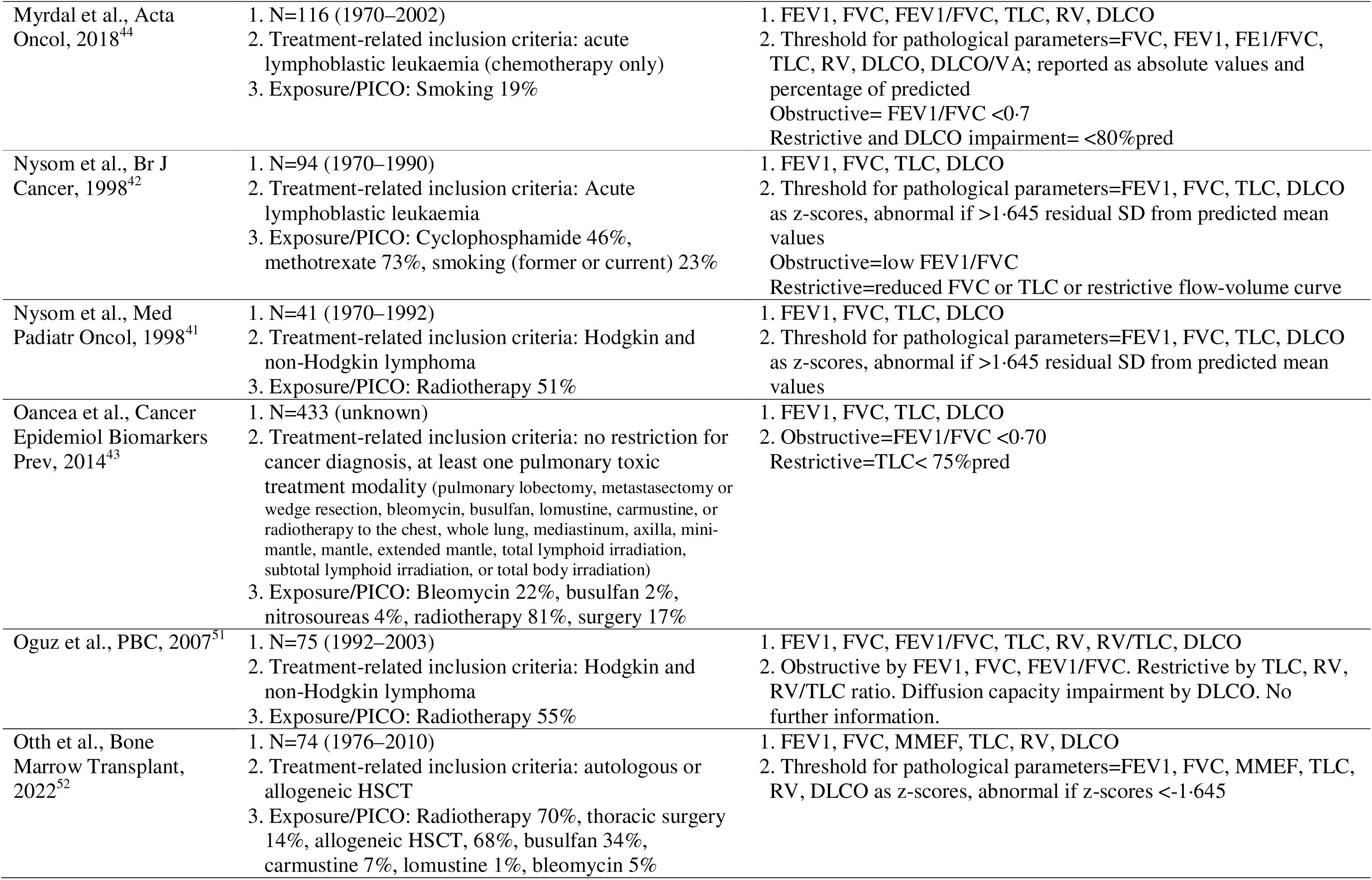
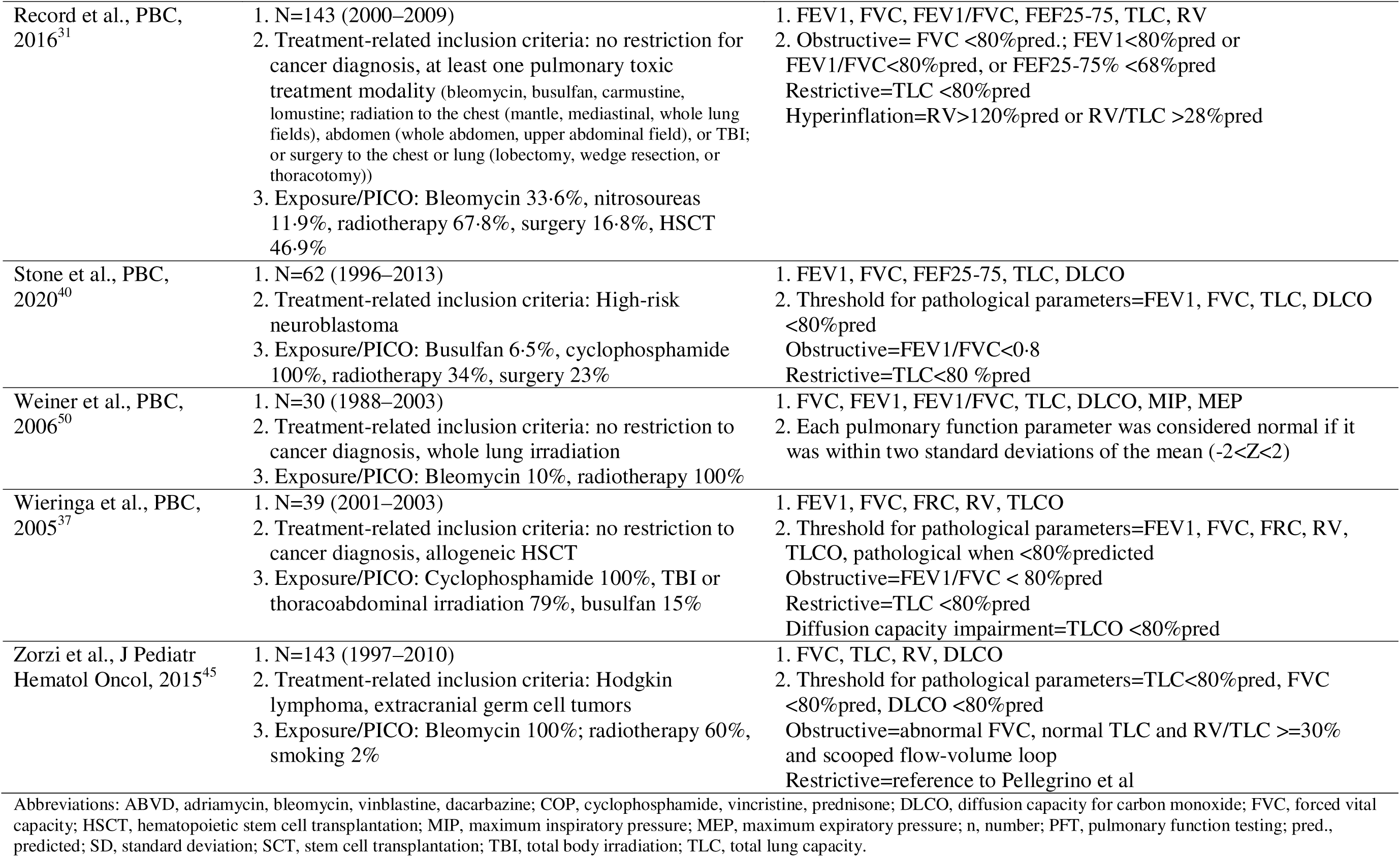
Key characteristics of included studies used for recommendation (n=26). Detailed information available in Appendix K.

**Table 2:**
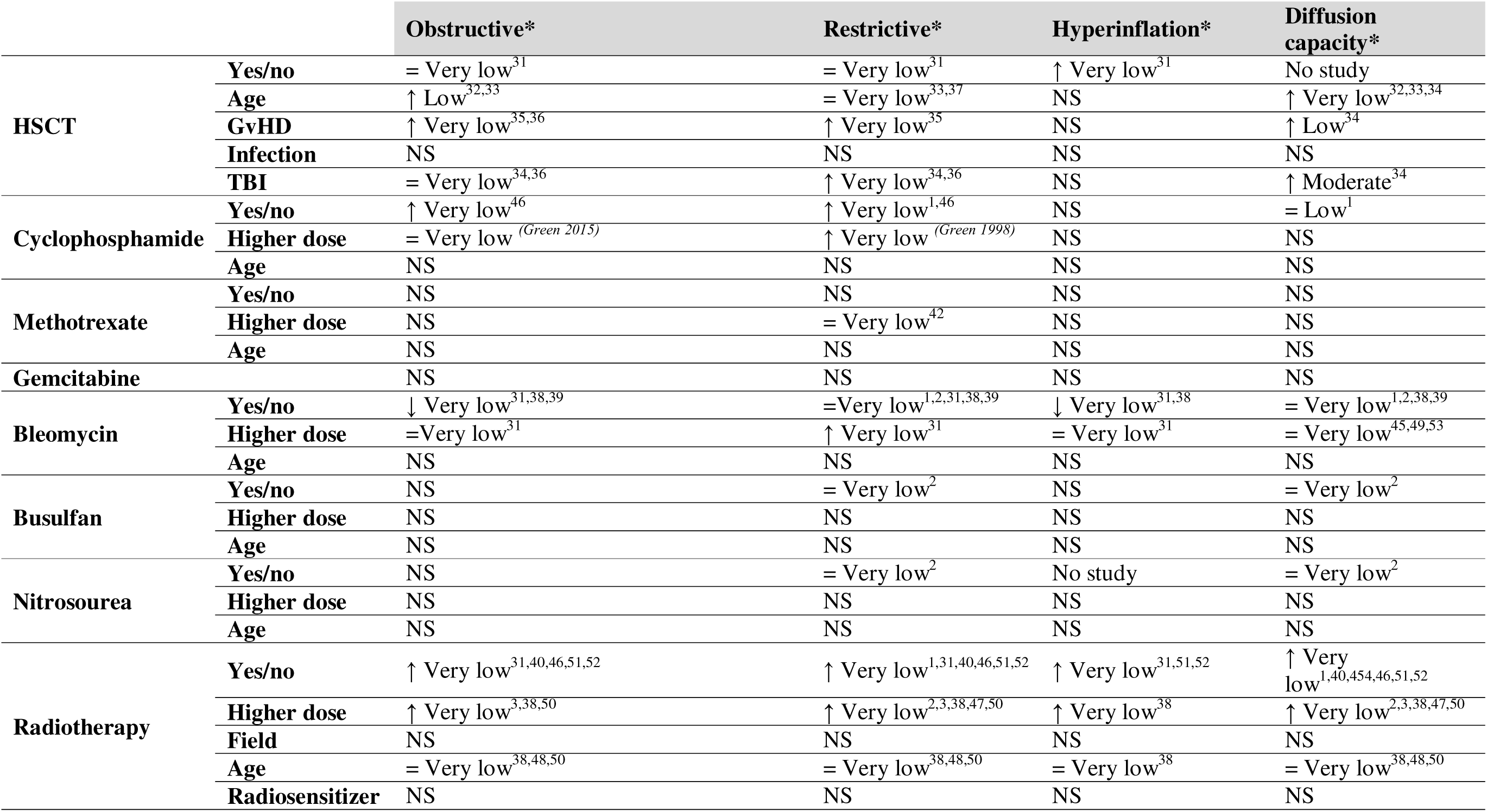
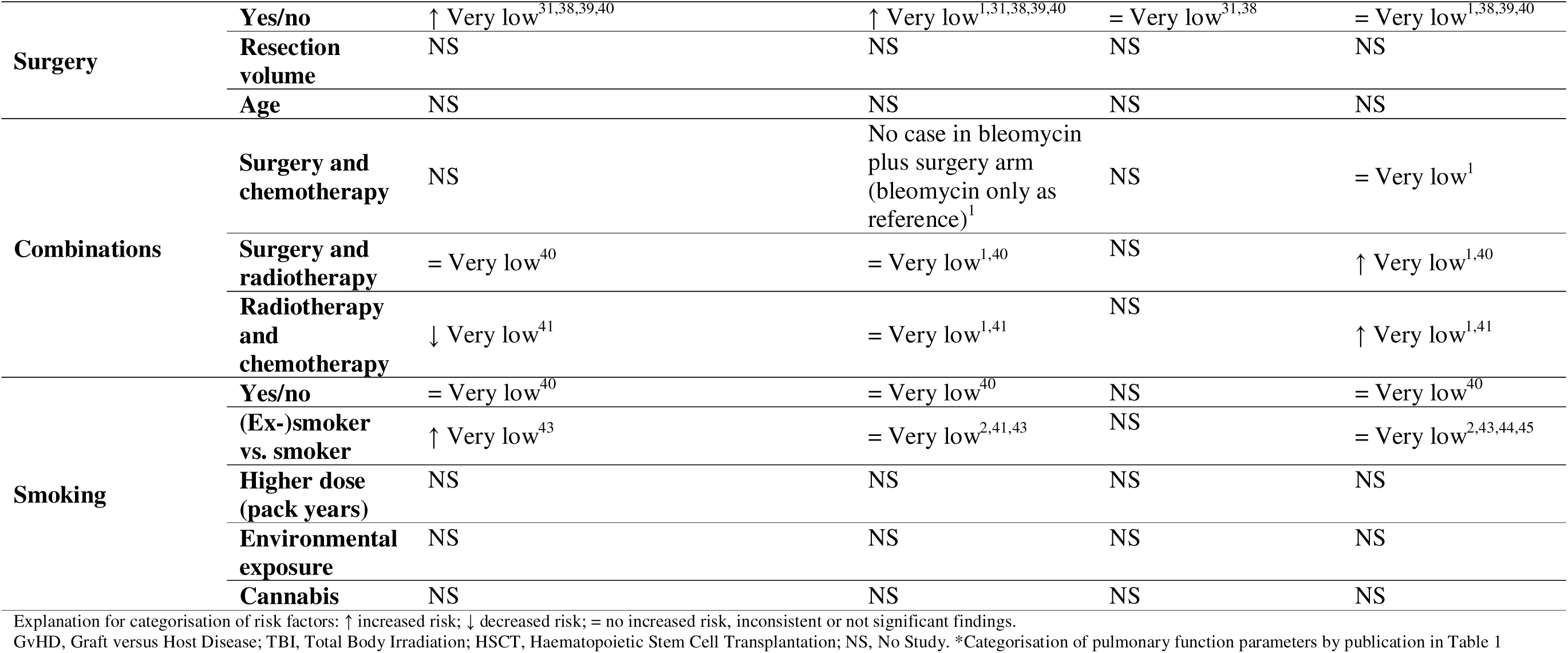
Conclusions and quality of the evidence for the risk and risk factors for pulmonary function impairment among childhood, adolescent, and young adult cancer survivors diagnosed up to age 30.

|  |  | **Obstructive\*** |  | **Restrictive\*** | **Hyperinflation\*** | **Diffusion capacity\*** |
| --- | --- | --- | --- | --- | --- | --- |
| **HSCT** | **Yes/no** | = Very low<sup>31</sup> |  | = Very low<sup>31</sup> | ↑ Very low<sup>31</sup> | No study |
|  | **Age** | ↑ Low<sup>32,33</sup> |  | = Very low<sup>33,37</sup> | NS | ↑ Very low<sup>32,33,34</sup> |
|  | **GvHD** | ↑ Very low<sup>35,36</sup> |  | ↑ Very low<sup>35</sup> | NS | ↑ Low<sup>34</sup> |
|  | **Infection** | NS |  | NS | NS | NS |
|  | **TBI** | = Very low<sup>34,36</sup> |  | ↑ Very low<sup>34,36</sup> | NS | ↑ Moderate<sup>34</sup> |
| **Cyclophosphamide** | **Yes/no** | ↑ Very low<sup>46</sup> |  | ↑ Very low<sup>1,46</sup> | NS | = Low<sup>1</sup> |
|  | **Higher dose** | = Very low<sup>(Green 2015)</sup> |  | ↑ Very low<sup>(Green 1998)</sup> | NS | NS |
|  | **Age** | NS |  | NS | NS | NS |
| **Methotrexate** | **Yes/no** | NS |  | NS | NS | NS |
|  | **Higher dose** | NS |  | = Very low<sup>42</sup> | NS | NS |
|  | **Age** | NS |  | NS | NS | NS |
| **Gemcitabine** |  | NS |  | NS | NS | NS |
| **Bleomycin** | **Yes/no** | ↓ Very low<sup>31,38,39</sup> |  | = Very low<sup>1,2,31,38,39</sup> | ↓ Very low<sup>31,38</sup> | = Very low<sup>1,2,38,39</sup> |
|  | **Higher dose** | = Very low<sup>31</sup> |  | ↑ Very low<sup>31</sup> | = Very low<sup>31</sup> | = Very low<sup>45,49,53</sup> |
|  | **Age** | NS |  | NS | NS | NS |
| **Busulfan** | **Yes/no** | NS |  | = Very low<sup>2</sup> | NS | = Very low<sup>2</sup> |
|  | **Higher dose** | NS |  | NS | NS | NS |
|  | **Age** | NS |  | NS | NS | NS |
| **Nitrosourea** | **Yes/no** | NS |  | = Very low<sup>2</sup> | No study | = Very low<sup>2</sup> |
|  | **Higher dose** | NS |  | NS | NS | NS |
|  | **Age** | NS |  | NS | NS | NS |
| **Radiotherapy** | **Yes/no** | ↑ Very low<sup>31,40,46,51,52</sup> |  | ↑ Very low<sup>1,31,40,46,51,52</sup> | ↑ Very low<sup>31,51,52</sup> | ↑ Very low<sup>1,40,454,46,51,52</sup> |
|  | **Higher dose** | ↑ Very low<sup>3,38,50</sup> |  | ↑ Very low<sup>2,3,38,47,50</sup> | ↑ Very low<sup>38</sup> | ↑ Very low<sup>2,3,38,47,50</sup> |
|  | **Field** | NS |  | NS | NS | NS |
|  | **Age** | = Very low<sup>38,48,50</sup> |  | = Very low<sup>38,48,50</sup> | = Very low<sup>38</sup> | = Very low<sup>38,48,50</sup> |
|  | **Radiosensitizer** | NS |  | NS | NS | NS |

|  |  |  |  |  |  |
| --- | --- | --- | --- | --- | --- |
| **Surgery** | **Yes/no** | ↑ Very low<sup>31,38,39,40</sup> | ↑ Very low<sup>1,31,38,39,40</sup> | = Very low<sup>31,38</sup> | = Very low<sup>1,38,39,40</sup> |
|  | **Resection volume** | NS | NS | NS | NS |
|  | **Age** | NS | NS | NS | NS |
| **Combinations** | **Surgery and chemotherapy** | NS | No case in bleomycin plus surgery arm (bleomycin only as reference)<sup>1</sup> | NS | = Very low<sup>1</sup> |
|  | **Surgery and radiotherapy** | = Very low<sup>40</sup> | = Very low<sup>1,40</sup> | NS | ↑ Very low<sup>1,40</sup> |
|  | **Radiotherapy and chemotherapy** | ↓ Very low<sup>41</sup> | = Very low<sup>1,41</sup> | NS | ↑ Very low<sup>1,41</sup> |
| **Smoking** | **Yes/no** | = Very low<sup>40</sup> | = Very low<sup>40</sup> | NS | = Very low<sup>40</sup> |
|  | **(Ex-)smoker vs. smoker** | ↑ Very low<sup>43</sup> | = Very low<sup>2,41,43</sup> | NS | = Very low<sup>2,43,44,45</sup> |
|  | **Higher dose (pack years)** | NS | NS | NS | NS |
|  | **Environmental exposure** | NS | NS | NS | NS |
|  | **Cannabis** | NS | NS | NS | NS |

Explanation for categorisation of risk factors: ↑ increased risk; ↓ decreased risk; = no increased risk, inconsistent or not significant findings.

**Table 3:** Main reasons for downgrading the quality of evidence.

|  |
| --- |
| **Representativeness uncertain:**<br>- 13 studies (50%) did not report original cohort sizes from where populations at-risk were selected (selection and attrition biases). |
| **Imprecision and indirectness in performing PFT:**<br>- Only 15 of 27 included studies (56%) used ERS/ATS recommendations on how to perform PFT. |
| **Imprecision and indirectness in reporting PFT results:**<br>- Only 20 of 27 studies (74%) reported the source of reference values used to standardise the survivor results. 22 different sources of reference values were used overall, with up to 10 different sources of reference values in the same study.<br>- Only 2 of 27 studies (7%) used the reference values from the global lung initiative, which is established as the international standard. |
| **Inconsistent reporting of PFT results:**<br>- 20 of 27 studies reported percentage of predicted values<br>- 5 of 27 studies reported z-scores<br>- 1 of 27 studies reported percentage of predicted values and z-scores<br>- 1 of 27 studies reported percentage of predicted and absolute values |
| **Inconsistent definitions of cut-off values** used to define normal values, obstruction, restriction, or diffusion capacity impairment |

Abbreviations: ATS, American Thoracic Society; ERS, European Respiratory Society; PFS, pulmonary function testing

### Evidence on risk factors for pulmonary dysfunction among CAYA cancer survivors

We identified seven studies examining allogeneic HSCT as a risk factor for pulmonary dysfunction (31–37); 13 studies for thoracic radiotherapy (1–3, 31, 38, 40, 45–48, 50–52); five studies for thoracic surgery (1, 31, 38–40); between one and eight studies for selected chemotherapeutic agents (bleomycin, busulfan, nitrosoureas, cyclophosphamide, methotrexate); no studies for gemcitabine; and six studies for tobacco exposure (2, 40, 41, 43–45) (Table 2, Appendix K). For busulfan and nitrosoureas, only one study of very low quality was available; it provided insufficient evidence to decide whether these agents significantly impact pulmonary function (2). Four studies of low to very low quality of evidence assessed pulmonary function after cyclophosphamide-containing treatment (1, 42, 46, 47). Studies examining effects from active tobacco smoking showed contradictory results (2, 41, 43–45). No studies examined impact from passive tobacco smoking or cannabis use. Our clinical questions and sub-questions aimed to investigate impact from exposure versus non-exposure and also impact from different dose levels; age at exposure; chronic graft versus host disease; infections; and total body irradiation among individuals treated with HSCT (Appendix B). Between one and four studies examined impact from different cyclophosphamide, methotrexate, and bleomycin doses; yet no studies were available for different doses of nitrosoureas and busulfan. Seven studies examined the impact of age at treatment with HSCT and radiotherapy.

Overall, we identified several potential sources of bias and methodological issues in most studies (Table 3). Study design was retrospective in more than half (n=14), increasing risks of bias and non-standardised measurements. Half of the studies did not describe original population sizes from where they selected participants at risk. This makes it uncertain if results are internally and externally representative and can be extrapolated for the entire population of CAYA cancer survivors. Only half of the studies described how they performed pulmonary function testing, such as by implementing the joint European Respiratory Society (ERS)-American Thoracic Society (ATS) recommendations. Even though most studies (74%) reported reference values used to standardize CAYA cancer survivor PFT results, studies used 22 different sources of reference values—with up to ten different sources in one study (47). Only two studies used internationally recommended all-age reference values from the Global Lung Initiative (GLI) (3, 52). Cut-off value definitions, such as for restrictive disease, were inconsistent, which made PFT results difficult to interpret and impossible to compare between studies and age groups. Additionally, findings from different studies on the same exposure were frequently inconsistent or contradictory, such as with bleomycin (31, 38, 39). Inconsistent and contradictory aspects precluded analysing quantitatively and interpreting findings, such as a meta-analysis; prevented formulation of recommendations for specific pulmonary function abnormalities, such as obstructive, restrictive, and diffusion impairment; confounded analysis of effects of specific chemotherapeutic agents on pulmonary function; or prevented definition of cut-off doses for chemotherapeutics or radiotherapy. However, studies we examined provided some evidence CAYA cancer survivors treated with allogeneic HSCT, thoracic radiotherapy, and thoracic surgery are at risk for pulmonary dysfunction as measured by PFT.

### Translating evidence and expert consensus into recommendations

#### Asymptomatic CAYA cancer survivors

Our panel concluded evidence was insufficient for recommending routine PFT for asymptomatic CAYA cancer survivors at present (Appendix L, Table 4). The current evidence is low quality with risks of participation bias and cannot be translated to represent the wider general situation in asymptomatic CAYA cancer survivors. Such factors are essential for formulating clear recommendations. For exposures with evidence of impacting pulmonary function, there is currently no effective intervention that reverses or delays pulmonary disease progression among asymptomatic survivors. Therefore, risk-benefit-assessments do not favour screening now. However, since extensive evidence shows smoking presents harmful effects on pulmonary health among the general population, our panel agreed about counselling CAYA cancer survivors not to smoke. The panel further recommended vaccinations for CAYA cancer survivors at risk for pulmonary-related pathogens, as appropriate for other vulnerable populations (Table 4). Influenza vaccination is recommended based on concordance between existing guidelines. Based on local or national recommendations for populations with increased vulnerability of pulmonary disease, such as pneumococcus and SARS-CoV-2, the panel recommended considering additional vaccinations against bacteria or viruses. All three LTFU care guidelines mentioned informing anaesthetists about previous bleomycin treatment, yet without further information or support from included studies; our additional search showed contradictory findings (54, 55).

**Table 4:** Harmonised recommendations for surveillance of pulmonary dysfunction for childhood, adolescent, and young adult cancer survivors.

|  |
| --- |
| **General recommendations** |
| CAYA cancer survivors and their healthcare providers should be aware of the risk of reduced pulmonary function, and pay attention to symptoms (shortness of breath on exertion, chronic cough) after treatment with:<br>- Allogeneic haematopoietic stem cell transplantation (very low quality of evidence)<br>- Radiotherapy to fields exposing lung tissue, including TBI (very low to moderate quality of evidence)<br>- Surgery to the lung or chest wall (very low quality of evidence)<br>(strong recommendation) |
| In at-risk* CAYA cancer survivors it is recommended to:<br>- Get a yearly influenza vaccination and additional vaccinations based on local or national recommendations<br>- Consider vaccination against viral pathogens that cause pneumonias according to local or national guidelines<br>For all CAYA cancer survivors it is recommended to:<br>- Avoid tobacco exposure, quit smoking, and/or reduce exposure to environmental smoke<br>(strong recommendation, expert opinion) |
| **Who needs surveillance for pulmonary dysfunction and what surveillance modality should be used?** |
| Routine pulmonary function testing is not recommended for <u>asymptomatic</u> at-risk* CAYA cancer survivors, due to lack of interventions to prevent the deterioration of asymptomatic pulmonary dysfunction (strong recommendation, lack of evidence, expert opinion) |

\*Survivors treated with allogeneic haematopoietic stem cell transplantation (very low quality of evidence); radiotherapy to fields exposing lung tissue, including TBI (very low to moderate quality of evidence); and surgery to the lung or chest wall (very low quality of evidence)
Abbreviations: CAYA, childhood adolescent and young adult; TBI, total body irradiation

#### Symptomatic CAYA cancer survivors

For symptomatic CAYA cancer survivors—especially among those treated with allogeneic HSCT, thoracic radiotherapy, and thoracic surgery—our panel agreed upon readily performing PFT with results evaluated by pulmonologists experienced with CAYA cancer populations. Health care professionals and CAYA cancer survivors should heed symptoms, such as chronic cough, chest tightness, dyspnoea, wheezing, or exercise intolerance. Consideration of differential diagnoses, such as cardiac dysfunction, should guide selection of appropriate investigations. Pulmonologists from our guideline panel recommended spirometry, body plethysmography, and DLCO measurements wherever possible. Breath washout tests can additionally be performed if available. Fractional exhaled nitric oxide, bronchodilator reversibility tests, or other specific investigations should be used for differential diagnoses of other pulmonary dysfunction causes, which—even among vulnerable populations—are arguably more frequent reasons for pathological findings than previous cancer treatment, such as asthma in cases of obstructive disease. Managing cases of abnormal findings and frequency of further PFT depends on local institutions and guidance from local pulmonologists; it is not part of our recommendations.

## DISCUSSION

Our review summarises existing guidelines, evidence from systematic literature searches, and harmonised recommendations for pulmonary dysfunction screening among CAYA cancer survivors diagnosed before age 30 with exposure to potentially lung-toxic cancer treatment modalities. Because current evidence is scarce with quality limitations and because there are no effective treatments for asymptomatic pulmonary dysfunction, our panel limited recommendations for PFT to symptomatic CAYA cancer survivors only. We recommend health care providers to be aware of increased risks for possible pulmonary dysfunction—especially among survivors treated with allogeneic HSCT, thoracic radiotherapy, and thoracic surgery; be vigilant for early clinical symptoms of pulmonary dysfunction; and refer symptomatic CAYA cancer survivors to pulmonologists experienced with the population. We also recommend counselling all CAYA cancer survivors about lifestyle factors relevant for pulmonary and general health.

Our recommendations are supported by two additional studies specific for children following HSCT (56, 57). Both studies were not considered in our final recommendation as they formulated follow-up recommendations independent of underlying diagnosis, including HSCT for malignant diseases, immune deficiencies, inherited bone marrow failure syndromes, and haemoglobinopathies. However, after two years of follow-up, neither study recommended regular PFT for asymptomatic children and adolescents; rather they advised considering follow-up PFT based on symptoms and past measurements (56, 57).

### Gaps in knowledge and future directions for research

With currently available evidence, we only answered a few of our original clinical questions and often to only a limited extent, such as any exposure to radiotherapy without differentiation for doses or volumes. To improve evidence on pulmonary dysfunction among CAYA cancer survivors, we outlined gaps in knowledge and methodological approaches for future research (Table 5).

**Table 5:** Gaps in knowledge and future directions for research.

| Current knowledge gaps |
| --- |
| **Risk factors for symptomatic and asymptomatic pulmonary dysfunction among CAYA cancer survivors**, including therapeutic exposures, medical conditions, and environmental exposures.<br>• Treatment with<br>• chemotherapeutic agents with reported pulmonary toxicity according to current LTFU guidelines, such as bleomycin, busulfan, carmustine, lomustine<br>• chemotherapeutic agents without reported pulmonary toxicity according to current LTFU guidelines and without clear evidence, such as cyclophosphamide, methotrexate<br>• targeted agents, such as tyrosine kinase inhibitors, checkpoint inhibitors, and monoclonal antibodies<br>• immunotherapy, such as chimeric antigen receptor T-cell therapy<br>• thoracic radiotherapy, such as proton versus photon therapy, and increasing radiation doses or volumes<br>• thoracic surgery, such as thoracotomy and pneumonectomy<br>• haematopoietic stem cell transplantation<br>• combination of the above-mentioned treatment modalities<br>• combination of bleomycin with additional oxygen (during anaesthesia)<br>• Impact of time from exposure on pulmonary function<br>• Impact of age at exposure on the risk of developing pulmonary dysfunction<br>• Impact of attained age at screening on outcome measures, such as PFT results, and clinical symptoms<br>• Impact of existing co-morbidities on pulmonary function, such as cardiac disease, impaired immune function, and neurological deficits<br>• Impact of genetic variants on pulmonary toxicity of cancer treatments<br>• Impact of acute treatment-related toxicities on pulmonary function, such as pulmonary infections, pulmonary GvHD<br>• Impact of inhaled substances, such as vaping, medicinal cannabis, alone or in combination with smoking cigarettes (during therapy and post-therapy)<br>**Detection of pulmonary dysfunction among CAYA cancer survivors**<br>• Benefit of novel PFT, such as multiple-breath washout tests, exhaled nitric oxide, impulse oscillometry, and lung imaging (MRI), including acceptability, sensitivity, specificity, practical consequences, and costs<br>• Longitudinal course of pulmonary dysfunction after cancer treatments, including onset and progression, to determine intervals for PFT<br>• Effects of the cancer itself and cancer treatments on physiological processes, including lung growth, peak attained lung function, and functional decline with ageing<br>• Association of functional outcomes (from PFT detected by screening) with clinical symptoms (onset, type, and severity)<br>• Predictive value of serial PFT to identify individuals who will develop pulmonary dysfunction in the future and who will become symptomatic<br>• Predictive value of serial lung function tests with relation to termination of screening (normal or stable results)<br>• Cost-effectiveness of different screening frequencies and modalities<br>• Potential harms associated with excessive screening and false-positive findings |
| **Interventions to prevent, reverse, or slow the decline in pulmonary function**<br>• Effect of variation in cancer treatments, such as radiotherapy, chemotherapy, or surgery on pulmonary dysfunction<br>• Effect of lifestyle and other preventive strategies on development or worsening of pulmonary dysfunction, such as lifestyle counselling and physical activity<br>• Effect of interventions, such as medical treatments and physiotherapy, to improve or reverse pulmonary dysfunction<br>• Benefit of optimal management of co-morbidities, such as cardiac dysfunction and chronic immunosuppression, on pulmonary dysfunction and pulmonary symptoms<br>• Effect of therapeutic targets used for other pulmonary diseases, such as antifibrotic drugs |
| **Factors to be considered in future studies assessing the risk of pulmonary dysfunction among CAYA cancer survivors**<br>• Close collaboration between oncologists and pulmonologists; paediatric and adult experts.<br>**Study design**<br>• Need for sufficient numbers of CAYA cancer survivors undergoing PFT to maximise statistical power and allow stratifying analyses into sub-groups<br>• Collaboration between different study groups working with harmonised protocols<br>• Avoidance of selection or attrition bias among study cohorts, by testing independent of pulmonary symptoms; access to testing independent of socioeconomic factors etc<br>• Longitudinal investigations including baseline before starting treatment and serial assessment of pulmonary function and correlation with symptoms and other patient-reported outcomes<br>**Standardisation of PFT**<br>• Performance of PFT by trained personnel in centres with expertise / accredited centres<br>• Performance of PFT according to standardised protocols, such as ERS/ATS guidelines<br>• Interpretation of PFT results according to standardised protocols for quality control, such as ERS/ATS guidelines, and standardised reference values, stratified by age and sex, such as GLI<br>• Reporting PFT results as raw data and z-scores instead of binary cut-offs, such as normal or abnormal, restrictive, obstructive, or diffusion capacity impairment<br>• Measurement of different lung function parameters to better describe the nature of lung function impairment<br>• Using novel PFT to facilitate detection of pulmonary dysfunction |

Abbreviations: ATS, American Thoracic Society; CAYA, childhood adolescent and young adult; ERS, European Respiratory Society; GvHD, Graft versus Host Disease; GLI, Global Lung Initiative; MRI, magnetic resonance imaging; LTFU, long-term follow-up; PFT, pulmonary function testing

We identified existing knowledge gaps for dose-response relationships of all studied exposures; newer chemotherapeutic or immunotherapeutic agents; other medical conditions, such as pulmonary complications during treatment, co-morbidities; impact of treatments on physiological processes affecting pulmonary function, such as lung growth and physiologic ageing; and also approaches for early assessment of pulmonary dysfunction and effects from preventive or curative interventions.

Most studies focused on well-established risk factors such as HSCT (31–37), thoracic radiotherapy (1–3, 31, 38, 40, 45–48, 50–52), and thoracic surgery (1, 31, 38–40). For chemotherapeutic agents—even those with previously reported pulmonary toxicity, such as bleomycin— we found clinical evidence for CAYA cancer survivors insufficient. Future studies evaluating other classical chemotherapeutic agents, targeted or immunotherapeutic agents, or pharmacovigilance data might identify new aspects of pulmonary dysfunction among CAYA cancer survivors. It is similar for radiotherapy, including a lack of data comparing photon and proton therapy where toxicity of protons might be lower from smaller irradiated volume compared with photons, possibly resulting in less lung-toxicity. For all exposures, we lack knowledge on how they interact with each other or how age at treatment or additional medical conditions modify impact from exposures; we also have little information about dose-response relationships.

Peak lung function attained in early adulthood and the trajectory of lung function decline with ageing are important for lung health across the life span. The impact of cancer itself, pulmonary complications during treatment, pulmonary co-morbidities, such as asthma, or impaired somatic growth, such as scoliosis, on peak attained lung function has not been examined. No studies examined whether CAYA cancer survivors start at a lower peak attained lung function or whether physiological ageing and decline in pulmonary function is faster and steeper than among the general population. Frailty and accelerated ageing were previously described for childhood cancer survivors (58–60). The definition of frailty is met when fulfilling three or more of five criteria: reduced lean muscle mass, weakness, slow walking speed, low energy expenditure, and fatigue (61). Ness et al. showed that components of frailty—reduced strength, walking speed, and increased fatigue—were as frequent among childhood cancer survivors from the St. Jude Lifetime Cohort at a median age of 33 years as among people age 65 years and older in the general population (62). By calculating the deficit accumulation index score, Williams et al. showed childhood cancer survivors acquire more damage and disease than community controls (60). Both studies suggest accelerated ageing among childhood cancer survivors. Factors contributing to accelerated ageing and frailty include more rapid cellular senescence, telomere length reduction, epigenetic modifications, somatic mutations, and mitochondrial DNA damage (58). These factors may also affect lung growth and function among children and adolescents or result in faster pulmonary ageing, but their potential impact among CAYA cancer survivors is unknown.

Another gap in knowledge concerns measuring early stages of pulmonary dysfunction. Prior studies primarily utilized spirometry, body plethysmography, or DLCO measurement. More sensitive tests, such as multiple breath washout tests, may identify pulmonary disease earlier and eventually contribute to better understanding of pulmonary dysfunction development among CAYA cancer survivors. Parisi et al. and Schindera et al. investigated pulmonary function of childhood cancer survivors using multiple breath washout tests (63, 64). Parisi et al. investigated 57 survivors with median follow-up time of 6.2 years from end of treatment; they did not show differences in ventilation homogeneity compared with controls (64). The 46 survivors evaluated by Schindera et al. were median 20 years from cancer diagnosis (63). Survivors defined as high risk (bleomycin, busulfan, nitrosoureas, HSCT, thoracic radiotherapy, or surgery) tended to have more ventilation inhomogeneity than those at standard risk (other cancer therapies), yet not significantly (63). In both studies, more survivors had abnormal washout tests than abnormal spirometry.

Available data made it impossible to reach conclusions about the longitudinal course of pulmonary function as survivors progress through childhood, puberty, and adulthood in their growth and development followed by a trajectory of ageing. Eight studies with repeated PFT results suffered from attrition bias, small sample sizes, and included sub-groups of CAYA cancer survivors, such as HSCT (2, 32, 33, 35, 37, 47, 52, 53). Ideally with baseline PFT before starting treatment, longitudinal data— ascertained at regular intervals from diagnosis—will be available improve knowledge about the onset of pulmonary dysfunction and its evolution.

PFT provides one way of assessing pulmonary health. Clinical symptoms or imaging are other possible modalities. Clinical symptoms lack objectivity and vary with age, which limits precise measurement. In addition, questions about clinical symptoms are worded differently between studies, which makes comparisons difficult. Louie et al validated selected self-reported complications from HSCT survivors (65). No data exist for other CAYA cancer survivor populations or for other questions about pulmonary dysfunction.

We suggest future studies take weaknesses and methodological problems we identified from the literature we reviewed into account and avoid them whenever possible (Table 5). Collaboration between paediatric oncologists and pulmonologists helps avoid shortcomings when conducting PFT and reporting results. Collaborative studies with harmonised protocols could maximise statistical power with larger numbers of CAYA cancer survivors and allow stratifying analyses into sub-groups defined by therapeutic exposures, age at treatment, cumulative doses, or genotype. Prospective rather than retrospective studies allow for standardising assessments, such as PFT at predefined time points, and minimise selection and attrition biases. Assessing patient-reported outcomes, such as symptoms, functional limitations, and quality of life, together with PFT, helps determine the clinical significance of findings and their impact on lives of patients and their families.

To obtain accurate measurements, PFT must be performed to high standards, by trained personnel explicitly applying published guidelines and standards, including ERS/ATS guidelines. Reporting, interpreting, and applying results in clinical practice are equally important. It is essential that future studies use GLI reference equations to standardise PFT results and make them comparable between age groups and regions (66, 67). Binary cut-offs—describing results as either normal or abnormal— reduce statistical power and introduce interpretations based on pre-defined threshold values. Since cut-off values differ between studies, such dichotomisation hampers comparisons of results, leading to conflicting and potentially misleading proportions of CAYA cancer survivors with pulmonary dysfunction. Reporting results as raw data and z-scores based on internationally agreed, age-adjusted reference values is preferred and allows comparing and pooling data.

We suggest studies investigating pulmonary function among CAYA cancer survivors be conducted in the knowledge that at present no curative treatments exist for suspected progressive inflammatory and fibrotic changes underlying pulmonary dysfunction. Therefore, we advise careful study of benefits and harms from repeat testing. However, awareness of impaired pulmonary function possibly leads to earlier treatment of bacterial infections with antibiotics, especially because excess pulmonary mortality and hospitalisations among CAYA cancer survivors are mainly from infection (68, 69). The U.S. Food and Drug Administration and European Medicines Agency approved two anti-fibrotic drugs—pirfenidone and nintedanib—for the treatment of idiopathic pulmonary fibrosis; patients with progressive-fibrosing unclassifiable interstitial lung disease possibly also benefit (70). Therefore, in the future the possible benefit from anti-fibrotic drugs could be an area for investigation among CAYA cancer survivors.

### Strengths and limitations

Strengths of our recommendation are multidisciplinary and international collaboration, which included perspectives from paediatric and adult specialists in CAYA cancer care and survivors; broad inclusion criteria; our thorough review process paired with in-depth quality assessment of included studies; and resulting evidence. Limitations mainly reflect lack of available evidence: studies with small sample sizes, heterogeneous PFT result reporting, use of different reference values; and scarce longitudinal data.

## Conclusion

CAYA cancer survivors treated with allogeneic HSCT, thoracic radiotherapy, and thoracic surgery were reported at risk for pulmonary dysfunction. However, our extensive literature search highlights the absence of robust evidence linking these exposures and pulmonary dysfunction because of small study sizes, high risks of bias, inconsistently assessing and reporting PFT results, and a lack of effective interventions to prevent the deterioration of asymptomatic pulmonary dysfunction. Therefore, our panel could not currently recommend routine PFT for asymptomatic CAYA cancer survivors. Yet, it is important for health care professionals and CAYA cancer survivors to be aware of possibly impaired pulmonary health and act vigilantly about appropriately investigating and following up when symptoms develop. We also recommend routine vaccinations recommended for people with pulmonary diseases and careful counselling relating to avoidance of tobacco products. Our results highlighted the current paucity of evidence, revealed relevant knowledge gaps, and emphasised that clearly defined, well-planned, harmonised, and collaborative studies and reports of pulmonary function outcomes are urgently needed to improve the body of evidence about pulmonary function among CAYA cancer survivors in the future.

## Data Availability

All data produced in the present work are contained in the manuscript.

## Acknowledgment

We thank Elio Castagnola and Cecile Ronckers for their contribution to the title and abstract and full text screening; Prof Thorsten Langer (Pediatric Oncology and Hematology, University Hospital for Children and Adolescents, Lübeck, Germany) and Daniel Mulrooney (St Jude Children’s Research Hospital, Memphis, TN, USA) for critically appraising the recommendations and manuscript as external reviewers; and Anna Apel (CEO of Saving Kids with Cancer Foundation, Poland; PanCare Board Member) and Katie Weyer as survivor representatives. Lastly, we thank Kristin Marie Bivens (Institute of Social and Preventive Medicine, University of Bern) for her editorial assistance.

## Author contributions

Concept and design: CEK, ACD, RLM, SHA, MMH, LK, RK

Literature search: CEK, RLM, RK, MO

Title and abstract screening: all authors

Full text screening: all authors

Data extraction: all authors

Risk of bias assessment: CEK, RLM, RK, MO, CS, MG, NW

Quality of evidence assessment: CEK, RLM, RK, MO, CS, MG, NW

Manuscript writing: all authors

Manuscript review: all authors

MO and RK have directly accessed and verified all underlying data reported in the manuscript.

All authors had full access to all the data in the study and accept responsibility for the submission for publication.

## Abbreviations

ATS: American Thoracic Society
CAYA: Childhood Adolescent and Young Adult
COG: Childrens’ Oncology Group
DCOG: Dutch Childrens’ Oncology Group
DLCO: Diffusion capacity for carbon-monoxide
ERS: European Respiratory Society
FEV1: Forced expiratory volume in the first second
FVC: Forced vital capacity
GLI: Global Lung Initiative
GRADE: Grading of Recommendations, Assessment, Development and Evaluation
GvHD: Graft versus Host Disease
HSCT: Haematopoietic Stem Cell Transplantation
IGHG: International Guideline Harmonization Group
LTFU: Long-term follow-up
MEF25-75%: Maximal expiratory flow between 25% and 75% of the FVC
PFT: Pulmonary function testing
PRISMA: Preferred Reporting Items for Systematic Reviews and Meta-Analyses
RV: Residual volume
TBI: Total Body Irradiation
TLC: Total lung capacity
UKCCLG: United Kingdom Children’s Cancer and Leukaemia Group

