## Supplementary material for "Recommendations for surveillance of pulmonary dysfunction among childhood, adolescent, and young adult cancer survivors: a report from the International Late Effects of Childhood Cancer Guideline Harmonization Group": Online Appendix

A) Guideline panel members	p. 3

B) Clinical questions	p. 4 – 5

C) Search strategies	p. 6 – 10

D) Inclusion and exclusion criteria	p. 11

E) Information extracted at fullt-text level – evidence summary tables	p. 12

F) Risk of Bias assessment criteria as stated by the IGHG Handbook	p. 13

G) GRADE quality assessment as stated by the IGHG Handbook	p. 14 – 15

H) Workflow on formulating recommendations for pulmonary function screening	p. 16

I) Concordances/discordances	p. 17 – 21

K) Evidence summary tables of included studies on	p. 22 – 71

“Who needs surveillance for pulmonary dysfunction?”

L) Summary of the evidence assessments and quality of data	p. 72 – 150

contributing to the recommendations

M) Evidence to decision Framework	p. 151 – 158

N) References	p. 159 – 161

### Abbreviations through the whole appendix

ATS	American Thoracic Society

BCNU	armustine

CAYA	Childhood Adolescent and Young Adult

CCNU	Lomustine

CCS	Childhood cancer survivors

CRT	Cranial radiotherapy

CSI	Craniospinal irradiation

COG	Childrens’ Oncology Group

CYC	Cyclophosphamide

DCOG	Dutch Childrens’ Oncology Group

DLCO	Diffusion capacity for carbon-monoxide

ERS	European Respiratory Society

FEV1	Forced expiratory volume in the first second

FTBI	fractionated total body irradiation

FVC	Forced vital capacity

GLI	Global Lung Initiative

GRADE	Grading of Recommendations, Assessment, Development and Evaluation

GvHD	Graft versus Host Disease

Gy	Gray

HDM	high-dose methotrexate

HSCT	Haematopoietic Stem Cell Transplantation

IGHG	International Guideline Harmonization Group

iMLD	individual lung dose

IU	international units

iV5	volume of individual lungs receiving 5Gy

KCO	carbon monoxide transfer coefficient

LLN	lower limit of normal

LTFU	Long-term follow-up

MAXRT	maximum of cumulative RT doses to left or right lungs

MEF25-75%	Maximal expiratory flow between 25% and 75% of the FVC

MLD	mean total lung dose

MTX	methotrexate

NS	not significant

PFT	Pulmonary function test

PRISMA	Preferred Reporting Items for Systematic reviews and Meta-Analyses

PY	pack years

RCT	Randomized clinical trail

RT	Radiotherapy

RV	Residual volume

SB	selection bias

SD	standard deviation

SFTBI	single-fraction total body irradiation

SIGN	Scottish Intercollegiate Guidelines Network

TBI	Total Body Irradiation

TLC	Total lung capacity

UKCCLG	United Kingdom Children’s Cancer and Leukaemia Group

V5	volume of both lungs receiving 5Gy

VA	Alveolar volume

WLI	whole lung irradiation

yr	year

%pred	percentage of predicted

### A) Guideline panel members

| **Name** | **Country** | **Area of expertise** | **Role** |
| --- | --- | --- | --- |
| **Core group** | | | |
| Claudia Kuehni | Switzerland | Paediatric Pneumology/ Epidemiology | Chair |
| Andrew Dietz | USA | Paediatric Oncology | Chair |
| Maria Otth | Switzerland | Paediatric Oncology | Coordinator |
| Rahel Kasteler | Switzerland | Paediatric Oncology | Coordinator |
| Leontien Kremer | The Netherlands | Expert guideline development | Advisor |
| Renée Mulder | The Netherlands | Expert guideline development | Advisor |
| Roderick Skinner | United Kingdom | Paediatric Oncology | Advisor |
| Melissa Hudson | USA | Pediatric Oncology | Advisor |
| Sandy Constine | USA | Radiation Oncology | Advisor |
| Kevin Oeffinger | USA | General Practitioner | Advisor |
| Saro Armenian | USA | Paediatric Oncology | Advisor |
| **Expert panel** | | | |
| Jennifer Agrusa | USA | Paediatric Oncology | WG member |
| Dana Barnea | Israel | General Practitioner | WG member |
| Anne Bergeron | Switzerland | Adult Pneumology | WG member |
| Neel Bhatt | USA | Paediatric Oncology | WG member |
| Stephen Bourke | United Kingdom | Adult Pneumology | WG member |
| Myrofora Goutaki | Switzerland | Paediatric Pneumology/ Epidemiology | WG member |
| Daniel Green | USA | Paediatric Oncology / Epidemiology | WG member |
| Ulrike Hennewig | Germany | Paediatric Oncology / Epidemiology | WG member |
| Véronique Houdouin | France | Paediatric Pneumology | WG member |
| Philipp Latzin | Switzerland | Paediatric Pneumology | WG member |
| Anthony Ng | United Kingdom | Paediatric Oncology | WG member |
| Cécile Ronckers | Germany | Epidemiology | WG member |
| Christina Schindera | Switzerland | Paediatric Oncology | WG member |
| Grit Sommer | Switzerland | Epidemiology | WG member |
| Saumini Srinivasan | USA | Paediatric Pneumology | WG member |
| Dennis Stokes | USA | Paediatric Pneumology | WG member |
| Birgitta Versluys | The Netherlands | Paediatric Oncology | WG member |
| Nicolas Waespe | Switzerland | Paediatric Oncology | WG member |
| Daniel Weiner | USA | Paediatric Pneumology | WG member |
| **Review of surveillance recommendation by patient stakeholders and external experts** | | | |
| Thorsten Langer | Germany | Paediatric Oncology | Ext. expert |
| Daniel Mulrooney | USA | Paediatric Oncology | Ext. expert |
| Anna Apel | Poland | Patient stakeholder | Ext. expert |
| Katie Weyer | USA | Patient stakeholder | Ext. expert |

### B) Clinical questions

Eleven clinical questions were formulated to cover all aspects and risk factors potentially associated with pulmonary dysfunction in childhood, adolescent, and young adult cancer survivors.

1.	What is the risk of pulmonary dysfunction in childhood and young adult cancer survivors (CAYA) treated with **allogeneic haematopoietic stem cell transplantation** compared to CAYA not treated with haematopoietic stem cell transplantation?

a.	What is the risk in younger compared to older age at treatment?

b.	What is the risk in patients with cGvHD compared to patients without cGvHD?

c.	What is the risk in patients who had a pulmonary infection during allogeneic HSCT compared to patients without pulmonary infection during allogeneic HSCT?

2.	What is the risk of pulmonary dysfunction in CAYA treated with **cyclophosphamide** compared to CAYA not treated with cyclophosphamide?

a.	What is the risk associated with different doses?

b.	What is the risk in younger compared to older age at treatment?

3.	What is the risk of pulmonary dysfunction in CAYA treated with **methotrexate** compared to CAYA not treated with methotrexate?

a.	What is the risk associated with different doses?

b.	What is the risk in younger compared to older age at treatment?

4.	What is the risk of pulmonary dysfunction in CAYA treated with **gemcitabine** compared to CAYA not treated with gemcitabine?

a.	What is the risk associated with different doses?

b.	What is the risk in younger compared to older age at treatment?

5.	What is the risk of pulmonary dysfunction in CAYA treated with **bleomycin** compared to CAYA not treated with bleomycin?

a.	What is the risk associated with different doses?

b.	What is the risk in younger compared to older age at treatment?

c.	What is the risk in patients with renal dysfunction versus patients without renal dysfunction?

i.	What is the threshold of renal dysfunction (GFR, creatinine, other)?

6.	What is the risk of pulmonary dysfunction in CAYA treated with **busulfan** compared to CAYA not treated with busulfan?

a.	What is the risk associated with different doses?

b.	What is the risk in younger compared to older age at treatment?

7.	What is the risk of pulmonary dysfunction in CAYA treated with **lomustine** (CCNU) or **carmustine** (BCNU) compared to CAYA not treated with lomustine (CCNU) or carmustine (BCNU)?

a.	What is the risk associated with different doses?

b.	What is the risk in younger compared to older age at treatment?

8.	What is the risk of pulmonary dysfunction in CAYA treated with **thoracic radiotherapy** compared to CAYA not treated with thoracic radiotherapy?

a.	What is the risk associated with different doses and volumes?

i.	Dose-volume relationship

ii.	the impact of the dose per fraction

b.	What is the risk in different radio therapeutic fields?

c.	What is the risk iassociated with patient age at the time of radiation?

d.	What is the risk of pulmonary dysfunction in CAYA treated with radiosensitizing/radiomimetic chemotherapy (doxorubicin, dactinomycin, busulfan, bleomycin, topotecan, irinotecan) combined with thoracic radiotherapy compared to CAYA not treated with radiomimetic chemotherapy combined with thoracic radiotherapy?

e.	What is the risk for patients treated with total body irradiation in the setting of stem cell transplantation?

9.	What is the risk of pulmonary dysfunction in CAYA treated with **surgery** (resection of lung tissue, thoracic cage or respiratory muscles) compared to CAYA not treated with surgery?

a.	What is the risk associated with different resection volumes?

b.	What is the risk in younger compared to older age at treatment?

10.	What is the risk of pulmonary dysfunction in CAYA treated with **combinations** of the therapies above?

a.	What is the risk of thoracic surgery combined with lung-toxic chemotherapy (bleomycin, CCNU, BCNU, busulfan, cyclophosphamide, methotrexate, gemcitabine)?

b.	What is the risk of thoracic surgery combined with thoracic radiotherapy?

c.	What is the risk of lung-toxic chemotherapy combined with thoracic radiotherapy?

11.	What is the risk of pulmonary dysfunction in CAYA who have a history of **tobacco exposure** compared to CAYA with no history of tobacco exposure?

a.	What is the risk in smokers/ex-smokers compared to non-smokers?

b.	What is the risk associated with different doses (pack-years)?

c.	What is the risk in patients exposed to environmental tobacco smoke compared to not exposed?

d.	What is the risk in marijuana smokers compared to non-smokers?

### C) Search strategies

**Search strategy for initial search in November 2016**

| Search 1a:  Childhood cancer cancer | (leukemia OR leukemi* OR leukaemi* OR (childhood ALL) OR AML OR lymphoma OR lymphom* OR hodgkin OR hodgkin* OR T-cell OR B-cell OR non-hodgkin OR sarcoma OR sarcom* OR sarcoma, Ewing's OR Ewing* OR osteosarcoma OR osteosarcom* OR wilms tumor OR wilms* OR nephroblastom* OR neuroblastoma OR neuroblastom* OR rhabdomyosarcoma OR rhabdomyosarcom* OR teratoma OR teratom* OR hepatoma OR hepatom* OR hepatoblastoma OR hepatoblastom* OR PNET OR medulloblastoma OR medulloblastom* OR PNET* OR neuroectodermal tumors, primitive OR retinoblastoma OR retinoblastom* OR meningioma OR meningiom* OR glioma OR gliom*) OR (pediatric oncology OR paediatric oncology) OR (childhood cancer OR childhood tumor OR childhood tumors)) OR (brain tumor* OR brain tumour* OR brain neoplasms OR central nervous system neoplasm OR central nervous system neoplasms OR central nervous system tumor* OR central nervous system tumour* OR brain cancer* OR brain neoplasm* OR intracranial neoplasm*) OR (leukemia, lymphocytic, acute) OR (leukemia, lymphocytic, acute*) |
| --- | --- |
| Search 1b:  Children, adolescents and young adults | Infan* OR toddler* OR minors OR minors* OR boy OR boys OR boyfriend OR boyhood OR girl* OR kid OR kids OR child OR child* OR children* OR schoolchild* OR schoolchild OR school child[tiab] OR school child*[tiab] OR adolescen* OR juvenil* OR youth* OR teen* OR under*age* OR pubescen* OR pediatrics[mh] OR pediatric* OR paediatric* OR peadiatric* OR school[tiab] OR school*[tiab] OR young adult[mh] OR young adult |
| Search 2:  Chemotherapy | Antineoplastic Protocols OR Antineoplastic Combined Chemotherapy Protocols OR Chemoradiotherapy OR Chemoradiotherapy, Adjuvant OR Chemotherapy, Adjuvant OR Consolidation Chemotherapy OR Induction chemotherapy OR Maintenance chemotherapy OR Chemotherapy, Cancer, Regional Perfusion OR Antineoplastic agents OR chemotherap* OR busulphan OR busulfan* OR Carmustine OR BCNU OR Chlorambucil OR cyclophosphamide OR cyclophosphane OR cytophosphan OR endox* OR cyclophospha* OR Lomustine OR CCNU OR lomustine* OR Mechlorethamine OR mechlorethamine* OR Chlormethine OR Mustine OR Chlorethazine OR doxorubicin OR doxorubic* OR bleomycin OR dactinomycin OR gemcitabine OR irinotecan OR methotrexate OR topotecan OR tacrolimus OR immunotherapy |
| Search 3:  Radiotherapy general  Field: thorax | (Radiotherapy OR radiation OR radiation therapy OR irradiation OR irradiat* OR radiation injuries OR injuries, radiation OR injury, radiation OR radiation injury OR radiation syndrome OR radiation syndromes OR syndrome radiation OR radiation sickness OR radiation sicknesses OR sickness radiation OR radiation* OR irradiation OR radiations)  AND  (TBI OR total body OR whole body OR total body* OR body whole* OR chest OR lung OR axilla OR mediastinal OR mantle OR supraclavicular OR susclavicular OR cranial axis OR total axis OR supra diaphragm[tiab] OR abdominal OR Inverted Y[tiab] OR Left Flank OR Hemiabdomen OR Left upper quadrant OR Paraaortic OR Spleen OR craniospinal) |
| Search 4:  HSCT | Stem cell transplant[mh] OR stem-cell transplant OR stem cell transplant*OR stem cell transplantation OR bone marrow transplantation[mh] OR transplantation, conditioning[mh] OR hematopoetic stem cell transplantation[mh] OR reduced-intensity conditioning regimen OR myeloablative agonists[mh] |
| Search 5: Pulmonary surgery | pulmonary metastasectomy OR pulmonary lobectomy OR thoracotomy OR sternotomy OR thoracoscopy OR rib resection[tiab] OR spinal surgery OR spinal fusion OR (resection AND (pulmonary wedge OR lung OR claviculae OR scapulae OR muscle tissue on thorax)) |
| Search 6:  Tobacco smoking | (tobacco OR nicotine OR cigarette OR e-cigarette OR cigar OR pipe OR environmental tobacco smoke OR second hand smoke OR ETS OR waterpipe OR narghile OR arghile OR shisha OR hookah OR marijuana OR joint OR MJ[tiab] OR spice OR thc OR cannabis) AND (smoking OR smoke OR smoke*) |
| Search 7:  Pulmonary disease | Pulmonary Fibrosis OR lung fibrosis OR (scarring AND (lung OR lungs*)) OR interstitial lung disease OR acute respiratory distress syndrome[tiab] OR ARDS OR respiratory distress syndrome OR shock lung[tiab] OR pneumonia OR COP[tiab] OR pneumonitis[tiab] OR pulmonitis[tiab] OR (lung AND (cancer OR carcinoma OR tumor)) OR lung neoplasms[mh] OR (lung AND (infection OR disease)) OR lung diseases[mh] OR (chest wall AND (abnormalit* OR disease)) OR kyphoscoliosis OR fibrothorax OR bronchitis OR bronchiectasis OR emphysema OR fibroelastosis OR Bronchiolitis OR BOS[tiab] OR BOOP OR cryptogenic organizing pneumonia[mh] OR cryptogenic organizing pneumonia[tiab] OR pulmonary disease OR pulmonary disease, chronic obstructive[mh] OR COPD OR pulmonary complications OR OSA OR respiratory tract diseases[mh] OR respiratory disease* OR low infectious respiratory disease OR respiratory defect OR apnea OR asthma |
| Search 8: Pulmonary functional consequences | ((Pulmonary OR respiratory) AND dysfunction) OR lung diseases, obstructive[mh] OR obstructive lung disease[tiab] OR restrictive lung disease[tiab] OR gas exchange impairment[tiab] OR ((ventilation OR respiration) AND (inhomogeneity OR inhomogeneous OR mismatch)) OR impaired diffusion capacity OR diffusion capacity impairment |
| Search 9:  Pulmonary symptoms | dyspnea OR cough OR mucus OR sputum OR hypoxia OR oygen requirement[tiab] OR exercise intolerance[tiab] OR respiratory sounds[mh] OR wheeze OR wheeze* OR breathlessness[tiab] OR shortness of breath OR chest pain OR chest discomfort[tiab] OR snore OR snoring OR hemoptysis OR oxygen requirement |
| Search 10:  Pulmonary diagnostic tests | respiratory function tests[mh] OR (function test AND (lung OR pulmonary OR respiratory)) OR spirometry OR bronchospasmolysis OR plethysmography OR DLCO OR diffusion capacity OR breath washout OR pulsoxymetry OR therapeutic irrigation[mh] OR broncho alveolar lavage[tiab] OR bronchoscopy OR blood gas analysis OR FEV1 OR forced expiratory volume OR LCI OR lung clearance index OR TLC OR total lung capacity OR FVC OR forced vital capacity OR PEF OR peak expiratory flow OR forced expiratory flow OR FEF OR maximum expiratory flow OR MEF OR KCO OR diffusion capacity OR maximal inspiratory pressure OR maximal expiratory pressure OR respiratory muscle pressure OR ((HR-CT OR MRI OR X-ray OR Biopsy OR lavage) AND (lung OR pulmonary OR chest OR thorax)) OR (transfer factor AND lung) |
| Search 11: Survivor | Survivor OR survivors OR survivor* OR long term survivor OR long term survivors OR long term survivor* OR survivo* OR surviving OR long term survival[tiab] OR survival[mh] |
| Search 12:  Late effects | "late effect" OR "late effects" OR "late effect*" OR "late side effect" OR "late side effects" OR "late side effect*" OR "late adverse effect" OR "late adverse effects" OR "late adverse effect*" OR long term effect[tiab] OR long term effect* OR long term adverse effects[mh] OR follow up studie* OR follow up study OR aftercare [mh] OR aftercare* OR after treatment [tiab] |
| **Filters: published since 1990; Humans; English language** | |

**Search strategy for search update in June 2019**

| 1. Cancer diagnoses in CAYA cancer patients | (leukemia OR leukemi* OR leukaemi* OR (childhood ALL) OR AML OR lymphoma OR lymphom* OR hodgkin OR hodgkin* OR T-cell OR B-cell OR non-hodgkin OR sarcoma OR sarcom* OR sarcoma, Ewing's OR Ewing* OR osteosarcoma OR osteosarcom* OR wilms tumor OR wilms* OR nephroblastom* OR neuroblastoma OR neuroblastom* OR rhabdomyosarcoma OR rhabdomyosarcom* OR teratoma OR teratom* OR hepatoma OR hepatom* OR hepatoblastoma OR hepatoblastom* OR PNET OR medulloblastoma OR medulloblastom* OR PNET* OR neuroectodermal tumors, primitive OR retinoblastoma OR retinoblastom* OR meningioma OR meningiom* OR glioma OR gliom*) OR (pediatric oncology OR paediatric oncology) OR (childhood cancer OR childhood tumor OR childhood tumors) OR (brain tumor* OR brain tumour* OR brain neoplasms OR central nervous system neoplasm OR central nervous system neoplasms OR central nervous system tumor* OR central nervous system tumour* OR brain cancer* OR brain neoplasm* OR intracranial neoplasm*) OR (leukemia, lymphocytic, acute) OR (leukemia, lymphocytic, acute*) |
| --- | --- |
| 2. Different age categories | Infan* OR toddler* OR minors OR minors* OR boy OR boys OR boyfriend OR boyhood OR girl* OR kid OR kids OR child OR child* OR children* OR schoolchild* OR schoolchild OR school child[tiab] OR school child*[tiab] OR adolescen* OR juvenil* OR youth* OR teen* OR under*age* OR pubescen* OR pediatrics[mh] OR pediatric* OR paediatric* OR peadiatric* OR school[tiab] OR school*[tiab] OR young adult[mh] OR young adult |
| 3. Combine | #1 AND #2 |
| 4. Potential pulmonary toxic chemotherapeutic agents | Antineoplastic Protocols OR Antineoplastic Combined Chemotherapy Protocols OR Chemoradiotherapy OR Chemoradiotherapy, Adjuvant OR Chemotherapy, Adjuvant OR Consolidation Chemotherapy OR Induction chemotherapy OR Maintenance chemotherapy OR Chemotherapy, Cancer, Regional Perfusion OR Antineoplastic agents OR hemotherapy* OR busulphan OR busulfan* OR Carmustine OR BCNU OR Chlorambucil OR cyclophosphamide OR cyclophosphane OR cytophosphan OR endox* OR cyclophospha* OR Lomustine OR CCNU OR lomustine* OR Mechlorethamine OR mechlorethamine* OR Chlormethine OR Mustine OR Chlorethazine OR doxorubicin OR doxorubic* OR bleomycin OR dactinomycin OR gemcitabine OR irinotecan OR methotrexate OR topotecan OR tacrolimus OR immunotherapy |
| 5. Radiotherapy involving the lung | (Radiotherapy OR radiation OR radiation therapy OR irradiation OR irradiat* OR radiation injuries OR injuries, radiation OR injury, radiation OR radiation injury OR radiation syndrome OR radiation syndromes OR syndrome radiation OR radiation sickness OR radiation sicknesses OR sickness radiation OR radiation* OR irradiation OR radiations) AND (TBI OR total body OR whole body OR total body* OR body whole* OR chest OR lung OR axilla OR mediastinal OR mantle OR supraclavicular OR susclavicular OR cranial axis OR total axis OR supra diaphragm[tiab] OR abdominal OR Inverted Y[tiab] OR Left Flank OR Hemiabdomen OR Left upper quadrant OR Paraaortic OR Spleen OR craniospinal) |
| 6. Hematopoietic stem cell transplantation | Stem cell transplant[mh] OR stem-cell transplant OR stem cell transplant* OR stem cell transplantation OR bone marrow transplantation[mh] OR transplantation, conditioning[mh] OR hematopoetic stem cell transplantation[mh] OR reduced-intensity conditioning regimen OR myeloablative agonists[mh] |
| 7. Surgery to the lung and thoracic cage | pulmonary metastasectomy OR pulmonary lobectomy OR thoracotomy OR sternotomy OR thoracoscopy OR rib resection[tiab] OR spinal surgery OR spinal fusion OR (resection AND (pulmonary wedge OR lung OR clavicula* OR scapula* OR muscle tissue OR thorax)) |
| 8. Combine | #4 OR #5 OR #6 OR #7 |
| 9. Smoking | (tobacco OR nicotine OR cigarette OR e-cigarette OR cigar OR pipe OR environmental tobacco smoke OR second hand smoke OR ETS OR waterpipe OR narghile OR arghile OR shisha OR hookah OR marijuana OR joint OR MJ[tiab] OR spice OR thc OR cannabis) AND (smoking OR smoke OR smoke*) |
| 10 Pulmonary disease | Pulmonary Fibrosis OR lung fibrosis OR (scarring AND (lung OR lungs*)) OR interstitial lung disease OR acute respiratory distress syndrome[tiab] OR ARDS OR respiratory distress syndrome OR shock lung[tiab] OR pneumonia OR COP[tiab] OR pneumonitis[tiab] OR pulmonitis[tiab] OR (lung AND (cancer OR carcinoma OR tumor)) OR lung neoplasms[mh] OR (lung AND (infection OR disease)) OR lung diseases[mh] OR (chest wall AND (abnormalit* OR disease)) OR kyphoscoliosis OR fibrothorax OR bronchitis OR bronchiectasis OR emphysema OR fibroelastosis OR Bronchiolitis OR BOS[tiab] OR BOOP OR cryptogenic organizing pneumonia[mh] OR cryptogenic organizing pneumonia[tiab] OR pulmonary disease OR pulmonary disease, chronic obstructive[mh] OR COPD OR pulmonary complications OR OSA OR respiratory tract diseases[mh] OR respiratory disease* OR low infectious respiratory disease OR respiratory defect OR apnea OR asthma |
| 11. Types of pulmonary function impairment | ((Pulmonary OR respiratory) AND dysfunction) OR lung diseases, obstructive[mh] OR obstructive lung disease[tiab] OR restrictive lung disease[tiab] OR gas exchange impairment[tiab] OR ((ventilation OR respiration) AND (inhomogeneity OR inhomogeneous OR mismatch)) OR impaired diffusion capacity OR diffusion capacity impairment |
| 12. Pulmonary symptoms | dyspnea OR cough OR mucus OR sputum OR hypoxia OR oygen requirement[tiab] OR exercise intolerance[tiab] OR respiratory sounds[mh] OR wheeze OR wheeze* OR breathlessness[tiab] OR shortness of breath OR chest pain OR chest discomfort[tiab] OR snore OR snoring OR hemoptysis OR oxygen requirement |
| 13. Pulmonary function parameters | respiratory function tests[mh] OR (function test AND (lung OR pulmonary OR respiratory)) OR spirometry OR bronchospasmolysis OR plethysmography OR DLCO OR diffusion capacity OR breath washout OR pulsoxymetry OR therapeutic irrigation[mh] OR broncho alveolar lavage[tiab] OR bronchoscopy OR blood gas analysis OR FEV1 OR forced expiratory volume OR LCI OR lung clearance index OR TLC OR total lung capacity OR FVC OR forced vital capacity OR PEF OR peak expiratory flow OR forced expiratory flow OR FEF OR maximum expiratory flow OR MEF OR KCO OR diffusion capacity OR maximal inspiratory pressure OR maximal expiratory pressure OR respiratory muscle pressure OR ((HR-CT OR MRI OR X-ray OR Biopsy OR lavage) AND (lung OR pulmonary OR chest OR thorax)) OR (transfer factor AND lung) |
| 14. Combine | #10 OR #11 OR #12 OR #13 |
| 15. Survivors | Survivor OR survivors OR survivor* OR long term survivor OR long term survivors OR long term survivor* OR survivo* OR surviving OR long term survival[tiab] OR survival[mh] |
| 16. Late effects | "late effect" OR "late effects" OR "late effect*" OR "late side effect" OR "late side effects" OR "late side effect*" OR "late adverse effect" OR "late adverse effects" OR "late adverse effect*" OR long term effect[tiab] OR long term effect* OR long term adverse effects[mh] OR follow up studie* OR follow up study OR aftercare [mh] OR aftercare* OR after treatment [tiab] |
| 17. Combine | #15 OR #16 |
| 18. Combine | #3 AND #8 AND #14 AND #17  (not taking smoking into account) |
| 19. Combine | #3 AND #9 AND #14 AND #17  (taking smoking into account instead of chemo, radio, surgery and transplant) |
| 20. Combine | #18 OR #19 |
| 21. Humans only | animals[mh] NOT humans[mh] |
| 22. Combine | #20 NOT #21 |
| 23. Date restriction | Search("2018/01/01"[Date - Publication] : "2019/02/28"[Date - Publication]) |
| 24. Combine | #22 AND #23 |

**Search strategy for search update in December 2020**

We used exactly the same search strategy as in June 2019 and restricted the search to: ("2019/02/28"[Date - Publication] : "2020/12/01"[Date - Publication])

**Search strategy for search update in June 2022**

We used exactly the same search strategy as in June 2019 and restricted the search to: ("2020/12/02"[Date - Publication] : "2022/06/01"[Date - Publication])

**Search strategy for search update in April 2023**

We used exactly the same search strategy as in June 2019 and restricted the search to: ("2022/06/02"[Date - Publication] : "2023/04/01"[Date - Publication])

### D) Inclusion and exclusion criteria

Inclusion criteria

- Survivors of childhood, adolescent, and young adult cancer (≥50% of population diagnosed prior to age 30 years)
- Outcome assessed ≥2 years after end of treatment
- Pulmonary function test results as pulmonary outcome
- Risk factor analysis performed
- Number of CAYA cancer survivors ≥ 20

Exclusion criteria

- Language other than English
- Case report, case series, abstracts
- Outcome reported as:
  - Self-reported pulmonary diseases or symptoms (e.g. Childhood Cancer Survivor Study)
  - Physician reported disease (e.g. bronchiolitis obliterans)
  - Combinations (e.g. “abnormal” as combination of symptoms, disease and/or pulmonary function parameter)
- Outcome reported as prevalence only but without risk factor analysis

### E) Information extracted at full-text level – evidence summary tables

| **General information**   - Author and year - Study design - Treatment era - Years of follow-up | **Participants**   - Study population - Diagnosis - Age at diagnosis - Age at follow-up |
| --- | --- |
| **Treatment**  HSCT, Cyclophosphamide, Methotrexate, Gemcitabine, Bleomycin, Busulfan, Lomustine, Carmustine, thoracic radiotherapy, surgery, combinations, tobacco exposure | **Main outcome**   - Pulmonary function test results: absolute values, z-scores, percentage of predicted, percentage with pathological test - Extraction of results |
| **Additional remarks**   - Longitudinal data available? - Control group mentioned? - Reference values stated? - Quality check of PFT performed? - Lung function procedure stated? - Cleaning of lung function data described? - Was person who analyzed PFT blinded to the exposure? - Describe analysis performed | |

### F) Risk of Bias assessment criteria as stated by the IGHG Handbook

Based on the studies resulting from the systematic literature search, we could include observational studies only, we therefore used the corresponding risk of bias assessment criteria below.

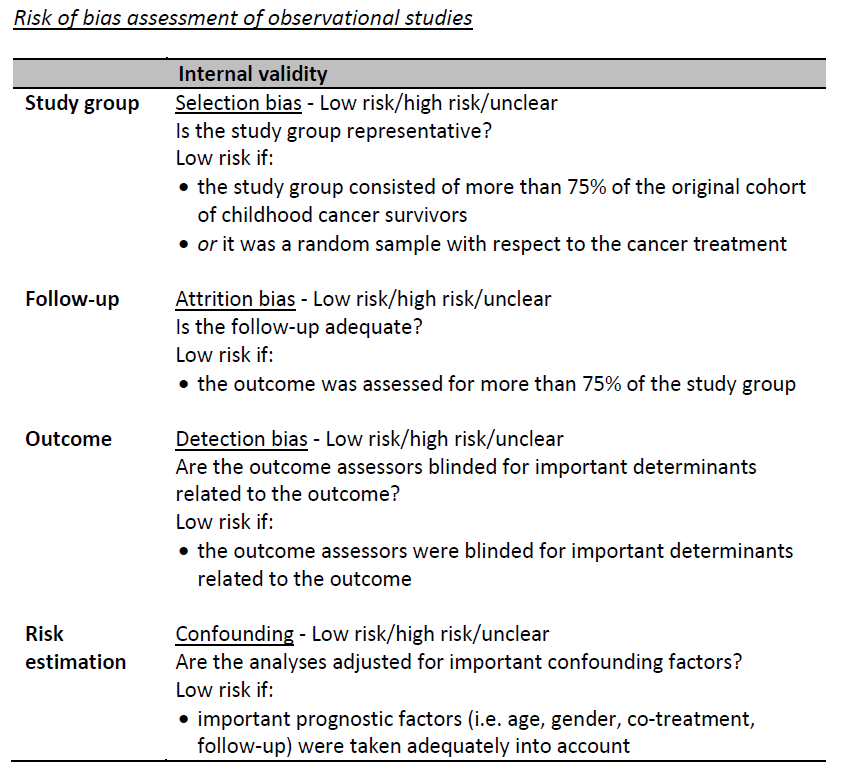

### G) GRADE quality assessment as stated by the IGHG Handbook

Initial score based on type of evidence

- +4: RCTs/ SR of RCTs
- +2: CCTs or observational evidence (e.g., cohort, case-control) for intervention questions
- +4: Observational evidence for etiologic, prognostic and diagnostic questions

**Factors that might decrease the quality of the body of evidence**

1. Study limitations: risk of bias based on selection bias, attrition bias, detection bias and confounding as defined in the risk of bias table.

- 0: No problems
- -1: Problem with 1 element
- -2: Problem with 2 elements
- -3: Problem with 3 or more elements

2. Consistency: degree of consistency of effect between or within studies

- 0: All/most studies show similar results
- -1: Lack of agreement between studies (statistical heterogeneity / conflicting result, e.g. effect sizes in different directions)

3. Directness: the generalizability of population and outcomes from each study to the population of interest

- 0: Population and outcomes broadly generalizable
- -1: Problem with 1 element (population different from the defined inclusion criteria OR outcomes different from the defined inclusion)
- -2: Problem with 2 elements (population and outcomes)

4. Precision: the precision of the results

- 0: No important imprecision when studies include many patients and many events and thus have narrow confidence intervals; determine with the chairs and advisors what is seen as many patients, many events and narrow confidence intervals
- -1: Important imprecision when studies include relatively few patients and few events and thus have wide confidence intervals (especially when the confidence interval cross the 0). Another criterion to consider is the clinical decision threshold. This is the threshold of the effect size that would change the decision whether or not to adopt a clinical action. Downgrade if the effect estimates and confidence intervals cross the clinical decision threshold. Determine with the chairs and advisors the clinical decision threshold.
- OR if only one study has been identified
- -2: If there is important imprecision (see -1) AND if only one study has been identified

5. Publication bias: if investigators fail to report studies and outcomes (typically those that show no effect)

- 0: Publication bias unlikely
- -1: Risk of publication bias when for example published evidence is limited to industry funded trials

**Factors that might increase the quality of the body of evidence**

1. Magnitude of effect:

- +1: Large magnitude of effect; all studies show significant effect sizes (point estimate) >2 or <0.5
- +2: Very large magnitude of effect; all studies show significant effect sizes (point estimate) >5 or <0.2

2. Dose response gradient:

- +1: Evidence of clear relation with increases in the outcome with higher exposure levels across or within studies

3. Plausible confounding:

- +1: If adjustment for confounders would have increased the effect size; for example, the estimate of effect is not controlled for the following possible confounders: smoking, degree of education, but the distribution of these factors in the studies is likely to lead to an underestimate of the true effect

**Total score**

⊕⊕⊕⊕ High quality evidence

⊕⊕⊕⊖ Moderate quality evidence

⊕⊕⊖⊖ Low quality evidence

⊕⊖⊖⊖ Very low quality evidence

### H) Workflow on formulating recommendations for pulmonary function screening

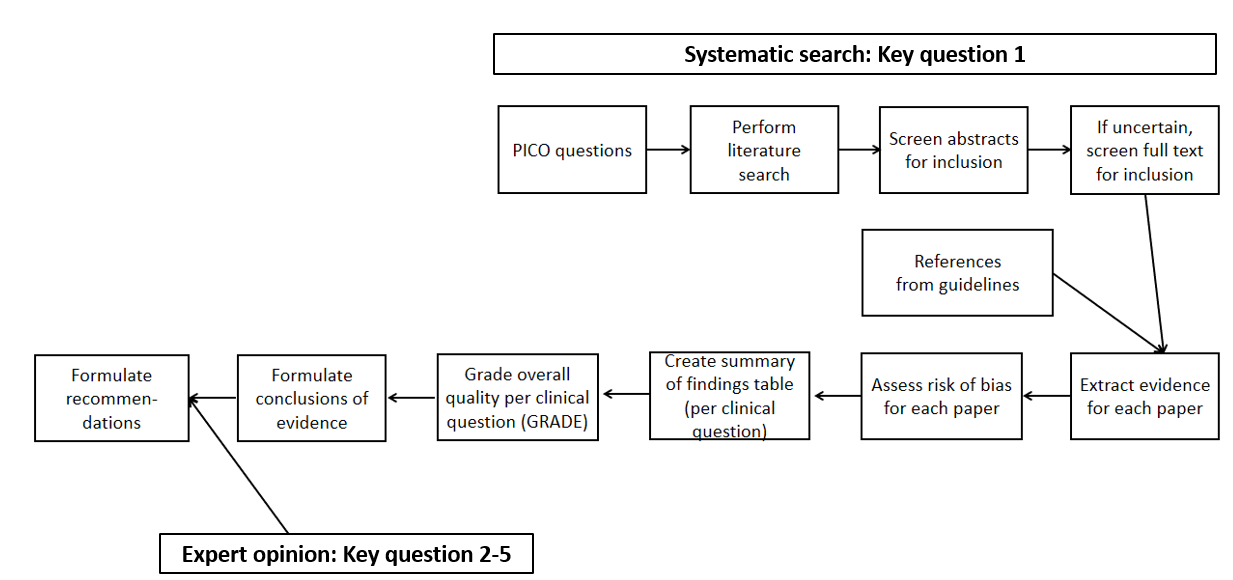

### I) Concordances/discordances

**Who needs surveillance?**

| **Who needs surveillance?** | | | | |
| --- | --- | --- | --- | --- |
|  | **North American Children’s**  **Oncology Group** | **Dutch Childhood**  **Oncology Group** | **UK Children’s Cancer and Leukaemia Group** | **Concordant/**  **Discordant** |
| **At risk** |  |  |  |  |
| Allogeneic HSCT | Yes (TBI, busulfan, cGvHD) | No (Not mentioned) | Yes | Discordant |
| Bleomycin | Yes | Yes | Yes (little evidence of late toxicity) | Concordant |
| Busulfan | Yes | Yes | Yes | Concordant |
| Nitrosureas | Yes | Yes | Yes | Concordant |
| Radiotherapy | Yes | Yes | Yes | Concordant |
| Surgery | Yes | Yes | Yes | Concordant |
| **At higher risk** (=treatment factors mentioned in the guidelines that may increase the risk) | | | | |
| HSCT | Yes (especially if cGvHD) | No (Not listed separately) | Yes (especially if TBI, busulfan, cGvHD) | Discordant |
| Higher cumulative bleomycin dose | Yes (≥400 U/m^2^) | No (No dose specified) | No (No dose specified)) | Discordant |
| Higher busulfan dose | Yes (≥500 mg – “HSCT dose”) | No (No dose specified) | No (No dose specified) | Discordant |
| Higher cumulative dose BCNU | Yes (≥600 mg/m^2^) | No (No dose specified) | Yes (No dose specified) | Discordant |
| Higher radiotherapy dose | Yes (RT ≥15 Gy; TBI ≥6 Gy single fraction, TBI ≥12 Gy fractionated) | No (No dose specified) | Yes (but: no dose specified) | Discordant |
| Larger radiotherapy treatment volume | No (Not mentioned) | No (No volume specified) | Yes (but: no volume specified) | Discordant |

|  | North American Children’s  Oncology Group | Dutch Childhood  Oncology Group | UK Children’s Cancer and Leukemia Group | Concordant/  Discordant |
| --- | --- | --- | --- | --- |
| Combinations of above | Yes  Busulfan, BCNU combined with chest RT or TBI  belomycin combined with chest RT or TBI  surgery combined with alkylating agents or bleomycin or chest RT or TBI | No (No combinations specified) | No (No combinations specified) | Discordant |
| Combination of radiotherapy and radiomimetic chemotherapy | Yes (doxorubicin, dactinomycin) | No (No combinations specified) | No (No combinations specified) | Discordant |
| Younger age | Yes (bleomycin, radiotherapy no age specified) | No (Not mentioned) | Yes (BCNU <5yrs, radiotherapy) | Discordant |
| Renal dysfunction (bleomycin) | Yes (not further specified) | No (Not mentioned) | No (Not mentioned) | Discordant |
| cGvHD / Immunosuppress. | Yes | No (Not mentioned) | Yes | Discordant |
| Pulmonary infection (HSCT) | No (Not mentioned) | No (Not mentioned) | Yes | Discordant |
| Tobacco: smoking /  marijuana | Yes | No (Not mentioned) | No (Not mentioned) | Discordant |

**Risk factors added by experts (currently not in guidelines)**

|  | **North American Children’s**  **Oncology Group** | **Dutch Childhood**  **Oncology Group** | **UK Children’s Cancer and Leukaemia Group** | **Concordant/**  **Discordant** |
| --- | --- | --- | --- | --- |
| Cyclophosphamide | Not included in guideline | Not included in guideline | Not included in guideline | - |
| Methotrexate | Not included in guideline | Not included in guideline | Not included in guideline | - |
| Gemcitabine | Not included in guideline | Not included in guideline | Not included in guideline | - |

**What surveillance modality should be used?**

**At what frequency should surveillance be performed?**

**When should surveillance be initiated?**

**What should be done when abnormalities are identified?**

|  | **North American Children’s**  **Oncology Group** | **Dutch Childhood**  **Oncology Group** | **UK Children’s Cancer and Leukaemia Group** | **Concordant/**  **Discordant** |
| --- | --- | --- | --- | --- |
| **What surveillance modality should be used?** | | | | |
| Clinical history | Yes (cough, shorness of breath, dyspnea on exertion, Wheezing) | No (Not mentioned) | Yes (exercise tolerance, smoking) | Discordant |
| Physical examination | Yes (pulmonary exam) | No (Not mentioned) | Yes (respiratory system) | Discordant |
| Pulmonary function tests | Yes (further specified) | Yes (further specified) | Yes (not further specified) | Concordant |
| Spirometry | Yes | Yes (flow volume curve) | Not specified | Discordant |
| Body plethismography | No (Not mentioned) | Yes (total lung capacity) | Not specified | Discordant |
| DLCO | Yes | Yes (DLCO/VA) | Not specified | Discordant |
| Radiology examination | Yes | No (Not mentioned) | Yes | Discordant |
| Chest x-ray | No (deleted in version 4.0) | No (Not mentioned) | Yes (if symptomatic or pulmonaty function test abnormal) | Discordant |
| CT | Yes (discuss for patient with high risk for lung cancer (chest RT and smoking)) | No (Not mentioned) | Yes (high-resolution CT if symptomatic or abnormal) | Discordant |

| **At what frequency should surveillance be performed and when be initiated?** | | | | |
| --- | --- | --- | --- | --- |
| Physical examination | Yes (Yearly) | No (Not mentioned) | Yes (at LTFU clinic, all patients) | Discordant |
| Clinical history | Yes (Yearly) | No (Not mentioned) | Yes (at LTFU clinic, all patients) | Discordant |
| Pulmonary function tests | At entry into LTFU, at least 2 years after end of cancer treatment, thereafter as clinically indicated in pat with abnormal results or progressive dysfunction | 5 and 10 years after diagnosis, then every 5 years if abnormal pulmonary function test (<75% predicted) | End of treatment, then after 1 year if symptomatic or abnormal pulmonary function test (<2SD below normal) | Discordant |
| Pulmonary function tests post-HSCT | At entry into LTFU, thereafter as clinically indicated | No (Not mentioned) | Pre-HSCT, 1 year post-HSCT, then every 1-3/5 years depending on symptoms and pulmonary function test results | Discordant |
| Radiology examination | Yes  Discuss for patient for high risk for Lung cancer (chest RT and smoking) | No (Not mentioned) | Yes (if symptomatic) | Discordant |

| **What should be done when abnormalities are identified?** | | | | |
| --- | --- | --- | --- | --- |
| Consider specialist referral | Yes (if symptomatic or progressive) | Yes (if symptomatic) | Yes (if symptomatic or abnormal pulmonary function test) | Concordant |
| Warn anaesthetists about previous bleomycin treatment | Yes  (consider repeated pulmonary function test before anesthesia if bleomycin busulfan, BCNU, CCNU) | Yes  (no exposure to FiO2>30% after bleomycin >400mg/m2 and/or RT to thorax) | Yes  (but nothing specified) | Concordant |
| Preventive measures | Yes | Yes  (FEV1/FVC or DLCOcorr/VA <75%pred or have >20% reduction from baseline) or recurrent respiratory infection/ chronic cough) | Yes | Partly concordant |
| Consider pneumococcal and influenza immunization | Yes (influenza and pneumococcal) | Yes (influenza if abnormalities in pulmonary function test as described above) | Yes (influenza and pneumococcal if established lung disease) | Partly concordant |
| Tobacco smoking | Yes (abstain) | Yes (abstain) | Yes (abstain) | Concordant |
| Inhaled drug use (marijuana) | Yes (abstain) | No (Not mentioned) | No (Not mentioned) | Discordant |
| Physical exercise | Yes (regular physical exercise) | No (Not mentioned) | No (Not mentioned) | Discordant |
| Environmental tobacco exposure (e.g., parents smoking) | Yes (avoid) | No (Not mentioned) | No (Not mentioned) | Discordant |
| Advice on career choice | Yes  Follow safety rules, don’t’ inhale toxic substances (chemicals, solvents, paints) use protective ventilators and report unsafe work conditions | Yes (avoid toxic substances) | No (Not mentioned) | Discordant |
| Scuba diving | Yes  (if busulfan, BCNU, CCNU, HSCT, RT to chest, bleomycin get advice from pulmonologist) | Yes (Not mentioned) | No (Not mentioned) | Discordant |
| Therapeutic approaches | No (Not mentioned) | No (Not mentioned) | Yes  (consider immunosuppression in chronic pulmonary disease with chronic GvHD) | Discordant |

### J) Evidence summary tables of included studies on “Who needs surveillance for pulmonary dysfunction?”

| **Main findings/message:**  **In univariate analysis hyperinflation significantly more frequent in pediatric cancer survivors in BMT versus no BMT (52.2% vs. 31.6%, P=0.01).**  **In univariate analysis any pulmonary abnormality, obstructive and hyperinflation are significantly more frequent in pediatric cancer survivors in not exposed to bleomycin versus exposed (72% vs 52%, p=0.02; 33% vs 12% p=0.01; 52% vs 21% p=<0.01).**  **In univariate analysis any pulmonary abnormality and obstructive disease are significantly more frequent in pediatric cancer survivors exposed to lung surgery versus no lung surgery (83.3% versus 61.3%, P=0.03).**  **In univariate analysis the prevalence of any pulmonary abnormality, obstructive, restrictive, and hyperinflation were not significantly different between exposed and non-exposed.** | | | | |
| --- | --- | --- | --- | --- |
| **Record, et al.** Analysis of Risk Factors for Abnormal Pulmonary Function in Pediatric Cancer Survivors. 2016; 63:1264-71. 10.1002/pbc.25969 | | | | |
| **Study design**  **Treatment era**  **Years of follow-up** | **Participants** | **Treatment**  (= treatment analyzed in the paper) | **Main outcomes** | **Additional remarks** |
| ☒ **Cohort**  ☐ **Cross-sectional**  ☐ **Case-control**  ☐ **Other:**  **__________**  ☒ **Retrospective**  ☐ **Prospective** | Study population (N)   - Original cohort: NA - Eligible cohort: 226 - Analyzed cohort:143 | ☒ 1 **HSCT**  ☐ **2 Cyclophosphamid**  ☐ **3 Methotrexate**  ☐ **4 Gemcitabine**  ☒ **5 Bleomycin**  ☐ **6 Busulfan**  ☐ **7 Lomustin (CCNU)**  ☐ **8 Carmustin (BCNU)**  ☒ **9 Radiotherapy lung**  ☒ **10 Surgery**  ☐ **11 Combinations**  ☐ **12 Tobacco exposure** | ☒ **Pulmonary diseases**  ☐ **Pulmonary symptoms**  ☒ **Pulmonary function test**  ☐ **Absolute values**  ☐ **Z-scores**  ☐ **Percentage predicted**  **☒ Percentage pathological tests**  (e.g. 24% with reduced FEV1) | ☐ **Longitudinal data available**  **☐ Control group mentioned**  **☒ Reference values stated**  Wang, Hankinson  **☐ Quality check performed**  **☒ Lung function procedure stated**  **☐ Cleaning of lung function data**  **described**  **☒ Person who analyzed PFT was**  **blinded to the exposure** |
| Centres:  Single center (Atlanta survivor clinic)  Country:  USA  Treatment era:  2000-2009  (7 pat dx. in late 1990)  Years of Follow-up:  Mean age: 14.1 ±4.8 yrs  2002-2012 | Study population:  Eligible (N): 226  Analysis (N): 143  (response rate 63.3%)  Inclusion criteria: therapy after 2000 with at least one pulmonary toxic treatment  Except children < 5 years and brain tumor  Cancer diagnosis:  Leukemia 28%  Hodgkin 28%  Non-hodgkin lymphoma 7%  Neuroblastoma: 9.8%  Renal tumor 10.5%  Sarcoma 0.1%  Other: 7.7%  Age at diagnosis:  Median 9 yr (0-21.8)  Age at follow-up:  Mean age at evaluation: 14.1 ± 4.8 yr | Chemotherapy:  Bleomycine 33.6%  Busulfan, Carnustine (BCNU), Lomustine (CCNU): 11.9%  Radiotherapy: 67.8%  Surgery: 16.8%  Bone Marrow Transplantation: 46.9% | Definition of outcome   1. Prevalence of pulmonary function abnormalities 2. Risk factors with these PF abnormalities (proportion)   PFT: spirometry, body plethysmography, DLco  Abnormal PFT if %-predicted pathological:  **Restrictive =** TLC <80%  **Obstructive =**FVC <80%, FEV1<80% or FEV1/FVC<80%, or FEF25-75% <68%  **Hyperinflation** = RV>120% or RV/TLC >28%  **Pulmonary vascular disease** =DLco/VAadj <4ml/mmHg  **Symptoms:** medical record abstraction  Results:  **Any abnormal PFT in n=93 (65%),** 21% having multiple abnormalities, **80% being asymptomatic**  Hyperinflation n=59/129 (41.3%)  Restrictive disease n=19/129 (13.3%)  Obstructive disease n=37/143 (25.9%)  Pulmonary vascular disease n=6/110 (5.5%)  **Risk factors**:  **BMT versus no BMT**:  Hyperinflation 52.2% vs. 31.6% (P=0.01)  **Bleomycin versus no Bleomycin:**  Bleomycin has sign. lower percentages of any abnormality, obstructive and hyperinflation compared to no Bleomycin  **Thoracic RT versus no thoracic RT:**  No sig. difference  **Lung surgery versus no lung surgery:**  Any abnormality 83.3% versus 61.3% (P=0.03)  Obstructive 50% versus 21% (P=0.01)  **Exact results available from publication, TABLE III: “**Univariate Comparison of Demographis, Diagnosis and Treatment Characteristics by PFT Abnormality to Those without the PFT Abnormality Among Pediatric Cancer Survivors at Risk for Pulmonary Late Effects” | Analysis: univariate comparison of treatment characteristics by PFT abnormality  Limitations:  - Only univariate analysis performed  - BMT not stratified into allo & auto, but most probably all allo BMT as GvHD reported  - Patients without RV or TLC classified as normal if no other abnormality |

| **Main findings/message:**  **In univariate analysis no significant association between bleomycin exposure and restrictive disease (OR 0.7, 95%CI 0.3-1.6) and DLco abnormalities (OR 0.8, 95%CI 0.4-1.7)**  **In univariate analysis no significant association between busulfan exposure and restrictive disease (OR 0.8, 95%CI 0.2-2.9) and DLco abnormalities (OR 0.4, 95%CI 0.1-1.6)**  **In univariate analysis no significant association between CCNU or BCNU exposure and restrictive disease (OR 1.1, 95%CI 0.3-4.2) and DLco abnormalities (OR 1.4, 95%CI 0.6-4.7)**  **In multivariable analysis significant association between increasing doses of chest radiation and restrictive disease (20 Gy: OR 5.6 (95%CI 1.5-21.0), p<0.05). Significant association between increasing doses of chest radiation and DLCO abnormality (≤20 Gy: OR 6.4 (95%CI 1.7-24.4), p<0.01; 20 Gy: OR 11.3 (95%CI 2.6-49.5). p<0.01). Increasing chest radiation doses are significant predictors of decline in DLco longitudinally (20 Gy: OR 24.4 (95%CI 5.7-38.3), p<0.01).**  **In univariate analysis no significant association between history of smoking and restrictive disease (OR 0.9, 95%CI 0.7-1.9) and DLco abnormalities (OR 0.9, 95%CI 0.2-5.3)** | | | | |
| --- | --- | --- | --- | --- |
| **S. H. Armenian, et al.** Long-term pulmonary function in survivors of childhood cancer. 2015;33:1592-600. 10.1200/jco.2014.59.8318 | | | | |
| **Study design**  **Treatment era**  **Years of follow-up** | **Participants** | **Treatment**  (= treatment analyzed in the paper) | **Main outcomes** | **Additional remarks** |
| ☒ **Cohort**  ☐ **Cross-sectional**  ☐ **Case-control**  ☐ **Other:**  **__________**  ☐ **Retrospective**  ☒ **Prospective** | Study population (N)   - Original cohort: NA - Eligible cohort: 155   Analysed cohort: 121 | ☐ 1 **HSCT a, b**  ☐ **2 Cyclophosphamid**  ☐ **3 Methotrexate**  ☐ **4 Gemcitabine**  ☒ **5 Bleomycin**  ☒ **6 Busulfan**  ☒ **7 Lomustin (CCNU)**  ☒ **8 Carmustin (BCNU)**  ☒ **9 Radiotherapy lung, 9a**  ☐ **10 Surgery**  ☐ **11 Combinations**  ☒ **12 Tobacco exposure** | ☐ **Pulmonary diseases**  ☐ **Pulmonary symptoms**  ☒ **Pulmonary function test**  ☐ **Absolute values**  ☐ **Z-scores**  ☒ **Percentage predicted**  **☐ Percentage pathological tests**  (e.g. 24% with reduced FEV1) | ☒ **Longitudinal data available**  **☒ Control group mentioned**  **☐ Reference values stated**  **☐ Quality check performed**  **☒ Lung function procedure stated**  **☐ Cleaning of lung function data**  **described**  **☒ Person who analyzed PFT was**  **blinded to the exposure** |
| Centres:  Single center, City of Hope Survivorship Clinic  Country: USA  Treatment era:  1972-2007  Years of Follow-up:  Time dx to t2: 17.1 yrs (range 6.3-40.1 yrs)  Time t1 to t2: median of 5 yrs (1-10.3 yrs) | Study population:  Eligible at t1 (N): 155  Analysis at t2 (N): 121  Response rate=78.1%  Controls:  General population, age- and sex-matched  Inclusion criteria:  Survivors diagnosed <age 22, with ≥2 yrs post diagnosis, treated with pulmonary-toxic chemotherapy and/or radiation and/or allogeneic HCT with cGVHD or pulmonary and/or surgery  Cancer diagnoses:  HL 34%  NHL 6%  Leukemia 36%  Sarcoma 11%  Other 14% (not specified)  Age at diagnosis (yrs):  Median (range): 16.5 (0.2-21.9)  Age at follow-up (t2) (yrs):  Median (range): 32.2 (14.6-58.9) | Chemotherapy (median doses) and %any  Bleomycin (60 IU/m2), 35%  Busulfan (436 mg/m2), 12%  BCNU/CCNU (450 mg/m2), 10%  Radiotherapy (median doses, range)  chest (13.2 Gy, 2-76):  26% no radiotherapy  50% ≤20 Gy  24% >20 Gy  Surgery  6% lobectomy, wedge resection or metastasectomy  HSCT (53%)  Autologous 17%  Allogeneic 36% | Pulmonary function assessment:  - PFT at baseline (t1) and at follow-up (t2)  - Compared with healthy controls (at t2)  - PFTs performed according to ATS protocols  - PFT parameters measured: TLC, FVC, FEV1, FEV1/FVC, DLco, DLco/Va  - %-predicted calculated by using established reference values (reference not stated)  - cut-offs:  obstructive FEV1/FVC<0.7, FEV1<80% predicted; restrictive TLC<75%, FEV1≥80% predicted  diffusion DLco<75% predicted  **Comparison survivors – survivors with risk factor analysis** (univariable analysis, if sig -> multivariable regression analysis)  **Bleomycin**:  - no significant association between bleomycin exposure and restrictive disease: univariable OR 0.7, 95%CI 0.3-1.6  - no significant association between bleomycin exposure and DLCO abnormality: univariable OR 0.8, 95%CI 0.4-1.7 (no multivariable anaylsis performed because not significant!)  **Busulfan**:  - no significant association between busulfan exposure and restrictive disease: univariable OR 0.8, 95%CI 0.2-2.9  - no significant association between busulfan exposure and DLCO abnormality: univariable OR 0.4, 95%CI 0.1-1.6 (no multivariable anaylsis performed because not significant!)  **BCNU or CCNU**:  - no significant association between BCNU or CCNU exposure and restrictive disease: univariable OR 1.1, 95%CI 0.3-4.2  - no significant association between BCNU or CCNU exposure and DLCO abnormality: univariable OR 1.4, 95%CI 0.6-4.7 (no multivariable anaylsis performed because not significant!)  **Smoking**  - no significant association between smoking history and restrictive disease: univariable OR 0.9, 95%CI 0.7-1.9  - no significant association between smoking history and DLCO abnormality: univariable OR 0.9, 95%CI 0.2-5.3 (no multivariable anaylsis performed because not significant!)  **Chest radiation**:  - significant association (multivariable) between increasing doses of chest radiation and restrictive disease:  - ≤20 Gy: OR 1.6 (95%CI 0.5-5.7), not sign.  - >20 Gy: OR 5.6 (95%CI 1.5-21.0), p<0.05  - significant association (multivariable) between increasing doses of chest radiation and DLCO abnormality:  - ≤20 Gy: OR 6.4 (95%CI 1.7-24.4), p<0.01  - >20 Gy: OR 11.3 (95%CI 2.6-49.5). p<0.01  **Longitudinal comparison t1 – t2 for DLco:**  - t1: 89 normal DLco patients  - t2: 23/89 (25.8%) abnormal DLco test  -> predictors for decline in DLco:  - ≤20 Gy: OR 6.4 (95%CI not stated), not sign.  - >20 Gy: OR 24.4 (95%CI 5.7-38.3), p<0.01 | **Analysis:**  - Cross-sectional and longitudinal analysis  - Univariable logistic regression  - Multivariable logistic regression, adjusted for race, health insurance status, smoking, heart failure  Limitations:  - Single center  - Data collection not clearly prospective/retrospective  - Selection bias – only survivors at follow-up at a tertiary center  - No lung function quality checks reported, no missing values reported  - Healthy control group not well characterized  - Time between t1 and t2 highly variable  Strength:  - Longitudinal PFT assessment  - PFT assessment blinded to exposure |

| **Main findings/message:**  **Hodgkin and NHL survivors treated with thoracic radiation and chemo have significantly lower FEV1 and FEV1/FVC compared to survivors treated with chemo-only.** | | | | |
| --- | --- | --- | --- | --- |
| **K. Nysom, et al.** Risk factors for reduced pulmonary function after malignant lymphoma in childhood. 1998;30:240-8. | | | | |
| **Study design**  **Treatment era**  **Years of follow-up** | **Participants** | **Treatment** | **Main outcomes** | **Additional remarks** |
| ☒ **Cohort**  ☐ **Cross-sectional**  ☐ **Case-control**  ☐ **Other:**  **__________**  ☒ **Retrospective**  ☐ **Prospective** | Study population (N)   - Original cohort: 118 - Eligible cohort: 63   Analysed cohort: 41 | ☐ 1 **HSCT**  ☐ **2 Cyclophosphamid**  ☐ **3 Methotrexate**  ☐ **4 Gemcitabine**  ☐ **5 Bleomycin**  ☐ **6 Busulfan**  ☐ **7 Lomustin (CCNU)**  ☐ **8 Carmustin (BCNU)**  ☒ **9 Radiotherapy lung:**  ☐ **10 Surgery**  ☐ **11 Combinations**  ☐ **12 Tobacco exposure** | ☐ **Pulmonary diseases**  ☒ **Pulmonary symptoms**  ☒ **Pulmonary function test**  ☐ **Absolute values**  ☒ **Z-scores**  ☐ **Percentage predicted**  **☒ Percentage pathological tests**  (e.g. 24% with reduced FEV1) | ☐ **Longitudinal data available**  **☐ Control group mentioned**  **☒ Reference values stated:**  Quanjer, Rosenthal  **☒ Quality check performed**  **☒ Lung function procedure stated** ERS  **☐ Cleaning of lung function data**  **described**  **☐ Person who analyzed PFT was**  **blinded to the exposure** |
| Centres:  Danish Cancer Registry  (Juliane Marie Centre, Rigshospitalet)  Country:  Denmark  Treatment era:  1970 to 1992 | Study population:  Patients diagnosed with HD & NHL < age 15yr  Reference values based on 348 healthy, non-smoking controls (age 13-24yr in a local population)   - 47 HD, 71 NHL (total 118)   Eligible & Analysis:  32 with HD, 31 with NHL;  **41** patients [22 with HD & 19 with NHL] were analysed  Inclusion criteria:  Alive and completed therapy; Informed consent  Cancer diagnosis:  Lymphoma (HD & NHL)  Age at diagnosis:  Median 11yr (3.9-15yr)  Age at follow-up:  Median 21.3yr (10.6-38.1yr)  Time since diagnosis:  Median 10.5yr (2.3- 23.7yr)  Smoking:  10 smokers, 4 ex-smokers | Name of protocol:  Various protocols (not all patients received all drugs in combinations)  Stratification in 2 exposure groups:   - **TI: Chemotherapy & thoracic RT** (can include RT to other sites) [21 patients]   - **noTI: Chemotherapy only** (included RT to other sites but not thoracic) [20 patients]  Chemotherapy (doses):   1. Bleomycin   Median 113mg/m^2^ (111-147) in Chemo only group;  Median 115mg/m^2^ (20-116) in Chemo & RT group   2. BCNU   Median 702mg/m^2^ (180-1064) in chemo only group;  Median 231mg/m^2^ (83-671) in chemo & RT group   3. CCNU   430 & 460mg/m^2^ in chemo only group;  Median 384mg/m^2^ (67-525) in chemo & RT group   4. Cyclophosphamide   7g/m^2^ in chemo only group;  7.2 & 7.8g/m^2^ in chemo & RT;   5. Doxorubicin   Median 421mg/m^2^ (113-528) in chemo only group;  Median 265mg/m^2^ (50-446) in chemo & RT group   6. Methotrexate (IV)   Median 24g/m^2^ (1-80) in chemo only group   7. Other drugs include:   Procarbazine, Dacarbazine, Mechlorethamine, Methotrexate intrathecal & oral  Radiotherapy (doses):   1. Mantle & thoracic   Median 37Gy (37-40)   2. Inverted Y/ Abdominal   Median 37Gy (20-40)   3. CNS   Median 24Gy (18-24) | How was outcome assessed?  Lung function (FEV1, FVC, TLC, DLCO) & heights were measured.  Results of lung function values were analysed as **standard residuals** (observed minus predicted values/residual standard deviation), **equivalent to** **SD (Z-scores)**. These were expressed as mean values with 95% CI & ranges, and compared with reference data and between treatment groups (chemotherapy & thoracic RT versus chemotherapy only).  Lung function results were considered **abnormal** if they were **>1.645 residual SD from predicted mean values**.  Information on respiratory symptoms, self-directed physical work capacity, & smoking were collected.  Main **descriptive results:**  **Comparing all patients with reference values:**  **- mean FEV1, FVC, TLC** significantly reduced when compared with reference values (-0.9 to -1.1 standard residual).  **- mean DLCO was significantly reduced** when compared with reference value (-1.3 standard residual)  **Comparing TI versus noTI:**  **- mean FEV1, FVC, FEV1/FVC, TLC** lower in TI versus noTI. FEV1 and FEV1/FVC significantly lower in TI versus noTI; FVC and TLC not significant.  Main results **multiple linear regression**:  - Lung volumes (FVC, FEV1, TLC) were significantly related to age at diagnosis when adjusted for treatment group and smoking status. | Analysis:  Student’s t-test, Chi-square & Mann-Whitney’s unpaired tests were used to evaluate any significant difference between:   - Patients and reference values; - TI/Chemo & RT group and noTI/chemo-only group; - Smokers and non-smokers (smaller N!)   Multiple linear regression models were used to evaluate possible predictive variables of lung function  Limitations and Potential bias/methodological problems:  Some differences in demographic data between chemo & RT and chemo only groups:   - More intrathoracic disease with HD (18 vs 4) in chemo & RT group - More smokers in chemo & RT group (9 vs 3) - Longer follow-up period from completion of therapy in chemo & RT group (11.3 vs 3yr) |

| **Main findings/message:**  **Lung function (FVC, DLCO, and DLCO/VA) decreased during first 6 months and improved thereafter. FVC and TLC were back to normal (>80% predicted) at 12 months, DLCO was normalized at 24 months, but DLCO/VA remained reduced. There is no sign. effect on lung function of cumualtive Bleomycine dose (DLCO, P=0.98; DLCO/VA, p=0.92), additional lung irradiation (bilateral full radiation, p>0.4) or smoking (p>0.25).** | | | | |
| --- | --- | --- | --- | --- |
| **N. M. Marina, et al.** Serial pulmonary function studies in children treated for newly diagnosed Hodgkin's disease with mantle radiotherapy plus cycles of cyclophosphamide, vincristine, and procarbazine alternating with cycles of doxorubicin, bleomycin, vinblastine, and dacarbazine. 1995;75:1706-11. | | | | |
| **Study design**  **Treatment era**  **Years of follow-up** | **Participants** | **Treatment** | **Main outcomes** | **Additional remarks** |
| ☒ **Cohort**  ☐ **Cross-sectional**  ☐ **Case-control**  ☐ **Other:**  **__________**  ☐ **Retrospective**  ☒ **Prospective** | **Study population (N)**   - Original cohort: 85 Hodgkin patients - Eligible cohort: 52 with mantle RT and COP/ABVD - Analyzed cohort: 37 | ☐ **1** **HSCT**  ☐ **2 Cyclophosphamid**  ☐ **3 Methotrexate**  ☐ **4 Gemcitabine**  ☒ **5 Bleomycin**  ☐ **6 Busulfan**  ☐ **7 Lomustin (CCNU)**  ☐ **8 Carmustin (BCNU)**  ☒ **9 Radiotherapy lung**  ☐ **10 Surgery**  ☐ **11 Combinations:**  ☒ **12 Tobacco exposure** | ☐ **Pulmonary diseases**  ☐ **Pulmonary symptoms**  ☒ **Pulmonary function test**  ☒ **Absolute values**  ☐ **Z-scores**  ☒ **Percentage predicted**  **☐ Percentage pathological tests**  (e.g. 24% with reduced FEV1) | ☒ **Longitudinal data available**  **☐ Control group mentioned**  **☐ Reference values stated**  **☐ Quality check performed**  **☐ Lung function procedure stated**  **☐ Cleaning of lung function data**  **described**  **☐ Person who analyzed PFT was**  **blinded to the exposure** |
| Centres:  Single-centre:  St. Jude’s Children’s Research Hospital, Departments of Pediatrics and Radiology University  Country:  USA  Treatment era:  1983-1988  Years of Follow-up:  Median (range) 93 (56-126) months  Pulmonary function follow-up:  Median 19 (3-79) months after end of therapy | Inclusion criteria:  Biopsy-proven Hodgkin’s disease  Pulmonary functions measured before, during and after treatment  Cancer diagnosis:  Hodgkin’s disease  Age at diagnosis:  Median 15 (range 6-20) years  Time since diagnosis  Lung functions from DX to 2 yrs after DX (all), up to 4 yrs after DX (some) | Name of protocol  COP/ABVD  Chemotherapy (doses)  **COP**: cyclophosphamide (200mg/m2 i.v. weeklyx4), vincristine (1.0mg/m2 i.v. weeklyx4, procarbazine (100mg/m2 orally daily for 2 weeks)  **ABVD** on days 1 and 14: doxorubicin (25mg/m2 i.v.), bleomycin (10mg/m2 i.v.), vinblastine (6.0mg/m2 i.v.), dacarbazine (250mg/m2 i.v.)  Radiotherapy (doses)  Absent pulmonary parenchymal disease: low dose mantle radiotherapy, when 18-20 Gy  Nodular parenchymal involvement: mantle radiotherapy plus 14- 16 Gy bilateral whole lung radiation  Surgery (kind of surgery)  NA | How was outcome assessed?  Medical history, physical examination, laboratory, diagnostic imaging, clinical staging (Ann Arbor), measurement of: DLCO, Spirometry, body plethysmography  **Assessed parameters:** FVC, TLC, diffusing capacity (DLCO), diffusing capacity per unit of alveolar volume (DLCO/VA).  All parameters presented as % predicted  **Time points of PFT:** before 1^st^ Bleomycin dose (Baseline lung function), after end of radiotherapy, after end of therapy, and in general also before each cycle of ABVD.  Average 7 PFT per patient, range 3 to 12  Prevalence  **FVC and TLC** decreased slightly at 1 yr post Dx (n.s.), back to baseline at 2 yrs post Dx.    **DLCO and DLCO/VA**: declined during first 6 months of therapy, gradual improvement over time. Both remained decreased at one year post-diagnosis, DLCO/VA remained also decreased at 2 yrs after Dx (= 1 yr after end of tt)  Risk factors  No sign. effect on lung function of:  - Cumualtive Bleomycine dose (but: bleomycin was omitted when DLCO/VA <50%): DLCO, P=0.98; DLCO/VA, p=0.92  - Additional lung irradiation (bilateral full radiation): p>0.4  - smoking: p>0.25 | Analysis:  Repeated-measures mixed-effects model  Limitations:  Small study population, 30% drop-out  PFT as % predicted  Follow-up for only 2 (to 4) years  Strength:  Carefully conducted and reported study  Longitudinal PFT  Baseline PFT before treatment  Homogeneous group (1 DX, 1 Treatment scheme)  Analysis fine (change from baseline)  Potential bias/methodological problems: |

| **Main findings/message:**  **Multiple thoracotomies in osteosarcoma patients predicted greater impairment of TLC. Prevalence of abnormal values not significantly different those exposed to Bleomycin or not.** | | | | |
| --- | --- | --- | --- | --- |
| **J. W. Denbo, et al.** Long-term pulmonary function after metastasectomy for childhood osteosarcoma: a report from the St Jude lifetime cohort study. 2014;219:265-71. 10.1016/j.jamcollsurg.2013.12.064 | | | | |
| **Study design**  **Treatment era**  **Years of follow-up** | **Participants** | **Treatment** | **Main outcomes** | **Additional remarks** |
| ☒ **Cohort**  ☐ **Cross-sectional**  ☐ **Case-control**  ☐ **Other:**  **__________**  ☐ **Retrospective**  ☒ **Prospective** | **Study population (N)**   - Original cohort: NA - Eligible cohort: 26 - Analyzed cohort: 21 | ☐ **1** **HSCT**  ☐ **2 Cyclophosphamid**  ☐ **3 Methotrexate**  ☐ **4 Gemcitabine**  ☒ **5 Bleomycin**  ☐ **6 Busulfan**  ☐ **7 Lomustin (CCNU)**  ☐ **8 Carmustin (BCNU)**  ☐ **9 Radiotherapy lung**  ☒ **10 Surgery**  ☐ **11 Combinations**  ☐ **12 Tobacco exposure** | ☐ **Pulmonary diseases**  ☐ **Pulmonary symptoms**  ☒ **Pulmonary function test**  ☐ **Absolute values**  ☐ **Z-scores**  ☒ **Percentage predicted**  **☐ Percentage pathological tests**  (e.g. 24% with reduced FEV1) | ☐ **Longitudinal data available**  **☐ Control group mentioned**  **☒ Reference values stated**  ATS guidelines  **☐ Quality check performed**  **☐ Lung function procedure stated**  **☐ Cleaning of lung function data**  **described**  **☐ Person who analyzed PFT was**  **blinded to the exposure** |
| Centres:  Single centre: SJLIFE  Country:  USA  Treatment era:  1968-1998  Years of Follow-up:  Mean 20 years (±9 yesr SD) | Study population:  Eligible (N): 26  Analysis (N): 21  Inclusion criteria: metastasectomy for osteosarcoma and available PFT results  Cancer diagnosis:  Osteosarcoma  Age at diagnosis:  Mean 13yeasr (± 5 years SD)  Age at follow-up:  Mean 35 years (±11 SD) | Chemotherapy (doses)  Bleomycin in n=6 (mean 107mg/m2; range 45-150mg/m2)  BCNU n=0  Radiotherapy (doses)  None  Surgery (kind of surgery)  Thoracotomy 100%  HSCT  None | How was outcome assessed?  Medical records, prospective measurements  Spirometry, body plethysmography, DLCO measurement  Abnormal PFT:  FVC <80%; FEV1 <80%; TLC <75%; DLCOcorr <75%  Incidence, Prevalence:  - Abnormal TLC: 29%  - Abnormal DLCOcorr: 47%  - Abnormal FVC 40%  - Abnormal FEV1 48%  - None with obstructive disease  - 29% (6/21) with restrictive disease  Risk factors (RR, OR…):  - After multiple thoracotomies higher prevalence of abnormal values for TLC, FVC, and FEV1 (only TLC statistically significant: p=0.031)  - Prevalence of abnormal values not significantly different when comparing ≤2 resected lesions vs >2 resected lesions  - Prevalence of abnormal values not significantly different those exposed to Bleomycin or not. | Analysis:  Fisher’s exact test  Limitations:  Small sample  Sample size too small for risk factor analysis  Strength:  Homogeneous cohort  No missing outcome data  Potential bias/methodological problems:  Confounding not assessed |
| **Main findings/message:**  **Long-term cancer survivors treated with potentially pulmonary-toxic therapy screened decades after treatment have a high prevalence of restrictive dysfunction (17.6%) and decreased DLCO (40%); Compared Bleomycin exposure, pulmonary radiotherapy and pulmonary surgery are all associated with pulm function impairment. Pulm RT, in combination with bleomycin or surgery is the most important risk factor.** | | | | |
| **R. L. Mulder, et al.** Pulmonary function impairment measured by pulmonary function tests in long-term survivors of childhood cancer. 2011;66:1065-71. 10.1136/thoraxjnl-2011-200618 | | | | |
| **Study design**  **Treatment era**  **Years of follow-up** | **Participants** | **Treatment** | **Main outcomes** | **Additional remarks** |
| ☒ **Cohort**  ☐ **Cross-sectional**  ☐ **Case-control**  ☐ **Other:**  **__________**  ☒ **Retrospective**  ☐ **Prospective** | **Study population (N)**   - Original cohort: 248 - Eligible cohort: 220 - Analyzed cohort: 193 | ☐ **1** **HSCT**  ☒ **2 Cyclophosphamid**  ☐ **3 Methotrexate**  ☐ **4 Gemcitabine**  ☒ **5 Bleomycin**  ☐ **6 Busulfan**  ☐ **7 Lomustin (CCNU)**  ☐ **8 Carmustin (BCNU)**  ☒ **9 Radiotherapy lung**  ☒ **10 Surgery**  ☒ **11 Combinations**  ☐ **12 Tobacco exposure** | ☐ **Pulmonary diseases**  ☐ **Pulmonary symptoms**  ☒ **Pulmonary function test**  ☐ **Absolute values**  ☐ **Z-scores**  ☐ **Percentage predicted**  **☒ Percentage pathological tests**  (e.g. 24% with reduced FEV1) | ☐ **Longitudinal data available**  **☐ Control group mentioned**  **☐ Reference values stated**  **☐ Quality check performed**  **☐ Lung function procedure stated**  **☐ Cleaning of lung function data**  **described**  **☐ Person who analyzed PFT was**  **blinded to the exposure** |
| Centres:  Single centre, Emma Children’s Hospital /academic medical center  Country:  Netherlands  Treatment era:  1966-1996  Years of Follow-up:  1996 to 2009  Median 17.9 years (range 5.6-36.8) | Inclusion criteria:  1) Diagnosed between 1966 and 1996  2) aged <18 years at dx  3) treated mainly at EKZ/AMC  4) survived ≥5 years after diagnosis at January 2007  5) Received pulmonary toxic therapy (Bleomycin, pulmonary radiation, pulmonary surgery)  6) ≥18 at PFT evaluation  Cancer diagnosis:  **All cancers** where pulmonary toxic therapy was used  Age at diagnosis:  Median 10 years (range 0-17)  Age at follow-up:  Median 27.3 years (range 18.2-47.0)  Time since diagnosis  Median 17.9 years (range 5.6-36.8) | Name of protocol: NA  Chemotherapy (doses)  **Bleomycin** (**57%**; dose: 60 [10-594/m2])  Not details on other chemotherapeutics  Radiotherapy(doses)  **Any (40.9%)**  Complete thorax (6.7%)  Part of thorax (13.5%)  Mediastinum (13.5%)  TBI (6.8%)  Surgery (kind of surgery)  **Any (16.6%)**  Metastectomy uni (4.7%)  Metastectomy bilat (5.7%)  Lobectomy (0)  Pneumonectomy (0)  Thoracic wall resection (2.6%)  Other (3.1%) | How was outcome assessed?  One-time assessment, at variable interval after Dx.  First “complete” pulmonary function test performed at least 5 yrs after Dx included.   - Diagnoses, graded per CTCAE and other standardized definitions - Pulmonary function test: - Spirometry: FVC, FEV1, FEV1% - Unclear how TLC was measured. (Plethysmography?) - DLCO and DLCO/AV   Prevalence at time of investigation (5 to 37 years after DX)  Overall,   - 21% with FEV1<80% - 3.1% with FEV1/FEV <70% - 2.1% with both parameters pathological (=obstructive) - 17.6% with TLC or FVC<75% predicted (=restrictive) - 39.9% with DLCO<75% pred - 4.3% with DLCO/AV <75% - **Total** **44% had ≥Grade 2 pulmonary impairment** - 73% had one or more mild pulmonary function impairments (grade 1) - 14.5% had both restrictive lung function and decreased DLCO   Risk factors (OR):  **Restrictive disease (≥Grade 2)**  Model 1:  Pulmonary radiotherapy (OR 12.87; 3.37-49.08); vs. no RT  Surgery yes vs. no (OR 3.79; 1.25-5.79)  High-dose cyclophosphamide and bleomycin not associated with restrictive disease  Multivariable Model 2 (ref: bleomycin alone):  Radiotherapy only (OR 6.99; 2.27-21.54)  Bleomycin + RT (OR 9.42; 1.71-51.86)  RT + surgery (OR 33.44; 7.81-143.09)  Surgery only not associated  **DLCO impairment** **(≥Grade 2)**  Model 1:  Pulmonary radiotherapy (OR 5.84; 1.88-18.14); vs. no RT  High-dose cyclophosphamide, bleomycin, and surgery not associated with DLCO impairment  Multivariable Model 2 (ref: bleomycin alone):  Radiotherapy only (OR 2.85; 1.32-6.19)  Bleomycin + RT (OR 6.17; 1.37-27.84)  RT + surgery (OR 5.98; 1.64-21.81)  Surgery only and bleomycin with surgery not associated with DLCO impairment  If longitudinal data available:  N/A  **Exact results available from publication, Table 3**: “Risk factors for pulmonary function impairment (grade 2 or higher) | Analysis:  Cross-sectional design  Multivariable analysis  Limitations:   - Retrospective study - No description of lung function testing and validation, likely heterogeneous data quality; no quality control described. - No description of normal values used for lung function tests - Lung RT info not dose-specific (any). - Variable length of Follow-up (5 - 37 years) - No original lung function data described, only the proportion with pathological results based on definitions.   Strength:   - good participation rate (>85%) - Long-term outcomes, but at very variable distance from Dx (5 to 37 years) - clearly defined severity grading   Potential bias/methodological problems: - retrospective study  - Limited informaiton on lung function  testing  - Limited information on chemotherapy and  radiation dosimetry  - No information on smoking and lifestyle  - Limited to survivors with known  pulmonary toxic agents |

| **Main findings/message:**  **Median 23 years after treatment for ALL with chemotherapy only, mean pulmonary function is within the lower predicted range for the whole group. Impaired DLCO (<80% predicted) was found in 22%. Smoking is a risk factor for impaired DLCO. No association found between cumulative dose of methotrexate and cyclophosphamide and impaired DLCO (only in text, no data shown).** | | | | |
| --- | --- | --- | --- | --- |
| **O. Myrdal, et al.** Risk factor for impaired pulmonary function and cardiorespiratory fitness in very long-term adult survivors of childhood acute lymphoblastic leukemia after treatment with chemotherapy only. (2018). Acta Oncologica, 57:5, 658-664 | | | | |
| **Study design**  **Treatment era**  **Years of follow-up** | **Participants** | **Treatment** | **Main outcomes** | **Additional remarks** |
| ☐ **Cohort**  ☒ **Cross-sectional**  ☐ **Case-control**  ☐ **Other:**  **__________**  ☐ **Retrospective**  ☒ **Prospective** | **Study population survivors (N)**   - Original cohort: NA - Eligible cohort: 210 - Analyzed cohort: 116 | ☐ **1** **HSCT**  ☐ **2 Cyclophosphamid**  ☐ **3 Methotrexate**  ☐ **4 Gemcitabine**  ☐ **5 Bleomycin**  ☐ **6 Busulfan**  ☐ **7 Lomustin (CCNU)**  ☐ **8 Carmustin (BCNU)**  ☐ **9 Radiotherapy lung: a**  ☐ **10 Surgery**  ☐ **11 Combinations**  ☒ **12 Tobacco exposure** | ☐ **Pulmonary diseases**  ☐ **Pulmonary symptoms**  ☒ **Pulmonary function test**  ☒ **Absolute values**  ☐ **Z-scores**  ☒ **Percentage predicted**  **☐ Percentage pathological tests**  (e.g. 24% with reduced FEV1) | ☐ **Longitudinal data available**  **☐ Control group mentioned**  **☒ Reference values stated:** Pellegrino  **☐ Quality check performed**  **☒ Lung function procedure stated**  According to ERS  **☐ Cleaning of lung function data**  **described**  **☐ Person who analyzed PFT was**  **blinded to the exposure** |
| Design  Prospective cross-sectional  Centres:  Oslo University Hospital  Country:  Norway  Treatment era:  1970 - 2002  Years of Follow-up from diagnosis:  Median 23.2 years  Range 7.4 – 40.0 years | Study population:  Survivors of acute lymphoblastic leukemia (ALL) treated with chemotherapy only  Inclusion criteria:  Diagnosed before age 16 years with ALL, treated with chemotherapy only (no CSI, no BMT), diagnosed 1970 to 2002, age >18 years and alive in 2009  Cancer diagnoses:  Acute lymphoblastic leukemia 100%  Age at diagnosis:  Median 5.4 years  Range 0.3 – 16 years  Age at follow-up:  Median 28.5 years  Range 18.6 – 46.5 years  Time since treatment:  Median 23.2 years  Range 7.4 – 40.0 years | Name of protocol  NA, different protocols  Chemotherapy (dose) median (range):  95% Methotrexate: 21g/m^2^ (1-64)  77% Anthracyclines: 120mg/m^2^ (40 – 510)  33% Cyclophosphamid: 3g/m^2^ (0.3 – 10)  Radiotherapy (dose):  No radiotherapy  Surgery  NA  Smoking: 19% | How was outcome assessed?  - Spirometry: FVC, FEV1, FE1/FVC  - Lung volumes: TLC, RV  - Gas diffusion capacity: DLCO, DLCO/VA  - PFT results as absolute values and percentage of predicted normal values  Prevalence:  - Mean value for all lung function variables >80% predicted  - 3% with restrictive impairment  - 6% with obstructive impairment  - 22% impaired DLCO  Risk factor analysis:  - No significant correlation between DLCO% predicted and cumulative dose of methotrexate or cyclophosphamide (only in text, no data shown)  - Multiple linear regression analysis: smoking associated with reduced DLCO% predicted: β -9.8; 95%CI -16.0, -3.6; p-value 0.002 | Analysis:  - Students t-test or Mann-Whitney-U test: comparison of group mean  - Chi-squared: comparison of categorical data  - Multiple linear regression analysis: to detect associations between pulmonary function and explanatory variables  - PFT according to ERS guidelines  - All measurements on same machine  - Reference values for PFT: Pellegrino et al  - Obstructive= FEV1/FVC <0.7 (GOLD criteria)  - Restrictive and DLCO impairment= <80% predicted (corresponds to lower 5^th^ percentiles acc. to Pellegrino et al)  Limitations:  - No control group  Strength:  - long follow-up period  - homogeneous population  - accurate treatment data  - all PFT performed with the same criteria  - high response rate  Potential bias/methodological problems:  - PFT values compared to normal values from 2005 |

| **Main findings/message:**  **Treatment of childhood ALL causes mild pulmonary toxicity on the long term (61% normal lung function pattern). Age at treatment and intensity of treatment protocols are risk factors for reduced total lung capacity (TLC). Higher cumulative doses of cyclophosphamide are related with changes in TLC (simple regression model: R^2^ =0.04, p=0.07; multiple regression model: R^2^ =0.1, p=0.02). Change in TLC is not associated with the number of high-dose methotrexate cycles (simple regression model: R^2^ =0.00, p=0.9). No increased pulmonary toxicity of tobacco smoking in survivors of childhood ALL compared with background population (simple regression model: R^2^ =0.02, p=0.2); CAVE: small sample size and mild tobacco exposure.** | | | | |
| --- | --- | --- | --- | --- |
| **6402. K. Nysom, et al.** Pulmonary function after treatment for acute lymphoblastic leukaemia in childhood. 1998;78:21-7. | | | | |
| **Study design**  **Treatment era**  **Years of follow-up** | **Participants** | **Treatment** | **Main outcomes** | **Additional remarks** |
| ☒ **Cohort**  ☐ **Cross-sectional**  ☐ **Case-control**  ☐ **Other:**  **__________**  ☒ **Retrospective**  ☐ **Prospective** | **Study population (N)**   - Original cohort: 304 - Eligible cohort: 162 - Analyzed cohort: 94 | ☐ **1** **HSCT**  ☒ **2 Cyclophosphamid**  ☒ **3 Methotrexate**  ☐ **4 Gemcitabine**  ☐ **5 Bleomycin**  ☐ **6 Busulfan**  ☐ **7 Lomustin (CCNU)**  ☐ **8 Carmustin (BCNU)**  ☐ **9 Radiotherapy lung**  ☐ **10 Surgery**  ☐ **11 Combinations**  ☒ **12 Tobacco exposure** | ☐ **Pulmonary diseases**  ☒ **Pulmonary symptoms**  ☒ **Pulmonary function test**  ☐ **Absolute values**  ☒ **Z-scores**  ☐ **Percentage predicted**  **☒ Percentage pathological tests**  (e.g. 24% with reduced FEV1) | ☐ **Longitudinal data available**  **☐ Control group mentioned**  **☒ Reference values stated**  reference values form own  laboratory by adjusting published  reference values  **☐ Quality check performed**  **☒ Lung function procedure stated:** ERS  **☐ Cleaning of lung function data**  **described**  **☒ Person who analyzed PFT was**  **blinded to the exposure** |
| Design:  Cross-sectional, retrospective  Centres:  Multicentre:  Data from population-based Danish Cancer Registry  Lung function test: Rigis hospital  Country:  Denmark  Treatment era:  1970-1990  Years of Follow-up from diagnosis:  Median: 10.6 years Range: 3.4-23.4 years | Study population:  After acute lymphoblastic leukemia  Inclusion criteria:  Diagnosis of ALL, alive, in first remission, treatment finished, not treated with HSCT  Cancer diagnosis:  Acute lymphoblastic leukemia  Age at diagnosis:  Median: 3.9 years  Range: 0.5-14.8 years  Age at follow-up:  Median: 16.2 years  Range: 5.3-34.2 years  Time since diagnosis  Median: 10.6 years  Range: 3.4-23.4 years | Name of protocol  Several NOPHO protocols  Chemotherapy (doses)  AraC, CYC, DNR, DOX, L-ASP, MTX, PRED, VM26, 6TG, VCR  CYC: 600 – 6700mg/m2  Radiotherapy (doses):  Cranial irradiation 15-18 Gy or 24 Gy in 39 children | How was outcome assessed?  Pulmonary function testing:  - pneumotachograph: FEV1, FVC, flow-volume curves  - Helium dilution technique: TLC  - Single breath technique: transfer factor for carbon monoxide  - comparison of values with reference values from laboratory, generated by adjusting published reference values (Quanjer and Tammeling et al)  - data as standardized residuals; abnormal if >1.645 residual standard deviation from the predicted mean value  PFT:  - For every parameter at least one patient showed significantly reduced results (standardized residuals <1.645): see “additional data” Table 2  Aggregated data:  - 26% (25/94) restrictive pattern  - 11% (10/94) reduced TLCO  - 2% (2/94) obstructive pattern  - 61% normal pattern  Risk factors (RR, OR…):  Simple regression model  - Cyclophosphamid: p=0.07  - High-dose MTX: p=0.9  - Smoking: p=0.2  - Younger age at treatment: p=0.045  - Younger age at follow-up: p=0.01  - Cranial irradiation: p=0.04  Multiple regression model  - Cyclophosphamid: p=0.02  **Exact results available from publication, Table 2**: “Pulmonary function test results” and **Table 3** “Regression models for total tlung capacity” | Analysis:  PFT results analyzed as standardized residuals (Z-scores)  Pearson, spearman  Simple and multiple linear regression models (step-down procedure)  Chi-square test and Mann-Whitney test to compare baseline characteristics  Limitations:  Strongly correlated risk factors, for which they applied a complex statistical work around  Small sample size of smokers  Assessed only change in TLC in simple and multiple regression  Strength:  Large and homogeneous cohort  All tests performed in same laboratory  Evaluation of PFT result without knowledge of treatment protocol  Potential bias/methodological problems:  Selection in study: 25% of eligible patients declined to participate; their characteristics are not described  Long period of time and therefore many changes in treatment protocols |

| **Main findings/message:**  **Significant PFT deficits in this population. Lower doses of radiotherapy (<23,45Gy) had larger DLCOcorr% predicted than those with higher dose (p=0.032) (univariable analysis). Cyclophosphamid dose is not significantly associated with change in FEV1% predicted (univariable analysis).** | | | | |
| --- | --- | --- | --- | --- |
| **D. M. Green, et al.** Pulmonary Function After Treatment for Embryonal Brain Tumors on SJMB03 That Included Craniospinal Irradiation. 2015;93:47-53. 10.1016/j.ijrobp.2015.05.019 | | | | |
| **Study design**  **Treatment era**  **Years of follow-up** | **Participants** | **Treatment** | **Main outcomes** | **Additional remarks** |
| ☒ **Cohort**  ☐ **Cross-sectional**  ☐ **Case-control**  ☐ **Other:**  **__________**  ☐ **Retrospective**  ☒ **Prospective** | **Study population (N)**   - Original cohort: 305 - Eligible cohort: 303 - Analyzed cohort: 260 | ☐ **1** **HSCT**  ☒ **2 Cyclophosphamid**  ☐ **3 Methotrexate**  ☐ **4 Gemcitabine**  ☐ **5 Bleomycin**  ☐ **6 Busulfan**  ☐ **7 Lomustin (CCNU)**  ☐ **8 Carmustin (BCNU)**  ☒ **9 Radiotherapy lung:**  ☐ **10 Surgery**  ☐ **11 Combinations**  ☐ **12 Tobacco exposure** | ☐ **Pulmonary diseases**  ☐ **Pulmonary symptoms**  ☒ **Pulmonary function test**  ☐ **Absolute values**  ☐ **Z-scores**  ☒ **Percentage predicted**  **☐ Percentage pathological tests**  (e.g. 24% with reduced FEV1) | ☒ **Longitudinal data available**  **☐ Control group mentioned**  **☒ Reference values stated**  10 different references for  standardization  **☐ Quality check performed**  **☒ Lung function procedure stated:** ATS  **☐ Cleaning of lung function data**  **described**  **☐ Person who analyzed PFT was**  **blinded to the exposure** |
| Desgin  Prospective cohort  Centres:  Multicentric  Country:  USA, Canada, Australia  Treatment era:  June 2003 - March 2010  Years of Follow-up from diagnosis:  minimum 2 years | Study population:  Embryonal brain tumors  Inclusion criteria:  Patients 3-21 years with embryonal brain tumors, treated on SJMB03 including CSI, minimum follow-up of 24 month, PFT data available  Cancer diagnosis patients with PFT yes:  Medulloblastoma 80%  PNET 8%  ATRT 7%  Pineoblastoma 5%  Medullomyoblastoma n=1  Age at diagnosis:  Median 8,9 years  Range 3,1-20,4 years  Age at follow-up: NA  Time since treatment:  minimum 2 years | Name of protocol  SJMB03 protocol  Chemotherapy (dose):  High-dose chemotherapy: CYC, Cisplatin, VCR and peripheral blood stem cell support  Median cumulative dose of CYC: 16,0 g/m2 (IQR: 15,7-16)  Radiotherapy (dose):  Median dose spinal radiation: 23,4 Gy IQR (23,4-36)  Spinal dose ≤2345 cGy: 66,3 %  Spinal dose >2345 cGy: 33,6%  Proton beam: 0,07% | How was outcome assessed?  Pulmonary function test after CSI, before each course of high-dose chemotherapy and 24 and 60 months after the completions of chemotherapy  - PFT predominantly in children 6 years and older  - Spirometry: FVC, FEV1  - Nitrogen washout method and body plethysmography: TLC  - Single breath method: DLCO  - values standardized to % predicted  - abnormal values if: FEV1<80% pred., FVC <80% pred., DLCO<75% pred, TLC<75% pred  Incidence, Prevalence:  - DLCO corr < 75% predicted: 23% and 25%  - FEV1 <80%: 20% and 29%  - FVC <80%: 27% and 28%  - TLC < 75%: 9%and 11%  Risk factor analysis:  - Higher **DLCO% predicted** in male (p<0.001), younger age at diagnosis (p=0.007), and treated with photon CSI (p=0.006)  - Time point is significant predictor of DLCO% predicted (p>0.001)  - Treatment with lower RT doses (≤2345 cGy) had larger DLCO (p=0.032)  - Significant predictors of higher **FEV1% predicted**: male sex (p=0.025) and time from diagnosis (p<0.001).  - Significant predictors of larger **TLC% predicted**: decreased time from diagnosis (p>0.001), male sex (p=0.009), white, non Hispanic race group (p=0.003), photon beam (p=0.002) and younger age (p=0.003)  - Significant predictors of larger **FVC% predicted**: male sex (p=0.003) and shorter elapsed time from diagnosis (p<0.001)  - No analysis of cyclophosphamide possible because all received CYC | Analysis:  - Associations between categorical variables: Fisher exact test and X^2^  - Differences between different time points: exact Wilcoxon signed-rank test  - Repeated measures models were used to examine predictors of pulmonary outcomes  Limitations:  No PFT in young children  No evaluation of scoliosis  No baseline PFT/before CSI  Many different references to standardize PFT results  Strength:  Large cohort with prospective and longitudinal data  Homogenous cohort, one treatment protocol only  Potential bias/methodological problems:  Selection bias: PFTs more likely to be performed in those >5 years of age and with M0 stage disease  Unclear how T1 and T6 are compared because number of eligible decreases from 295 to 214 |

ATRT=atypical teratoid rhabdoid tumor; CSI=craniospinal irradiation; PNET=primitive neuroectodermal tumor

| **Main findings/message:**  **In a cohort of 10-year survivors of childhood cancer, current and former smokers had lower FEV1/ FVC and DLCOcorr compared to non-smoker.** | | | | |
| --- | --- | --- | --- | --- |
| **S. C. Oancea, et al.** Cigarette smoking and pulmonary function in adult survivors of childhood cancer exposed to pulmonary-toxic therapy: results from the St. Jude lifetime cohort study.  2014;23:1938-43. 10.1158/1055-9965.epi-14-0266 | | | | |
| **Study design**  **Treatment era**  **Years of follow-up** | **Participants** | **Treatment** | **Main outcomes** | **Additional remarks** |
| ☒ **Cohort**  ☐ **Cross-sectional**  ☐ **Case-control**  ☐ **Other:**  **__________**  ☒ **Retrospective**  ☐ **Prospective** | Study population (N)   - Original cohort: NA - Eligible cohort: 433 - Analyzed cohort: 433 | ☐ 1 **HSCT**  ☐ **2 Cyclophosphamid**  ☐ **3 Methotrexate**  ☐ **4 Gemcitabine**  ☐ **5 Bleomycin**  ☐ **6 Busulfan**  ☐ **7 Lomustin (CCNU)**  ☐ **8 Carmustin (BCNU)**  ☐ **9 Radiotherapy lung**  ☐ **10 Surgery**  ☐ **11 Combinations**  ☒ **12 Tobacco exposure** | ☐ **Pulmonary diseases**  ☐ **Pulmonary symptoms**  ☒ **Pulmonary function test**  ☐ **Absolute values**  ☐ **Z-scores**  ☒ **Percentage predicted**  **☐ Percentage pathological tests**  (e.g. 24% with reduced FEV1 | ☐ **Longitudinal data available**  **☐ Control group mentioned**  **☐ Reference values stated**  **☐ Quality check performed**  **☐ Lung function procedure stated**  **☐ Cleaning of lung function data**  **described**  **☐ Person who analyzed PFT was**  **blinded to the exposure** |
| Centres:  Single, St. Jude (SJLIFE)  Country:  US  Treatment era: unspecified  Years of Follow-up:  Not specified | Inclusion criteria:  >10 years from diagnosis, >18 years of age at assessment; risk-based assessment of pulmonary function acc. to COG guidelines  Cancer diagnosis: not specified  Age at diagnosis:  Median (range) yr  0-4: n= 60  5-9: n=88  10-14: n=139  15-22: n=146  Age at follow-up:  Median (range) yr  35 (IQR 30-41)  Time since diagnosis  Median (range) yr 23 years IQR 18-29 years | Name of protocol  not specified  Chemotherapy (doses)  bleomycin, busulfan, lomustine (BCNU), carmustine (CCNU), doses not provided  Radiotherapy(doses):  radiotherapy to the chest (thorax), whole lung, mediastinum, axilla, mini-mantle, mantle, extended mantle, total lymphoid irradiation, subtotal lymphoid irradiation, or total body irradiation, doses not provided  Surgery (kind of surgery)  pulmonary lobectomy, pulmonary metastasectomy or pulmonary wedge resection,  HSCT (allo/auto)  none | How was outcome assessed?  - Single breath diffusion capacity for carbon monoxide corrected for haemoglobin (DLCOcorr)  - Spirometry: FEV1, FVC  - Body plethysmography: TLC  Definitions  - Obstructive: FEV1/FVC < 0.70; acc. to GOLD criteria  - Restrictive: TLC < 75% predicted; acc. to Guide to the Evaluation of Permanent Impairment  - Smoker: smoked >100 cigarettes in their life  Incidence, Prevalence:  - 1 former smoker, 1 never smoker met criteria for obstructive lung disease (FEV1/ FVC <0.7)  - Restrictive lung disease (TLC <75%) in 25.3% of current, 30.4% in former, 34.6% in never smokers.  - Median TLC, FEV1, and FVC values not significantly different between never, former, and current smoker and between those who ever smoked more of less than 6 pack years (py)  - Median DLCOcorr only significantly only different between never and current smoker (p=0.02) and those who ever smoked >6 py (p=0.03)  - FEV1/FVC median values among current (p=0.03) and former smoker (p=0.01) significantly lower compared to median values of never smoker  If longitudinal data available:  None  **Exact results available from publication, Table 2**: “Pulmonary function among adult survivors of childhood cancer” | Analysis:  - Kruskal wallis test  - Dwass, Steel, Critchlow-Flinger multiple comparison procedure (DSCF) for pairwise comparisons between the groups  - Chisq tests or Exact chisq tests for associations for categorical variables  Limitations:  Smoking assessed by self-report, no control group (used normative values)  Strength:  Large number, treatment exposure (although doses not provided)  PFT performed according to ATS standards  Potential bias/methodological problems:  Critical risk of bias in classification of intervention due to self-reported outcome |

| **Main findings/message: Comparing lung function parameters of leukemia survivors with age- and sex-matched controls a median of 4 years since diagnosis, FEV1, FVC, FRC, TLC, DLCO and VA are significantly lower in survivors than in controls. Risk factors for reduced parameters is cyclophosphamide.** | | | | |
| --- | --- | --- | --- | --- |
| **6698. M. E. Jenney, et al.** Lung function and exercise capacity in survivors of childhood leukaemia. 1995;24:222-30. | | | | |
| **Study design**  **Treatment era**  **Years of follow-up** | **Participants** | **Treatment** | **Main outcomes** | **Additional remarks** |
| ☐ **Cohort**  ☒ **Cross-sectional**  ☒ **Case-control**  ☐ **Other:**  **__________**  ☐ **Retrospective**  ☒ **Prospective** | Study population (N)   - Original cohort: NA - Eligible cohort: 178 - Analyzed cohort: 70   69 with PFT | ☐ 1 **HSCT**  ☒ **2 Cyclophosphamid**  ☐ **3 Methotrexate**  ☐ **4 Gemcitabine**  ☐ **5 Bleomycin**  ☐ **6 Busulfan**  ☐ **7 Lomustin (CCNU)**  ☐ **8 Carmustin (BCNU)**  ☒ **9 Radiotherapy lung**  ☐ **10 Surgery**  ☐ **11 Combinations**  ☐ **12 Tobacco exposure** | ☐ **Pulmonary diseases**  ☒ **Pulmonary symptoms**  ☒ **Pulmonary function test**  ☐ **Absolute values**  ☐ **Z-scores**  ☒ **Percentage predicted**  **☐ Percentage pathological tests**  (e.g. 24% with reduced FEV1) | ☐ **Longitudinal data available**  **☒ Control group mentioned**  **☐ Reference values stated**  **☐ Quality check performed**  **☐ Lung function procedure stated**  **☐ Cleaning of lung function data**  **described**  **☐ Person who analyzed PFT was**  **blinded to the exposure** |
| Centres:  Multi-Centre  Country:  UK  Treatment era:  1954-1988  Years of Follow-up:  Median (range) yr  Follow-up: August 1992 | Inclusion criteria:  Children with ALL or ANLL, diagnosed in the North West Region since 1953, completed treatment > 6 months without relapse, and age at study 6-30 years  Cancer diagnosis:  Acute lymphoblastic and non-lymphoblastic leukaemia (ALL, ANLL)  Age at diagnosis:  Median 5.8 (range 1.6-14.9) years  Age at follow-up:  Median 14.6 (range 13.3 – 15.9) years  Time since diagnosis  Median 4.2 (range 0.60 – 18.5) years | Name of protocol  ALL: MRC UKALL protocol, prior to 1980: UKALL II, III, V, VII; after 1980: UKALL 8 and UKALL 10  ANLL: UKALL AML 9 or 10 trials  Allogenic and autologous bone marrow transplantation was introduced for treatment of higher risk ALL and ANLL  Chemotherapy (doses)  Not specified  Radiotherapy(doses)  Cranial RT: n=45; dose 1800-2400cGy  Craniospinal RT: n=10; dose 1200-2400cGy  TBI: n=14; dose 1100-1440cGy  Surgery (kind of surgery): NA  HSCT: unclear if allogeneic or autologous HSCT, unclear number of patients with HSCT 🡪 not included as PICO | How was outcome assessed?  Medical record: age at diagnosis, age at completion of therapy, details of cytotoxic chemotherapy and radiotherapy received, incidence of lower respiratory tract infections requiring hospitalisation  Examination: cardiorespiratory system, resting BP and HR, height, weight and arm span  Pulmonary function tests: FEV1, FVC, RV, FRC, ITGV, RAW, SGAW, TLC, DLCO  Risk factor analysis  - Cyclophposphamid leads to reduction in FEV1, FVC, TLC: p<0.001  - Craniospinal irradiation leads in reduction in FEV1, FVC, TLC: p<0.001 and DLCO=0.03  **Exact results available from publication, Table Va**: “Lung Function Results of Survivors of Leukemia” and **Table VI** “Independent Variables Which Led to a Reducation in Indices of Lung Function” | Analysis:  - Multiple regression analysis on lung function data from control group to obtain predictive equations  - Comparison survivors vs controls by student’s unpaired t-tests  - Interrelationships between variables examined by Pearson correlation coefficient and stepwise multiple regression  Statistical significance was set at 5%  146 age- and sex-matched controls; close friend or siblings  Limitations:  No longitudinal data  No pretreatment Pulmonary function  Strength:  Control group  Potential bias/methodological problems:  Table VI (“Independent variables which lead to a reduction in indices of lung function”) shows significant results only |

| **Main findings/message:**  **Lung function test results of survivors treated with bleomycin after a median of 2.3 years after treatment, bleomycin dose is associated with abnormal spirometry and smoking is associated with reduced DLCO.** | | | | |
| --- | --- | --- | --- | --- |
| **A. P. Zorzi, et al.** Bleomycin-associated Lung Toxicity in Childhood Cancer Survivors. 2015;37:e447-52. 10.1097/mph.0000000000000424 | | | | |
| **Study design**  **Treatment era**  **Years of follow-up** | **Participants** | **Treatment** | **Main outcomes** | **Additional remarks** |
| ☐ **Cohort**  ☒ **Cross-sectional**  ☐ **Case-control**  ☐ **Other:**  **__________**  ☒ **Retrospective**  ☐ **Prospective** | Study population (N)   - Original cohort: NA - Eligible cohort: 195 - Analyzed cohort 143 | ☐ 1 **HSCT**  ☐ **2 Cyclophosphamid**  ☐ **3 Methotrexate**  ☐ **4 Gemcitabine**  ☒ **5 Bleomycin**  ☐ **6 Busulfan**  ☐ **7 Lomustin (CCNU)**  ☐ **8 Carmustin (BCNU)**  ☐ **9 Radiotherapy lung**  ☐ **10 Surgery**  ☐ **11 Combinations**  ☐ **12 Tobacco exposure** | ☐ **Pulmonary diseases**  ☒ **Pulmonary symptoms**  ☒ **Pulmonary function test**  ☐ **Absolute values**  ☐ **Z-scores**  ☒ **Percentage predicted**  **☐ Percentage pathological tests**  (e.g. 24% with reduced FEV1) | ☐ **Longitudinal data available**  **☐ Control group mentioned**  **☒ Reference values stated**  Stanojevic, Weng and Levison, reference equations from Sick Children, Pellegrino et al  **☐ Quality check performed**  **☒ Lung function procedure stated:** ATS  **☐ Cleaning of lung function data**  **described**  **☐ Person who analyzed PFT was**  **blinded to the exposure** |
| Centres:  Single center, Hospital for Sick Children, Toronto  Country:  Canada  Treatment era:  1997-2010  Years of Follow-up:  Median (range) yr  4,4 y (2,-7,4) | Inclusion criteria:  Bleomycin containing regimen  Cancer diagnosis:  Hodgkin’s disease 86%  Extracranial germ cell tumor: 14%  Age at diagnosis:  Median (range) yr  All < 18 years  63% aged between 11-15 years  Age at follow-up:  Median (range) yr  4,4 y (2,2-7,4)  Time since treatment  PFT median 2,3 years (1,4-4,9) after treatment | Name of protocol  Median cumulative bleomycin dose= 60 U/m2 (20-180)  MOPP/ABV 18%  COPP/ABV 17%  COPE/ABV 14%  ABVD 1%  AHOD0031 28%  CCG59704 7%  AHOD0831 1%  PEB 14%  Radiotehrapy  Chest radiation 60%  Dose median (range): 2100 cGy (1500-3000) | How was outcome assessed?  - Medical records  - Most recent post-treatment PFT (spirometry, body plethysmography, DLCO)  - PFT performed according to ATS criteria  - References: Stanojevic (spirometry); Weng and Levison (body plethysmography); unpublished reference equations from data collected at Sick Children (DLCO)  Outcome definitions:  Percent predicted for abnormal   - TLC <80% - FVC <80% - DLCO <80%   - Obstructive if: abnormal FVC, normal TLC and RV/TLC >=30% and scooped flow volume loop.  - Restrictive: acc. to Pellegrino et al  Prevalence:  - Abnormal spirometry: 41% (n=58)  - obstructive: 70% (n=42)  - restrictive: 18% (n=11)  - mixed: 9% (n=5)  - Abnormal DLCO: 19% (n=27)  Risk factors  No association between ventilator defects (obstr., restr., or mixed) and smoking (p=0.8) or lung radiation (p=0.13)  OR for developing abnormal spirometry for each 1 U/m2 increase of bleomycin was 1.01 (95%CI 1.00-1.02)  Association between being a smoker and abnormal DLCO (p=0.04). Cumulative bleomycin dose (p=0.07) and radiation (p=0.83) were not associated with reduced DLCO | Analysis:  Chi-squared, Fishers exact test  Univariate analysis  No multivariable model due to low event rates  Limitations:  Retrospective study  PFT was performed with a median of 2,3 years (1,4-4,9)  Strength:  Homogeneous cohort  Well defined PFTs  Potential bias/methodological problems:  Serious risk of confounding due to unadjusted analysis |

| **Main findings/message:**  **With longer follow-up time, the proportion of survivors after HSCT having a lung disease increases. Risk factors for restrictive lung disease in multivariate analysis were single fraction TBI (OR 22, 95%-CI 3.9-120), fractionated TBI 1.2Gy (OR 2.5, 95%-CI 0.4-16), fractionated TBI 2.0-2.25Gy (OR 2.8, 95%-CI 0.6-13) with no-TBI as reference. Risk factors for obstructive disease in univariate analysis were prior cyclophosphamide (p=0.05) and chronic GvHD (p=0.02). Risk factors for obstructive disease in multivariate analysis were cGvHD (OR 4.4, 95%-CI 1.6-12); SFTBI (OR 0.1, 95%-CI 0.5-0.5) seemed negatively associated, FTBI 1.2Gy (OR 0.1, 95%-CI 0.0-1.4) and FTBI 2.0-2.25Gy (OR 0.9, 95%-CI 0.3-2.8) were not associated.** | | | | |
| --- | --- | --- | --- | --- |
| **P. A. Hoffmeister, et al.** Pulmonary function in long-term survivors of pediatric hematopoietic cell transplantation. 2006;47:594-606. | | | | |
| **Study design**  **Treatment era**  **Years of follow-up** | **Participants** | **Treatment** | **Main outcomes** | **Additional remarks** |
| ☐ **Cohort**  ☒ **Cross-sectional**  ☐ **Case-control**  ☐ **Other:**  **__________**  ☒ **Retrospective**  ☐ **Prospective** | Study population (N)   - Original cohort: 472 - Eligible cohort: 260 - Analyzed cohort: 215 | ☒ 1 **HSCT b, d**  ☒ **2 Cyclophosphamid**  ☐ **3 Methotrexate**  ☐ **4 Gemcitabine**  ☐ **5 Bleomycin**  ☐ **6 Busulfan**  ☐ **7 Lomustin (CCNU)**  ☐ **8 Carmustin (BCNU)**  ☐ **9 Radiotherapy lung a**  ☐ **10 Surgery**  ☐ **11 Combinations**  ☐ **12 Tobacco exposure** | ☐ **Pulmonary diseases**  ☒ **Pulmonary symptoms**  ☒ **Pulmonary function test**  ☐ **Absolute values**  ☐ **Z-scores**  ☐ **Percentage predicted**  **☒ Percentage pathological tests**  (e.g. 24% with reduced FEV1) | ☐ **Longitudinal data available**  **☐ Control group mentioned**  **☒ Reference values stated**  reference equations of Rosenthal  for children <18 years old and of Crapo for adults 18 years and older  **☐ Quality check performed**  **☒ Lung function procedure stated**  ATS  **☐ Cleaning of lung function data**  **described**  **☐ Person who analyzed PFT was**  **blinded to the exposure** |
| Centres:  Single center  Institution:  Fred Hutchinson Cancer Research Center (FHCRC)  Country:  USA  Treatment era:  1969-1995 | Inclusion criteria:  Myeloablative HSCT at FHCRC, at least 5 years survival after HSCT and at least 6y old  Cancer diagnosis:  ALL 36%  AML 21%  CML 10%  MDS 6%  JMML 1.5%  Non-malignant 19%  Age at diagnosis:  Median (range) yr  At HSCT 8.3y (0.3-18)  Age at follow-up:  Median (range) yr  19y (6.5-40.5)  Time since diagnosis  Median (range) yr  After HSCT: 10.5y (5-27.5) | Name of protocol  Chemotherapy (doses)/ Radiotherapy(doses)  a) 9.2 – 10.0 Gy single-fraction TBI (SFTBI) (13%)  b) fractionated TBI (FTBI) with exposures of 2.0 – 2.75 Gy for 6 to 7 consecutive days or hyperfractionated exposures of 1.2 Gy 2 to 3 times daily for 4 consecutive days (62%).  Between 1986 and 1990, some patients received 14.4 Gy TBI from a linear accelerator with lung shielding.  Overall median dose not stated.  Most TBI regimens included CY 60 mg/kg/day for 2 days.  Most chemotherapy-only regimens utilized CY 50 mg/kg/day for 4 days for aplastic anemia (AA) patients and CY (50 mg/kg/day for 4 days or 60 mg/kg/day for 2 days) combined with BU 4 mg/kg/day for 4 days for patients with a hematologic malignancy and some patients with non-malignant hematologic disorders.  Non-TBI regimens: Cyclophosphamide (CY) with/ without procarbazine (n=24, 10%), busulfan with CY or CY dimethylmyleran (12%)  Surgery (kind of surgery):  NA  HSCT (allo/auto/donor specifics/Conditioning):  5% autologous  95% allogeneic | How was outcome assessed?  Medical records, prospective measurements, clinical examination, pulmonary function tests  Incidence, prevalence:  Obstruction (O)/ restriction (R), resp.:  After 5-10 years FU: 8% (O)/ 20% (R);  After 10-15 years: 12% (O)/ 36% (R);  After 15-20 years: 9% (O)/ 43% (R);  After >20 years: 38% (O)/ 50% (R)  Risk factors:  Univariate analysis:  Restrictive: no PICO-relevant variable analyzed  Obstructive: prior cyclophosphamide (p=0.05), chronic GvHD (p=0.02)  Multivariate analysis:  Restrictive: SFTBI (OR 22, 95%-CI 3.9-120), FTBI 1.2 Gy (OR 2.5, 95%-CI 0.4-16), FTBI 2.0-2.25Gy (OR 2.8, 95%-CI 0.6-13) versus non-TBI  Obstructive: cGvHD (OR 4.4, 95%-CI 1.6-12); SFTBI (OR 0.1, 95%-CI 0.5-0.5) seemed negatively associated, FTBI 1.2Gy (OR 0.1, 95%-CI 0.0-1.4) and FTBI 2.0-2.25Gy (OR 0.9, 95%-CI 0.3-2.8) were not associated.  If longitudinal data available:  Mean/Median duration until pulmonary disease develops:  NA  **Exact results available from publication, TABLE VI**: “Univariate Analysis of Risk Factors Associated With Restrictive Lung Disease and Obstructive Lung Disease” and **TABLE VII** “Multivariate Risk Factors for Restrictive Lung Disease” and **TABLE VIII** “Multivariate Risk Factors for Obstructive Lung Disease” | Analysis:  Multivariate analysis for restrictive and obstructive lung disease  Limitations: Different exposures and cross- sectional design limit the strength of the study  Strength: For HSCT studies, large number of participants.  Potential bias/methodological problems: Follow-up data is aggregate data and not follow-up information of specific patients. |

| **Main findings/message:**  **Long-term follow-up of lung function after bone marrow transplantation frequently remains abnormal. Post-transplant survival is related to pre-transplant DLCO. Older age at transplantation is associated with significant decrease of FVC z-score (p=0.026) and DLCO z-score (p=0.039) from pre-transplant to post-transplant.** | | | | |
| --- | --- | --- | --- | --- |
| **J. P. Ginsberg, et al.** Pre-transplant lung function is predictive of survival following pediatric bone marrow transplantation. 2010;54:454-60. http://dx.doi.org/10.1002/pbc.22337 | | | | |
| **Study design**  **Treatment era**  **Years of follow-up** | **Participants** | **Treatment** | **Main outcomes** | **Additional remarks** |
| ☒ **Cohort**  ☐ **Cross-sectional**  ☐ **Case-control**  ☐ **Other:**  **__________**  ☒ **Retrospective**  ☐ **Prospective** | Study population (N)   - Original cohort: 457 - Eligible cohort: - Analyzed cohort:   317 (post-HSCT)  273 (pre-HSCT)  133 (both pre- and post-HSCT) | ☒ 1 **HSCT: 1a**  ☐ **2 Cyclophosphamid**  ☐ **3 Methotrexate**  ☐ **4 Gemcitabine**  ☐ **5 Bleomycin**  ☐ **6 Busulfan**  ☐ **7 Lomustin (CCNU)**  ☐ **8 Carmustin (BCNU)**  ☐ **9 Radiotherapy lung**  ☐ **10 Surgery**  ☐ **11 Combinations**  ☐ **12 Tobacco exposure** | ☐ **Pulmonary diseases**  ☐ **Pulmonary symptoms**  ☒ **Pulmonary function test**  ☐ **Absolute values**  ☒ **Z-scores**  ☐ **Percentage predicted**  **☒ Percentage pathological tests**  (e.g. 24% with reduced FEV1) | ☒ **Longitudinal data available**  Only aggregate, not single patient  **☐ Control group mentioned**  **☒ Reference values stated**  Rosenthal, Hankinson  **☐ Quality check performed**  **☒ Lung function procedure stated:**  ATS  **☐ Cleaning of lung function data**  **described**  **☐ Person who analyzed PFT was**  **blinded to the exposure** |
| Centres:  Multicentre: Children’s Hospital of Philadelphia; Hospital for Sick Children (Toronto)  Country: USA/Canada  Treatment era:  1978-2005  Years of Follow-up:  Only time point of last lung function measurement available | Inclusion criteria:  Treatment with stem cell transplant, at least one lung function test result available  Exclusion:  >1 transplantation  Cancer diagnosis:  Those with post-HSCT PFTs:  ALL 84 (26%)  AML 83 (26%)  Immune 7 (2%)  Other leukemia 14 (5%)  Lymphoma 34 (11%)  Non-malignant 47(15%)  Other 1 (0%)  Solid 37 (12%)  Age at diagnosis:  Categories only:  < 6 years: 45 (14%)  6–12 years: 129 (41%)  12–18 years: 122 (38%)  18+ years: 21 (7%)  Age at follow-up:  NA  Time since diagnosis  NA | Name of protocol:  NA  Chemotherapy (doses)  no dose  Radiotherapy(doses):  no dose  Surgery (kind of surgery):  no further information  Type of transplantation:  For those with post-HSCT PFTs:  Allogeneic 76%  Autologous 24%  Conditioning  Busulfan/ CYC 74 (23%)  TBI/ CYC 127 (40%)  CTX+ other 49 (15%)  TBI+ other 47 (15%)  Other 16 (5%)  Duration of protocol:  PFTs at the following time points in post-transplant analysis:  Pre-transplant: 133 (42%);  0–6 months post: 44 (14%);  6–12 months post: 107 (34%);  1–2 years post: 142 (45%);  2–5 years post: 156 (49%);  5+ years post: 98 (31%) | How was outcome assessed?  Medical databases of transplanted patients and of the respiratory department for PFTs  Lung Function Score (LFS) calculated with pre-transplant values:  - FEV1 % and DLCO >80% predicted=1, 70 – 80% predicted=2, 60–70% predicted=3, <60% predicted=4.  - FEV1 and DLCO scores were summed (maximum value of 8)  - Sum assigned to one of four categories as the pre-transplant lung function score (LFS): LFS=2: Category I, LFS=3–4: Category II, LFS=5–6: Category III, LFS=7–8: Category IV  Risk factors (RR, OR…):  Predictors of post-HSCT PFTs (at the last time of measurement):  - Older age at transplantation is associated with decrease of:  - FVC z-score (p=0.026)  - DLCO z-score (p=0.039)  - FEV1 z-score (p=0.079)  - TLC z-score (p=0.432)  from pre-transplant to post-transplant (ANOVA).  If longitudinal data available:  2-5 years post-transplant compared to pre-transplant (156 patients)  FVC z-scores: mean -1.55  FEV1 z-scores: mean -1.56  TLC z-scores mean -0.70  DLCO z-scores mean -1.71  >5 years post-transplant compared to pre-transplant (98 patients)  FVC z-scores: mean -1.68  FEV1 z-scores: mean -1.70  TLC z-scores mean -1.12  DLCO z-scores mean -1.54  Mean/Median duration until pulmonary disease develops  NA  **Exact results available from publication, TABLE III**: “Correlates of Last Post-Transplant Pulmonary Function” | Analysis:  - One-way analysis of variance: for differences in pulmonary function by type of transplant, diagnosis, conditioning, and age at transplant  - Kaplan–Meier curves and proportional hazards model: relationships between post-transplant survival and LFS, age, diagnosis, study site, race, and sex.  - standard errors: differences between pre- and post-transplant populations  - P-values  Limitations:  Retrospective data  PFTs at different intervals  Incomplete data on pre- and post-treatment PFTs  Strength:  Relatively large cohort  Longitudinal data  Results as percentage predicted and z-scores  Potential bias/methodological problems:  Not all the patients have PFTs >2 years after transplantation  Unknown if patients without post-PFT differ significantly than the ones with post-PFT, e.g. had better lung function |

| **Main findings/message:**  **59% of pulmonary dysfunction after allogeneic HSCT with most events occurring during the first year post-HSCT. Risk factors were extensive chronic GVHD and abnormal pre-treatment PFTs. Limited chronic GvHD and age at HSCT were not risk factors.** | | | | |
| --- | --- | --- | --- | --- |
| **L. M. Madanat-Harjuoja, et al.** Pulmonary function following allogeneic stem cell transplantation in childhood: a retrospective cohort study of 51 patients. 2014;18:617-24. 10.1111/petr.12313 | | | | |
| **Study design**  **Treatment era**  **Years of follow-up** | **Participants** | **Treatment** | **Main outcomes** | **Additional remarks** |
| ☒ **Cohort**  ☐ **Cross-sectional**  ☐ **Case-control**  ☐ **Other:**  **__________**  ☒ **Retrospective**  ☐ **Prospective** | Study population (N)   - Original cohort: 163 - Eligible cohort: 96 - Analyzed cohort: 51 | ☒ 1 **HSCT 1a, 1b**  ☐ **2 Cyclophosphamid**  ☐ **3 Methotrexate**  ☐ **4 Gemcitabine**  ☐ **5 Bleomycin**  ☐ **6 Busulfan**  ☐ **7 Lomustin (CCNU)**  ☐ **8 Carmustin (BCNU)**  ☐ **9 Radiotherapy lung**  ☐ **10 Surgery**  ☐ **11 Combinations**  ☐ **12 Tobacco exposure** | ☐ **Pulmonary diseases**  ☒ **Pulmonary symptoms**  ☐ **Pulmonary function test**  ☐ **Absolute values**  ☐ **Z-scores**  ☒ **Percentage predicted**  **☒ Percentage pathological tests**  (e.g. 24% with reduced FEV1) | ☒ **Longitudinal data available**  **☐ Control group mentioned**  **☒ Reference values stated**  Quanjer (ERS)  **☒ Quality check performed**  **☒ Lung function procedure stated**  ATS  **☒ Cleaning of lung function data**  **described**  **☐ Person who analyzed PFT was**  **blinded to the exposure** |
| Centres:  Single center: Children’s Hospital, Helsinki University Central Hospital  Country:  Finland  Treatment era:  1993-2005 | Inclusion criteria:  Allogeneic HSCT.  Final cohort (n=51) had at least one visit >1 yr post-HSCT and a baseline PFT.  No metabolic disease, >6 mts post-HSCT, age >6 yrs  Cancer diagnosis:  ALL 55%  AML or MDS 28%  Non-malignant 8%  CML 6%  CGD 4%  Lymphoma 2%  Age at HSCT:  Median (range) yr: 11.2 (6.2-19)  Age at follow-up:  Median (range) yr  Time since HSCT:  Median (range) yr: 4.1 | Name of protocol  NA  Chemotherapy (doses)  Cyclophosphamide 47%  Cytarabine 47%  Other 6%  Radiotherapy(doses)  Fractionated TBI 10-14 Gy 98%  Total nodal irradiation 6 Gy 2%  HSCT:  Cyclophosphamide or cytarabine-based conditioning 94% +  TBI 98%, Total nodal irradiation 2% | How was outcome assessed?  Medical records.  PFTs performed at baseline (pre-HSCT) and at follow-up visits starting at 6 month post-HSCT.  RLD= FVC<80% + FEV1/FVC>80%  OLD=FEV1<80% + FEV1/FVC<80%  Patients with FVC<60% underwent lung biopsy.  Incidence, Prevalence:  15.7% (8/51) had an abnormal pre-treatment PFT (5 restrictive, 3 mild obstructive)  59% developed abnormal PFT; 73% restrictive, 26% obstructive  PFT at baseline, 1-yr post HSCT, 5-yr post-HSCT  FVC: mean 93%, 75%, 77%  FVC: median 92%, 82%, 83%  FEV1: mean 95%, 75%, 75%  FEV1: median 95%, 87%, 83%  FEV1/FVC: mean 0.98, 0.90, 0.84  Significant reduction within 12 months in FEV1 and FVC in patients with normal baseline. After 12 months reduction not significant.  Risk factors:  Extensive chronic GVHD associated with a decline in FVC and FEV1.  🡪 Chronic GVHD (none=reference)  Extensive: HR=10.20 (CI 2.42-43.03); p=0.002  Limited: HR=0.42 (CI 0.10-1.83); p=0.247  🡪 Age at HSCT (6-11 years= reference):  HR=1.14 (CI 0.40-3.26); p=0.804 (12-19yr)  12/51pts had FVC<60%, of which 8 had lung biopsies.  11/12 had extensive GVHD including lung involvement, as verified on lung biopsy. 4/12 (33%) died of pulmonary complications.  **Exact results available from publication, Table 3**: “Risk factors for pulmonary dysfunction in survivors of allogeneic stem cell transplantation in childhood” | Analysis:  Fisher exact test for evaluation of risk factors.  Cox proportional hazards model for analysis of risk factors on pulmonary function.  Longitudinal models for repeated measurements.  Limitations:  Retrospective analysis, small cohort, Only about half of eligible population had PFT; single center  No data regarding doses of chemotherapy.  Strength:  Longitudinal data regarding timing of abnormal PFT.  PFTs were reviewed according to ATS protocol.  Potential bias/methodological problems:  Selection bias – only patients with PFTs after treatment were included. |

| **Main findings/message:**  **Abnormal pulmonary function test results are present in up to 64% of survivors of pediatric allo-HSCT (DLCO). Older age (continuous) at allogeneic HSCT is associated with obstructive dysfunction measured by FEF 25-75% (HR 1.1; p=0.038) and impaired diffusion capacity (HR 1.1; p=0.005).** | | | | |
| --- | --- | --- | --- | --- |
| **H. Inaba, et al**. Pulmonary dysfunction in survivors of childhood hematologic malignancies after allogeneic hematopoietic stem cell transplantation. 2010;116:2020-30. 10.1002/cncr.24897 | | | | |
| **Study design**  **Treatment era**  **Years of follow-up** | **Participants** | **Treatment** | **Main outcomes** | **Additional remarks** |
| ☒ **Cohort**  ☐ **Cross-sectional**  ☐ **Case-control**  ☐ **Other:**  **__________**  ☒ **Retrospective**  ☐ **Prospective** | Study population (N)   - Original cohort: NA - Eligible cohort: 208 - Analyzed cohort: 89 | ☒ 1 **HSCT 1a**  ☐ **2 Cyclophosphamid**  ☐ **3 Methotrexate**  ☐ **4 Gemcitabine**  ☐ **5 Bleomycin**  ☐ **6 Busulfan**  ☐ **7 Lomustin (CCNU)**  ☐ **8 Carmustin (BCNU)**  ☐ **9 Radiotherapy lung**  ☐ **10 Surgery**  ☐ **11 Combinations**  ☐ **12 Tobacco exposure** | ☐ **Pulmonary diseases**  ☐ **Pulmonary symptoms**  ☒ **Pulmonary function test**  ☐ **Absolute values**  ☐ **Z-scores**  ☒ **Percentage predicted**  **☒ Percentage pathological tests**  (e.g. 24% with reduced FEV1) | ☒ **Longitudinal data available**  **☐ Control group mentioned**  **☒ Reference values stated**  Hankinson  **☐ Quality check performed**  **☒ Lung function procedure stated**  ATS  **☐ Cleaning of lung function data**  **described**  **☐ Person who analyzed PFT was**  **blinded to the exposure** |
| Centres:  Single  Country:  USA  Treatment era:  1990-2005  Years of Follow-up:  Median 8.9 (range 1.7-16.4) years | Inclusion criteria:  Allo-HSCT at St. Jude, at least 6 years old, available pre-HSCT PFT for comparison  Cancer diagnosis:  ALL, AML, MDS, CML  Age at HSCT:  Median 12.7 (range 6.6-21.3) years  Age at follow-up:  Not stated  Time since diagnosis  Not stated | Name of protocol: Not stated  Chemotherapy (doses): Mostly cyclophophamide, no pre-HSCT details known  Radiotherapy(doses): Mostly TBI of 12 or 14 Gy  Surgery (kind of surgery): N/A  HSCT (allo/auto/donor specifics/Conditioning): all allo-HSCT patients, 46.1% unrelated, 41.6% matched sibling, 12.3% parent, 88.8% bone marrow, 11.2% PBSC, 78.6% fully matched, mostly Cy/TBI +/- other additions | How was outcome assessed?  PFT per American Thoracic Society guidelines  Incidence, Prevalence:  Respiratory dysfunction in up to 64%, FEV1/FVC – 22.5%, FEV1 – 36%, FEF25-75 – 49.4%, RRV/TLC – 38.2%, FVC – 39.3%, TLC 43.8%, DLCO – 64%  Risk factors (HR):  Obstructive: male (2.1), respiratory event within 1 year of HSCT (2.7-3.2) PBSC (2.5), High risk disease (1.8), older age (1.1; p=0.038)  Restrictive: PBSC (2.7), respiratory event <1 year of HSCT (2.3), acute GVHD (1.9)  DLCO: high risk disease (2.6), CMV positive (1.7), older age (1.1)  If longitudinal data available:  Gradual decline over time | Analysis:  Longitudinal methods, Cox proportional hazards models, Fisher’s exact test, Cumulative incidences accounting for competitive risks; Independent analyses were performed for the 9 response variables  Limitations:  Restricted to ages 6 and above, restricted to those with good PFT results pre-HSCT  Strength:  Medium size, long follow-up, longitudinal data  Potential bias/methodological problems: survivor bias, participation bias, testing bias |

| **Main findings/message:**  **More than 1 year after allogeneic SCT 62% show impaired diffusion capacity (TLCO), 41% restrictive, and 11% obstructive disorder. Restrictive disease > 1 year after SCT associated with female sex (p=0.002) and younger age at SCT (p=0.08). Neither radiotherapy (TBI/TAI) nor donor-type were identified as risk factors for restrictive and/or obstructive disease. Significant reduction >1year after SCT compared to baseline only for TLCO. Significant reduction in FVC, FEV1 and TLC <1 year improve but not to normal again.** | | | | |
| --- | --- | --- | --- | --- |
| **J. Wieringa, et al.** Pulmonary function impairment in children following hematopoietic stem cell transplantation. 2005;45:318-23. 10.1002/pbc.20304 | | | | |
| **Study design**  **Treatment era**  **Years of follow-up** | **Participants** | **Treatment** | **Main outcomes** | **Additional remarks** |
| ☒ **Cohort**  ☐ **Cross-sectional**  ☐ **Case-control**  ☐ **Other:**  **__________**  ☐ **Retrospective**  ☒ **Prospective** | **Study population (N)**   - Original cohort: NA - Eligible cohort: 106 - Analysed cohort: 39 | ☒ **1** **HSCT a, d**  ☐ **2 Cyclophosphamid**  ☐ **3 Methotrexate**  ☐ **4 Gemcitabine**  ☐ **5 Bleomycin**  ☐ **6 Busulfan**  ☐ **7 Lomustin (CCNU)**  ☐ **8 Carmustin (BCNU)**  ☒ **9 Radiotherapy lung**  ☐ **10 Surgery**  ☐ **11 Combinations**  ☐ **12 Tobacco exposure** | ☐ **Pulmonary diseases**  ☐ **Pulmonary symptoms**  ☒ **Pulmonary function test**  ☐ **Absolute values**  ☐ **Z-scores**  ☒ **Percentage predicted**  **☒ Percentage pathological tests**  (e.g. 24% with reduced FEV1) | ☒ **Longitudinal data available**  **☒ Control group mentioned**  **☒ Reference values stated**  Polgar and Weng  **☐ Quality check performed**  **☐ Lung function procedure stated**  **☐ Cleaning of lung function data**  **described**  **☐ Person who analyzed PFT was**  **blinded to the exposure** |
| Design  Prospective cohort  Centres:  Single centre, Leiden  Country: Netherlands  Treatment era:  seen in late effects clinic 2001-2003  Years of Follow-up:  Median 4.5 years Range 0.5-10 years | Study population:  After allogeneic SCT  Inclusion criteria:  Allogenic SCT, visit outpatient late effect clinic, PFT before and at least twice after SCT  Cancer diagnosis:  Malignant: 30/39 (77%)  - ALL 39%  - AML 18%  - CML 2%  - MDS 16%  - JMML 2%  Benign: (23%)  - SAA 14%  - Thalassemia 2%  - Fanconi anemia 2%  - X-linked Adrenoleuko-  dystrophy 5%  Age at SCT:  Median (range) 10 years (4-18)  Time since SCT  Median 4.5 years  Range 0.5-10 years | Name of protocol  According to disease:  DCLSG protocols ALL-6, 7 and 8, relapse ALL 90 and 98 and ANLL 87, 92, 94 and 97  Chemotherapy (doses)  CYC 120 – 200 mg/m2 and TBI or thoracoabdominal irradiation in 31 patients  Busulfan (6.20 mg/kg) and CYC in 6 patients  CYC only in 2 patients  Radiotherapy dose:  TBI: 7 - 12Gy  Thoracoabdominal: 4 - 5Gy  GvHD prophylaxis: NA  HSCT  Graft type: allogeneic, not specified | How was outcome assessed?  PFT: spirometry (FVC, FEV1), helium dilution method (FRC, RV), single breath method using helium (TLCO)  Parameters recorded as percentage predicted  Reference for PFT: age, sex, length matched; paper Polgar and Weng  Pathological when <80% predicted  Prevalence:  Significant reduction >1year after SCT compared to baseline only for TLCO  Significant reduction in FVC, FEV1 and TLC <1 year improve but not to normal again  Risk factors:  -Restrictive lung disease > 1 year after SCT associated with female sex (p=0.002) and younger age (p=0.004)  -Decrease in TLCO < 1 year after SCT more pronounced in boys and in those with malignant disease (p=0.009)  - Decrease in TLCO > 1 year after SCT more in patients with malignant disease (p=0.05)  - age at HSCT: trend towards a higher TLC at SCTpost2 for patients older than 10 years (P=0.08), w  Longitudinal data:  See table III in additional information  Mean/Median duration until pulmonary disease develops NA | Analysis:  Changes in lung function values between pre-SCT, < 1 year post SCT and > 1 year post SCT: student paired t-test  Risk factor analysis: linear regression  Limitations:  Small cohort  Heterogeneous exposure before SCT which can influence “baseline” PFT  Strength:  Assessment via pulmonary function test  Longitudinal data  Potential bias/methodological problems:  Bias due to confounding  No information on 67 patients who had SCT in the study period and visited the outpatient clinic but did not have PFT at the given time points.  PFT procedure not mentioned |

| **Main findings/message:**  **Cumulative incidence of pulmonary dysfunction at 10 years is 63.2%. TBI, age at HSCT (per year), and chronic GvHD are associated with reduced DLCO% predicted. TBI is associated with reduced TLC% predicted and FEV1/FVC predicted.** | | | | | | | | | | | | |
| --- | --- | --- | --- | --- | --- | --- | --- | --- | --- | --- | --- | --- |
| **W. Leung, et al.** A prospective cohort study of late sequelae of pediatric allogeneic hematopoietic stem cell transplantation. 2007;86:215-24. 10.1097/MD.0b013e31812f864d | | | | | | | | | | | | |
| **Study design**  **Treatment era**  **Years of follow-up** | | **Participants** | | | **Treatment** | | | **Main outcomes** | | | **Additional remarks** | |
| ☒ **Cohort**  ☐ **Cross-sectional**  ☐ **Case-control**  ☐ **Other:**  **__________**  ☐ **Retrospective**  ☒ **Prospective** | Study population (N)   - Original cohort: NA - Eligible cohort: 204 - Analyzed cohort: 155 | | | ☒ 1 **HSCT: 1a, 1b, 1d**  ☐ **2 Cyclophosphamid**  ☐ **3 Methotrexate**  ☐ **4 Gemcitabine**  ☐ **5 Bleomycin**  ☐ **6 Busulfan**  ☐ **7 Lomustin (CCNU)**  ☐ **8 Carmustin (BCNU)**  ☐ **9 Radiotherapy lung**  ☐ **10 Surgery**  ☐ **11 Combinations**  ☐ **12 Tobacco exposure** | | | ☐ **Pulmonary diseases**  ☐ **Pulmonary symptoms**  ☒ **Pulmonary function test**  ☐ **Absolute values**  ☐ **Z-scores**  ☒ **Percentage predicted**  **☒ Percentage pathological tests**  (e.g. 24% with reduced FEV1) | | | ☐ **Longitudinal data available**  **☐ Control group mentioned**  **☐ Reference values stated**  **☐ Quality check performed**  **☐ Lung function procedure stated**  **☐ Cleaning of lung function data**  **described**  **☐ Person who analyzed PFT was**  **blinded to the exposure** | | |
| Centres:  Single; St. Jude Children’s Research Hospital (Memphis, TN)  Country:  USA  Treatment era:  1990-2003  Years of Follow-up:  Median (range) yr  9 (3.1-15.9) | | Inclusion criteria:  Surviving 1 or more year after allogeneic HSCT  Cancer diagnosis:  Myeloid malignancy 84 (54%), lymphoid malignancy 40 (26%), non-malignant 31 (20%)  Age at diagnosis:  Median (range) yr 9.7 (0.5 – 21.4)  Age at follow-up:  Median (range) yr  18.5 (4.6 – 36.1)  Time since HSCT  Median (range) 9 (3.1-15.9) | | | Name of protocol: NA  Radiotherapy (doses)  TBI 123 (79%)  Dose of TBI 14.4Gy in n=59  Dose of TBI 8-12Gy in n=64  Dose of TBI none in n=32  Surgery (kind of surgery)  HSCT (allo/auto/donor specifics/Conditioning)  Alkylator-based conditioning 32 (21%) | | | How was outcome assessed?  Prospective pulmonary function test  Incidence, Prevalence:  At least 1 parameter abnormal /77 survivors  Cumulative incidence at 10 years 63.2%  Risk factors:  DLCO <80% predicted:  - TBI: HR=2.2 (95%CI 1.07-5.09) p=0.026  - Age, per year: 1.1 (95%CI 1.04-1.17)  p=<0.001  - chronic GVHD: HR=1.96 (95%CI 1.12-3.44)  p=0.025  TLC <80% predicted:  - TBI: HR=2.4 (95%CI 1.04-4.95) p=0.03  FEV1/ FVC predicted:  - TBI: HR=2.4 (95%CI 1.1-5.74) ü=0.02  Longitudinal data available:  No  **Exact results available from publication, TABLE 4**: “Risk Factors for Late Sequelae” | | | Analysis:  Cumulative incidence function of each late event was estimated  Comparison by Kalbfleisch and Prentice and Gray.  Limitations:  Exposure data very limited, unclear how outcomes were assessed, procedures not clearly described. Only associations described that were found to be significant.  Strength:  Prospective cohort of exclusively childhood cancer survivors  Potential bias/methodological problems:  Recommendations:  - if age > 8 at HSCT, TBI:  🡪 PFTs biennilally started 3 years after HSCT – pulmonary referral, flue shot, counselling for smoking, job, relocation | |
| **Main findings/message:**  **The odds of developing restrictive and hyperinﬂation defects increased with increasing mean and maximum lung dose but also with lung volume receiving at least 10Gy and 20Gy of irradiation. Thoracic surgery prior to radiation increased the odds of reduced FEV1 and RV/TLC. Bleomycin was found to have a protective effect but those patients also had less radiation why this finding was questioned.** | | | | | | | | | | | | |
| **A. De, et al.** Correlation of pulmonary function abnormalities with dose volume histograms in children treated with lung irradiation. 2015;50:596-603. | | | | | | | | | | | | |
| **Study design**  **Treatment era**  **Years of follow-up** | | | **Participants** | | | **Treatment** | | | **Main outcomes** | | | **Additional remarks** |
| ☒ **Cohort**  ☐ **Cross-sectional**  ☐ **Case-control**  ☐ **Other:**  **__________**  ☒ **Retrospective**  ☐ **Prospective** | | | Study population (N)   - Original cohort: 170 - Eligible cohort: 139 - Analyzed cohort: 49 | | | ☐ 1 **HSCT:**  ☐ **2 Cyclophosphamid**  ☐ **3 Methotrexate**  ☐ **4 Gemcitabine**  ☒ **5 Bleomycin**  ☐ **6 Busulfan**  ☐ **7 Lomustin (CCNU)**  ☐ **8 Carmustin (BCNU)**  ☒ **9 Radiotherapy lung: 9, 9ai**  ☒ **10 Surgery**  ☐ **11 Combinations**  ☐ **12 Tobacco exposure** | | | ☒ **Pulmonary diseases**  ☒ **Pulmonary symptoms**  ☒ **Pulmonary function test**  ☐ **Absolute values**  ☐ **Z-scores**  ☒ **Percentage predicted**  **☒ Percentage pathological tests**  (e.g. 24% with reduced FEV1) | | | ☐ **Longitudinal data available**  **☐ Control group mentioned**  **☒ Reference values stated**  Hankinson, Wang  **☐ Quality check performed:**  **☒ Lung function procedure stated:**  ATS  **☐ Cleaning of lung function data**  **described**  **☐ Person who analyzed PFT was**  **blinded to the exposure** |
| Centres: monocentric: Children’s Hospital Los Angeles (CHLA), USA  Country:  Los Angeles  Treatment era:  1999 - 2009  Years of Follow-up:  Median: 2,91 Range: 0,01-8,28 | | | Inclusion criteria:  - Radiotherapy to the lungs.  - One PFT post irradiation  - Exclusion: TBI or palliative radiation  Cancer diagnosis:  Hodgkin Lymphoma (78%)  Wilms tumor (2%)  Ewing sarcoma (8%) Rhabdomyosarcoma (4%)  Non-Hodgkin lymphoma (2%)  Neuroblastoma (2%) Thymoma (2%)  Synovial sarcoma (2%)  Age at radiotherapy  Median: 13,8 Range: 4,02-20,98  Age of pulmonary function test after irradiation  Median: 2,91 Range: 0,01-8,28 | | | Name of protocol:  not specified, doses not specified  Bleomycin in 78% of patients, Cyclophosphamide in 82%  Doxorubicin in 94%  Dosimetric parameter Median (range)  Prescribed dose of radiation (Gy) 21 (10.5–1)  Mean lung dose (Gy) 8.95 (1.1–1.1)  Maximum lung dose (Gy) 23.04 (12–8.2)  Surgery (kind of surgery)  18% of chest surgery (chest wall surgery, thoracoscopic biopsy of mediastinal or lung mass, incisional biopsy, and lung parenchymal resection)  Duration of protocol:  Not specified | | | How was outcome assessed?  Retrospective review of medical records, functional tests  Last pulmonary function test selected for the study  % abnormal; Median value (range)  FVC %pred: 24% - 94 (22–144)  FEV1 %pred: 29%- 91 (24–131)  FEV1/FVC %pred: 14% - 86 (67–105)  FEF25–5% %pred: 20% - 87 (19–142)  RV %pred: 21%- 98 (17–246)  TLC %pred: 15%- 99 (28–165)  RV/TLC: 21% - 21 (5–49)  Phase II N2 %N 2/L: 27% 1.7 (0–7.2)  DLCO adj %pred: 9% - 92.7 (28–157)  DLCO adj/VA ml/mmHg/min/L: 5% - 5.5 (3.7-9)  Risk factors:  Abnormal FVC:  - Thoracic surgery: OR 8.0; p = <0.01  - Bleomycin: OR 0.15; p=<0.05  - Age at radiation: OR1.13; p=NS  - Mean dose (in Gy): OR 1.22; p=<0.01  - Max dose (on Gy): OR 1.10; p=<0.01  Abnormal FEV1:  - Thoracic surgery: OR 3.2; p=NS  - Bleomycin: OR 0.07; p=<0.01  - Age at radiation: OR 1.03; p=NS  - Mean dose (in Gy): OR 1.20; p=<0.01  - Max dose (on Gy): OR 1.12; p=<0.01  Abnormal FEF25-75%:  - Thoracic surgery: OR 2.35; p=NS  - Bleomycin: OR 0.18; p=<0.05  - Age at radiation: OR 1.09; p=NS  - Mean dose (in Gy): OR 1.18; p=<0.01  - Max dose (on Gy): OR 1.06; p=<0.05  Abnormal TLC:  - Thoracic surgery: OR 1.94; p=NS  - Bleomycin: OR 0.27; p=NS  - Age at radiation: OR 1.14; p=NS  - Mean dose (in Gy): OR 1.30; p=<0.01  - Max dose (on Gy): OR 1.07; p=<0.05  Abnormal RV/TLC  - Thoracic surgery: OR 8.5; p=<0.01  - Bleomycin: OR 0.15; p=<0.05  - Age at radiation: OR 1.05; p=NS  - Mean dose (in Gy): OR 1.30; p=<0.01  - Max dose (on Gy): OR 1.26; p=<0.05  Abnormal DLCO adj:  - Thoracic surgery: OR 1.89; p=NS  - Bleomycin: OR 0.06; p=<0.05  - Age at radiation: OR 1.01; p=NS  - Mean dose (in Gy): OR 1.27; p=<0.01  - Max dose (on Gy): OR 1.07; p=<0.05  *mean dose = mean lung dose  The odds of developing restrictive and hyperinﬂation defects increased with increasing Vdose beginning at V10 and V20  Obstructive disease  - Thoracic surgery: OR 5.89; p<0.05  - Bleomycin: OR 0.27; p=NS  - Mean dose (in Gy): OR 0.99; p=NS  - Max dose (on Gy): OR 1.03; p=NS  - Prescribed dose (in Gy): OR 1.05; p=NS  Restrictive disease  - Thoracic surgery: OR 1.94; p=NS  - Bleomycin: OR 0.27; p=NS  - Mean dose (in Gy): OR 1.30; p<0.01  - Max dose (on Gy): OR 1.07; p<0.05  - Prescribed dose (in Gy): OR 1.04; p=NS  Hyperinflation  - Thoracic surgery: OR 8.5; p<0.01  - Bleomycin: OR 0.15; p<0.05  - Mean dose (in Gy): OR 1.29; p<0.01  - Max dose (on Gy): OR 1.26; p<0.01  - Prescribed dose (in Gy): OR 1.27; p<0.01  Diffusion defect  - Thoracic surgery: OR 1.07; p=NS  - Bleomycin: OR 0.08; p<0.01  - Mean dose (in Gy): OR 1.16; p<0.05  - Max dose (on Gy): OR 1.05; p=NS  - Prescribed dose (in Gy): OR 1.05; p=NS | | | Analysis:  Univariate analysis/ logistic regression  Limitations:  Only patients who underwent pulmonary function testing were included in the study: 49/170 28,8%  The majority of patients received radiotherapy because they are treated for lymphoma  Only univariate (unadjusted) analyses were done  Different timepoints assessed (last test available) with a wide range.  Strength:  Potential bias/methodological problems: only patients who have functional testing are included |

| **Main findings/message:**  **Clinical and functional signs of pulmonary dysfunction in survivors of HL and NHL. None of the patients has subjective symptoms and signs of pulmonary dysfunction. FEV1, RV, RV/TLC, DLCO were significantly lower in the group treated with chemotherapy and radiotherapy to the chest compared to chemotherapy only.** | | | | |
| --- | --- | --- | --- | --- |
| A. Oguz, et al. Long-term pulmonary function in survivors of childhood Hodgkin disease and non-Hodgkin lymphoma. 2007;49:699-703. 10.1002/pbc.21175 | | | | |
| Study design  Treatment era  Years of follow-up | Participants | Treatment  (= treatment analyzed in the paper) | Main outcomes | Additional remarks |
| **☐ Cohort**  **☒ Cross-sectional**  **☐ Case-control**  **☐ Other:**  **__________**  **☐ Retrospective**  **☒ Prospective** | **Study population (N)**  **Original cohort:** NA  **Eligible cohort:**  **Analyzed cohort:** 75 | **☐ 1 HSCT a, b**  **☐ 2 Cyclophosphamid**  **☐ 3 Methotrexate**  **☐ 4 Gemcitabine**  **☐ 5 Bleomycin**  **☐ 6 Busulfan**  **☐ 7 Lomustin (CCNU)**  **☐ 8 Carmustin (BCNU)**  **☒ 9 Radiotherapy lung**  **☐ 10 Surgery**  **☐ 11 Combinations**  **☐ 12 Tobacco exposure** | **☒ Pulmonary diseases**  **☐ Pulmonary symptoms**  **☐ Pulmonary function test**  **☐ Absolute values**  **☐ Z-scores**  **☒ Percentage predicted**  **☐ Percentage pathological tests**  **(e.g. 24% with reduced FEV1)** | **☐ Longitudinal data available**  **☐ Control group mentioned**  **☒ Reference values stated**  reference equations  recommended by the European  Coal and Steel Community  Severity scoring in accordance  with ATS pulmonary function  laboratory guidelines  **☐ Quality check performed**  **☒ Lung function procedure stated**  ATS  **☐ Cleaning of lung function data**  **described**  **☐ Person who analyzed PFT was**  **blinded to the exposure** |
| Centres:  Single centre  Country:  Turkey  Treatment era:  1992-2003 | Cancer diagnosis:  Hodgkin Lymphoma  Non-Hodgkin Lymphoma  Group 1:  N=23 chemotherapy and thoracic chemotherapy  Group 2:  N=52 chemotherapy only  Age at diagnosis  median (range):  8 years (1.8-15.0)  Age at end of therapy  Median (range):  8.25 years (2.3-16)  Age at follow-up  Median (range):  13 (7.5-25)  Time since diagnosis  Median (range):  5 years (2-13) | Name of protocol  BFM 90 n=19  BFM 95 n=8  LSA2L2 n=3  LMT89 n=7  COMP n=1  COPP n=13  ABVD n=5  COPP/ABVD n=18  MOPP n=1  Radiotherapy (doses)  Hodgkin Lymphoma:  n=34/37  Non-Hodgkin Lymphoma:  n=7/38  Doses:  2400cGY (range 1500-4000cGy)  17/75 other than thoracic radiotherapy  Radiotherapy field:  Mantle/minimantle n=16  Mediasten n=3  Paraaortic/Abdomen n=5  Cervical n=13  Cranial n=2  Liver n=1 | How was outcome assessed?  Lung function: spirometry, lung volumes, and diffusion capacity measurements in all patients using the Sensor Medics Vmax22 spirometry and gas dilution system.  Definition pulmonary toxicity:  - obstructive disorder by FEV1, FVC, FEV1/FVC  - restrictive disorder by TLC, RV, RV/TLC ratio  - interstitial involvement: diffusion capacity for carbon monoxide (DLCO)  Incidence, Prevalence:  Abnormal PFT: n=10/75 (13%)  Group 1:  - 5/23 low DLCO  - 1/23 restrictive lung disease (low RV)  - 1/23 restrictive lung disease (low TLC)  - No obstructive disease  Group 2:  -4/52 low DLCO  - 2/52 restrictive lung disease (low RV)  - 1/52 restrictive lung disease (low TLC)  - No obstructive disease  Percent predicted values, compared by student t-test  - FVC  Group 1: 101.17 +- 19.93  Group 2: 102.94 +- 18.11  p=0.706  - FEV1  Group 1: 95.43 +-16.47  Group 2: 105.09 +- 19.01  p=0.038  - FEV1/FVC  Group 1: 96.43+-9.15  Group 2: 99.88 +- 11.93  p=0.221  - TLC  Group 1: 102.74 +- 15.63  Group 2: 106.73 +- 17.46  p=0.349  - RV  Group 1: 113.35 +-28.53  Group 2: 126.71 +- 24.63  p=0.043  - RV/TLC  Group 1: 25.39 +- 5.31  Group 2: 27.71 +- 4.92  p=0.062  - DLCO  Group 1: 101.35+-22.17  Group 2: 112.65 +- 4.92  p=0.025  **Exact results available from publication, TABLE II: “**Pulmonary Function Tests of the Patients” | Analysis:  Student t-test to compare percent predicted of lung function in group 1 and group 2  Limitations*:*  No data on long-term outcome of these patients  No cut offs for pathological disease  Did not take other risk factors between group1 and group2 into account, such as smoking or lung toxic chemotherapy  Strength:  Prospective design  Potential bias/methodological problems:  Reporting bias (retrospective)  Did not take chemotherapy and other risk factors into account |

| **Main findings/message:**  **In a cohort of children nearly 3 years after treatment with whole lung irradiation, the severity of the most recent pulmonary function abnormality (Z-scores for FEV1, TLC, and DLCO) did not correlate with age at the time of radiation (r2<0.001, r2=0.08, and r2=0.08 respectively) and total radiation dose (r2=0.002, r2=0.06, and r2=0.13, respectively)** | | | | |
| --- | --- | --- | --- | --- |
| D. J. Weiner, et al. Pulmonary function abnormalities in children treated with whole lung irradiation. 2006;46:222-7. | | | | |
| Study design  Treatment era  Years of follow-up | Participants | Treatment  (= treatment analyzed in the paper) | Main outcomes | Additional remarks |
| **☒ Cohort**  **☐ Cross-sectional**  **☐ Case-control**  **☐ Other:**  **__________**  **☒ Retrospective**  **☐ Prospective** | **Study population (N)**  **Original cohort:** NA  **Eligible cohort:** 63  **Analyzed cohort:** 30 with PFT | **☐ 1 HSCT a, b**  **☐ 2 Cyclophosphamid**  **☐ 3 Methotrexate**  **☐ 4 Gemcitabine**  **☐ 5 Bleomycin**  **☐ 6 Busulfan**  **☐ 7 Lomustin (CCNU)**  **☐ 8 Carmustin (BCNU)**  **☒ 9 Radiotherapy lung**  **☐ 10 Surgery**  **☐ 11 Combinations**  **☐ 12 Tobacco exposure** | **☐ Pulmonary diseases**  **☒ Pulmonary symptoms**  **☐ Pulmonary function test**  **☐ Absolute values**  **☒ Z-scores**  **☒ Percentage predicted**  **☐ Percentage pathological tests**  **(e.g. 24% with reduced FEV1)** | **☐ Longitudinal data available**  **☐ Control group mentioned**  **☐ Reference values stated**  **☐ Quality check performed**  **☒ Lung function procedure stated**  **☐ Cleaning of lung function data**  **described**  **☐ Person who analyzed PFT was**  **blinded to the exposure** |
| Centres:  Single center  Country:  USA, Department of Radiation Oncology at the Hospital of the University of Pennsylvania  Treatment era:  1988 – 2003  Years of Follow-up  median (range):  2.79 years (0–13.7) | Inclusion criteria:  - pediatric oncology patients  - whole lung irradiation  - pulmonary function testing during follow-up  Cancer diagnosis:  Wilms tumor (n=15), Hodgkin disease (n= 3), Sarcoma (n=11)  Hepatoblastoma (n=1)  Age at radiation  median (range):  7.6 (1.8 – 18)  Age at follow-up (most recent PFT)  median (range):  12.1 (range 5.3–19)  Time since diagnosis  Median (range) yr  NA | Name of protocol  not mentioned  Chemotherapy, doses, median (range):  Bleomycin: N=3 received Bleomycin; 20–40 U/m2  Radiotherapy, median (range):  Whole lung irradiation  1,200 cGy (1,050 - 1,760)  median fraction of 150cGy | How was outcome assessed?  Medical records and PFT database  Spirometry, body plethysmography, and diffusing capacity performed according to ATS protocols  Equipment used: Sensormedics 6200 Body Plethysmograph and Sensormedics Vmax22 spirometer and gas dilution system (Sensormedics,Yorba Linda, CA).  Definition of pulmonary function impairment:  normal: -2<Z<2  mildly reduced -4<Z<-2  moderately reduced -6<Z<-4  severely reduced Z<-6  Assessed values: FVC, FEV1, FEV1/FVC, TLC, DLCO, MIP (maximum inspiratory pressure), MEP (maximum expiratory pressure)  Incidence, Prevalence:  Pulmonary function Test Standard Deviation z-scores: Mean, Median, (range):  FVC (Z) n=30: -2.82, -2.37, (-13.3, 1.72)  FEV1 (Z) n=30: -2.47, -1.79, (-14.2, 0.72)  FEV1/FVC (%): 92, 93, (76, 100)  TLC (Z) n=23: -3.95, -3.25, (-25.3, 9.31)  DLCO (Z) n=21: -3.59, 3.32, (-10.32, 2.39)  MIP (%pred), n=23: 96.2, 88.8  MEP (%pred): 90.9, 92.3  FVC (N=30)  Normal n=14 (47%) ; mildly reduced n=10 (33%)  moderately reduced n=3 (10%) ; severely reduced n=3 (10%)  FEV1 (N=30)  Normal n=15 (50%) ; mildly reduced n=9 (30%)  moderately reduced n=3 (10%) ; severely reduced n=3 (10%)  TLC (N=23)  Normal n=9 (40%) ; mildly reduced n=4 (17%)  moderately reduced n=3 (13%) ; severely reduced n=7 (30%)  DLCO  Normal n=15 (50%) ; mildly reduced n=9 (30%)  moderately reduced n=3 (10%) ; severely reduced n=3 (10%)  Risk Factors  - Severity of the most recent pulmonary function abnormality (Z-scores for FEV1, TLC, and DLCO) did not correlate with:  age at the time of radiation (r^2^<0.001, r^2^=0.08, and r^2^=0.08 respectively)  total radiation dose (r^2^=0.002, r^2^=0.06, and r^2^=0.13, respectively)  - Severity of the most recent pulmonary function abnormality did not correlate with radiation dose/body length :  (r^2^=0.002, r^2^=0.027, and r^2^=0.03, respectively)  If longitudinal data available:  N=15 with two or more tests (too small to answer PICO) | Analysis:  - One-way analysis of variance: Differences in pulmonary function according to diagnostic groups  - Spearman correlation: correlation of pulmonary function abnormalities and continuous variables including radiation dose and age at radiation exposure  - Fisher exact test: presence of symptoms and pulmonary function  Limitation*:*  - Retrospective analysis  - Small heterogenous group  Strength:  Longitudinal data with Z-scores.    Potential bias/methodological problems:  Participation rate |

| **Main findings/message:**  **In survivors exposed to potential lung toxic treatment modalities according to COG guidelines, only irradiation of an increasing percentage of the lung with 10 Gy or more increases the risk for reduced FEV1, FVC, TLC, and DLCOcorr percentage predicted** | | | | |
| --- | --- | --- | --- | --- |
| D. M. Green, et al. Pulmonary Function after Treatment for Childhood Cancer. A Report from the St. Jude Lifetime Cohort Study (SJLIFE).  2016;13:1575-85. 10.1513/AnnalsATS.201601-022OC | | | | |
| Study design  Treatment era  Years of follow-up | Participants | Treatment  (= treatment analyzed in the paper) | Main outcomes | Additional remarks |
| **☒ Cohort**  **☐ Cross-sectional**  **☐ Case-control**  **☐ Other:**  **__________**  **☐ Retrospective**  **☒ Prospective** | **Study population (N)**  **Original cohort:** 4421  **Eligible cohort:** 989  **Analyzed cohort:** 606 (FEV1, FVC), 597 (TLC, DLco_corr_) | **☐ 1 HSCT**  **☐ 2 Cyclophosphamid**  **☐ 3 Methotrexate**  **☐ 4 Gemcitabine**  **☐ 5 Bleomycin**  **☐ 6 Busulfan**  **☐ 7 Lomustin (CCNU)**  **☐ 8 Carmustin (BCNU)**  **☒ 9 Radiotherapy lung**  **☐ 10 Surgery**  **☐ 11 Combinations**  **☐ 12 Tobacco exposure** | **☐ Pulmonary diseases**  **☐ Pulmonary symptoms**  **☒ Pulmonary function test**  **☐ Absolute values**  **☐ Z-scores**  **☒ Percentage predicted**  **☒ Percentage pathological tests**  **(e.g. 24% with reduced FEV1)** | **☐ Longitudinal data available**  **☐ Control group mentioned**  **☒ Reference values stated**  Hakinson, Goldman, Boren, Miller, GLI  **☒ Quality check performed**  **☒ Lung function procedure stated**: ATS  **☐ Cleaning of lung function data**  **described**  **☐ Person who analyzed PFT was**  **blinded to the exposure** |
| Centres:  Single centre, St Jude Lifetime Cohort  Country:  USA  Treatment era:  -  Years of Follow-up, median (range):  -  Time since diagnosis  median (range):  21.9 years | Inclusion criteria:  - >10 years from diagnosis  - age >=18 years  - pulmonary toxic treatment acc. to COG Guidelines (Bleomycin, Busulfan, CCNU, BCNU, Radiation therapy to the chest [including whole lung, mediastinum, axilla, mini-mantle, mantle, extended mantle, total lymphoid irradiation, subtotal lymphoid irradiation])  complete PFT  Cancer diagnosis:  ALL 5%  AML 3,5%  Other leukemia 3,5%  CNS tumor 1,3%  Hodgkin’s disease 49,3%  NHL 4%  Neuroblastoma 2,3%  Wilms tumor 5,8%  Osteosarcoma 5,8%  Ewing tumor 4,6%  Germ cell tumor 5,1%  Rhabdomyosarcoma 2,6%  Non rhabdomyosarcoma 2,6%  Other cancer 2,7%  Age at diagnosis  median (range): 13.0 years  Age at follow-up  median (range): 34.2 years | Name of protocol:  Not mentioned  Chemotherapy, %, doses  Cyclophosphamide: n=391 (64,5%)  Bleomycin: n=129 (21,3%)  Busulfan: n=16 (2,6%)  BCNU: n=11 (1,8%)  CCNU: n=12 (2%)  Chemotherapy, mean (SD)  Cyclophosphamide: 7,1g/m2 ( 5.3)  Bleomycin: 69.9mg/m2 (41.3)  Busulfan: 442.9 mg/m2 (406)  BCNU: 213.1 mg/m2 (159.99  CCNU: 459.7 mg/m2 (224.7)  Chemotherapy median (IQR)  Cyclophosphamide: 5.1g/m2 (3.6 – 9.2)  Bleomycin: 60.1mg/m2 (4.1 – 79.5)  Busulfan: 406.0mg/m2 (374.1 – 529.7)  BCNU: 147.6mg/m2 (100.0 – 300.0)  CCNU: 437.9mg/m2 (325.3 – 597.2)  Radiotherapy to chest, doses  n=450 (76,7%)  Lung radiation doses were estimated for the total lung and reported as the volume of lung receiving 10 Gy (V10), 20 Gy (V20), and 24 Gy (V24), reported as a percentage of the total lung volume  Mean (SD) proportions (%) of the lungs that received:  10 Gy: 0.58 (0.27)  20 Gy: 0.23 (0.19),  24 Gy: 0.15 (0.17)  Surgery: 19,7%  (Thoracotomy, rib resection, chest wall resection)  HSCT:  Allo: 6,6%, auto: 1,7%, both: 0,3% | How was outcome assessed?  Spirometry: FEV1, FVC, FEV1/FVC  Body plethysmography: TLC, DLcocorr  Definition of pulmonary function impairment:  - FEV1%predicted < 80% (GLI, race and sex specific by Wanger et al, 2005, EurRespirJ)  - FVC%predicted < 80% (GLI, race and sex specific by Wanger et al, 2005, EurRespirJ)  - TLC%predicted < 75% (sex specific equations (Goldman et al., 1959, AmRevTuberc, Boren et al1966, AmJMed)  - DLco_corr_ %predicted < 75% (sex specific equations Miller et al, 1983, AmRevRespirDis)  - FEV1/FVC: 0.8% percent predicted less than 0.7  Prevalence of abnormal parameters:  - FEV1: 50.7%  - FVC: 47.2%  - TLC: 31.2%  - DLco_corr:_ 44.6%  Risk factors (RR) *(here: parameters listed only for %predicted and not LLN)*:  FEV1%predicted <80%  V10 (per 10% increase) RR= 1.07 (1.04–1.09); p<0.001  FVC%predicted <80% and  V10 (per 10% increase) RR=1.08 (1.05–1.11); p<0.001  TLC% predicted <75% and  V10 (per 10% increase) 1.07 (1.01–1.13); p0.019  DLCOcorr%predicted <75% and  V10 (per 10% increase) 1.07 (1.04–1.10); p<0.001  In multivariable models selected by  BMA, all PFT results except TLC were  worse with increasing percentages of  the lungs that received 10 Gy or more  **Exact results available from publication, Table 3: “**Multivariable log-binominal regression models” | Analysis:  Relation between each PFT outcome and treatment modeled by using  multivariable log-binomial regression  Limitations:  No data on long-term outcome of these patients  Strength:  High participation rate  All tests performed with same standards  Prospective evaluation    Potential bias/methodological problems: |

| **Main findings/message:**  **Younger age at treatment is associated with an increased risk of developing pulmonary dysfunction (age categorical).** | | | | |
| --- | --- | --- | --- | --- |
| **F. Khan, et al.** Impact of Respiratory Developmental Stage on Sensitivity to Late Effects of Radiation in Pediatric Cancer Survivors. 2020 5, 426-433/Advances in Radiation Oncology | | | | |
| **Study design**  **Treatment era**  **Years of follow-up** | **Participants** | **Treatment** | **Main outcomes** | **Additional remarks** |
| ☒ **Cohort**  ☒ **Cross-sectional**: last PFT  ☐ **Case-control**  ☐ **Other:**  **__________**  ☒ **Retrospective**  ☐ **Prospective** | Study population (N)   - Original cohort: 136 with RT - Eligible cohort: 61 with PFT - Analysed cohort: 61 | ☐ 1 **HSCT**  ☐ **2 Cyclophosphamid**  ☐ **3 Methotrexate**  ☐ **4 Gemcitabine**  ☐ **5 Bleomycin**  ☐ **6 Busulfan**  ☐ **7 Lomustin (CCNU)**  ☐ **8 Carmustin (BCNU)**  ☒ **9 Radiotherapy lung**  ☐ **10 Surgery**  ☐ **11 Combinations**  ☐ **12 Tobacco exposure** | ☐ **Pulmonary diseases**  ☐ **Pulmonary symptoms**  ☒ **Pulmonary function test**  ☐ **Absolute values**  ☒ **Z-scores**  ☐ **Percentage predicted**  **☐ Percentage pathological tests**  (e.g. 24% with reduced FEV1) | ☐ **Longitudinal data available**  **☐ Control group mentioned**  **☒ Reference values stated**  Rosenthal  **☐ Quality check performed**  **☒ Lung function procedure stated:** ATS  **☐ Cleaning of lung function data**  **described**  **☐ Person who analyzed PFT was**  **blinded to the exposure** |
| Centres: single centre  Country: USA  Treatment era:  1995-2016  Years of Follow-up:  Mean 9 years (range, 1-20) | Study population:  Eligible (N): 61  Analysis (N): 61  Diagnoses:  All childhood cancer diagnoses  Age at radiation:  Mean 12.7 years (range 1.1-22.3)  Age at most recent PFT:  Mean 18.2 (range 7-27) | Name of protocol: NA  Chemotherapy: unknown except for bleomycin  Radiotherapy (doses):  Average dose 19.7±7.25 Gy (range 10.5-50.4Gy)  34% RT to whole lung (n=21)  66% partial RT to lung (n=40)  RT field included: thorax, upper and total abdomen, total body irradiation  Surgery: NA  HSCT:  Not stratified into auto/allo (n=17) | How was outcome assessed: Pulmonary function test (PFT) results; spirometry, body plethysmography, DLCO  Pulmonary function parameters normal if within 1.645 standard deviations above or below the mean predicted value.  Pulmonary outcomes:  - obstructive: FVC z-score >-1.645, FEV1 z-score <-1.645, FEV1/FVC ratio z-score <-1.645  - restrictive: TLC z-score <-1.645  - hyperinflation: RV/TLC ratio z-score >+1.645  - DLCO z-score <-1.645  Results  - Any abnormality: n=21  - Diffusion abnormality: n=12  - Restrictive abnormality: n=11  - Obstructive abnormality: n=5  Risk factor analysis for age at radiotherapy (multivariable logistic regression, crude model, OR (95%CI)):  - Any abnormality  - <5 years: 7.71 (1.17-51.06)  - >5 or <13 years: 3.51 (1.06-11.57)  - >13 years: 1.0 (ref)  - Diffusing abnormality  - <5 years: 3.75 (0.51-27.5)  - >5 or <13 years: 3.00 (0.73-12.27)  - >13 years: 1.0 (ref)  - Restrictive abnormality  - <5 years: 3.75 (0.51-27.50)  - >5 or <13 years: 2.34 (0.55-9.97)  - >13 years: 1.0 (ref)  - Obstructive abnormality  - <5 years: 3.20 (0.24-42.19)  - >5 or <13 years: 1.68 (0.22-12.96)  - >13 years: 1.0 (ref)  Risk factor analysis for age at radiotherapy (multivariable logistic regression, adjusted for time since treatment, OR (95%CI)):  - Any abnormality  - <5 years: 4.45 (0.38-51.79)  - >5 or <13 years: 3.09 (0.86-10.77)  - >13 years: 1.0 (ref)  - Diffusing abnormality  - <5 years: 4.27 (0.28-64.08)  - >5 or <13 years: 3.09(0.71-13.45)  - >13 years: 1.0 (ref)  - Restrictive abnormality  - <5 years: 2.22 (0.15-33.44)  - >5 or <13 years: 2.06 (0.45-9.51)  - >13 years: 1.0 (ref)  - Obstructive abnormality  - <5 years: 11.35 (0.20-634.6)  - >5 or <13 years: 2.10 (0.26-16.98)  - >13 years: 1.0 (ref)  Risk factor analysis for age at radiotherapy (multivariable logistic regression, adjusted for time since treatment and bleomycin exposure, OR (95%CI)):  - Any abnormality  - <5 years: 1.91 (0.13-29.04)  - >5 or <13 years: 1.63 (0.35-7.58)  - >13 years: 1.0 (ref)  - Diffusing abnormality  - <5 years: 3.64 (0.18-72.86)  - >5 or <13 years: 2.74 (0.46-16.18)  - >13 years: 1.0 (ref)  - Restrictive abnormality  - <5 years: 1.26 (0.06-25.63)  - >5 or <13 years: 1.30 (0.19-8.72)  - >13 years: 1.0 (ref)  - Obstructive abnormality  - <5 years: 6.57 (0.08-571.7)  - >5 or <13 years: 1.44 (0.11-19.21)  - >13 years: 1.0 (ref) | Analysis:  Risk-factor analysis for age at RT (<5 years, 5-13 years, >13 years) using multivariable regression model, crude model and model adjusted for time since treatment and additional bleomycin exposure  Limitations:  - retrospective design  - Few suervivors with pulmonary function abnormalities  Strength:  - z-scores  - use of ATS guidelines, single center (one PFT laboratory only)  - multivariate analysis  Potential bias/methodological problems:  - only 45% of initial cohort with PFT |

| **Main findings/message:**  **Radiotherapy: exposure to thoracic radiotherapy associated with significant higher odds for abnormal FVC and FEV1 and trend for TLC and DLCO (large CI).**  **Surgery: exposure to thoracic surgery associated with significant higher odds for abnormal FVC, FEV1 and TLC, and trend for DLCO (large CI).**  **Combinations (radio PLUS surgery versus no radio and surgery): exposure to combination associated with significant higher odds for abnormal FVC, FEV1, and TLC and trend for DLCO (large CI)**  **Smoking with trend to lower odds for FVC, FEV1, TLC and DLCO (large CI)** | | | | |
| --- | --- | --- | --- | --- |
| **A. Stone, et al.** Assessment of Pulmonary Outcomes, Exercise Capacity, and Longitudinal Changes in Lung Function in Pediatric Survivors of High-Risk Neuroblastoma. 2020, PBC, 2019 November ; 66(11): e27960. doi:10.1002/pbc.27960. | | | | |
| **Study design**  **Treatment era**  **Years of follow-up** | **Participants** | **Treatment** | **Main outcomes** | **Additional remarks** |
| ☒ **Cohort**  ☒ **Cross-sectional**:  ☐ **Case-control**  ☒ **Other:**  **longitudinal**  ☐ **Retrospective**  ☒ **Prospective** | Study population (N)   - Original cohort: NA - Eligible cohort: 62 - Analysed cohort: 62; 23 for longitudinal analysis (2 PFT) | ☐ 1 **HSCT**  ☐ **2 Cyclophosphamid**  ☐ **3 Methotrexate**  ☐ **4 Gemcitabine**  ☐ **5 Bleomycin**  ☐ **6 Busulfan**  ☐ **7 Lomustin (CCNU)**  ☐ **8 Carmustin (BCNU)**  ☒ **9 Radiotherapy lung**  ☒ **10 Surgery**  ☒ **11 Combinations**  ☒ **12 Tobacco exposure** | ☐ **Pulmonary diseases**  ☐ **Pulmonary symptoms**  ☒ **Pulmonary function test**  ☐ **Absolute values**  ☐ **Z-scores**  ☒ **Percentage predicted**  **☒ Percentage pathological tests**  (e.g. 24% with reduced FEV1) | ☐ **Longitudinal data available**  **☐ Control group mentioned**  **☒ Reference values stated**  NHANES III (Pellegrino)  **☐ Quality check performed**  **☒ Lung function procedure stated:** ATS  **☐ Cleaning of lung function data**  **described**  **☐ Person who analyzed PFT was**  **blinded to the exposure** |
| Centres: single centre, long-term follow-up clinic  Country: USA  Treatment era:  1996-2013  Years of Follow-up:  Median 5.29 years (range 0.24 – 15.24) | Study population:  Eligible (N): 62  Analysis (N): 62; 23 for longitudinal analysis (2 PFT)  Diagnoses:  High-risk neuroblastoma (stage 3 or 4)  Age at diagnosis:  Median 2.75 years (range 0.03 – 10.86)  Age at study:  Median 10.92 years (range 6.37 – 17.53) | Name of protocol: NA  Chemotherapy:  Cyclophosphamide 100%  Busulfan 6.5%  Radiotherapy (doses):  34% chest radiation therapy: radiation fields mentioned, no doses  Surgery:  23% thoracic surgery  HSCT:  50% received autologous HSCT | How was outcome assessed: Pulmonary function test (PFT) results; spirometry, body plethysmography, DLCO  Pulmonary outcomes:  Abnormalities in FVC and FEV1: (1) mild: 70 −79%pred; (2) moderate: 60–69%pred; (3) moderately severe: 50–59%pred; (4) severe: 35–49%pred; (5) very severe: <35%pred  Abnormalities TLC: (1) mild: 70–79%pred; (2) moderate: 60–69%pred; (3) severe: <60%pred  Abnormalities in DLCO: (1) mild: 61–79%pred; (2) moderate: 40–60%pred; (3) severe: <40%pred  Obstructive disease: FEV1/FVC<0.8  Restrictive disease: TLC<80 %pred  Results  - 77% with PFT abnormalities  - Restriction in 35%, obstruction in 6%, and mixed in 6%  - Decreased FVC in 53%, FEV1 in 47%, TLC in 42%, DLCO in 71%  - longitudinal analyses: 2^nd^ PFT median 2.97 years (range 1.07-5.55) after enrollment  - decline from t1 to t2 in FVC: 79.9%pred to 70.0%pred, p<0.05  - decline from t1 to t2 in FEV1: 81.6%pred to 69.9%pred, p<0.05  Results of univariable logistic analysis:  FVC (OR, 95%CI)  - Thoracic surgery yes/no and normal/abnormal parameter: 18.20 (2.20 – 150.58), p=0.001  - Radiotherapy yes/no and normal/abnormal parameter: 4.40 (1.34 – 14.51) p=0.010  - Thoracic surgery + radiotherapy yes/no and normal/abnormal parameter: 14.00 (1.68 – 116.85),  p=0.003  - Smoking yes/no and normal/abnormal parameter: 0.69 (0.19 – 2.53), p=0.569  FEV1 (OR, 95%CI)  - Thoracic surgery yes/no and normal/abnormal parameter: 10.94 (2.19 – 54.71), p=0.001  - Radiotherapy yes/no and normal/abnormal parameter: 4.29 (1.35 – 13.58), p=0.005  - Thoracic surgery + radiotherapy yes/no and normal/abnormal parameter: 19.56 (2.33 – 164.05), p=0.001  - Smoking yes/no and normal/abnormal parameter: 0.59 (0.16 – 2.28), p=0.446  TLC (OR, 95%CI)  - Thoracic surgery yes/no and normal/abnormal parameter: 3.28 (0.95 – 11.38), p=0.054  - Radiotherapy yes/no and normal/abnormal parameter: 4.33 (1.39 – 13.50), p=0.005  - Thoracic surgery + radiotherapy yes/no and normal/abnormal parameter: 5.82 (1.39 – 24.38), p=0.010  - Smoking yes/no and normal/abnormal parameter: 0.75 (0.20 – 2.90), p=0.748  DLCO (OR, 95%CI)  - Thoracic surgery yes/no and normal/abnormal parameter: 2.33 (0.45 – 12.09), p=0.475  - Radiotherapy yes/no and normal/abnormal parameter: 2.05 (0.49 – 8.62), p=0.339  - Thoracic surgery + radiotherapy yes/no and normal/abnormal parameter: 1.75 (0.33 – 9.31), p=0.70  - Smoking yes/no and normal/abnormal parameter: 0.39 (0.10 – 1.52), p=0.263 | Analysis:  Descriptive (t-test, chi-squared, Fisher’s exact test). Unadjusted logistic regression modelling.  Limitations:  - univariable analysis  - 37% with longitudinal assessment  - %predicted with fix cutoff values  - very large 95%CI for some outcomes  Strength:  - results of single PFT parameters reported  Potential bias/methodological problems:  - size of original cohort unclear |

| **Main findings/message:**  **Higher cumulative dose of bleomycin (>80mg/m2) is associated with reduced DLCO (<80% predicted; OR 2.12 (95%CI 0.99 – 4.49))** | | | | |
| --- | --- | --- | --- | --- |
| **A. Mittal, et al.** Late effects in pediatric Hodgkin lymphoma survivors after uniform treatmentwith ABVD with orwithout radiotherapy. 2021, PBC, March2021; DOI: 10.1002/pbc.29293 | | | | |
| **Study design**  **Treatment era**  **Years of follow-up** | **Participants** | **Treatment** | **Main outcomes** | **Additional remarks** |
| ☒ **Cohort**  ☐ **Cross-sectional**:  ☐ **Case-control**  ☐ **Other:**  **longitudinal**  ☐ **Retrospective**  ☒ **Prospective** | Study population (N)   - Original cohort: 223 - Eligible cohort: 154 - Analysed cohort: 125 with PFT - Analysed cohort: 119 with DLCO | ☐ 1 **HSCT**  ☐ **2 Cyclophosphamid**  ☐ **3 Methotrexate**  ☐ **4 Gemcitabine**  ☒ **5 Bleomycin**  ☐ **6 Busulfan**  ☐ **7 Lomustin (CCNU)**  ☐ **8 Carmustin (BCNU)**  ☐ **9 Radiotherapy lung**  ☐ **10 Surgery**  ☐ **11 Combinations**  ☐ **12 Tobacco exposure** | ☐ **Pulmonary diseases**  ☐ **Pulmonary symptoms**  ☒ **Pulmonary function test**  ☐ **Absolute values**  ☐ **Z-scores**  ☒ **Percentage predicted**  **☐ Percentage pathological tests**  (e.g. 24% with reduced FEV1) | ☐ **Longitudinal data available**  **☐ Control group mentioned**  **☒ Reference values stated**  Quanjer, Pellegrino  **☐ Quality check performed**  **☒ Lung function procedure stated:** ERS/ATS  **☐ Cleaning of lung function data**  **described**  **☐ Person who analyzed PFT was**  **blinded to the exposure** |
| Centres:  Single center  Country:  India  Treatment era:  2003 – 2013  Years of Follow-up:  Median 10.3yr (6.04-16.8) | Study population:  Eligible (N):  Analysis (N): 154  Diagnoses:  Hodgkin lymphoma  Age at diagnosis:  Median 10 years | Name of protocol:  ABVD (doxorubicin, bleomycin, vinblastine, dacarbazine)  Chemotherapy:  Bleomycin 100%  Radiotherapy (doses):  Radiotherapy total: 107 (69.5%)  Radiotherapy to mediastinum/chest: 12 (7.8%)  Radiotherapy to neck: 91 (59.1%)  Radiotehrapy to other sites: 7 (4.5%)  Surgery:  Not stated  HSCT:  Not stated | How was outcome assessed:  Spirometry (FEV1, FVC) best of three efforts, DLCO  Pulmonary outcomes:  FEV1: Normal (>80 % pred), mild decrease (70-79% pred), moderate decrease (60-69% pred), moderate severe decrease (50-59% pred), severe decrease (35-49% pred), very severe decrease (<35% pred)  FVC: normal (≥80 % pred), decreased (<80% pred)  Restrictive pattern: FVC<80%, FEV1/FVC≥85  Mixed pattern: FVC<80%, FEV1/FVC<85  DLCO: normal (>80 % pred), mild decrease (60-79% pred), moderate decr. (40-59% pred), severe decr. (<40% pred)  Results  - Restrictive: 36 (28.8%)  - Mixed pattern: 18 (14.4%)  - Moderate DLCO impairment: 7 (5.9%)  - Severe DLCO impairment: 1 (0.8%)  Risk factor analysis  Cumulative bleomycin dose ≤80mg/m2 vs >80mg/m2:  OR 2.12 (95%CI 0.99 – 4.49), p=0.051 | Analysis:  Multivariate analysis to estimate the effect of higher bleomyinc dose (>80mg/m2) on DLCO  Limitations:  - Impact of radiotherapy not taken into account  - only categories of percentage of predicted used  Strength:  - homogeneous cohort  - prospective design  Potential bias/methodological problems:  - 53% of cohort had DLCO assessed |
| **Main findings/message:**  **Exposure to radiotherapy was associated with a significantly lower FEV1 and FVC and a trend towards lower MME, TLC and DLCO in a cohort of transplanted survivors** | | | | |
| **M. Otth, et al.** Longitudinal lung function in childhood cancer survivors ater hematopoietic stem cell transplantation. 2021, Bone Marrow Transplantation, November 2021; DOI: 10.1038/s41409-021-01509-1 | | | | |
| **Study design**  **Treatment era**  **Years of follow-up** | **Participants** | **Treatment** | **Main outcomes** | **Additional remarks** |
| ☒ **Cohort**  ☐ **Cross-sectional**:  ☐ **Case-control**  ☒ **Other:**  **longitudinal**  ☒ **Retrospective**  ☐ **Prospective** | Study population (N)   - Original cohort: 142 - Eligible cohort: - Analysed cohort: 72 with 2 PFT of good quality | ☒ 1 **HSCT**  ☐ **2 Cyclophosphamid**  ☐ **3 Methotrexate**  ☐ **4 Gemcitabine**  ☐ **5 Bleomycin**  ☐ **6 Busulfan**  ☐ **7 Lomustin (CCNU)**  ☐ **8 Carmustin (BCNU)**  ☐ **9 Radiotherapy lung**  ☐ **10 Surgery**  ☐ **11 Combinations**  ☐ **12 Tobacco exposure** | ☐ **Pulmonary diseases**  ☐ **Pulmonary symptoms**  ☒ **Pulmonary function test**  ☐ **Absolute values**  ☒ **Z-scores**  ☐ **Percentage predicted**  **☐ Percentage pathological tests**  (e.g. 24% with reduced FEV1) | ☐ **Longitudinal data available**  **☐ Control group mentioned**  **☒ Reference values stated**  GLI 2021, Zapletal, ECCS  **☐ Quality check performed**  **☐ Lung function procedure stated:**  **☐ Cleaning of lung function data**  **described**  **☐ Person who analyzed PFT was**  **blinded to the exposure** |
| Centres:  Multicenter, national  Country:  Switzerland  Treatment era:  1976 – 2010  Years of Follow-up:  9.4 years (6.1 – 12.3) | Study population  Eligible (N): 142  Analysis (N): 72 with 2 PFT of good quality  Diagnoses:  Leukemia: 69%  Lymphoma: 16%  Other: 15%  Age at diagnosis:  7.4 years (3.5 – 12.2)  Age at last PFT:  16.2 years (14.2 – 20.0) | Name of protocol:  Not mentioned, different  Chemotherapy:  Busulfan: n=25; median 422mg/m2 (324-470)  Bleomycin: n=4; median 41mg/m2 (30-46)  Carmustine: n=5; median 300mg/m2 (300-300)  Lomustine: n=1; median 190mg/m2  Radiotherapy (doses):  RT to thorax, n=52 (70%)  Surgery:  Thoracic surgery: n=10 (14%)  HSCT:  Allogeneic: 50 (68%)  Autologous: 24 (32%) | How was outcome assessed:  Spirometry (FEV1, FVC, MMEF), body plethysmography (RV. TLC), DLCO  Pulmonary outcomes:  Pulmonary function test results: FEV1, FVC, MMEF, RV, TLC, DLCO  Results for radiotherapy (exposure yes/no) on intercept:  FEV1  - Coefficient -1.306; 95%CI -2.055 - -0.558; p=0.001  FVC  - Coefficient -1.473; 95%CI -2.207 - -0.739; p=<0.001  MMEF  - Coefficient -0.664; 95%CI -1.583 – 0.253; p=0.156  TLC  - Coefficient -0.717; 95%CI -2.051 – 0.616; p=0.292  RV  - Coefficient 0.663; 95%CI -0.307 – 1.634; p=0.181  DLCO  - Coefficient -1.279; 95%CI -2.773 - 0.213; p=0.093 | Analysis:  Mixed effect multivariable linear regression analysis with random intercept and slope  Limitations:  - retrospective  - Lung function procedures not stated  Strength:  - z-scores  - GLI 2021 references for FEV1, FVC, DLCO  Potential bias/methodological problems:  - 52% of initially eligible population with PFT results |

### K) Summary of the evidence assessments and quality of data contributing to the recommendations

**General comments/explanations**

Abbreviations used in the column “risk of bias”

- SB, selection bias
- AB, attrition bias
- DB, detection bias
- CF, confounding

Numbering used to assess quality of pulmonary function test results in the column “PFT quality”

1. Was control group mentioned?

**2. Were reference values stated?**

3. Was a quality check performed?

**4. Were lung function procedure stated?**

5. Was the cleaning of lung function data described?

6. Was the person who analysed PFT blinded to the exposure?

🡪 Focus was on point 2 and 4 when evaluating quality of PFT

🡪 Especially point 6 but also point 5 are not relevant when studies have been performed retrospectively/ with	existing data

🡪 Point 1 not relevant when reference equations have been used

References are added in **Appendix M)**

#### PICO 1: Allogeneic Hematopoietic Stem Cell Transplantation (HSCT)

| PICO | Study | | | No. of participants | | Follow-up (median/mean, range) yr | Allogeneic HSCT  n (%) | Pulmonary function Outcomes | Effect size | PFT quality | Risk of bias |
| --- | --- | --- | --- | --- | --- | --- | --- | --- | --- | --- | --- |
| 1 What is the risk of obstructive abnormalities in CAYA survivors treated with allogeneic HSCT compared to CAYA not treated with HSCT? | Record 2016 (1) | | | 143 CCS | | Mean 14.1 ± 4.8 (SD) | 67 (46.9%) | Obstructive  (FVC, FEV1, FEV1/FVC <80%pred or FEF25–75% <68%)  30% (20/67) HSCT  22% (17/76) no HSCT | Univariable comparison Chi2 HSCT Yes/No  0.30 | 1. No  2. Yes  Wang X, Pediatr Pulmonol 2005; Hankinson JL, Am J Respir Crit Care Med 1999  3. No  4. Yes  5. No  6. Yes | Retrospective cohort  SB: high risk  AB: low risk  DB: low risk  CF: high risk |
| GRADE assessment: | |  |  | |  | | | | | | |
| Study design: | |  | +4 | Retrospective cohort study | | | | | | | |
| Study limitations: | |  | -2 | Some limitations: Selection bias high in 1/1; Attrition bias low in 1/1; Detection bias low in 1/1; Confounding high in 1/1 | | | | | | | |
| Consistency: | |  | 0 | One study only | | | | | | | |
| Directness: | |  | -1 | Results are direct, population and outcomes broadly generalizable; PFT quality unsure (no control group mentioned, no quality checks performed, no cleaning of lung function data described) | | | | | | | |
| Precision: | |  | -2 | One study only, univariable comparison and no effect measure | | | | | | | |
| Publication bias: | |  | 0 | Unlikely | | | | | | | |
| Effect size: | |  | 0 | No effect measure, only univariable comparison | | | | | | | |
| Dose-response: | |  | 0 | Not applicable | | | | | | | |
| Plausible confounding: | |  | 0 | Only univariable comparison | | | | | | | |
| Quality of evidence: | | | ⊕⊖⊖⊖ VERY LOW | | | | | | | | |
| Conclusion: | | | No significant effect on obstructive abnormalities (FEV1, FEV1/FVC <80%pred or FEF25–75% <68%) in CAYA cancer survivors after allogeneic HSCT vs. no allogeneic HSCT.  (1 study; 1 non-significant effect; 143 participants; 37 obstructive) | | | | | | | | |
| Comment: | | | Only univariable comparison between CCS exposed to allogeneic HSCT and not exposed with Chi2 test and no effect measure. | | | | | | | | |

| PICO | Study | | | No. of participants | | Follow-up (median/mean, range) yr | Allogeneic HSCT  n (%) | Pulmonary function Outcomes | Effect size | PFT quality | Risk of bias |
| --- | --- | --- | --- | --- | --- | --- | --- | --- | --- | --- | --- |
| 1 What is the risk of restrictive abnormalities in CAYA survivors treated with allogeneic HSCT compared to CAYA not treated with HSCT? | Record 2016 (1) | | | 143 CCS | | Mean 14.1 ± 4.8 (SD) | 67 (46.9%) | Restrictive  (TLC<80% pred)  13% (9/67) HSCT  13% (10/76) no HSCT | Univariable comparison Chi2 HSCT Yes/No  0.96 | 1. No  2. Yes  Wang X, Pediatr Pulmonol 2005; Hankinson JL, Am J Respir Crit Care Med 1999  3. No  4. Yes  5. No  6. Yes | Retrospective cohort  SB: high risk  AB: low risk  DB: low risk  CF: high risk |
| GRADE assessment: | |  |  | |  | | | | | | |
| Study design: | |  | +4 | Retrospective cohort study | | | | | | | |
| Study limitations: | |  | -2 | Some limitations: Selection bias high in 1/1; Attrition bias low in 1/1; Detection bias low in 1/1; Confounding high in 1/1 | | | | | | | |
| Consistency: | |  | 0 | One study only | | | | | | | |
| Directness: | |  | -1 | Results are direct, population and outcomes broadly generalizable; PFT quality unsure (no control group mentioned, no quality checks performed, no cleaning of lung function data described) | | | | | | | |
| Precision: | |  | -2 | One study only, univariable comparison and no effect measure | | | | | | | |
| Publication bias: | |  | 0 | Unlikely | | | | | | | |
| Effect size: | |  | 0 | No effect measure, only univariable comparison | | | | | | | |
| Dose-response: | |  | 0 | Not applicable | | | | | | | |
| Plausible confounding: | |  | 0 | Only univariable comparison | | | | | | | |
| Quality of evidence: | | | ⊕⊖⊖⊖ VERY LOW | | | | | | | | |
| Conclusion: | | | No significant effect on restrictive abnormalities (TLC<80% pred) in CAYA cancer survivors after allogeneic HSCT vs. no allogeneic HSCT.  (1 study; 1 non-significant effect ; 143 participants; 19 restrictive) | | | | | | | | |
| Comment: | | | Only univariable comparison between CCS exposed to allogeneic HSCT and not exposed with Chi2 test and no effect measure. | | | | | | | | |

| PICO | Study | | | No. of participants | | Follow-up (median/mean, range) yr | Allogeneic HSCT  n (%) | Pulmonary function Outcomes | Effect size | PFT quality | Risk of bias |
| --- | --- | --- | --- | --- | --- | --- | --- | --- | --- | --- | --- |
| 1 What is the risk of hyperinflation in CAYA survivors treated with allogeneic HSCT compared to CAYA not treated with HSCT? | Record 2016 (1) | | | 143 CCS | | Mean 14.1 ± 4.8 (SD) | 67 (46.9%) | Hyperinflation  (RV >120%pred or RV/TLC >28%pred)  52% (35/67) HSCT  32% (24/76) no HSCT | Univariable comparison Chi2 HSCT Yes/No  0.01 | 1. No  2. Yes  Wang X, Pediatr Pulmonol 2005; Hankinson JL, Am J Respir Crit Care Med 1999  3. No  4. Yes  5. No  6. Yes | Retrospective cohort  SB: high risk  AB: low risk  DB: low risk  CF: high risk |
| GRADE assessment: | |  |  | |  | | | | | | |
| Study design: | |  | +4 | Retrospective cohort study | | | | | | | |
| Study limitations: | |  | -2 | Some limitations: Selection bias high in 1/1; Attrition bias low in 1/1; Detection bias low in 1/1; Confounding high in 1/1 | | | | | | | |
| Consistency: | |  | 0 | One study only | | | | | | | |
| Directness: | |  | -1 | Results are direct, population and outcomes broadly generalizable; PFT quality unsure (no control group mentioned, no quality checks performed, no cleaning of lung function data described) | | | | | | | |
| Precision: | |  | -2 | One study only, univariable comparison and no effect measure | | | | | | | |
| Publication bias: | |  | 0 | Unlikely | | | | | | | |
| Effect size: | |  | 0 | No effect measure, only univariable comparison | | | | | | | |
| Dose-response: | |  | 0 | Not applicable | | | | | | | |
| Plausible confounding: | |  | 0 | Only univariable comparison | | | | | | | |
| Quality of evidence: | | | ⊕⊖⊖⊖ VERY LOW | | | | | | | | |
| Conclusion: | | | Increased risk for hyperinflation (RV >120%pred or RV/TLC >28%pred) in CAYA cancer survivors after allogeneic HSCT vs. no allogeneic HSCT  (1 study; 1 significant effect; 143 participants; 59 hyperinflation) | | | | | | | | |
| Comment: | | | Only univariable comparison between CCS exposed to allogeneic HSCT and not exposed with Chi2 test and no effect measure. | | | | | | | | |

##### 1a Age at hematopoietic stem cell transplantation (HSCT)

| PICO | Study | | | No. of participants | Follow-up (median/mean, range) yr | Allogeneic HSCT  n (%) | Pulmonary function Outcomes | Effect size | PFT quality | Risk of bias |
| --- | --- | --- | --- | --- | --- | --- | --- | --- | --- | --- |
| 1a What is the risk of obstructive abnormalities in younger compared to older age at HSCT? | Inaba 2010 (2) | | | 89 CSS with hematological disease | Median 8.9 (range 1.7-16.4) | 89 (100%) | % of CCS below predicted values for FEF_25%-75%_  49% FEF_25%-75%_ (<67%pred) | Hazard Ratio  (p-value)  Older age at HSCT continuously, per year  1.082 (0.038) | 1. No  2. Yes  Hankinson JL, Am J Respir Crit Care Med, 1999  3. No  4. Yes: ATS  5. No  6. No | Prospective cohort  SB: High risk  AB: Low risk  DB: Low risk  CF: Unclear |
|  | Ginsberg, 2010 (3) | | | 317 CCS  (PFT post HSCT)  133 CCS  (PFT pre and post HSCT) | 0 - >5 years | 241 (76%)  Age at HSCT:  a. <7.8 yr  (n=77)  b. 7.8 – 11.4 yr  (n=79)  c. 11.4-14.6 yr  (n=79)  d. >14.6 yr  (n=79) | Z-score Mean (SD) for FEV1 at last post-transplant test  a. -1.270 (1.495)  b. -1.862 (1.469)  c. -1.730 (1.800)  d. -1.817 (1.936) | P-value of ANOVA  0.0790 | 1. No  2. Yes  Rosenthal M, Thorax, 1993, Hankinson JL, Am J Respir Crit Care Med, 1999  3. No  4. Yes: ATS  5. No  6. No | Retrospective cohort  SB: High risk  AB: Low risk  DB: Low risk  CF: Unclear |
| GRADE assessment: | |  |  |  | | | | | | |
| Study design: | |  | +4 | 1 prospective cohort study, 1 retrospective cohort study | | | | | | |
| Study limitations: | |  | -1 | Some limitations: Selection bias high in 2/2; Attrition bias low in 2/2; Detection bias low in 2/2; unclear in 2/2 | | | | | | |
| Consistency: | |  | 0 | No important inconsistency, both studies show increased risk with older age at HSCT, one p-value significant but hazard ration without confidence interval | | | | | | |
| Directness: | |  | 0 | Results are direct, population and outcomes broadly generalizable; PFT quality good (2/2 stated reference values and 2/2 the use if ATS guidelines) | | | | | | |
| Precision: | |  | -1 | Precision cannot be judged as 1/2 show results with p-value only and 1/2 shows results with Hazard rate but without 95%CI | | | | | | |
| Publication bias: | |  | 0 | Unlikely | | | | | | |
| Effect size: | |  | 0 | No large magnitude of effect | | | | | | |
| Dose-response: | |  | 0 | No clear age response relationship | | | | | | |
| Plausible confounding: | |  | 0 | No plausible confounding | | | | | | |
| Quality of evidence: | | | ⊕⊕⊖⊖ LOW | | | | | | | |
| Conclusion: | | | Increased risk for obstructive abnormalities (FEF_25%-75%_) in CAYA cancer survivors older vs. younger at allogeneic HSCT.  (2 studies; 1 significant effect [FEF_25%-75%_], 1 non-significant effect [FEV1]; 406 participants) | | | | | | | |
| Comment: | | | Only univariable comparison between CCS older vs. younger at allogeneic HSCT and effect measure in one study only. | | | | | | | |

| PICO | Study | | | | No. of participants | | Follow-up (median/mean, range) yr | Allogeneic HSCT  n (%) | Pulmonary function Outcomes | Effect size | PFT quality | Risk of bias |
| --- | --- | --- | --- | --- | --- | --- | --- | --- | --- | --- | --- | --- |
| 1a What is the risk of restrictive abnormalities in younger compared to older age at HSCT? | Ginsberg, 2010 (3) | | | | 317 CCS  (PFT post HSCT)  133 CCS  (PFT pre and post HSCT) | | 0 - >5 years | 241 (76%)  Age at transplant categorized:  a. <7.8 yr  (n=77)  b. 7.8 – 11.4 yr  (n=79)  c. 11.4-14.6 yr  (n=79)  d. >14.6 yr  (n=79) | Z-score Mean (SD) at last post-transplant test  **FVC**  a. -1.202 (1.234)  b. -1.707 (1.410)  c. -1.720 (1.668)  d. -17.796 (1.770)  **TLC**  a. - 0.587 (1.709)  b. - 1.041 (1.248)  c. - 0.812 (1.411)  d. - 0.836 (1.197) | P-value of ANOVA  0.0263  0.4319 | 1. No  2. Yes  Rosenthal M, Thorax, 1993, Hankinson JL, Am J Respir Crit Care Med, 1999  3. No  4. Yes: ATS  5. No  6. No | Retrospective cohort  SB: High risk  AB: Low risk  DB: Low risk  CF: Unclear |
|  | Wieringa 2005 (4) | | | | 39 CSS with hematological disease | | Median 4.5 years | 39 (100%)  Age at HSCT >10yr vs. <10yr | Higher **TLC** when older at HSCT *(no numbers stated)* | P-value from Student paired t-test,  >10yr vs. <10yr  0.08 | 1. Yes  2. Yes  Polgar G, Rev Resp Dis, 1979  3. No  4. No  5. No  6. No | Retrospective cohort  SB: High risk  AB: Low risk  DB: Low risk  CF: High risk |
| GRADE assessment: | |  |  | | |  | | | | | | |
| Study design: | |  | +4 | 2 retrospective cohort studies | | | | | | | | |
| Study limitations: | |  | -2 | Some limitations: Selection bias high in 2/2; Attrition bias low in 2/2; Detection bias low in 2/2; Confounding high in 1/2, unclear in 1/2 | | | | | | | | |
| Consistency: | |  | 0 | No important inconsistency. No significant effect on TLC in 2 studes and significant effect on FVC in 1 study | | | | | | | | |
| Directness: | |  | -1 | Population and outcomes broadly generalizable; PFT quality unsure (2/2 stated reference values and 1/2 the use if ATS guidelines) | | | | | | | | |
| Precision: | |  | -1 | Precision cannot be judged as 2/2 show results with p-value only | | | | | | | | |
| Publication bias: | |  | 0 | Unlikely | | | | | | | | |
| Effect size: | |  | 0 | No large magnitude of effect | | | | | | | | |
| Dose-response: | |  | 0 | No clear age response relationship | | | | | | | | |
| Plausible confounding: | |  | 0 | No plausible confounding | | | | | | | | |
| Quality of evidence: | | | ⊕⊖⊖⊖ VERY LOW | | | | | | | | | |
| Conclusion: | | | Inconsistent findings for restrictive abnormalities (TLC) in CAYA cancer survivors older vs. younger at allogeneic HSCT.  (2 studies; 1 significant effect [FVC], 2 non-significant effects [TLC]; 356 participants) | | | | | | | | | |
| Comment: | | | Only univariable comparison between CCS older vs. younger at allogeneic HSCT and no effect measure | | | | | | | | | |

| PICO | Study | No. of participants | Follow-up (median/mean, range) yr | Allogeneic HSCT  n (%) | Pulmonary function Outcomes | Effect size | PFT quality | Risk of bias |
| --- | --- | --- | --- | --- | --- | --- | --- | --- |
| 1a What is the risk of hyperinflation in younger compared to older age at HSCT? | | | | | | | | |

**No study**

| PICO | Study | | | No. of participants | | Follow-up (median/mean, range) yr | Allogeneic HSCT  n (%) | Pulmonary function Outcomes | Effect size | PFT quality | Risk of bias |
| --- | --- | --- | --- | --- | --- | --- | --- | --- | --- | --- | --- |
| 1a What is the risk of diffusion capacity impairment in younger compared to older age at treatment? | Inaba 2010 (2) | | | 89 CSS with hematological disease | | Median 8.9 (range 1.7-16.4) | 89 (100%) | % of CCS below predicted values for DLCO_corr_  64% DLCO_corr_ (<80%pred) | Hazard Ratio  (p-value)  Older age at HSCT continuously, per year  1.102 (0.005) | 1. No  2. Yes  Hankinson JL, Am J Respir Crit Care Med, 1999  3. No  4. Yes: ATS  5. No  6. No | Prospective cohort  SB: High risk  AB: Low risk  DB: Low risk  CF: Unclear |
|  | Ginsberg, 2010 (3) | | | 317 CCS  (PFT post HSCT)  133 CCS  (PFT pre and post HSCT) | | 0 - >5 years | 241 (76%)  Age at HSCT:  a. <7.8 yr  (n=77)  b. 7.8 – 11.4 yr  (n=79)  c. 11.4-14.6 yr  (n=79)  d. >14.6 yr  (n=79) | Z-score Mean (SD) for DLCO at last post-transplant test  a. -1.649 (1.830)  b. -1.889 (1.531)  c. -1.791 (1.665)  d. -2.182 (1.341) | P-value of ANOVA  0.432 | 1. No  2. Yes  Rosenthal M, Thorax, 1993, Hankinson JL, Am J Respir Crit Care Med, 1999  3. No  4. Yes: ATS  5. No  6. No | Retrospective cohort  SB: High risk  AB: Low risk  DB: Low risk  CF: Unclear |
|  | Leung 2007 (5) | | | 155 CCS | | Median 9 (range 3.1-15.9) | 155 (100%) | 35% DLCO (<80%pred) | Hazard Ratio (95%CI)  Older age at HSCT continuously, per year  1.1 (1.04-1.17) | 1. No  2. No  3. No  4. No  5. No  6. No | Prospective cohort  SB: Low risk  AB: Low risk  DB: Low risk  CF: Low risk |
| GRADE assessment: | |  |  | |  | | | | | | |
| Study design: | |  | +4 | 2 prospective cohort studies, 1 retrospective cohort study | | | | | | | |
| Study limitations: | |  | -1 | Some limitations: Selection bias high in 2/3, low in 1/3; Attrition bias low in 3/3; Detection bias low in 3/3; Confounding unclear in 2/3, low in 1/3 | | | | | | | |
| Consistency: | |  | 0 | No important inconsistency, two studies show increased risk with older age at HSCT, 2/3 showed significant effect | | | | | | | |
| Directness: | |  | -1 | Population and outcomes broadly generalizable; PFT quality unsure (not homogeneous across studies, 2/3 stated reference values and 2/3 the use if ATS guidelines) | | | | | | | |
| Precision: | |  | -1 | Moderate imprecision, 1/3 show precise results with small confidence interval, in 2/3 precision cannot be judged results shown as p-value only | | | | | | | |
| Publication bias: | |  | 0 | Unlikely | | | | | | | |
| Effect size: | |  | 0 | No large magnitude of effect | | | | | | | |
| Dose-response: | |  | 0 | No clear age response relationship | | | | | | | |
| Plausible confounding: | |  | 0 | No plausible confounding | | | | | | | |
| Quality of evidence: | | | ⊕⊖⊖⊖ VERY LOW | | | | | | | | |
| Conclusion: | | | Increased risk for diffusion impairment (DLCO) in CAYA cancer survivors older vs. younger at allogeneic HSCT.  (3 studies; 2 significant effects, 1 non-significant effect; 561 participants) | | | | | | | | |
| Comment: | | | One study with high precision, in two studies precision cannot be judged | | | | | | | | |

##### 1b Chronic Graft versus Host Disease (cGvHD)

| PICO | Study | | | | No. of participants | | Follow-up (median/mean, range) yr | Allogeneic HSCT | Pulmonary function Outcomes | Effect size | PFT quality | Risk of bias |
| --- | --- | --- | --- | --- | --- | --- | --- | --- | --- | --- | --- | --- |
| 1b What is the risk of obstructive abnormalities in patients with cGvHD compared to patients without cGvHD? | Madanat-Harjuoja, 2014 (6) | | | | 51 CSS with hematological disease | | Median 4.1 years  Chronic GvHD:  - 55% No  - 22% limited  - 23% extensive  Acute GvHD:  - 43% No/Grade I  - 57% Grade II-IV | 51 (100%)  Anlyses:  a. No vs limited  cGvHD  b. No vs  extensive  cGvHD  c. No/Grade I vs  Grade II-IV  aGvHD | FEV1  FEV1/FVC | Random effect modeling for longitudinal analysis  (estimates of coefficient, p-value)  a: 8.0871 (0.314)  b: - 27.8368 (0.003)  c:- 13.8726 (0.015)  a: 0.0292 (0.582)  b:- 0.1366 (0.026)  c: - 0.0081 (0.830) | 1. No  2. Yes  Quanjer PH, Eur Respir J Suppl,, 1997  3. Yes  4. Yes: ATS  5. Yes  6. No | Retrospective cohort  SB: High risk  AB: Low risk  DB: Low risk  CF: Low risk |
|  | Hoffmeister, 2006 (7) | | | | 215 CSS with hematological disease | | Median 10.5 (range 5-27.5) | 202 (94%)  cGvHD:  Yes: n=71  No: n=144 | Total 26 **Obstructive**  FEv1/FVC<80%, FEV1<100%pred  20% with cGvHD  8% without GvHD | Odds Ratio (95%CI)  cGVHD yes/no  Multivariable analysis 4.4 (1.6-12) | 1. No  2. Yes  Rosenthal M, Thorax, 1993; Crapo RO, Am Rev Respir Dis, 1981; Crapo RO, Bulletin Europeen de Physiopathologie Respiratoire, 1982  3. No  4. Yes: ATS  5. No  6. No | Retrospective cross-sectional  SB: High risk  AB: Low risk  DB: Low risk  CF: High risk |
| GRADE assessment: | |  |  | | |  | | | | | | |
| Study design: | |  | +4 | 1 retrospective cohort study, 1 retrospective cross-sectional study | | | | | | | | |
| Study limitations: | |  | -2 | Some limitations: Selection bias high in 2/2; Attrition bias low in 2/2; Detection bias low in 2/2; Confounding high in 1/2, low in 1/2 | | | | | | | | |
| Consistency: | |  | 0 | No important inconsistency; both studies show increased risk of obstructive abnormalities with development of cGvHD; one study showed significant effect of aGvHD on obstructive abnormalities | | | | | | | | |
| Directness: | |  | 0 | Population and outcomes broadly generalizable; PFT quality good (2/2 stated reference values and 2/2 the use if ATS guidelines) | | | | | | | | |
| Precision: | |  | -1 | Important imprecision, 1/2 with results from multivariable analysis but with large confidence interval, 1/2 precision cannot be judged as results shown as p-value only | | | | | | | | |
| Publication bias: | |  | 0 | Unlikely | | | | | | | | |
| Effect size: | |  | 0 | No large magnitude of effect | | | | | | | | |
| Dose-response: | |  | 0 | Not applicable | | | | | | | | |
| Plausible confounding: | |  | 0 | No plausible confounding | | | | | | | | |
| Quality of evidence: | | | ⊕⊖⊖⊖ VERY LOW | | | | | | | | | |
| Conclusion: | | | Increased risk for obstructive abnormalities (FEV1, FEV1/FVC) in CAYA cancer survivors after chronic GvHD vs. no GvHD, especially extensive cGvHD.  Increased risk for obstructive abnormalities (FEV1, FEV1/FVC) in CAYA cancer survivors after acute GvHD Grade II-IV vs. no GvHD/Grade I (onse study).  (2 studies; 2 significant effects, 266 participants) | | | | | | | | | |
| Comments: | | | Two studies with important imprecision | | | | | | | | | |

| PICO | Study | | | | No. of participants | Follow-up (median/mean, range) yr | Allogeneic HSCT | Pulmonary function Outcomes | Effect size | PFT quality | Risk of bias |
| --- | --- | --- | --- | --- | --- | --- | --- | --- | --- | --- | --- |
| 1b What is the risk of restrictive abnormalities in patients with cGvHD compared to patients without cGvHD? | Madanat- Harjuoja, 2014 (6) | | | | 51 CSS with hematological disease | Median 4.1 years  Chronic GvHD:  - 55% No  - 22% limited  - 23% extensive  Acute GvHD:  - 43% No/Grade I  - 57% Grade II-IV | 51 (100%)  Anlyses performed:  a. No vs limited  cGvHD  b. No vs  extensive  cGvHD  c. No/Grade I vs  Grade II-IV  aGvHD | **FVC** | Random effect modeling for longitudinal analysis  (estimates of coefficient, p-value)  a: 7.5973 (0.243)  b: - 18.90747 (0.012)  c: - 13.1761 (0.004) | 1. No  2. Yes  Quanjer PH, Eur Respir J Suppl,, 1997  3. Yes  4. Yes: ATS  5. Yes  6. No | Retrospective cohort  SB: High risk  AB: Low risk  DB: Low risk  CF: Low risk |
| GRADE assessment: | |  |  |  | | | | | | | |
| Study design: | |  | +4 | 1 retrospective cohort study | | | | | | | |
| Study limitations: | |  | -1 | Some limitations: Selection bias high in 1/1; Attrition bias low in 1/1; Detection bias low in 1/1; Confounding low in 1/1 | | | | | | | |
| Consistency: | |  | 0 | One study only | | | | | | | |
| Directness: | |  | 0 | Population and outcomes broadly generalizable; PFT quality good (2/2 stated reference values and 2/2 the use if ATS guidelines) | | | | | | | |
| Precision: | |  | -1 | Some imprecision, only 1 study, no effect size not shown | | | | | | | |
| Publication bias: | |  | 0 | Unlikely | | | | | | | |
| Effect size: | |  | 0 | No large magnitude of effect | | | | | | | |
| Dose-response: | |  | 0 | Not applicable | | | | | | | |
| Plausible confounding: | |  | 0 | No plausible confounding | | | | | | | |
| Quality of evidence: | | | ⊕⊖⊖⊖ VERY LOW | | | | | | | | |
| Conclusion: | | | Increased risk of restrictive abnormalities (FVC) in CAYA cancer survivors after extensive chronic GvHD and acute GvHD Grade II-IV vs. no GvHD.  (1 study; 1 significant effect; 51 participants) | | | | | | | | |
| Comments: | | | One study only with some limitations | | | | | | | | |

| PICO | Study | | | | No. of participants | | Follow-up (median/mean, range) yr | Allogeneic HSCT | Pulmonary function Outcomes | Effect size | PFT quality | Risk of bias |
| --- | --- | --- | --- | --- | --- | --- | --- | --- | --- | --- | --- | --- |
| 1b What is the risk of diffusion capacity impairment in patients with cGvHD compared to patients without cGvHD? | Leung 2007 (5) | | | | 155 CCS | | Median 9 (range 3.1-15.9) | 155 (100%)  cGvHD (26%)  No cGvHD (74%) | 34% DLCO (<80%pred) | Hazard Ratio (95%CI)  1.96 (1.12-3.44) | 1. No  2. No  3. No  4. No  5. No  6. No | Prospective cohort  SB: Low risk  AB: Low risk  DB: Low risk  CF: Low risk |
| GRADE assessment: | |  |  | | |  | | | | | | |
| Study design: | |  | +4 | 1 prospective cohort study | | | | | | | | |
| Study limitations: | |  | 0 | No limitations: Selection bias low in 1/1; Attrition bias low in 1/1; Detection bias low in 1/1; Confounding low in 1/1 | | | | | | | | |
| Consistency: | |  | NA | One study only | | | | | | | | |
| Directness: | |  | -1 | No statement on generalizability possible (no reference values and no use of ATS guidelines stated) | | | | | | | | |
| Precision: | |  | -1 | Some imprecision, only 1 study but precise results with small confidence interval | | | | | | | | |
| Publication bias: | |  | 0 | Unlikely | | | | | | | | |
| Effect size: | |  | 0 | No large magnitude of effect | | | | | | | | |
| Dose-response: | |  | 0 | Not applicable | | | | | | | | |
| Plausible confounding: | |  | 0 | No plausible confounding | | | | | | | | |
| Quality of evidence: | | | ⊕⊕⊖⊖ LOW | | | | | | | | | |
| Conclusion: | | | Increased risk for diffusion capacity impairment (DLCO) in CAYA cancer survivors with chronic GvHD vs. no cGvHD.  (1 study; 1 significant effect, 155 participants) | | | | | | | | | |
| Comments: | | | One study only with some limitations | | | | | | | | | |

##### 1c Infection during hematopoietic stem cell transplantation

| PICO | Study | No. of participants | Follow-up (median/mean, range) yr | Allogeneic HSCT | Pulmonary function Outcomes | Effect size | Risk of bias |
| --- | --- | --- | --- | --- | --- | --- | --- |
| 1c What is the risk in patients who had a pulmonary infection during HSCT compared to patients without pulmonary infection during HSCT? | | | | | | | |

**No study**

##### 1d Total body irradiation (TBI) as conditioning for hematopoietic stem cell transplantation (HSCT)

| PICO | Study | | | No. of participants | | Follow-up (median/mean, range) yr | Allogeneic HSCT  N (%) | Pulmonary function Outcomes | Effect size | PFT quality | Risk of bias |
| --- | --- | --- | --- | --- | --- | --- | --- | --- | --- | --- | --- |
| 1d What is the risk of obstructive abnormalities for patients treated with total body irradiation as conditioning for HSCT? | Leung 2007 (5) | | | 155 CCS | | Median 9 (range 3.1-15.9) | Allogeneic:  155 (100%)  TBI: 123 (85%) | Number of CCS with respective parameter below predicted values  41 FEV1/FVC (<85%pred) | Hazard Ratio (95%CI)  2.39 (1.10-5.74) | 1. No  2. No  3. No  4. No  5. No  6. No | Prospective cohort  SB: Low risk  AB: Low risk  DB: Low risk  CF: Low risk |
|  | Hoffmeister, 2006 (7) | | | 215 CSS with hematological disease | | Median 10.5 (range 5-27.5)  No TBI: n=53  FTBI: n=133  1.2Gy: n=37  >1.2Gy: n=96  SFTBI: n=29 | Allogeneic:  202 (94%)  Analyses performed:  a. No TBI vs  FTBI 1.2Gy  b. No TBI vs  FTBI 2.0-2.25  Gy  c. No TBI vs  SFTBI | Total 26 obstructive  FEv1/FVC<80%, FEV1<100%pred  3% (1/37): FTBI 1.2Gy  13% (12/96): FTBI 2.0-2.25Gy  7% (2/29): SFTBI | Multivariate Analysis Odds Ratio (95%CI)  a. 0.1 (0.0-1.4)  b. 0.9 (0.3-2.8)  c. 0.1 (0.0-0.5) | 1. No  2. Yes:  Rosenthal M, Thorax, 1993; Crapo RO, Am Rev Respir Dis, 1981; Crapo RO, Bulletin Europeen de Physiopathologie Respiratoire, 1982  3. No  4. Yes: ATS  5. No  6. No | Retrospective cross-sectional  SB: High risk  AB: Low risk  DB: Low risk  CF: High risk |
| GRADE assessment: | |  |  | |  | | | | | | |
| Study design: | |  | +4 | 1 prospective cohort study, 1 retrospective cross-sectional study | | | | | | | |
| Study limitations: | |  | -2 | Some limitations: Selection bias high in 1/2, low in 1/2; Attrition bias low in 2/2; Detection bias low in 2/2; Confounding high in 1/2, low in 1/2 | | | | | | | |
| Consistency: | |  | 0 | No important inconsistency. One study reports significant reduction in FEV1/FVC in CCS exposed to TBI. One study compares different TBI regimens to non-TBI with no significant association. | | | | | | | |
| Directness: | |  | -1 | Population and outcomes broadly generalizable; PFT quality unsure (not homogeneous, 1/2 stated reference values and 1/2 the use if ATS guidelines) | | | | | | | |
| Precision: | |  | 0 | No imprecision, 2/2 show precise results with small confidence interval. | | | | | | | |
| Publication bias: | |  | 0 | Unlikely | | | | | | | |
| Effect size: | |  | 1 | Large magnitude of effect in one study | | | | | | | |
| Dose-response: | |  | 0 | Not applicable | | | | | | | |
| Plausible confounding: | |  | 0 | No plausible confounding | | | | | | | |
| Quality of evidence: | | | ⊕⊖⊖⊖ VERY LOW | | | | | | | | |
| Conclusion: | | | Inconsistent findings. Increased risk of obstructive abnormalities (FEV1/FVC) in CAYA cancer survivors after TBI vs. no TBI (one study only, fractioning not mentioned). No significant effect of different fractioning (one study).  (2 studies; 2 significant effect [TBI yes/no], 1 non-significant effect [different fractioning of TBI]; 370 participants) | | | | | | | | |
| Comments: | | | Two studies with some limitations but high precision | | | | | | | | |

| PICO | Study | | | No. of participants | Follow-up (median/mean, range) yr | Allogeneic HSCT  N (%) | Pulmonary function Outcomes | Effect size | PFT quality | Risk of bias |
| --- | --- | --- | --- | --- | --- | --- | --- | --- | --- | --- |
| 1d What is the risk of restrictive abnormalities for patients treated with total body irradiation as conditioning for HSCT? | Leung 2007 (5) | | | 155 CCS | Median 9 (range 3.1-15.9) | Allogeneic:  155 (100%)  TBI: 123 (85%) | Number of CCS with respective parameter below predicted values  48 **TLC** (<80%pred) | Hazard Ratio (95%CI)  2.26 (1.04-4.95) | 1. No  2. No  3. No  4. No  5. No  6. No | Prospective cohort  SB: Low risk  AB: Low risk  DB: Low risk  CF: Low risk |
|  | Hoffmeister, 2006 (7) | | | 215 CSS with hematological disease | Median 10.5 (range 5-27.5)  No TBI: n=53  FTBI: n=133  1.2Gy: n=37  >1.2Gy: n=96  SFTBI: n=29 | Allogeneic:  202 (94%)  Analyses performed:  a. No TBI vs FTBI  1.2Gy  b. No TBI vs FTBI  2.0-2.25 Gy  c. No TBI vs  SFTBI | Total 67 **restrictive**  TLC <80%pred  19% (7/37) when FTBI 1.2Gy  31% (30/96) when FTBI 2.0-2.25Gy  72% (21/29) when SFTBI | Multivariate Analysis Odds Ratio (95%CI)  a. 2.5 (0.4-16)  b. 2.8 (0.6-13)  c. 22.0 (3.9-120) | 1. No  2. Yes:  Rosenthal M, Thorax, 1993; Crapo RO, Am Rev Respir Dis, 1981; Crapo RO, Bulletin Europeen de Physiopathologie Respiratoire, 1982  3. No  4. Yes: ATS  5. No  6. No | Retrospective cross-sectional  SB: High risk  AB: Low risk  DB: Low risk  CF: High risk |
| GRADE assessment: | |  |  | |  | | | | | |
| Study design: | |  | +4 | 1 prospective cohort study, 1 retrospective cross-sectional study | | | | | | |
| Study limitations: | |  | -2 | Some limitations: Selection bias high in 1/2, low in 1/2; Attrition bias low in 2/2; Detection bias low in 2/2; Confounding high in 1/2, low in 1/2 | | | | | | |
| Consistency: | |  | 0 | No inconsistency, both studies report reduction in TLC in CAYA cancer survivors exposed to TBI. | | | | | | |
| Directness: | |  | -1 | Population and outcomes broadly generalizable; PFT quality unsure (1/2 stated reference values and 1/2 the use if ATS guidelines) | | | | | | |
| Precision: | |  | -1 | Important imprecision, both studies show precise results, but the 95%CI but is large in one study | | | | | | |
| Publication bias: | |  | 0 | Unlikely | | | | | | |
| Effect size: | |  | 1 | Large magnitude of effect on one study | | | | | | |
| Dose-response: | |  | 0 | Not applicable | | | | | | |
| Plausible confounding: | |  | 0 | No plausible confounding | | | | | | |
| Quality of evidence: | | | ⊕⊖⊖⊖ VERY LOW | | | | | | | |
| Conclusion: | | | Increased risk for restrictive abnormalities (TLC) in CAYA cancer survivors after TBI as conditioning for HSCT vs. no TBI. No significant effect of different fractioning (one study).  (2 studies; 2 significant effects [TBI yes/no], 1 non-significant effect [different fractioning of TBI]; 370 participants) | | | | | | | |
| Comments: | | | Results of one study with large confidence intervals. | | | | | | | |

| PICO | Study | | | No. of participants | | Follow-up (median/mean, range) yr | Allogeneic HSCT  N (%) | Pulmonary function Outcomes | Effect size | PFT quality | Risk of bias |
| --- | --- | --- | --- | --- | --- | --- | --- | --- | --- | --- | --- |
| 1d What is the risk of diffusion capacity impairment for patients treated with total body irradiation as conditioning for HSCT? | Leung 2007 (5) | | | 155 CCS | | Median 9 (range 3.1-15.9) | Allogeneic:  155 (100%)  TBI: 123 (85%) | Number of CCS with respective parameter below predicted values  52/155 DLCO (<80%pred) | Hazard Ratio (95%CI)  2.24 (1.07-5.09) | 1. No  2. No  3. No  4. No  5. No  6. No | Prospective cohort  SB: Low risk  AB: Low risk  DB: Low risk  CF: Low risk |
| GRADE assessment: | |  |  | |  | | | | | | |
| Study design: | |  | +4 | 1 prospective cohort study | | | | | | | |
| Study limitations: | |  | 0 | No limitations: Selection bias low in 1/1; Attrition bias low in 1/1; Detection bias low in 1/1; Confounding low in 1/1 | | | | | | | |
| Consistency: | |  | NA | One study only | | | | | | | |
| Directness: | |  | -1 | No information on generalizability; PFT quality unsure (no reference values and no information on guidelines stated) | | | | | | | |
| Precision: | |  | -1 | Some imprecision, only 1 study but precise results with small confidence interval | | | | | | | |
| Publication bias: | |  | 0 | Unlikely | | | | | | | |
| Effect size: | |  | 1 | Large magnitude of effect | | | | | | | |
| Dose-response: | |  | 0 | Not applicable | | | | | | | |
| Plausible confounding: | |  | 0 | No plausible confounding | | | | | | | |
| Quality of evidence: | | | ⊕⊕⊕⊖ MODERATE | | | | | | | | |
| Conclusion: | | | Increased risk for diffusion capacity impairment (DLCO) in CAYA cancer survivors after TBI as conditioning for HSCT vs. no TBI.  (1 study; 1 significant effect, 155 participants) | | | | | | | | |
| Comments: | | | One study with high precision | | | | | | | | |

#### PICO 2: Cyclophosphamide (CYC)

| PICO | Study | | | No. of participants | Follow-up (median/mean, range) yr | Cyclophosphamide  (CYC) | Pulmonary function Outcomes | Effect size | PFT quality | Risk of bias |
| --- | --- | --- | --- | --- | --- | --- | --- | --- | --- | --- |
| 2 What is the risk of obstructive abnormalities in CAYA treated with CYC compared to CAYA not treated with CYC? | Jenney 1995 (8) | | | 70 leukemia CCS | Median 4.2 (range 0.6-18.5) | Proportion receiving CYC unclear | Number of CCS with FEV1 below predicted values  36/69 FEV1 (<85% pred)  23/69 FEV1 (<80% pred) | p-value  CYC yes vs no  p<0.001 | 1. Yes  2. No  3. No  4. No  5. No  6. No | Prospective cross-sectional  SB: high risk  AB: low risk  DB: unclear  CF: unclear |
| GRADE assessment: | |  |  |  | | | | | | |
| Study design: | |  | +4 | 1 retrospective cross-sectional study | | | | | | |
| Study limitations: | |  | -2 | Some limitations: Selection bias high in 1/1; Attrition bias low in 1/1; Detection bias unclear in 1/1; Confounding unclear in 1/1 | | | | | | |
| Consistency: | |  | NA | One study only | | | | | | |
| Directness: | |  | -1 | Population and outcomes broadly generalizable; PFT quality unsure (no reference values and guidelines stated) | | | | | | |
| Precision: | |  | -1 | Important imprecision, precision cannot be judged as 1/1 shows p-value only | | | | | | |
| Publication bias: | |  | 0 | Unlikely | | | | | | |
| Effect size: | |  | 0 | No large magnitude of effect | | | | | | |
| Dose-response: | |  | NA | Not applicable | | | | | | |
| Plausible confounding: | |  | 0 | No plausible confounding | | | | | | |
| Quality of evidence: | | | ⊕⊖⊖⊖ VERY LOW | | | | | | | |
| Conclusion: | | | Increased risk for obstructive abnormalities (FEV1) in CAYA cancer survivors after cyclophosphamide vs. no cyclophosphamide  (1 study; 1 significant effect; 70 participants) | | | | | | | |
| Comments: | | | One study and precision cannot be judged as result is shown as p-value only | | | | | | | |

| PICO | Study | | | | No. of participants | | Follow-up (median/mean, range) yr | Cyclophosphamide  (CYC) | Pulmonary function Outcomes | Effect size | PFT quality | Risk of bias |
| --- | --- | --- | --- | --- | --- | --- | --- | --- | --- | --- | --- | --- |
| 2 What is the risk of restrictive abnormalities in CAYA treated with CYC compared to CAYA not treated with CYC? | Jenney 1995 (8) | | | | 70 leukemia CCS | | Median 4.2 (range 0.6-18.5) | Proportion receiving CYC unclear | Number of CCS with respective parameter below predicted values  32/69 FVC (<85% pred)  20/69 FVC (<80% pred)  26/69 TLC (<85% pred)  20/69 TLC (<80% pred) | CYC leads to reduction in FVC, and TLC:  p<0.001  p<0.001 | 1. Yes  2. No  3. No  4. No  5. No  6. No | Prospective cross-sectional  SB: high risk  AB: low risk  DB: unclear  CF: unclear |
|  | Mulder 2011 (9) | | | | 193 CCS | | Median 17.9 (range 5.6-36.8) | High-dose CYC  43 (22.3%) | 34/193 Restrictive disease  (TLC or FVC <75% pred) | Odds Ratio (95%CI)  High-dose CYC vs no  2.15 (0.80-5.79) | 1. No  2. No  3. No  4. No  5. No  6. No | Retrospective cohort  SB: Low risk  AB: Low risk  DB: Low risk  CF: Low risk |
| GRADE assessment: | |  |  | | |  | | | | | | |
| Study design: | |  | +4 | 1 retrospective cohort study, 1 prospective cross-sectional study | | | | | | | | |
| Study limitations: | |  | -2 | Some limitations: Selection bias high in 1/2, low in 1/2; Attrition bias low in 2/2; Detection bias low in 1/2, unclear 1/2; Confounding low in 1/2, unclear 1/2 | | | | | | | | |
| Consistency: | |  | 0 | No inconsistency. Both studies show more restrictive abnormalities in CCS exposed to CYC, but in one study this effect was not significant | | | | | | | | |
| Directness: | |  | -1 | Population and outcomes broadly generalizable; PFT quality unsure (both studies do not report reference values and guidelines used) | | | | | | | | |
| Precision: | |  | -1 | Important imprecision, 1/2 show precise results with small confidence interval, in 1/2 precision cannot be judged as p-value is shown only | | | | | | | | |
| Publication bias: | |  | 0 | Unlikely | | | | | | | | |
| Effect size: | |  | 0 | No large magnitude of effect | | | | | | | | |
| Dose-response: | |  | 0 | Not applicable | | | | | | | | |
| Plausible confounding: | |  | 0 | No plausible confounding | | | | | | | | |
| Quality of evidence: | | | ⊕⊖⊖⊖ VERY LOW | | | | | | | | | |
| Conclusion: | | | Increased risk for restrictive abnormalities (FVC, TLC) in CAYA cancer survivors after cyclophosphamide vs. no cyclophosphamide  (2 studies; 1 significant effects [FVC, TLC], 1 non-significant effects [FVC]; 263 participants) | | | | | | | | | |
| Comments: | | | One study shows significant p-value only, one study shows effect measure by OR but with non-significant 95%CI. | | | | | | | | | |

| PICO | Study | | | No. of participants | | Follow-up (median/mean, range) yr | Cyclophosphamide  (CYC) | Pulmonary function Outcomes | Effect size | PFT quality | Risk of bias |
| --- | --- | --- | --- | --- | --- | --- | --- | --- | --- | --- | --- |
| 2 What is the risk of diffusion capacity impairment in CAYA treated with CYC compared to CAYA not treated with CYC? | Mulder 2011 (9) | | | 193 CCS | | Median 17.9 (range 5.6-36.8) | High-dose CYC  43 (22.3%) | 85/193 Diffusion impairment (DLCO <75% pred) | Odds Ratio (95%CI) High-dose CYC vs no  1.25 (0.58-2.71) | 1. No  2. No  3. No  4. No  5. No  6. No | Retrospective cohort  SB: Low risk  AB: Low risk  DB: Low risk  CF: Low risk |
| GRADE assessment: | |  |  | |  | | | | | | |
| Study design: | |  | +4 | 1 retrospective cohort study | | | | | | | |
| Study limitations: | |  | 0 | No limitations: Selection low in 1/1; Attrition bias low in 1/1; Detection bias low 1/1; Confounding low in 1/1 | | | | | | | |
| Consistency: | |  | 0 | One study only | | | | | | | |
| Directness: | |  | -1 | Population and outcomes broadly generalizable; PFT quality unsure (no reference values and use of guidelines stated) | | | | | | | |
| Precision: | |  | -1 | Some imprecision, only 1 study but with precise results with small confidence interval | | | | | | | |
| Publication bias: | |  | 0 | Unlikely | | | | | | | |
| Effect size: | |  | 0 | No large magnitude of effect | | | | | | | |
| Dose-response: | |  | 0 | Not applicable | | | | | | | |
| Plausible confounding: | |  | 0 | No plausible confounding | | | | | | | |
| Quality of evidence: | | | ⊕⊕⊖⊖ LOW | | | | | | | | |
| Conclusion: | | | No significant effect on diffusion capacity impairment (DLCO) in CAYA cancer survivors after cyclophosphamide vs. no cyclophosphamide.  (1 study; 1 non-significant effect; 193 participants) | | | | | | | | |
| Comments: | | | One study only with non-significant 95%CI and no information on PFT quality. | | | | | | | | |

##### 2a Different doses

| PICO | Study | | | No. of participants | Follow-up (median/mean, range) yr | Cyclophosphamide | Pulmonary function Outcomes | Effect size | | PFT quality | Risk of bias |
| --- | --- | --- | --- | --- | --- | --- | --- | --- | --- | --- | --- |
| 2a What is the risk of obstructive abnormalities in CAYA survivors treated with different doses of CYC? | Green 2015 (10) | | | 260 embryonal brain tumors | Minimum 2 yr | 260 (100%) | Proportion of CCS with FEV1 below predicted after 60 months  29% **FEV1** (<80% pred) | | CYC dose was not found to be a significant predictor of FEV1 % predicted. (univariable model) | 1. No  2. Yes  Newth CJ, Eur Respir J, 1997; Stocks J, Eur Respir J, 1995; Paoletti P, Am Rev Respir Dis, 1985; Hankinson JL, Am J Respir Crit Care Med, 1999; Wang X, Pediatr Pulmonol, 1993; Knudosn RJ, Am Rev Respir Dis, 1983; Eigen H, Am J Respir Crit Care Med, 2001; Polgar G, 1971; Kim YJ, Pediatr Pulmonol, 2012; Zapletal A, 1987; HIbbert ME, Pediatr Pulmonol, 1989; Stanojevic S, Am J Respir Crit Care Med, 2008  3. No  4. Yes: ATS  5. No  6. No | Prospective cohort  SB: low risk  AB: high risk  DB: unclear  CF: high risk |
| GRADE assessment: | |  |  |  | | | | | | | |
| Study design: | |  | +4 | 1 prospective cohort study | | | | | | | |
| Study limitations: | |  | -3 | Severe limitations: Selection bias low in 1/1; Attrition bias high 1/1 ; Detection bias unclear 1/1; Confounding high in 1/1 | | | | | | | |
| Consistency: | |  | 0 | One study only | | | | | | | |
| Directness: | |  | -1 | Population and outcomes broadly generalizable; PFT quality unsure (10 different references stated und use of ATS guidelines) | | | | | | | |
| Precision: | |  | -2 | Important imprecision, univariable analysis only, no effect size mentioned, one study only | | | | | | | |
| Publication bias: | |  | 0 | Publication bias unlikely | | | | | | | |
| Effect size: | |  | 0 | No large magnitude of effect | | | | | | | |
| Dose-response: | |  | 0 | No clear dose response relationship | | | | | | | |
| Plausible confounding: | |  | 0 | No plausible confounding | | | | | | | |
| Quality of evidence: | | | ⊕⊖⊖⊖ VERY LOW | | | | | | | | |
| Conclusion: | | | No significant effect on obstructive abnormalities with increasing doses of cyclophosphamide  (1 study; 260 participants) | | | | | | | | |
| Comments: | | | One study only, univariable analysis, and no effect size mentioned or p-value. | | | | | | | | |

| PICO | Study | | | No. of participants | Follow-up (median/mean, range) yr | Cyclophosphamide | Pulmonary function Outcomes | Effect size | PFT quality | Risk of bias |
| --- | --- | --- | --- | --- | --- | --- | --- | --- | --- | --- |
| 2a What is the risk of restrictive abnormalities in CAYA survivors treated with different doses of CYC? | Nysom 1998 (11) | | | 94 leukemia CCS | Median 10.6 (range 3.4-23.4) | 43 (46%)  Cumulative dose of CYC as continuous variable | 17% (15/89) with reduced or raised **TLC** | Regression coefficient (95%CI), p-value:  Simple regression  -0.11 (-0.23 – 0.01), 0.07  Multiple regression:  -0.14 (-0.25 – -0.02), 0.02 | 1. No  2. Yes  Reference form own laboratory by adjusting published  reference values (Quanjer PH, Pediatr Pulmonol. 1995; Rosenthal M, Thorax, 1993; Quanjer PH, Bull Eur Physiopathol Respir, 1983: Stam H, Pediatr Pulmonl, 1996)  3. No  4. Yes  5. No  6. Yes | Retrospective cohort  SB: High risk  AB: Low risk  DB: Unclear  CF: High risk |
| GRADE assessment: | |  |  |  | | | | | | |
| Study design: | |  | +4 | 1 retrospective cohort study | | | | | | |
| Study limitations: | |  | -2 | Severe limitations: Selection bias high in 1/1; Attrition bias low in 1/1 ; Detection bias unclear 1/1; Confounding high in 1/1 | | | | | | |
| Consistency: | |  | 0 | One study only | | | | | | |
| Directness: | |  | -1 | Population and outcomes broadly generalizable; PFT quality good (reference values and ATS guidelines stated) | | | | | | |
| Precision: | |  | -1 | Some imprecision, precise results with small confidence interval, only one study | | | | | | |
| Publication bias: | |  | -1 | Publication bias likely, as not for all lung function parameters assessed the are shown (FEV1, FVC) | | | | | | |
| Effect size: | |  | 0 | No large magnitude of effect | | | | | | |
| Dose-response: | |  | 1 | Dose response relationship | | | | | | |
| Plausible confounding: | |  | 0 | No plausible confounding | | | | | | |
| Quality of evidence: | | | ⊕⊖⊖⊖ VERY LOW | | | | | | | |
| Conclusion: | | | Increased risk for restrictive abnormalities (TLC) with increasing doses of cyclophosphamide in CAYA cancer survivors.  (1 study; 89 participants with TLC measurements) | | | | | | | |
| Comments: | | | One study only with effect size and 95%CI mentioned. | | | | | | | |

##### 2b Age at exposure to cyclophosphamide

**No study**

#### PICO 3: Methotrexate (MTX)

| PICO | Study | No. of participants | Follow-up (median/mean, range) yr | Methotrexate (MTX) | Pulmonary function Outcomes | Effect size | PFT quality | Risk of bias |
| --- | --- | --- | --- | --- | --- | --- | --- | --- |
| 3 What is the risk of pulmonary dysfunction in CAYA treated with methotrexate compared to CAYA not treated with methotrexate? | | | | | | | | |

**No study**

##### 3a Different doses

| PICO | Study | No. of participants | Follow-up (median/mean, range) yr | Methotrexate (MTX) | Pulmonary function Outcomes | Effect size | PFT quality | Risk of bias |
| --- | --- | --- | --- | --- | --- | --- | --- | --- |
| 3a What is the risk of obstructive abnormalities in CAYA survivors treated with different doses of MTX? | | | | | | | | |

**No study**

| PICO | Study | | | No. of participants | Follow-up (median/mean, range) yr | Methotrexate (MTX) | Pulmonary function Outcomes | Effect size | PFT quality | Risk of bias |
| --- | --- | --- | --- | --- | --- | --- | --- | --- | --- | --- |
| 3a What is the risk of restrictive abnormalities in CAYA survivors treated with different doses of MTX? | Nysom 1998 (11) | | | 94 leukemia survivors | Median 10.6 (range 3.4-23.4) | 16 (17%) with high-dose MTX (HDM)    Number of HDM cycles (cont.) | 17% (15/89) with reduced or raised **TLC** | Regression coefficient (95%CI), p-value:  Simple regression:  -0.005 (-0.08 - 0.07) 0.9 | 1. No  2. Yes  Reference form own laboratory by adjusting published  reference values (Quanjer PH, Pediatr Pulmonol. 1995; Rosenthal M, Thorax, 1993; Quanjer PH, Bull Eur Physiopathol Respir, 1983: Stam H, Pediatr Pulmonl, 1996)  3. No  4. Yes  5. No  6. Yes | Retrospective cohort  SB: High risk  AB: Low risk  DB: Unclear  CF: High risk |
| GRADE assessment: | |  |  | |  | | | | | |
| Study design: | |  | +4 | 1 retrospective cohort study | | | | | | |
| Study limitations: | |  | -2 | Severe limitations: Selection bias high in 1/1; Attrition bias low in 1/1 ; Detection bias unclear 1/1; Confounding high in 1/1 | | | | | | |
| Consistency: | |  | 0 | One study only | | | | | | |
| Directness: | |  | -1 | Population and outcomes broadly generalizable; PFT quality unsure (“own” reference values generated but lung function procedure stated) | | | | | | |
| Precision: | |  | -1 | Some imprecision, precise results with small confidence interval, only one study | | | | | | |
| Publication bias: | |  | -1 | Publication bias likely, as not for all lung function parameters assessed the are shown (FEV1, FVC) | | | | | | |
| Effect size: | |  | 0 | No large magnitude of effect | | | | | | |
| Dose-response: | |  | 0 | No clear dose response relationship | | | | | | |
| Plausible confounding: | |  | 0 | No plausible confounding | | | | | | |
| Quality of evidence: | | | ⊕⊖⊖⊖ VERY LOW | | | | | | | |
| Conclusion: | | | No significant effect on restrictive abnormalities (TLC) after increasing doses of methotrexate in CAYA cancer survivors.  (1 study; 89 with TLC measurements) | | | | | | | |
| Conclusion: | | | One study, small number of participants in whole study and only 16 exposed to high-dose methotrexate. | | | | | | | |

| PICO | Study | No. of participants | Follow-up (median/mean, range) yr | Methotrexate (MTX) | Pulmonary function Outcomes | Effect size | PFT quality | Risk of bias |
| --- | --- | --- | --- | --- | --- | --- | --- | --- |
| 3a What is the risk of hyperinflation in CAYA survivors treated with different doses of MTX? | | | | | | | | |

**No study**

| PICO | Study | No. of participants | Follow-up (median/mean, range) yr | Methotrexate (MTX) | Pulmonary function Outcomes | Effect size | PFT quality | Risk of bias |
| --- | --- | --- | --- | --- | --- | --- | --- | --- |
| 3a What is the risk of diffusion capacity impairment in CAYA survivors treated with different doses of MTX? | | | | | | | | |

**No study**

##### 3b Age at exposure

| PICO | Study | No. of participants | Follow-up (median/mean, range) yr | Methotrexate | Pulmonary function Outcomes | Effect size | Risk of bias |
| --- | --- | --- | --- | --- | --- | --- | --- |
| 3b What is the risk in younger compared to older age at treatment? | | | | | | | |

**No study**

#### PICO 4: Gemcitabine

| PICO | Study | No. of participants | Follow-up (median/mean, range) yr | Methotrexate | Pulmonary function Outcomes | Effect size | Risk of bias |
| --- | --- | --- | --- | --- | --- | --- | --- |
| 4 What is the risk of pulmonary dysfunction in CAYA treated with gemcitabine compared to CAYA not treated with gemcitabine? | | | | | | | |

**No study**

#### PICO 5: Bleomycin

| PICO | Study | | | No. of participants | | Follow-up (median/mean, range) yr | Bleomycin exposure | Pulmonary function Outcomes | Effect size |  | Risk of bias |
| --- | --- | --- | --- | --- | --- | --- | --- | --- | --- | --- | --- |
| 5 What is the risk of obstructive abnormalities in CAYA treated with bleomycin compared to CAYA not treated with bleomycin? | Record, 2016 (1) | | | 143 CCS | | Mean 14.1 ± 4.8 (SD) | 48 (33.6%) | Obstructive  (FEV1, FEV1/FVC <80% pred or FEF25–75% <68%)  12.5% (6/48) bleomycin  32.6% (31/95) no bleomycin | Univariable analysis comparing bleomycin yes/no  (p-value)  0.01  **More in non-exposed** | 1. No  2. Yes  Wang X, Pediatr Pulmonol 2005; Hankinson JL, Am J Respir Crit Care Med 1999  3. No  4. Yes  5. No  6. Yes | Retrospective cohort  SB: high risk  AB: low risk  DB: low risk  CF: high risk |
|  | De 2015 (12) | | | 49 Osteosarcoma survivors | | Median 2.91 (range 0.01-8.28) | 38 (78%) | Proportion of CCS with abnormal results per lung function parameter in whole cohort  29% (14/49) FEV1 <80% pred  20% (10/49) FEF25–5% <68% pred  24% (12/49) Obstructive  (FEV1/FVC <80%pred, FEV1<80%pred or FEF25-75<68%pred with normal TLC) | Univariable logistic regression comparing bleomycin yes/no  Odds Ratio (p-value)  0.07 (<0.01)  0.18 (<0.05)  0.27 (NS)  **More in non-exposed** | 1. No  2. Yes  Hankinson JL, Am J Respir Crit Care Med, 1999;  Wang X, Pediatr Pulmonol, 1993  3. No  4. Yes: ATS  5. No  6. No | Retrospective cohort  SB: High Risk  AB: High Risk  DB: Low risk  CF: High Risk |
|  | Denbo, 2014 (13) | | | 21  Osteo-sarcoma survivor | | Mean 20 yr (SD +/-9) | 6 (28%) | Number of CCS with abnormal results per parameter  FEV1 <80% pred  50% (3/6) bleomycin  50% (7/15) no bleomycin | Univariable analysis comparing bleomycin yes/no  (p-value)  1.00 | 1. No  2. Yes  Hankinson JL, Am J Respir Crit Care Med, 1999; Miller A, Am Rev Respir Dis, 1983  3. No  4. Yes: ATS, Morris AH, 1984  5. No  6. No | Prospective cohort  SB: low risk  AB: low risk  DB: unclear  CF: high risk |
| GRADE assessment: | |  |  | |  | | | | | | |
| Study design: | |  | +4 | 2 retrospective cohort studies, 1 prospective cohort study | | | | | | | |
| Study limitations: | |  | -2 | Severe limitations: Selection bias high in 2/1, low in 1/3; Attrition bias high in 1/3, low in 2/3 ; Detection bias low in 2/3, unclear in 1/3; Confounding high in 3/3 | | | | | | | |
| Consistency: | |  | 0 | No inconsistency. All studies show no increased risk for obstructive abnormalities in CAYA survivors exposed to bleomycin. | | | | | | | |
| Directness: | |  | 0 | Population and outcomes broadly generalizable; PFT quality good (all studies mention reference values and lung function procedures) | | | | | | | |
| Precision: | |  | -1 | Precision cannot be judged as 2/3 show p-value only and 1/3 shows OR but without 95%CI. All studies performed univariable analysis only. | | | | | | | |
| Publication bias: | |  | 0 | Publication bias unlikely | | | | | | | |
| Effect size: | |  | 0 | No large magnitude of effect | | | | | | | |
| Dose-response: | |  | 0 | Not applicable | | | | | | | |
| Plausible confounding | |  | 0 | No plausible confounding | | | | | | | |
| Quality of evidence: | | | ⊕⊖⊖⊖ VERY LOW | | | | | | | | |
| Conclusion: | | | Deacreased risk for obstructive abnormalities (FEV1, FEF25-75, FEV1/FVC) after bleomycin vs. no bleomycin in CAYA cancer survivors.  (3 studies; 2 significant effects, 1 non-significant effect; 213 participants) | | | | | | | | |
| Comment: | | | All three studies with univariable analysis only and all report results with p-values only and without confidence intervals. Two studies with less than 50 participants. | | | | | | | | |

| PICO | Study | | | | No. of participants | | Follow-up (median/mean, range) yr | Bleomycin exposure | Pulmonary function Outcomes | Effect size |  | Risk of bias |
| --- | --- | --- | --- | --- | --- | --- | --- | --- | --- | --- | --- | --- |
| 5 What is the risk of restrictive abnormalities in CAYA treated with bleomycin compared to CAYA not treated with bleomycin? | Record, 2016 (1) | | | | 143 CCS | | Mean 14.1 ± 4.8 (SD) | 48 (33.6%) | Restrictive  (TLC<80% pred)  12.5% (6/48) bleomycin  13.7% (13/95) no bleomycin | Univariable analysis comparing bleomycin yes/no  (p-value)  0.84 | 1. No  2. Yes  Wang X, Pediatr Pulmonol 2005; Hankinson JL, Am J Respir Crit Care Med 1999  3. No  4. Yes  5. No  6. Yes | Retrospective cohort  SB: high risk  AB: low risk  DB: low risk  CF: high risk |
|  | Armenian 2015 (14) | | | | 121 CCS | | Median 17.1 (6.3-40.1) | 42 (34.7%) | Restrictive  TLC <75%pred and FEV1 >80%pred)  19% (8/42) bleomycin  27% (21/79) no bleomycin | Univariable logistic regression  Odds Ratio (95%CI)  0.7 (0.3-1.6) | 1. Yes  2. No  3. No  4. Yes: ATS, Miller MR, Eur Respir J, 2005  5. No  6. Yes | Prospective cohort  SB: Low risk  AB: low risk  DB: low risk  CF: high risk |
|  | De 2015 (12) | | | | 49  Osteo-sarcoma survivors | | Median 2.91 (range 0.01-8.28) | 38 (78%) | Proportion of CCS with abnormal results per lung function parameter in whole cohort  24% (12/49) FVC <80% pred  15% (7/49) TLC <77% pred  15% (7/49) Restrictive disease  (TLC <77%) | Univariable logistic regression comparing bleomycin yes/no  Odds Ratio (p-value)  0.15 (<0.05)  0.27 (NS)  0.27 (NS) | 1. No  2. Yes  Hankinson JL, Am J Respir Crit Care Med, 1999;  Wang X, Pediatr Pulmonol, 1993  3. No  4. Yes: ATS  5. No  6. No | Retrospective cohort  SB: High Risk  AB: High Risk  DB: Low risk  CF: High Risk |
|  | Denbo, 2014 (13) | | | | 21  Osteo-sarcoma survivor | | Mean 20 yr (SD +/-9) | 6 (28%) | Number of CCS with abnormal results per parameter  FVC <80%predicted  50% (3/6) bleomycin  36% (5/14) no bleomycin  TLC <75%predicted  17% (1/6) bleomycin  33% (7/15) no bleomycin | Univariate analysis comparing bleomycin yes/no (p-value)  0.642  0.623 | 1. No  2. Yes  Hankinson JL, Am J Respir Crit Care Med, 1999; Miller A, Am Rev Respir Dis, 1983  3. No  4. Yes: ATS, Morris AH, 1984  5. No  6. No | Prospective cohort  SB: low risk  AB: low risk  DB: unclear  CF: high risk |
|  | Mulder 2011 (9) | | | | 193 CCS | | Median 17.9 (range 5.6-36.8) | 110 (57%) | Total 34/193 Restrictive  (TLC or FVC <75%) | Comparison of bleomycin yes/no  Odds Ratio (95%CI)  1.5 (0.38-5.97) | 1. No  2. No  3. No  4. No  5. No  6. No | Retrospective cohort  SB: Low risk  AB: Low risk  DB: Low risk  CF: Low risk |
| GRADE assessment: | |  |  | | |  | | | | | | |
| Study design: | |  | +4 | 3 retrospective cohort studies, 2 prospective cohort studies | | | | | | | | |
| Study limitations: | |  | -3 | Severe limitations: Selection bias high in 2/5, low in 3/5; Attrition bias high in 1/5, low in 4/5 ; Detection bias low in 4/5, unclear in 1/5; Confounding high in 4/5, low in 1/5 | | | | | | | | |
| Consistency: | |  | -1 | Important inconsistency. Three studies show more restrictive abnormalities in CAYA survivors not exposed to bleomycin; in one study it depends on the outcome factor assessed whether exposed CAYA survivors are at risk or not (FVC vs TLC); in one study exposed CYAY survivors are more at risk than non-exposed. | | | | | | | | |
| Directness: | |  | 0 | Population and outcomes broadly generalizable; PFT quality unsure (not homogeneous across studies, two studies do not mention reference values and one does not mention lung function procedures used) | | | | | | | | |
| Precision: | |  | -1 | Important imprecision, 2/5 shows precise results with small confidence interval, in 3/5 precision cannot be judged as results are shown with p-value only; 4/5 report univariable analysis only | | | | | | | | |
| Publication bias: | |  | 0 | Publication bias unlikely | | | | | | | | |
| Effect size: | |  | 0 | No large magnitude of effect | | | | | | | | |
| Dose-response: | |  | 0 | Not applicable | | | | | | | | |
| Plausible confounding | |  | 0 | No plausible confounding | | | | | | | | |
| Quality of evidence: | | | ⊕⊖⊖⊖ VERY LOW | | | | | | | | | |
| Conclusion: | | | No significant effect on restrictive abnormalities (TLC or FVC) after bleomycin vs. no bleomycin in CAYA cancer survivors.  (5 studies; 5 non-significant effects; 527 participants) | | | | | | | | | |
| Comment: | | | Four studies with univariable analysis only, three reported results with p-values only and without confidence intervals. Two studies with < 50 participants. | | | | | | | | | |

| PICO | Study | | | No. of participants | | | Follow-up (median/mean, range) yr | Bleomycin exposure | Pulmonary function Outcomes | Effect size |  | Risk of bias |
| --- | --- | --- | --- | --- | --- | --- | --- | --- | --- | --- | --- | --- |
| 5 What is the risk of hyperinflation in CAYA treated with bleomycin compared to CAYA not treated with bleomycin? | Record, 2016 (1) | | | 143 CCS | | | Mean 14.1 ± 4.8 (SD) | 48 (33.6%) | Hyperinflation  (RV >120%pred or RV/TLC >28%pred)  20.8% (10/48) with bleomycin  51.6% (49/95) without bleomycin | Univariable analysis comparing bleomycin yes/no  (p-value)  <0.01  **More in non-exposed** | 1. No  2. Yes  Wang X, Pediatr Pulmonol 2005; Hankinson JL, Am J Respir Crit Care Med 1999  3. No  4. Yes  5. No  6. Yes | Retrospective cohort  SB: high risk  AB: low risk  DB: low risk  CF: high risk |
|  | De 2015 (12) | | | 49  Osteo-sarcoma survivors | | | Median 2.91 (range 0.01-8.28) | 38 (78%) | Proportion of CCS with abnormal RV/TLC in whole cohort  21% (10/49) RV/TLC >28% | Univariable logistic regression comparing bleomycin yes/no  Odds Ratio (p-value)  0.15 (<0.05)  **More in non-exposed** | 1. No  2. Yes  Hankinson JL, Am J Respir Crit Care Med, 1999;  Wang X, Pediatr Pulmonol, 1993  3. No  4. Yes: ATS  5. No  6. No | Retrospective cohort  SB: High Risk  AB: High Risk  DB: Low risk  CF: High Risk |
| GRADE assessment: | |  |  | | |  | | | | | | |
| Study design: | |  | +4 | | 2 retrospective cohort studies | | | | | | | |
| Study limitations: | |  | -3 | | Severe limitations: Selection bias high in 2/2; Attrition bias high in 1/2, low in 1/2 ; Detection bias low in 2/2; Confounding high in 2/2 | | | | | | | |
| Consistency: | |  | 0 | | No inconsistency. Both studies show that hyperinflation is not associated with bleomycin exposure. | | | | | | | |
| Directness: | |  | 0 | | Population and outcomes broadly generalizable; PFT quality good (both studies mention reference values and lung function procedures) | | | | | | | |
| Precision: | |  | -1 | | Precision cannot be judged as 2/2 show results with p-value only and 2/2 univariable analysis only | | | | | | | |
| Publication bias: | |  | 0 | | Publication bias unlikely | | | | | | | |
| Effect size: | |  | 0 | | No large magnitude of effect | | | | | | | |
| Dose-response: | |  | 0 | | Not applicable | | | | | | | |
| Plausible confounding | |  | 0 | | No plausible confounding | | | | | | | |
| Quality of evidence: | | | ⊕⊖⊖⊖ VERY LOW | | | | | | | | | |
| Conclusion: | | | Decreased risk for hyperinflation after bleomycin vs. no bleomycin in CAYA cancer survivors  (2 studies with significant effect; 192 participants) | | | | | | | | | |
| Comment: | | | Both studies with univariable analysis and results as p-values only. One study with < 50 participants. | | | | | | | | | |

| PICO | Study | | | | No. of participants | | Follow-up (median/mean, range) yr | Bleomycin exposure | Pulmonary function Outcomes | Effect size |  | Risk of bias |
| --- | --- | --- | --- | --- | --- | --- | --- | --- | --- | --- | --- | --- |
| 5 What is the risk of diffusion capacity impairment in CAYA treated with bleomycin compared to CAYA not treated with bleomycin? | Armenian 2015 (14) | | | | 121 CCS | | Median 17.1 (6.3-40.1) | 42 (34.7%) | Diffusion abnormality  (DLCO <75%pred)  31% (13/42) bleomycin  37% (29/79) no bleomycin | Univariable logistic regression  Odds Ratio (95%CI)  0.8 (0.4-1.7) | 1. Yes  2. No  3. No  4. Yes: ATS, Miller MR, Eur Respir J, 2005  5. No  6. Yes | Prospective cohort  SB: Low risk  AB: low risk  DB: low risk  CF: high risk |
|  | De 2015 (12) | | | | 49 Osteosarcoma survivors | | Median 2.91 (range 0.01-8.28) | 38 (78%) | 9% (4/49) DLCO adj <65% pred  14% (6/49) Diffusion impairment  (DLCO <65% or DLCOadj/VA <4ml(mmHg/min/l)) | Univariable logistic regression comparing bleomycin yes/no  Odds Ratio (p-value)  0.06 (<0.05)  0.08 (<0.01) | 1. No  2. Yes  Hankinson JL, Am J Respir Crit Care Med, 1999;  Wang X, Pediatr Pulmonol, 1993  3. No  4. Yes: ATS  5. No  6. No | Retrospective cohort  SB: High Risk  AB: High Risk  DB: Low risk  CF: High Risk |
|  | Denbo, 2014 (13) | | | | 21 Osteosarcoma survivor | | Mean 20 yr (SD +/-9) | 6 (28%) | Number of CCS with abnormal results per parameter  DLCO_corr_ <75%predicted  50% (3/6) bleomycin  46% (6/13) no bleomycin | Univariable analysis comparing bleomycin yes/no  (p-value)  1.00 | 1. No  2. Yes  Hankinson JL, Am J Respir Crit Care Med, 1999; Miller A, Am Rev Respir Dis, 1983  3. No  4. Yes: ATS, Morris AH, 1984  5. No  6. No | Prospective cohort  SB: low risk  AB: low risk  DB: unclear  CF: high risk |
|  | Mulder 2011 (9) | | | | 193 CCS | | Median 17.9 (range 5.6-36.8) | 110 (57%) | Total 85/193 diffusion impairment  (DLCO <75%) | Comparison of bleomycin yes/no  Odds Ratio (95%CI)  1.99 (0.56-7.07) | 1. No  2. No  3. No  4. No  5. No  6. No | Retrospective cohort  SB: Low risk  AB: Low risk  DB: Low risk  CF: Low risk |
| GRADE assessment: | |  |  | | |  | | | | | | |
| Study design: | |  | +4 | 2 retrospective cohort studies, 2 prospective cohort studies | | | | | | | | |
| Study limitations: | |  | -1 | Severe limitations: Selection bias high in 1/4, low in 3/4; Attrition bias high in 1/4, low in 3/4 ; Detection bias low in 3/4, unclear in 1/4; Confounding high in 3/4, low in 1/4 | | | | | | | | |
| Consistency: | |  | -1 | Important inconsistency. Two studies show higher DLCO impairment in CAYS survivors not exposed to bleomycin, one study shows no difference, and one study shows a not significant association between bleomycin exposure and diffusion capacity impairment | | | | | | | | |
| Directness: | |  | -1 | Population and outcomes broadly generalizable; PFT quality unsure (not homogeneous across studies, two studies do not mention reference values and one does not mention lung function procedures used) | | | | | | | | |
| Precision: | |  | -1 | Important imprecision, 2/4 shows precise results with small confidence interval, in 2/4 precision cannot be judged as results are shown as p-value only; 3/4 report univariable analysis only | | | | | | | | |
| Publication bias: | |  | 0 | Publication bias unlikely | | | | | | | | |
| Effect size: | |  | 0 | No large magnitude of effect | | | | | | | | |
| Dose-response: | |  | 0 | Not applicable | | | | | | | | |
| Plausible confounding | |  | 0 | No plausible confounding | | | | | | | | |
| Quality of evidence: | | | ⊕⊖⊖⊖ VERY LOW | | | | | | | | | |
| Conclusion: | | | Inconsistent findings for diffusion capacity impairment after bleomycin vs. no bleomycin in CAYA cancer survivors.  (4 studies, 1 significant effect, 3 non-significant effects; 384 participants) | | | | | | | | | |
| Comment: | | | Three studies with univariable analysis only, two reported results with p-values only and without confidence intervals. Outcome and cutoff value defined identical in three studies (DLCO< 75%predicted) and different in one study. | | | | | | | | | |

##### 5a Different doses

| PICO | Study | | | | No. of participants | Follow-up (median/mean, range) yr | Bleomycin exposure | Pulmonary function Outcomes | Effect size | PFT quality | Risk of bias |
| --- | --- | --- | --- | --- | --- | --- | --- | --- | --- | --- | --- |
| 5a What is the risk of obstructive abnormalities in CAYA survivors associated with different doses of bleomycin? | Record, 2016 (1) | | | | 143 CCS | Mean 14.1 ± 4.8 (SD) | 13 low dose (<60IU/m^2^)  35 high dose (>= 60IU/m^2^) | Obstructive disease  (FEV1, FEV1/FVC <80% predicted or FEF25–75% <68%)  11% (4/35) high  15% (2/13) low | Univariable comparison Chi2 low dose/high dose  0.72 | 1. No  2. Yes  Wang X, Pediatr Pulmonol 2005; Hankinson JL, Am J Respir Crit Care Med 1999  3. No  4. Yes  5. No  6. Yes | Retrospective cohort  SB: high risk  AB: low risk  DB: low risk  CF: high risk |
| GRADE assessment: | |  |  |  | | | | | | | |
| Study design: | |  | +4 | 1 retrospective cohort study | | | | | | | |
| Study limitations: | |  | -2 | Severe limitations: Selection bias high in 1/1; Attrition bias low in 1/1; Detection bias low in 1/1; Confounding high in 1/1 | | | | | | | |
| Consistency: | |  | 0 | One study only | | | | | | | |
| Directness: | |  | 0 | Population and outcomes broadly generalizable; PFT quality good (reference values and lung function procedures mentioned) | | | | | | | |
| Precision: | |  | -1 | Precision cannot be judged as results are shown as p-value only, univariable analysis, only one study | | | | | | | |
| Publication bias: | |  | 0 | Publication bias unlikely | | | | | | | |
| Effect size: | |  | 0 | No large magnitude of effect | | | | | | | |
| Dose-response: | |  | 0 | No clear dose response relationship | | | | | | | |
| Plausible confounding: | |  | 0 | No plausible confounding | | | | | | | |
| Quality of evidence: | | | ⊕⊖⊖⊖ VERY LOW | | | | | | | | |
| Conclusion: | | | No significant effect for obstructive abnormalities in CAYA cancer survivors exposed to higher doses (≥60IU/m^2^) of bleomycin vs. lower doses (<60IU/m^2^).  (1 study; 143 participants, 34 exposed to bleomycin) | | | | | | | | |
| Comments: | | | Results reported from univariable analysis and as p-values only. Small sample size exposed to bleomycin in total. | | | | | | | | |

| PICO | Study | | | No. of participants | Follow-up (median/mean, range) yr | Bleomycin exposure | Pulmonary function Outcomes | Effect size | PFT quality | Risk of bias |
| --- | --- | --- | --- | --- | --- | --- | --- | --- | --- | --- |
| 5a What is the risk of restrictive abnormalities in CAYA survivors associated with different doses of bleomycin? | Record, 2016 (1) | | | 143 CCS | Mean 14.1 ± 4.8 (SD) | 13 low dose (<60IU/m^2^)  35 high dose (>= 60IU/m^2^) | Restrictive disease  (TLC<80% predicted)  17.1% (6/35) high  0% low | Univariable comparison Chi2 low dose/high dose  0.05 | 1. No  2. Yes  Wang X, Pediatr Pulmonol 2005; Hankinson JL, Am J Respir Crit Care Med 1999  3. No  4. Yes  5. No  6. Yes | Retrospective cohort  SB: high risk  AB: low risk  DB: low risk  CF: high risk |
| GRADE assessment: | |  |  | |  | | | | | |
| Study design: | |  | +4 | 1 retrospective cohort study | | | | | | |
| Study limitations: | |  | -2 | Severe limitations: Selection bias high in 1/1; Attrition bias low in 1/1; Detection bias low in 1/1; Confounding high in 1/1 | | | | | | |
| Consistency: | |  | 0 | One study only | | | | | | |
| Directness: | |  | 0 | Population and outcomes broadly generalizable; PFT quality good (reference values and lung function procedures mentioned) | | | | | | |
| Precision: | |  | -1 | Precision cannot be judged as results are shown as p-value only, univariable analysis | | | | | | |
| Publication bias: | |  | 0 | Publication bias unlikely | | | | | | |
| Effect size: | |  | 0 | No large magnitude of effect | | | | | | |
| Dose-response: | |  | 0 | No clear dose response relationship | | | | | | |
| Plausible confounding: | |  | 0 | No plausible confounding | | | | | | |
| Quality of evidence: | | | ⊕⊖⊖⊖ VERY LOW | | | | | | | |
| Conclusion: | | | Increased risk for restrictive abnormalities (TLC) in CAYA cancer survivors after higher doses (≥60IU/m^2^) of bleomycin vs. lower doses (<60IU/m^2^) of bleomycin in CAYA cancer survivors.  (1 study; 143 participants) | | | | | | | |
| Comments: | | | Results reported from univariable analysis and as p-values only. Small sample size exposed to bleomycin in total. | | | | | | | |

| PICO | Study | | | | No. of participants | Follow-up (median/mean, range) yr | Bleomycin exposure | Pulmonary function Outcomes | Effect size | PFT quality | Risk of bias |
| --- | --- | --- | --- | --- | --- | --- | --- | --- | --- | --- | --- |
| 5a What is the risk of hyperinflation in CAYA survivors associated with different doses of bleomycin? | Record, 2016 (1) | | | | 143 CCS | Mean 14.1 ± 4.8 (SD) | 13 low dose (<60IU/m^2^)  35 high dose  (≤60IU/m^2^) | Hyperinflation  (RV >120%predicted or RV/TLC >28% predicted)  25.7% (9/35) high  7.7% (1/13) low | Univariable comparison Chi2 low dose/high dose  0.14 | 1. No  2. Yes  Wang X, Pediatr Pulmonol 2005; Hankinson JL, Am J Respir Crit Care Med 1999  3. No  4. Yes  5. No  6. Yes | Retrospective cohort  SB: high risk  AB: low risk  DB: low risk  CF: high risk |
| GRADE assessment: | |  |  | | |  | | | | | |
| Study design: | |  | +4 | 1 retrospective cohort study | | | | | | | |
| Study limitations: | |  | -2 | Severe limitations: Selection bias high in 1/1; Attrition bias low in 1/1; Detection bias low in 1/1; Confounding high in 1/1 | | | | | | | |
| Consistency: | |  | 0 | One study only | | | | | | | |
| Directness: | |  | 0 | Population and outcomes broadly generalizable; PFT quality good (reference values and lung function procedures mentioned) | | | | | | | |
| Precision: | |  | -1 | Precision cannot be judged as results are shown as p-value only, univariable analysis | | | | | | | |
| Publication bias: | |  | 0 | Publication bias unlikely | | | | | | | |
| Effect size: | |  | 0 | No large magnitude of effect | | | | | | | |
| Dose-response: | |  | 0 | No clear dose response relationship | | | | | | | |
| Plausible confounding: | |  | 0 | No plausible confounding | | | | | | | |
| Quality of evidence: | | | ⊕⊖⊖⊖ VERY LOW | | | | | | | | |
| Conclusion: | | | No significant effect on hyperinflation (RV, RV/TLC) in CAYA cancer survivors after higher doses (≥60IU/m^2^) of bleomycin vs. lower doses (<60IU/m^2^) of bleomycin.  (1 study; 143 participants, 34 exposed to bleomycin) | | | | | | | | |
| Comments: | | | Results reported as univariable analysis and p-values only. Small sample size exposed to bleomycin in total. | | | | | | | | |

| PICO | Study | | | No. of participants | Follow-up (median/mean, range) yr | Bleomycin exposure | Pulmonary function Outcomes | Effect size | PFT quality | Risk of bias |
| --- | --- | --- | --- | --- | --- | --- | --- | --- | --- | --- |
| 5a What is the risk of diffusion capacity impairment in CAYA survivors associated with different doses of bleomycin? | Marina 1995 (15) | | | 37 Hodgkin Lymphoma CCS | Median 7.7 (range 4.7-10.5) | 37 (100%) | Cumulative dose of bleomycin and change in DLCO% predicted  (DLCO <80% pred)  Cumulative dose of bleomycin and change in DLCO/VA% predicted  (DLCO <80% pred) | Cumulative dose of bleomycin (cont.)  p=0.98  p=0.92 | 1. No  2. Yes  Polgar G, 1971; Hsu KH, J Pediatr 1979; Goldman HI, Am Rev Tuberc, 1959; Morris JF, Am Rev Respir Dis, 1971; Weng TR, Am Rev Respir Dis, 1969; Miller A, Am Rev Respir Dis, 1983  3. No  4. Yes  5. No  6. No | Prospective cohort  SB: Low risk  AB: Low risk  DB: Unclear  CF: High risk |
|  | Zorzi 2015 (16) | | | 143 CCS | Median 4.4  (2 – 7.4) | 86 (60%) | Total 19% (27/143) with abnormal DLCO  (DLCO <80% pred) | Cumulative dose of bleomycin (cont.) (1U/m2 increase of bleomycin)  OR (95%CI)    No association with abnormal DLCO (p=0.07) | 1. No  2. Yes  Stanojevic S, Am J Respir Crit Care Med, 2008; Wanger J, Eur Respir J, 2005; Weng TR, Am Rev Respir Dis, 1969; Pellegrino R, Eur Respir J, 2005; reference equations from Sick Children  3. No  4. No  5. No  6. No | Retrospective cross-sectional  SB: high risk  AB: low risk  DB: low risk  CF: Unclear |
|  | Mittal 2021  (17) | | | 119 Hodgkin lymphoma CCS with DLCO | Median 10.3yr (6.04-16.8) | 100% | DLCO in CCS exposed to <80mg/m2 vs. >80mg/m2 bleomycin | OR (95%CI)  OR 2.12 (95%CI 0.99 – 4.49), p=0.051 | 1. Yes  2. Yes  Quanjer, Pellegrino  3. No  4. Yes (ERS/ATS)  5. No  6. No | Prospective cohort  SB: high risk  AB: low risk  DB: unclear  CF: high risk |
| GRADE assessment: | |  |  | |  | | | | | |
| Study design: | |  | +4 | 2 prospective cohort studies, 1 retrospective cross-sectional study | | | | | | |
| Study limitations: | |  | -2 | Severe limitations: Selection bias high in 2/3, low in 1/2; Attrition bias low in 3/3; Detection bias low in 1/3, unclear in 2/3; Confounding high in 2/3, unclear in 1/2 | | | | | | |
| Consistency: | |  | 0 | No inconsistency. All studies show no significant association between cumulative dose of bleomycin and diffusion capacity impairment. | | | | | | |
| Directness: | |  | -1 | Population and outcomes broadly generalizable; PFT quality unsure (all studies mention reference values, one mentions lung function procedures used) | | | | | | |
| Precision: | |  | -1 | Important imprecision, 2/3 show show p-value only, 1/3 report Odds Ratio and 95%CI, allreport univariable analysis only | | | | | | |
| Publication bias: | |  | 0 | Publication bias unlikely | | | | | | |
| Effect size: | |  | 0 | No large magnitude of effect | | | | | | |
| Dose-response: | |  | 0 | No clear dose response relationship | | | | | | |
| Plausible confounding: | |  | 0 | No plausible confounding | | | | | | |
| Quality of evidence: | | | ⊕⊖⊖⊖ VERY LOW | | | | | | | |
| Conclusion: | | | No significant effect on diffusion capacity impairment after higher doses of bleomycin vs. lower doses of bleomycin in CAYA cancer survivors.  (3 studies; 3 non-significant effects; 299 participants, 242 participants exposed to bleomycin) | | | | | | | |
| Comments: | | | All studies report their results as univariable analysis only. Homogeneous outcome and cutoff definition across all studies. | | | | | | | |

##### 5b Age at exposure

| PICO | Study | No. of participants | Follow-up (median/mean, range) yr | Bleomycin exposure | Pulmonary function Outcomes | Effect size | Risk of bias |
| --- | --- | --- | --- | --- | --- | --- | --- |
| 5b What is the risk in younger compared to older age at treatment? | | | | | | | |

**No study**

#### PICO 6: Busulfan

| PICO | Study | No. of participants | Follow-up (median/mean, range) yr | Busulfan exposure | Pulmonary function Outcomes | Effect size | PFT quality | Risk of bias |
| --- | --- | --- | --- | --- | --- | --- | --- | --- |
| 6 What is the risk of obstructive abnormalities in CAYA treated with busulfan compared to CAYA not treated with busulfan? | | | | | | | | |

**No study**

| PICO | Study | | | | No. of participants | Follow-up (median/mean, range) yr | Busulfan exposure | Pulmonary function Outcomes | Effect size | PFT quality | Risk of bias |
| --- | --- | --- | --- | --- | --- | --- | --- | --- | --- | --- | --- |
| 6 What is the risk of restrictive abnormalities in CAYA treated with busulfan compared to CAYA not treated with busulfan? | Armenian, 2015 (14) | | | | 121 CCS | Median 17.1 (6.3-40.1) | 15 (12.4%) | 29 Restrictive  (TLC<75% and FEV1≥80% predicted)  13% (3/15) busulfan  24% (26/106) no busulfan | Univariable logistic regression  Odds Ratio (95%CI)  0.8 (0.2-2.9) | 1. Yes  2. No  3. No  4. Yes: ATS, Miller MR, Eur Respir J, 2005  5. No  6. Yes | Prospective cohort  SB: Low risk  AB: low risk  DB: low risk  CF: high risk |
| GRADE assessment: | |  |  |  | | | | | | | |
| Study design: | |  | +4 | Prospective cohort study | | | | | | | |
| Study limitations: | |  | -1 | Some limitations: Selection bias low in 1/1; Attrition bias low in 1/1; Detection bias low in 1/1; Confounding high in 1/1 | | | | | | | |
| Consistency: | |  | 0 | One study only | | | | | | | |
| Directness: | |  | -1 | Population and outcomes broadly generalizable; PFT quality unsure (no references mentioned, lung function procedure mentioned) | | | | | | | |
| Precision: | |  | -1 | One study only, univariable comparison, results shown as OR and 95%CI | | | | | | | |
| Publication bias: | |  | 0 | Unlikely | | | | | | | |
| Effect size: | |  | 0 | No large magnitude of effect | | | | | | | |
| Dose-response: | |  | 0 | Not applicable | | | | | | | |
| Plausible confounding: | |  | 0 | No plausible confounding | | | | | | | |
| Quality of evidence: | | ⊕⊖⊖⊖ VERY LOW | | | | | | | | | |
| Conclusion: | | No significant effect on restrictive abnormalities (TLC<75% and FEV1≥80% predicted) after busulfan vs. no busulfan in CAYA cancer survivors.  (1 study; 121 participants; 15 participants exposed to busulfan) | | | | | | | | | |
| Conclusion: | | Only univariable comparison between CCS exposed to busulfan and not; small sample size exposed to busulfan (12%) | | | | | | | | | |

| PICO | Study | No. of participants | Follow-up (median/mean, range) yr | Busulfan exposure | Pulmonary function Outcomes | Effect size | PFT quality | Risk of bias |
| --- | --- | --- | --- | --- | --- | --- | --- | --- |
| 6 What is the risk of hyperinflation in CAYA treated with busulfan compared to CAYA not treated with busulfan? | | | | | | | | |

**No study**

| PICO | Study | | | | No. of participants | Follow-up (median/mean, range) yr | Busulfan exposure | Pulmonary function Outcomes | Effect size | PFT quality | Risk of bias |
| --- | --- | --- | --- | --- | --- | --- | --- | --- | --- | --- | --- |
| 6 What is the risk of diffusion capacity impairment in CAYA treated with busulfan compared to CAYA not treated with busulfan? | Armenian, 2015 (13) | | | | 121 CCS | Median 17.1 (6.3-40.1) | 15 (12.4%) | 42 Diffusion impairment  (DLCOcorr<75% predicted)  13% (3/15) busulfan  37% (39/106) no busulfan | Univariable logistic regression  Odds Ratio (95%CI)  0.4 (0.1-1.6) | 1. Yes  2. No  3. No  4. Yes: ATS, Miller MR, Eur Respir J, 2005  5. No  6. Yes | Prospective cohort  SB: Low risk  AB: low risk  DB: low risk  CF: high risk |
| GRADE assessment: | |  |  | | |  | | | | | |
| Study design: | |  | +4 | Prospective cohort study | | | | | | | |
| Study limitations: | |  | -1 | Some limitations: Selection bias low in 1/1; Attrition bias low in 1/1; Detection bias low in 1/1; Confounding high in 1/1 | | | | | | | |
| Consistency: | |  | 0 | One study only | | | | | | | |
| Directness: | |  | -1 | Population and outcome broadly generalizable; PFT quality unsure (no references mentioned, lung function procedure mentioned) | | | | | | | |
| Precision: | |  | -1 | One study only, univariable comparison, results shown as OR and 95%CI | | | | | | | |
| Publication bias: | |  | 0 | Unlikely | | | | | | | |
| Effect size: | |  | 0 | No large magnitude of effect | | | | | | | |
| Dose-response: | |  | 0 | Not applicable | | | | | | | |
| Plausible confounding: | |  | 0 | No plausible confounding | | | | | | | |
| Quality of evidence: | | | ⊕⊖⊖⊖ VERY LOW | | | | | | | | |
| Conclusion: | | | No significant effect on diffusion capacity impairment (DLCO) after busulfan vs. no busulfan in CAYA cancer survivors.  (1 study; 121 participants; 15 participants exposed to busulfan) | | | | | | | | |
| Conclusion: | | | Only univariable comparison between CCS exposed to busulfan and not exposed | | | | | | | | |

##### 6a Different doses

| PICO | Study | No. of participants | Follow-up (median/mean, range) yr | Busulfan exposure | Pulmonary function Outcomes | Effect size | Risk of bias |
| --- | --- | --- | --- | --- | --- | --- | --- |
| 6a What is the risk associated with different doses? | | | | | | | |

**No study**

##### 6b Age at exposure

| PICO | Study | No. of participants | Follow-up (median/mean, range) yr | Busulfan exposure | Pulmonary function Outcomes | Effect size | Risk of bias |
| --- | --- | --- | --- | --- | --- | --- | --- |
| 6b What is the risk in younger compared to older age at treatment? 🡪 No study | | | | | | | |

**No study**

#### PICO 7: Nitrosureas

| PICO | Study | No. of participants | Follow-up (median/mean, range) yr | Nitrosurea exposure | Pulmonary function Outcomes | Effect size | PFT quality | Risk of bias |
| --- | --- | --- | --- | --- | --- | --- | --- | --- |
| 7 What is the risk of obstructive abnormalities in CAYA treated with nitrosureas compared to CAYA not treated with nitrosureas? | | | | | | | | |

**No study**

| PICO | Study | | | No. of participants | Follow-up (median/mean, range) yr | Nitrosurea exposure | Pulmonary function Outcomes | Effect size | PFT quality | Risk of bias |
| --- | --- | --- | --- | --- | --- | --- | --- | --- | --- | --- |
| 7 What is the risk of restrictive abnormalities in CAYA treated with nitrosureas compared to CAYA not treated with nitrosureas? | Armenian, 2015 (13) | | | 121 CAYA | Median 17.1 yrs (6.3-40.1 yrs) | 9.9% | Total 29 restrictive  (TLC<75% and FEV1≥80% predicted)  25% (3/12) nitrosurea  24% (26/109) no nitrosurea | Univariable logistic regression  Odds Ratio (95%CI)  1.1 (0.3-4.2) | 1. Yes  2. No  3. No  4. Yes: ATS  5. No  6. Yes | Prospective cohort  SB: Low risk  AB: low risk  DB: low risk  CF: high risk |
| GRADE assessment: | |  |  |  | | | | | | |
| Study design: | |  | +4 | 1 prospective cohort study | | | | | | |
| Study limitations: | |  | -1 | Some limitations: Selection bias low in 1/1; Attrition bias low in 1/1; Detection bias low in 1/1; Confounding high in 1/1 | | | | | | |
| Consistency: | |  | 0 | One study only | | | | | | |
| Directness: | |  | -1 | Population and outcomes broadly generalizable, PFT quality unsure (no reference mentioned, lung function procedure mentioned) | | | | | | |
| Precision: | |  | -1 | One study only, univariable analysis, results shown as OR with 95%CI | | | | | | |
| Publication bias: | |  | 0 | Unlikely | | | | | | |
| Effect size: | |  | 0 | No large magnitude of effect | | | | | | |
| Dose-response: | |  | 0 | Not applicable | | | | | | |
| Plausible confounding: | |  | 0 | No plausible confounding | | | | | | |
| Quality of evidence: | | | ⊕⊖⊖⊖ Very low | | | | | | | |
| Conclusion: | | | No significant effect on restrictive abnormalities (TLC and FEV1) after nitrosureas vs. no nitrosureas in CAYA cancer survivors.  (1 study; 121 participants; 12 exposed to nitrosureas) | | | | | | | |
| Comment: | | | Only one univariable comparison, small sample size exposed to nitrosureas | | | | | | | |

| PICO | Study | No. of participants | Follow-up (median/mean, range) yr | Nitrosurea exposure | Pulmonary function Outcomes | Effect size | PFT quality | Risk of bias |
| --- | --- | --- | --- | --- | --- | --- | --- | --- |
| 7 What is the risk of hyperinflation in CAYA treated with nitrosureas compared to CAYA not treated with nitrosureas? | | | | | | | | |

**No study**

| PICO | Study | | | No. of participants | | Follow-up (median/mean, range) yr | Nitrosurea exposure | Pulmonary function Outcomes | Effect size | PFT quality | Risk of bias |
| --- | --- | --- | --- | --- | --- | --- | --- | --- | --- | --- | --- |
| 7 What is the risk of diffusion capacity impairment in CAYA treated with nitrosureas compared to CAYA not treated with nitrosureas? | Armenian, 2015 (13) | | | 121 CAYA | | Median 17.1 yrs (6.3-40.1 yrs) | 9.9% | Total 42 diffusion abnormality  (DLCO<75% predicted)  42% (5/12) nitrosureas  34% (37/109) no nitrosureas | Univariable logistic regression  Odds Ratio (95%CI)  0.4 (0.6-4.7) | 1. Yes  2. No  3. No  4. Yes: ATS  5. No  6. Yes | Prospective cohort  SB: Low risk  AB: low risk  DB: low risk  CF: high risk |
| *GRADE assessment:* | |  |  | |  | | | | | | |
| Study design: | |  | +4 | 1 prospective cohort study | | | | | | | |
| Study limitations: | |  | -1 | Some limitations: Selection bias low in 1/1; Attrition bias low in 1/1; Detection bias low in 1/1; Confounding high in 1/1 | | | | | | | |
| Consistency: | |  | 0 | One study only | | | | | | | |
| Directness: | |  | -1 | Population and outcomes broadly generalizable, PFT quality unsure (no reference mentioned, lung function procedure mentioned) | | | | | | | |
| Precision: | |  | -1 | One study only, univariable analysis, results shown as OR with 95%CI | | | | | | | |
| Publication bias: | |  | 0 | Unlikely | | | | | | | |
| Effect size: | |  | 0 | No large magnitude of effect | | | | | | | |
| Dose-response: | |  | 0 | Not applicable | | | | | | | |
| Plausible confounding: | |  | 0 | No plausible confounding | | | | | | | |
| Quality of evidence: | | | ⊕⊖⊖⊖ Very low | | | | | | | | |
| Conclusion: | | | No significant effect on diffusion capacity impairment (DLCO) after nitrosureas vs. no nitrosureas in CAYA cancer survivors.  (1 study; 121 participants; 12 exposed to nitrosureas) | | | | | | | | |
| Comment: | | | Only one univariable comparison, small sample size exposed to nitrosureas | | | | | | | | |

##### 7a Different doses

| PICO | Study | No. of participants | Follow-up (median/mean, range) yr | Nitrosurea exposure | Pulmonary function Outcomes | Effect size | Risk of bias |
| --- | --- | --- | --- | --- | --- | --- | --- |
| 7a What is the risk associated with different doses? | | | | | | | |

**No study**

##### 7b Age at exposure

| PICO | Study | No. of participants | Follow-up (median/mean, range) yr | Nitrosurea exposure | Pulmonary function Outcomes | Effect size | Risk of bias |
| --- | --- | --- | --- | --- | --- | --- | --- |
| 7b What is the risk in younger compared to older age at treatment? | | | | | | | |

**No study**

#### PICO 8: Radiotherapy

| PICO | Study | | | | No. of participants | Follow-up (median/mean, range) yr | Radiotherapy exposing lung tissue | Pulmonary function Outcomes | Effect size | PFT quality | Risk of bias |
| --- | --- | --- | --- | --- | --- | --- | --- | --- | --- | --- | --- |
| 8 What is the risk of obstructive abnormalities in CAYA treated with radiotherapy exposing lung tissue compared to CAYA not treated with radiotherapy exposing lung tissue? | Oguz 2007 (18) | | | | 75 Lymphoma survivors | Median  5 (2-13) | Group 1:  Chemo and  Radio (n=23)  Group 2:  Chemo only (n=52) | Mean (±SD) of selected % predicted values  **FEV1**  Group 1: 95.43 (± 16.47)  Group 2: 105.09 (± 19.01)  **FEV1/FVC**  Group 1: 96.43 (± 9.15)  Group 2: 99.88 (± 11.93) | Comparison  Group I vs Group II (student t-test)    p=0.038  p=0.221 | 1. No  2. Yes:  References recommended by European Coal and Steel Community;  Severity acc. to ATS pulmonary function laboratory guidelines  3. No  4. No  5. No  6. No | Retrospective cross-sectional  SB unclear  AB: low risk  DB: unclear  CF: unclear |
|  | Jenney 1995 (8) | | | | 70 Leukemia survivors | Median 4.2 (0.6-18.5) | 14% (CSI, n=10)  20% (TBI, n=14) | Number of CCS with respective parameter below predicted values  36/69 FEV1 <85% predicted  23/69 FEV1 <80% predicted | Multivariable analysis, CSI (yes/no) leads to reduction in FEV1: p<0.001 | 1. Yes  2. No  3. No  4. No  5. No  6. No | Prospective cross-sectional  SB: high risk  AB: low risk  DB: unclear  CF: unclear |
|  | Record 2016 (1) | | | | 143 CCS | Mean 14.1 ±4.8 | 67.8%  (n=97) | Obstructive  (FVC, FEV1, FEV1/FVC <80% predicted or FEF25–75% <68%)  25% (24/97)radiotherapy  28% (13/46) no radiotherapy | Univariable comparison Chi2 radiation yes/no  p=0.66 | 1. No  2. Yes: Wang X, Pediatr Pulmonol 2005; Hankinson JL, Am J Respir Crit Care Med 1999  3. No  4. Yes  5. No  6. Yes | Retrospective cohort  SB: high risk  AB: low risk  DB: low risk  CF: high risk |
|  | Stone 2020  (19) | | | | 62 high-risk neuroblastoma | Median 5.29 (0.24-15.24) | 34%  (n=21) | Comparison of CCS treated with radiotherapy versus no radiotherapy  **FEV1**  (FEV1 <80% pred)  RT yes: 71.4% abnormal  RT no: 34.2% abnormal | OR, 95%CI  4.29 (1.35 – 13.58), p=0.005 | 1. No  2. Yes  3. No  4. Yes (ATS)  5. No  6. No | Prospective cohort  SB: high risk  AB: low risk  DB: low risk  CF: high risk |
|  | Otth 2021  (20) | | | | 72 CCS exposed to HSCT | Median 9.4  (6.1 – 12.3) | 70%  (n=52) | **FEV1**  Effect of radiotherapy vs. no radiotherapy on longitudinal changes in FEV1 (intercept)  **MMEF**  Effect of radiotherapy vs. no radiotherapy on longitudinal changes in MMEF (intercept) | mixed effects multivariable linear  regression analysis  Coefficient -1.306  95%CI -2.055 - -0.558 p=0.001  Coefficient -0.664  95%CI -1.583 – 0.253 p=0.156 | 1. Yes  2. Yes  3. No  4. No  5. Yes  6. No | Retrospective cohort  SB: high risk  AB: low risk  DB: low risk  CF: low risk |
| GRADE assessment: | |  |  |  | | | | | | | |
| Study design: | |  | +4 | 2 retrospective cohort study, 1 retrospective cross-sectional study, 1 prospective cross-sectional study, 1 prospective cohort study | | | | | | | |
| Study limitations: | |  | -2 | Some limitations: Selection bias high in 4/5, unclear in 1/3; Attrition bias low in 4/5; Detection bias low in 3/5, unclear in 2/3; Confounding high in 2/5, low in 1/5, unclear in 2/5 | | | | | | | |
| Consistency: | |  | -1 | Some inconsistency. Four studies show significant effect of radiotherapy exposing lung tissue on FEV1, one study on MMEF, no significant association for FEV1/FVC and a non-significant inverse effect on “obstructive”, where non-exposed CAYA cancer survivors show more often obstructive abnormalities than exposed | | | | | | | |
| Directness: | |  | -1 | Population and outcomes broadly generalizable, PFT quality unsure (reference mentioned in 5/5, lung function procedure mentioned in 0/5) | | | | | | | |
| Precision: | |  | -1 | Important imprecision, 3/5 report p-values only, 2/5 report 95%CI, 1/5 performed univariable regression analysis, 2/5 performed multivariable analysis | | | | | | | |
| Publication bias: | |  | 0 | Unlikely | | | | | | | |
| Effect size: | |  | 0 | No large magnitude of effect | | | | | | | |
| Dose-response: | |  | 0 | Not applicable | | | | | | | |
| Plausible confounding: | |  | 0 | No plausible confounding | | | | | | | |
| Quality of evidence: | | | ⊕⊖⊖⊖ Very low | | | | | | | | |
| Conclusion: | | | Increased risk for obstructive abnormalities (FEV1, MMEF) after radiotherapy exposing the lung tissue vs. no radiotherapy in CAYA cancer survivors.  (5 studies; 3 studies significant effect [FEV1] and 1 study on MMEF, 1 study non-significant effect [“obstructive]; 422 participants; 217 exposed to radiotherapy exposing lung tissue) | | | | | | | | |
| Comment: | | | Outcome assessed differently (FEV1, MMEF, “obstructive”) and cutoff values differ between studies. | | | | | | | | |

| PICO | Study | | | No. of participants | | Follow-up (median/mean, range) yr | Radiotherapy exposing lung tissue | Pulmonary function Outcomes | Effect size | PFT quality | Risk of bias |
| --- | --- | --- | --- | --- | --- | --- | --- | --- | --- | --- | --- |
| 8 What is the risk of restrictive abnormalities in CAYA treated with radiotherapy exposing lung tissue compared to CAYA not treated with radiotherapy exposing lung tissue? | Oguz 2007 (17) | | | 75 Lymphoma survivors | | Median  5 (2-13) | Group 1:  Chemo and  Radio (n=23)  Group 2:  Chemo only (n=52) | Mean (±SD) of selected % predicted values  **FVC**  Group 1: 101.17 (± 19.93)  Group 2: 102.94 (± 18.11)  **TLC**  Group 1: 102.74 (± 15.63)  Group 2: 106.73 (± 17.46) | Comparison  Group I vs Group II (student t-test)    p=0.706  p=0.349 | 1. No  2. Yes:  references recommended by European Coal and Steel Community; Severity acc. to ATS pulmonary function laboratory guidelines  3. No  4. No  5. No  6. No | Retrospective cross-sectional  SB unclear  AB: low risk  DB: unclear  CF: unclear |
|  | Jenney 1995 (8) | | | 70 Leukemia survivors | | Median 4.2 (0.6-18.5) | 14% (CSI, n=10)  20% (TBI, n=14) | Number of CCS with respective parameter below predicted values  32/69 FVC <85% predicted  26/69 TLC <85% predicted  20/69 FVC <80% predicted  20/69 TLC <80% predicted | Multivariable analysis, CSI leads to reduction in FVC and TLC: p<0.001 | 1. Yes  2. No  3. No  4. No  5. No  6. No | Prospective cross-sectional  SB: high risk  AB: low risk  DB: unclear  CF: unclear |
|  | Record 2016 (1) | | | 143 CCS | | Mean 14.1 ±4.8 yrs | 67.8%  (n=97) | Restrictive  (TLC<80% predicted)  11% (11/97) radiotherapy  17% (8/46) no radiotherapy | Univariable comparison Chi2 radiation yes/no  p=0.33 | 1. No  2. Yes  Wang X, Pediatr Pulmonol 2005; Hankinson JL, Am J Respir Crit Care Med 1999  3. No  4. Yes  5. No  6. Yes | Retrospective cohort  SB: high risk  AB: low risk  DB: low risk  CF: high risk |
|  | Mulder 2011 (9) | | | 193 CCS | | Median 17.9 (5.6-36.8) | 40.9%  (n=79) | Total 28 restrictive  (TLC OR FVC <75%)  Of those Exposed: 35% | Odds Ratio (95%CI) for radiotherapy yes/no  12.87 (3.37-49.08) | 1. No  2. No  3. No  4. No  5. No  6. No | Retrospective cohort  SB: low risk  AB: low risk  DB: unclear  CF: low risk |
|  | Stone 2020  (19) | | | 62 high-risk neuroblastoma | | Median 5.29 (0.24-15.24) | 34%  (n=21) | **FVC**  (FVC <80% pred)  RT yes: 76.2% abnormal  RT no: 41.7% abnormal  **TLC**  (TLC <80% pred)  RT yes: 66.7% abnormal  RT no: 35.4% abnormal | OR, 95%CI  4.40 (1.34 – 14.51) p=0.010  4.33 (1.39 – 13.50), p=0.005 | 1. No  2. Yes  3. No  4. Yes (ATS)  5. No  6. No | Prospective cohort  SB: high risk  AB: low risk  DB: low risk  CF: high risk |
|  | Otth 2021  (20) | | | 72 CCS exposed to HSCT | | Median 9.4  (6.1 – 12.3) | 70%  (n=52) | **FVC**  Effect of radiotherapy vs. no radiotherapy on longitudinal changes in FVC  **TLC**  Effect of radiotherapy vs. no radiotherapy on longitudinal changes in TLC | mixed effects multivariable linear  regression analysis  Coefficient -1.473  95%CI -2.207 –  -0.739  p=<0.001  Coefficient -0.717 95%CI -2.051 – 0.616; p=0.292 | 1. Yes  2. Yes  3. No  4. No  5. Yes  6. No | Retrospective cohort  SB: high risk  AB: low risk  DB: low risk  CF: low risk |
| GRADE assessment: | |  |  | |  | | | | | | |
| Study design: | |  | +4 | 3 retrospective cohort studies, 1 retrospective cross-sectional study, 1 prospective cross-sectional study, 1 prospective cohort study | | | | | | | |
| Study limitations: | |  | -2 | Some limitations: Selection bias high in 4/6, low in 1/6, unclear in 1/6; Attrition bias low in 6/6; Detection bias low in 3/6, unclear in 3/6; Confounding high in 3/6, low in 1/6, unclear in 2/6 | | | | | | | |
| Consistency: | |  | 0 | Some inconsistency. Four studies show significant effect of radiotherapy exposing lung tissue on restrictive parameter (FVC, TLC, “restrictive”), one has only small sample of exposed CAYA survivors and the second has a large confidence interval. No significant association in the other two studies. | | | | | | | |
| Directness: | |  | -1 | Population and outcomes broadly generalizable, PFT quality unsure (reference mentioned in 4/6, lung function procedure mentioned in 2/6) | | | | | | | |
| Precision: | |  | -1 | Important imprecision, precision cannot be judged as 3/6 report p-values only, 3/6 shows OR and 95%CI but some with large confidence interval, 2/6 performed multivariable analysis, 1/6 performed univariable regression analysis | | | | | | | |
| Publication bias: | |  | 0 | Unlikely | | | | | | | |
| Effect size: | |  | 1 | Large magnitude of effect in one study | | | | | | | |
| Dose-response: | |  | 0 | Not applicable | | | | | | | |
| Plausible confounding: | |  | 0 | No plausible confounding | | | | | | | |
| Quality of evidence: | | | ⊕⊖⊖⊖ Very low | | | | | | | | |
| Conclusion: | | | Increased risk for restrictive abnormalities (FVC or TLC) after radiotherapy exposing lung tissue vs. no radiotherapy in CAYA cancer survivors.  (6 studies; 4 studies significant effect, 2 studies non-significant effect; 617 participants; 296 exposed to radiotherapy exposing lung tissue) | | | | | | | | |
| Comment: | | | 3/6 studies show p-value only. Outcome and cutoff values differ between the studies. | | | | | | | | |

| PICO | Study | | | No. of participants | | | Follow-up (median/mean, range) yr | Radiotherapy exposing lung tissue | Pulmonary function Outcomes | Effect size | PFT quality | Risk of bias |
| --- | --- | --- | --- | --- | --- | --- | --- | --- | --- | --- | --- | --- |
| 8 What is the risk of hyperinflation in CAYA treated with radiotherapy exposing lung tissue compared to CAYA not treated with radiotherapy exposing lung tissue? | Oguz 2007 (17) | | | 75 Lymphoma survivors | | | Median  5 (2-13) | Group 1:  Chemo and  Radio (n=23)  Group 2:  Chemo only (n=52) | Mean (±SD) of selected % predicted values  **RV**  Group 1: 113.35 (± 28.53)  Group 2: 126.71 (± 24.63)  **RV/TLC**  Group 1: 25.39 (± 5.31)  Group 2: 27.71 (± 4.92) | Comparison  Group I vs Group II (student t-test)    p=0.043    p=0.062 | 1. No  2. Yes:  references recommended by European Coal and Steel Community; Severity acc. to ATS pulmonary function laboratory guidelines  3. No  4. No  5. No  6. No | Retrospective cross-sectional  SB unclear  AB: low risk  DB: unclear  CF: unclear |
|  | Record 2016 (1) | | | 143 CCS | | | Mean 14.1 ±4.8 | 67.8%  (n=97) | hyperinflation  (RV >120%pred or RV/TLC >28% pred)  46% (45/97) radiotherapy  30% (14/46) no radiotherapy | Univariable comparison Chi2 radiation yes/no  p=0.07 | 1. No  2. Yes: Wang X, Pediatr Pulmonol 2005; Hankinson JL, Am J Respir Crit Care Med 1999  3. No  4. Yes  5. No  6. Yes | Retrospective cohort  SB: high risk  AB: low risk  DB: low risk  CF: high risk |
|  | Otth 2021  (20) | | | 72 CCS exposed to HSCT | | | Median 9.4  (6.1 – 12.3) | 70%  (n=52) | **RV**  Effect of radiotherapy vs. no radiotherapy on longitudinal changes in TLC | mixed effects multivariable linear  regression analysis  Coefficient 0.663 95%CI -0.307 – 1.634; p=0.181 | 1. Yes  2. Yes  3. No  4. No  5. Yes  6. No | Retrospective cohort  SB: high risk  AB: low risk  DB: low risk  CF: low risk |
| GRADE assessment: | |  |  | | |  | | | | | | |
| Study design: | |  | +4 | | 1 retrospective cohort studies, 1 retrospective cross-sectional study, 1 retrospective cohort study | | | | | | | |
| Study limitations: | |  | -2 | | Some limitations: Selection bias high in 2/3, unclear in 1/3; Attrition bias low in 3/3; Detection bias low in 2/3, unclear in 1/3; Confounding high in 1/3, low in 1/3, unclear in 1/3 | | | | | | | |
| Consistency: | |  | 0 | | Some inconsistency between studies. Two studies show an association between hyperinflation and radiotherapy exposing the lung tissue. One study shows no association between exposure and longitudinal changes. | | | | | | | |
| Directness: | |  | -1 | | Population and outcomes broadly generalizable, PFT quality unsure (reference mentioned in 3/3, lung function procedure mentioned in 1/3) | | | | | | | |
| Precision: | |  | -1 | | Important imprecision, precision cannot be judged as 2/2 report p-values only and 2/2 performed unviable analysis only | | | | | | | |
| Publication bias: | |  | 0 | | Unlikely | | | | | | | |
| Effect size: | |  | 0 | | No large magnitude of effect | | | | | | | |
| Dose-response: | |  | 0 | | Not applicable | | | | | | | |
| Plausible confounding: | |  | 0 | | No plausible confounding | | | | | | | |
| Quality of evidence: | | | ⊕⊖⊖⊖ Very low | | | | | | | | | |
| Conclusion: | | | Increased risk for hyperinflation (RV, RV/TLC) after radiotherapy exposing the lung tissue vs. no radiotherapy in CAYA cancer survivors in 2 studies and a trend in one study  (3 studies; 292participants; 172participants with radiotherapy exposing the lung tissue) | | | | | | | | | |
| Comment: | | | 2/3 studies reported p-values only with a trend is towards more hyperinflation in exposed CAYA cancer survivors. Outcome definition differs between the studies. | | | | | | | | | |

| PICO | Study | | | | No. of participants | | Follow-up (median/mean, range) yr | Radiotherapy exposing lung tissue | Pulmonary function Outcomes | Effect size | PFT quality | Risk of bias |
| --- | --- | --- | --- | --- | --- | --- | --- | --- | --- | --- | --- | --- |
| 8 What is the risk of diffusion capacity impairment in CAYA treated with radiotherapy exposing lung tissue compared to CAYA not treated with radiotherapy exposing lung tissue? | Oguz 2007 (17) | | | | 75 Lymphoma survivors | | Median  5 (2-13) | Group 1:  Chemo and  Radio (n=23)  Group 2:  Chemo only (n=52) | Mean (±SD) of selected % predicted values  DLCO  Group 1: 101.35 (± 22.17)  Group 2: 112.65 (± 4.92) | Comparison  Group I vs Group II (student t-test)    p=0.025 | 1. No  2. Yes:  references recommended by European Coal and Steel Community; Severity acc. to ATS pulmonary function laboratory guidelines  3. No  4. No  5. No  6. No | Retrospective cross-sectional  SB unclear  AB: low risk  DB: unclear  CF: unclear |
|  | Jenney 1995 (8) | | | | 70 Leukemia survivors | | Median 4.2 (0.6-18.5) | 14% (CSI, n=10)  20% (TBI, n=14) | Number of CCS with respective parameter below predicted values  29/69 DLCO <85% predicted  19/69 DLCO <80% predicted | Multivariable analysis, CSI leads to reduction in DLCO: p<0.030 | 1. Yes  2. No  3. No  4. No  5. No  6. No | Prospective cross-sectional  SB: high risk  AB: low risk  DB: unclear  CF: unclear |
|  | Zorzi 2015 (16) | | | | 143 CCS (Hodgkin, extracranial germ cell tumor) | | Median 4.4  (2 – 7.4) | 60%  (n=86) | 19% (27/143) with abnormal DLCO  (DLCO <80%) | No association (p=0.83) | 1. No  2. Yes  Stanojevic S, Am J Respir Crit Care Med, 2008; Wanger J, Eur Respir J, 2005; Weng TR, Am Rev Respir Dis, 1969; Pellegrino R, Eur Respir J, 2005; reference equations from Sick Children  3. No  4. No  5. No  6. No | Retrospective  cross-sectional  SB: high risk  AB: low risk  DB: low risk  CF: unclear |
|  | Mulder 2011 (9) | | | | 193 CCS | | Median 17.9 (5.6-36.8) | 40.9%  (n=79) | 75/188 Diffusion impairment  (DLCO <75%) | Odds Ratio (95%CI) for radiotherapy yes/no  5.84 (1.88-18.14) | 1. No  2. No  3. No  4. No  5. No  6. No | Retrospective cohort  SB: low risk  AB: low risk  DB: unclear  CF: low risk |
|  | Stone 2020  (19) | | | | 62 high-risk neuroblastoma | | Median 5.29 (0.24-15.24) | 34%  (n=21) | **DLCO**  (DLCO <80% pred)  RT yes: 2.4% abnormal  RT no: 66.7% abnormal | OR, 95%CI  2.05 (0.49 – 8.62), p=0.339 | 1. No  2. Yes  3. No  4. Yes (ATS)  5. No  6. No | Prospective cohort  SB: high risk  AB: low risk  DB: low risk  CF: high risk |
|  | Otth 2021  (20) | | | | 72 CCS exposed to HSCT | | Median 9.4  (6.1 – 12.3) | 70%  (n=52) | **DLCO**  Effect of radiotherapy vs. no radiotherapy on longitudinal changes in DLCO | mixed effects multivariable linear  regression analysis  Coefficient -1.279  95%CI -2.773 - 0.213; p=0.093 | 1. Yes  2. Yes  3. No  4. No  5. Yes  6. No | Retrospective cohort  SB: high risk  AB: low risk  DB: low risk  CF: low risk |
| GRADE assessment: | |  |  | | |  | | | | | | |
| Study design: | |  | +4 | 2 retrospective cohort study, 2 retrospective cross-sectional studies, 1 prospective cross-sectional study, 1 prospective cohort study | | | | | | | | |
| Study limitations: | |  | -2 | Some limitations: Selection bias high in 4/6, low in 1/4, unclear in 1/4; Attrition bias low in 6/6; Detection bias low in 3/6, unclear in 3/6; Confounding low in 2/6, unclear in 4/6 | | | | | | | | |
| Consistency: | |  | 0 | No important inconsistency. Most studies show diffusion capacity impairment in CAYA cancer survivors exposed to radiotherapy exposing lung tissue compared to CAYA cancer survivors not exposed. | | | | | | | | |
| Directness: | |  | -1 | Population and outcomes broadly generalizable, PFT quality unsure (reference mentioned in 4/6, lung function procedure mentioned in 1/6) | | | | | | | | |
| Precision: | |  | -1 | Important imprecision, in 3/6 as results are shown with p-value only, 1/6 has large confidence interval, 2/6 perforemd multivariable analysis, 1/5 performed univariable regression analysis | | | | | | | | |
| Publication bias: | |  | 0 | Unlikely | | | | | | | | |
| Effect size: | |  | 0 | No large magnitude of effect | | | | | | | | |
| Dose-response: | |  | 0 | Not applicable | | | | | | | | |
| Plausible confounding: | |  | 0 | No plausible confounding | | | | | | | | |
| Quality of evidence: | | | ⊕⊖⊖⊖ Very low | | | | | | | | | |
| Conclusion: | | | Increased risk for diffusion capacity impairment (DLCO) after radiotherapy exposing the lung tissue vs. no radiotherapy in CAYA cancer survivors.  (6 studies; 3 studies significant effect, 3 study non-significant effect; 617participants; 296 participants with radiotherapy exposing the lung tissue) | | | | | | | | | |
| Comment: | | | Three studies show p-value only. Outcome definition and cutoff values differ between the studies. | | | | | | | | | |

##### 8a Different doses

| PICO | Study | | | | No. of participants | | Follow-up (median/mean, range) yr | Radiotherapy exposing lung tissue | Pulmonary function Outcomes | Effect size | PFT quality | Risk of bias |
| --- | --- | --- | --- | --- | --- | --- | --- | --- | --- | --- | --- | --- |
| 8a What is the risk for obstructive abnormalities associated with different doses and volumes of radiotherapy?  - dose-volume relationship  - impact of dose per fraction | Weiner 2006 (21) | | | | 30 CSS (Wilms tumor, Hodgkin disease, Sarcoma, Hepato-blastoma) | | Median 2.79 (range 0-13.7) | 100%  (n=30) | - FEV1 z-score  - FEV1 z-score | No correlation between severity of abnormal FEV1 z-score and total radiation dose:   - r^2^=0.002   (very weak)  No correlation between severity of abnormal FEV1 z-score and total radiation dose after taking body length into account:   - r^2^=0.002   (very weak) | 1. No  2. Yes:  Wang X, Pediatr Pulmonol, 1993; Rosenthal M, Thorax 1993  3. No  4. Yes  5. No  6. No | Retrospective cohort  SB: High Risk  AB: High risk  DB: Low risk  CF: High Risk |
|  | Green 2016 (22) | | | | 606 CCS (FEV1, FVC)  597 CCS  (TLC, DLCO) | | Median 21.9 | 76.7%  (n=465) | Proportion of CCS with pulmonary function parameter below %pred or LLN for whole cohort  51% FEV1 <80% pred  49% FEV1 <LLN | Multivariable log-binomial regression:  Outcome: V10 (per 10% increase)  Relative Risk (95%CI, p-value)  1.07 (1.04–1.09, <0.001)  1.06 (1.04-1.09, <0.001) | 1. No  2. Yes  Wanger J, Eur Respir J, 2005; Goldman HI, Am Rev Tuberc, 1959; Boren HG, Am J Med, 1966; Miller A, Am Rev Respir Dis, 1983; Quanjer PH, lookup table, accessed 2015; Quanjer PH, Eur Respir J, 2012  3. Yes  4. Yes: ATS  5. No  6. No | Prospective cohort  SB: High risk  AB: Low risk  DB: Unclear  CF: Low risk |
|  | De 2015 (12) | | | | 49  Osteo  sarcoma | | Median 2.91 (range 0.01-8.28) | 100%  (n=49) | Proportion of CCS with abnormal results per lung function parameter  FEV1 <80% pred: 29% (14/49)  FEF25–75% <68% pred: 20% (10/49)  Obstructive disease  (FEV1/FVC <80%, FEV1<80% or FEF25-75<68% with normal TLC): 24% (12/49) | Logistic regression with radiation dose in Gy (cont.) and normal/ abnormal parameter  Odds Ratio (p-value)  Mean dose: 1.20; 0.01  Max dose: 1.12; <0.01  Mean dose: 1.18; <0.01  Max dose: 1.06; <0.05  Mean dose: 0.99; NS  Max dose: 1.03; NS  Prescribed dose: 1.05; NS | 1. No  2. Yes  Hankinson JL, Am J Respir Crit Care Med, 1999;  Wang X, Pediatr Pulmonol, 1993  3. No  4. Yes: ATS  5. No  6. No | Retrospective cohort  SB: High Risk  AB: High Risk  DB: Low risk  CF: High Risk |
| GRADE assessment: | |  |  | | |  | | | | | | |
| Study design: | |  | +4 | 2 retrospective cohort studies, 1 prospective cohort study | | | | | | | | |
| Study limitations: | |  | -3 | Some limitations: Selection bias high in 3/3; Attrition bias high in 2/3, low in 1/3; Detection bias low in 2/3, unclear in 1/2; Confounding high in 2/3 | | | | | | | | |
| Consistency: | |  | 0 | Most studies show slightly increased risk for indicators of obstructive abnormalities below predicted or LLN with higher doses of radiotherapy. | | | | | | | | |
| Directness: | |  | -1 | Results are direct, population and outcomes broadly generalizable, PFT quality good (in 3/3 reference values and guidelines mentioned) | | | | | | | | |
| Precision: | |  | -1 | Important imprecision 1/3 show precise results with small confidence interval; 1/3 shows correlation coefficient only; 1/3 shows effect size but without 95%CI | | | | | | | | |
| Publication bias: | |  | 0 | Unlikely | | | | | | | | |
| Effect size: | |  | 0 | No large magnitude of effect | | | | | | | | |
| Dose-response: | |  | +1 | Dose-response relationship | | | | | | | | |
| Plausible confounding: | |  | 0 | No evidence of possible confounding | | | | | | | | |
| Quality of evidence: | | | ⊕⊖⊖⊖ Very low | | | | | | | | | |
| Conclusion: | | | Increased risk for obstructive abnormalities (FEV1, FEF25.75%) after increasing doses radiotherapy exposing lung tissue in CAYA cancer survivors  (3 studies; 2 significant effects [FEV1, FEF25.75%], 1 non-significant effect [obstructive, FEV1]; 685 participants; 544 participants exposed to radiotherapy) | | | | | | | | | |
| Comments: | | | One study shows correlation coefficient only | | | | | | | | | |

| PICO | Study | | | | No. of participants | Follow-up (median/mean, range) yr | Radiotherapy exposing lung tissue | Pulmonary function Outcomes | Effect size | PFT quality | Risk of bias |
| --- | --- | --- | --- | --- | --- | --- | --- | --- | --- | --- | --- |
| 8a What is the risk for restrictive abnormalities associated with different doses and volumes of radiotherapy?  - dose-volume relationship  - impact of dose per fraction | Weiner 2006 (18) | | | | 30 CSS (Wilms tumor, Hodgkin disease, Sarcoma,  Hepato-blastoma) | Median 2.79 (range 0-13.7) | 100%  (n=30) | -TLC z-score (n=23)  - TLC z-score (n=23) | No correlation between severity of abnormal FEV1, TCL, and DLCO (z-score) and total radiation dose:   - r^2^=0.06   (very weak)  No correlation between severity of abnormal TCL (z-score) and total radiation dose after taking body length into account:   - r^2^=0.027   (very weak) | 1. No  2. Yes:  Wang X, Pediatr Pulmonol, 1993; Rosenthal M, Thorax 1993  3. No  4. Yes  5. No  6. No | Retrospective cohort  SB: High Risk  AB: High risk  DB: Low risk  CF: High Risk |
|  | Green 2016 (19) | | | | 606 CCS (FEV1, FVC)  597 CCS  (TLC, DLCO) | Median 21.9 | 76.7%  (n=465) | Proportion of CCS with pulmonary function parameter below %pred or LLN in the whole cohort  47.2% FVC <80% pred  45.4% FVC < LLN  31.2% TLC <75% pred | Multivariable log-binomial regression:  Outcome: V10 (per 10% increase)  Relative Risk (95%CI, p-value)  1.08 (1.05–1.11, <0.001)  1.07 (1.04-1.10, <0.001)  1.07 (1.01–1.13, 0.019) | 1. No  2. Yes  Wanger J, Eur Respir J, 2005; Goldman HI, Am Rev Tuberc, 1959; Boren HG, Am J Med, 1966; Miller A, Am Rev Respir Dis, 1983; Quanjer PH, lookup table, accessed 2015; Quanjer PH, Eur Respir J, 2012  3. Yes  4. Yes: ATS  5. No  6. No | Prospective cohort  SB: high risk  AB: low risk  DB: unclear  CF: high risk |
|  | Armenian, 2015 (14) | | | | 121 CAYA | Median 17.1 yrs (6.3-40.1) | 73.6%  (n=89)  Categories:  26.4% No (Ref.)  49.6% ≤20Gy  24.0% >20Gy | Total 29 restrictive  13% (4/32) no radiation  45% (13/60) ≤20Gy,  41% (12/29) >20 Gy | Multivariable logistic regression  Odds Ratio (95%CI)  1  ≤20Gy 1.6 (0.5-5.7)  >20Gy 5.6 (1.5-2.1) | 1. Yes  2. No  3. No  4. Yes: ATS  5. No  6. Yes | Prospective cohort  SB: Low risk  AB: low risk  DB: low risk  CF: high risk |
|  | Green 2015 (10) | | | | 260 embryonal brain tumors | Minimum 2 yr | 100% CSI  (n=260) | Proportion of CCS with TLC below predicted after 60 months  TLC < 75%: 11%  *Unclear how many received proton and photon beam, but of initially 303 eligible patients only 20 had proton beam* | Larger **TLC% predicted**:  photon beam CSI (p=0.002) | 1. No  2. Yes: 10 different references for standardization  3. No  4. Yes: ATS  5. No  6. No | Prospective cohort  SB: low risk  AB: high risk  DB: unclear  CF: high risk |
|  | De 2015 (12) | | | | 49  Osteo  sarcoma | Median 2.91 (range 0.01-8.28) | 100%  (n=49) | Proportion of CCS with abnormal results per lung function parameter  FVC <80% pred: 24% (12/49)  TLC <77% pred: 15% (7/49)  Restrictive (TLC <77%)  15% (7/49) | Logistic regression analysis with radiation dose in Gy (cont.) and normal/ abnormal parameter  Odds Ratio (p-value)  Mean dose: 1.22; <0.01  Max dose: 1.10; <0.01  Mean dose: 1.30; <0.01  Max dose: 1.07; <0.05  The odds of developing restrictive abnormalities increased with increasing V dose beginning at V10  Mean dose: 1.30; <0.01  Max dose: 1.07; <0.05  Prescribed dose: 1.04; NS | 1. No  2. Yes  Hankinson JL, Am J Respir Crit Care Med, 1999;  Wang X, Pediatr Pulmonol, 1993  3. No  4. Yes: ATS  5. No  6. No | Retrospective cohort  SB: High Risk  AB: High Risk  DB: Low risk  CF: High Risk |
| GRADE assessment: | |  |  |  | | | | | | | |
| Study design: | |  | +4 | 2 retrospective cohort studies 3 prospective cohort studies | | | | | | | |
| Study limitations: | |  | -3 | Some limitations: Selection bias high in 3/5, low in 2/5; Attrition bias high in 3/5, low in 2/5; Detection bias low in 3/5, unclear in 2/5; Confounding high in 5/5, | | | | | | | |
| Consistency: | |  | 0 | Most studies show more restrictive abnormalities in CAYA cancer survivors exposed to increasing doses of radiotherapy to the thorax. | | | | | | | |
| Directness: | |  | -1 | Results are direct, population and outcomes broadly generalizable, PFT quality unsure (in 1/5 no reference values stated and in 1/5 10 different references; in 1/5 no guidelines mentioned) | | | | | | | |
| Precision: | |  | -1 | Important imprecision. 2/5 with multivariable analysis; 2/5 with effect size and small confidence intervals; in 2/5 precision cannot be judged as results are shown as coefficient or p-value only | | | | | | | |
| Publication bias: | |  | 0 | Unlikely | | | | | | | |
| Effect size: | |  | 0 | No large magnitude of effect | | | | | | | |
| Dose-response: | |  | +1 | Dose-response relationship | | | | | | | |
| Plausible confounding: | |  | 0 | No evidence of possible confounding | | | | | | | |
| Quality of evidence: | | | ⊕⊖⊖⊖ Very low | | | | | | | | |
| Conclusion: | | | Increased risk for restrictive abnormalities after increasing doses of radiotherapy exposing lung tissue in CAYA cancer survivors.  (5 studies; 1066 participants; 893 participants exposed to radiotherapy) | | | | | | | | |
| Comments: | | | Different cutoff values used between studies to define parameters as abnormal. One study shows correlation coefficient only | | | | | | | | |

| PICO | Study | | | | No. of participants | | Follow-up (median/mean, range) yr | Radiotherapy exposing lung tissue | Pulmonary function Outcomes | Effect size | PFT quality | Risk of bias |
| --- | --- | --- | --- | --- | --- | --- | --- | --- | --- | --- | --- | --- |
| 8a What is the risk for hyperinflation associated with different doses and volumes of radiotherapy?  - dose-volume relationship  - impact of dose per fraction | De 2015 (12) | | | | 49  Osteo-sarcoma | | Median 2.91 (range 0.01-8.28) | 100%  (n=49) | Proportion of CCS with abnormal results per lung function parameter  RV/TLC: >123% pred: 20% (10/49)  Hyperinflation  (RV/TLC >28%): 20% (10/49) | Logistic regression with radiation dose in Gy (cont.) and normal/ abnormal parameter  Odds Ratio (p-value)  Mean dose: 1.30; <0.01  Max dose: 1.26; <0.05  The odds of developing hyperinﬂation increased with increasing V dose beginning at V20  Mean dose: 1.29; <0.01  Max dose: 1.26; <0.01  Prescribed dose: 1.27; <0.01 | 1. No  2. Yes  Hankinson JL, Am J Respir Crit Care Med, 1999;  Wang X, Pediatr Pulmonol, 1993  3. No  4. Yes: ATS  5. No  6. No | Retrospective cohort  SB: High Risk  AB: High Risk  DB: Low risk  CF: High Risk |
| GRADE assessment: | |  |  | | |  | | | | | | |
| Study design: | |  | +4 | 1 retrospective cohort study | | | | | | | | |
| Study limitations: | |  | -3 | Important limitations: Selection bias high in 1/1; Attrition bias high in 1/1; Detection bias low in 1/1; Confounding high in 1/1 | | | | | | | | |
| Consistency: | |  | 0 | One study only | | | | | | | | |
| Directness: | |  | 0 | Population and outcomes broadly generalizable, PFT quality good (reference values and guidelines used stated) | | | | | | | | |
| Precision: | |  | -1 | Important imprecision 1/1 sow results with OR but without 95%CI | | | | | | | | |
| Publication bias: | |  | 0 | Unlikely | | | | | | | | |
| Effect size: | |  | 0 | No large magnitude of effect | | | | | | | | |
| Dose-response: | |  | +1 | Dose response relationship | | | | | | | | |
| Plausible confounding: | |  | 0 | No plausible confounding | | | | | | | | |
| Quality of evidence: | | | ⊕⊖⊖⊖ Very low | | | | | | | | | |
| Conclusion: | | | Increased risk for hyperinflation after increasing doses of radiotherapy exposing lung tissue in CAYA cancer survivors  (1 study; 49 participants; 49 participants exposed to radiation exposing the lung tissue) | | | | | | | | | |
| Comment: | | | One study only with small sample size and effect size without confidence interval. | | | | | | | | | |

| PICO | Study | | | | No. of participants | Follow-up (median/mean, range) yr | Radiotherapy exposing lung tissue | Pulmonary function Outcomes | Effect size | PFT quality | Risk of bias |
| --- | --- | --- | --- | --- | --- | --- | --- | --- | --- | --- | --- |
| 8a What is the risk for diffusion capacity impairment associated with different doses and volumes of radiotherapy?  - dose-volume relationship  - impact of dose per fraction | Weiner 2006 (18) | | | | 30 CSS (Wilms tumor, Hodgkin disease, Sarcoma,  Hepato-blastoma) | Median 2.79 (range 0-13.7) | 100%  (n=30) | - DLCO z-score (n=21)  - DLCO z-score (n=21) | No correlation between severity of abnormal FEV1, TCL, and DLCO (z-score) and total radiation dose:   - r^2^=0.13   (very weak)  No correlation between severity of abnormal DLCO (z-score) and total radiation dose after taking body length into account:   - r^2^=0.03   (very weak) | 1. No  2. Yes:  Wang X, Pediatr Pulmonol, 1993; Rosenthal M, Thorax 1993  3. No  4. Yes  5. No  6. No | Retrospective cohort  SB: High Risk  AB: High risk  DB: Low risk  CF: High Risk |
|  | Green 2016 (19) | | | | 606 CCS (FEV1, FVC)  597 CCS  (TLC, DLCO) | Median 21.9 | 76.7%  (n=465) | Proportion of CCS with pulmonary function parameter below %pred or LLN in the whole cohort  44.6% DLCO_corr_ <75% pred | Multivariable log-binomial regression:  Outcome: V10 (per 10% increase)  Relative Risk (95%CI, p-value)  1.07 (1.04–1.10, <0.001) | 1. No  2. Yes  Wanger J, Eur Respir J, 2005; Goldman HI, Am Rev Tuberc, 1959; Boren HG, Am J Med, 1966; Miller A, Am Rev Respir Dis, 1983; Quanjer PH, lookup table, accessed 2015; Quanjer PH, Eur Respir J, 2012  3. Yes  4. Yes: ATS  5. No  6. No | Prospective cohort  SB: high risk  AB: low risk  DB: unclear  CF: high risk |
|  | Armenian, 2015 (15) | | | | 121 CAYA | Median 17.1 (6.3-40.1) | 73.6%  (n=89)  Categories:  26.4% No (Ref.)  49.6% ≤20Gy  24.0% >20Gy | Total 42 diffusion abnormality  9% (3/32) no radiation,  40% (24/60) ≤20Gy,  52% (15/29) >20Gy | Multivariable logistic regression  Odds Ratio (95%CI)  1  6.4 (1.7-2.4)  11.3 (2.6-49.5) | 1. Yes  2. No  3. No  4. Yes: ATS  5. No  6. Yes | Prospective cohort  SB: Low risk  AB: low risk  DB: low risk  CF: high risk |
|  | Green 2015 (10) | | | | 260 embryonal brain tumors | Minimum 2 yr | 100% CSI  (n=260) | Proportion of CCS with DLCO below predicted 60 months after treatment  - DLCO corr < 75% predicted 25%  Unclear how many received proton and photon beam, but of initially 303 eligible patients only 20 had proton beam | Higher **DLCO% predicted** when treated with lower RT doses (≤2345 cGy) (p=0.032) | 1. No  2. Yes: 10 different references for standardization  3. No  4. Yes: ATS  5. No  6. No | Prospective cohort  SB: low risk  AB: high risk  DB: unclear  CF: high risk |
|  | De 2015 (12) | | | | 49  Osteo-sarcoma | Median 2.91 (range 0.01-8.28) | 100%  (n=49) | Proportion of CCS with abnormal results per lung function parameter  DLCO_adj_ <65% pred: 9% (4/49)  DLCO <65%pred: 14% (6/49) | Logistic regression analysis with radiation dose in Gy (cont.) and normal/ abnormal parameter  Odds Ratio (p-value)  Mean dose: 1.27; <0.01  Max dose: 1.07; <0.05  Mean dose : 1.16; <0.05  Max dose: 1.05; NS  Prescribed dose: 1.05; NS | 1. No  2. Yes  Hankinson JL, Am J Respir Crit Care Med, 1999;  Wang X, Pediatr Pulmonol, 1993  3. No  4. Yes: ATS  5. No  6. No | Retrospective cohort  SB: High Risk  AB: High Risk  DB: Low risk  CF: High Risk |
| GRADE assessment: | |  |  |  | | | | | | | |
| Study design: | |  | +4 | 2 retrospective cohort studies 3 prospective cohort studies | | | | | | | |
| Study limitations: | |  | -3 | Important limitations: Selection bias high in 3/5, low in 2/5; Attrition bias high in 3/5, low in 2/5; Detection bias low in 3/5, unclear in 2/5; Confounding high in 5/5 | | | | | | | |
| Consistency: | |  | 0 | No important inconsistency. Most studies show risk for more diffusion capacity impairment in CAYA cancer survivors treated with higher radiation doses. Only one study shows no correlation | | | | | | | |
| Directness: | |  | -1 | Results are direct, population and outcomes broadly generalizable, PFT quality unsure (1/5 does not mention reference values used; 5/5 mention the use of guidelines) | | | | | | | |
| Precision: | |  | -1 | Important imprecision; 2/5 performed multivariable analysis;21/5 shows effect size with small 95%CI; 1/5 shows effect size with large 95%CI; 1/5 shows effect size but without 95%CI, in 1/5 precision cannot be judged as result is shown with p-value only; 1/5 shows correlation coefficient only  2/6 have large confidence interval, 3/6 show no effect estimate but p-values 1/6 is a correlatin only | | | | | | | |
| Publication bias: | |  | 0 | Unlikely | | | | | | | |
| Effect size: | |  | 0 | No large magnitude of effect | | | | | | | |
| Dose-response: | |  | +1 | Dose-repsonse relationship | | | | | | | |
| Plausible confounding: | |  | 0 | No evidence of possible confounding | | | | | | | |
| Quality of evidence: | | | ⊕⊖⊖⊖ Very low | | | | | | | | |
| Conclusion: | | | Increased risk for diffusion capacity impairment after increasing doses of radiotherapy exposing the lung tissue in CAYA cancer survivors.  (5 studies; 4 significant effects, 1 non-significant effect; 1057 participants; 893 participants exposed to radiotherapy) | | | | | | | | |
| Comments: | | | Different cutoff values used between studies to define parameters as abnormal. One study shows correlation coefficient only | | | | | | | | |

##### 8b Different radiotherapeutic fields

| PICO | Study | No. of participants | Follow-up (median/mean, range) yr | Radiotherapy exposing lung tissue | Pulmonary function Outcomes | Effect size | Risk of bias |
| --- | --- | --- | --- | --- | --- | --- | --- |
| 8b What is the risk in different radiotherapeutic fields? 🡪 No study | | | | | | | |

##### 8c Age at exposure

| PICO | Study | | | | | No. of participants | | Follow-up (median/mean, range) yr | Radiotherapy exposing lung tissue | Pulmonary function Outcomes | Effect size | PFT quality | Risk of bias |
| --- | --- | --- | --- | --- | --- | --- | --- | --- | --- | --- | --- | --- | --- |
| 8c What is the risk for obstructive abnormalities associated with patient age at time of radiation? | Weiner 2006 (18) | | | | | 30 CSS (Wilms tumor, Hodgkin disease, Sarcoma,  Hepato-blastoma) | | Median 2.79 (range 0-13.7) | 100%  (n=30) | No correlation between severity of abnormal FEV1 (z-score) and age at time of radiation:  - FEV1 z-score (n=30) | Spearman Correlation:  No correlation with age at time of radiation  r^2^<0.001  (very weak) | 1. No  2. Yes:  Wang X, Pediatr Pulmonol, 1993; Rosenthal M, Thorax 1993  3. No  4. Yes  5. No  6. No | Retrospective cohort  SB: High Risk  AB: High risk  DB: Low risk  CF: High Risk |
|  | De 2015 (12) | | | | | 49  Osteo-sarcoma | | Median 2.91 (range 0.01-8.28) | 100%  (n=49) | Proportion of CCS with abnormal results per parameter  FEV1 <80% pred: 29% (14/49)  FEF25–5% <68% pred: 20% (10/49) | Univariable logistic regression analysis with age at radiation (cont.)  OR (p-value)  1.03 (NS)  1.09 (NS) | 1. No  2. Yes  Hankinson JL, Am J Respir Crit Care Med, 1999;  Wang X, Pediatr Pulmonol, 1993  3. No  4. Yes: ATS  5. No  6. No | Retrospective cohort  SB: High Risk  AB: High Risk  DB: Low risk  CF: High Risk |
|  | Khan 2020  (23) | | | | | 66 CCS exposed to radiotherapy | | Mean 9 years (range, 1-20) | 100%  (n=66) | <5 years  >5 or <13 years  >13 years: 1.0 (ref)  <5 years  >5 or <13 years  >13 years: 1.0 (ref)  <5 years  >5 or <13 years  >13 years: 1.0 (ref) | Multivariable logistic regression, crude model OR (95%CI)  3.20 (0.24-42.19) (NS)  1.68 (0.22-12.96) (NS)  Multivariable logistic regression, adjusted for time since treatment  OR (95%CI)  11.35 (0.20-634.6) (NS)  2.10 (0.26-16.98) (NS)  Multivariable logistic regression, adjusted for time since treatment and bleomycin exposure  OR (95%CI)  6.57 (0.08-571.7) (NS)  1.44 (0.11-19.21) (NS) | 1. No  2. Yes  Rosenthal M, Thorax, 1993  3. No  4. Yes: ATS  5. No  6. No | Retrospective cohort  SB: High Risk  AB: Low Risk  DB: Low risk  CF: Low Risk |
| GRADE assessment: | |  | |  | | |  | | | | | | |
| Study design: | | |  | +4 | 3 retrospective cohort studies | | | | | | | | |
| Study limitations: | | |  | -3 | Important limitations: Selection bias high in 3/3; Attrition bias high in 2/3; Detection bias low in 3/3; Confounding high in 2/3 | | | | | | | | |
| Consistency: | | |  | 0 | No inconsistency. Two studies show no correlation and non-significant association between obstructive abnormalities and age at radiotherapy. One study shows an increased risk with younger age at radiotherapy, but the associations were not significant and confidence intervals very large. | | | | | | | | |
| Directness: | | |  | 0 | Results broadly generalizable for CCS treated with radiotherapy exposing lung tissue. PFT quality is good (3/3 report reference values and guidelines used) | | | | | | | | |
| Precision: | | |  | -1 | Important imprecision; 1/3 reports correlation coefficient only, 1/3 studies shows effect size but without 95%CI and p-value not significant, 1/3 studies shows 95%CI which are very large and not significant | | | | | | | | |
| Publication bias: | | |  | 0 | Unlikely | | | | | | | | |
| Effect size: | | |  | 0 | No large magnitude of effect | | | | | | | | |
| Dose-response: | | |  | 0 | No age-respnce relationship | | | | | | | | |
| Plausible confounding: | | |  | 0 | No evidence of possible confounding | | | | | | | | |
| Quality of evidence: | | | | ⊕⊖⊖⊖ Very low | | | | | | | | | |
| Conclusion: | | | | No significant effect for obstructive abnormalities (FEV1, FEF25-75%) of older vs. younger age at radiotherapy exposing lung tissue in CAYA cancer survivors.  (3 studies; 145 participants; 145 participants exposed to radiotherapy exposing lung tissue) | | | | | | | | | |
| Comment | | | | Important imprecision | | | | | | | | | |

| PICO | Study | | | | No. of participants | | Follow-up (median/mean, range) yr | Radiotherapy exposing lung tissue | Pulmonary function Outcomes | Effect size | PFT quality | Risk of bias |
| --- | --- | --- | --- | --- | --- | --- | --- | --- | --- | --- | --- | --- |
| 8c What is the risk for restrictive abnormalities associated with patient age at time of radiation? | Weiner 2006 (18) | | | | 30 CSS (Wilms tumor, Hodgkin disease, Sarcoma,  Hepato-blastoma) | | Median 2.79 (range 0-13.7) | 100%  (n=30) | No correlation between severity of abnormal TCL (z-score) and age at time of radiation:  -TLC z-score (n=23) | Spearman Correlation:  No correlation with age at time of radiation  r^2^=0.08  (very weak) | 1. No  2. Yes:  Wang X, Pediatr Pulmonol, 1993; Rosenthal M, Thorax 1993  3. No  4. Yes  5. No  6. No | Retrospective cohort  SB: High Risk  AB: High risk  DB: Low risk  CF: High Risk |
|  | De 2015 (12) | | | | 49  Osteo-sarcoma | | Median 2.91 (range 0.01-8.28) | 100%  (n=49) | Proportion of CCS with abnormal results per parameter  FVC <80% pred: 24% (12/49)  TLC <77% pred: 15% (7/49) | Univariable logistic regression analysis with age at radiation (cont.)  OR (p-value)  1.13 (NS)  1.14 (NS) | 1. No  2. Yes  Hankinson JL, Am J Respir Crit Care Med, 1999;  Wang X, Pediatr Pulmonol, 1993  3. No  4. Yes: ATS  5. No  6. No | Retrospective cohort  SB: High Risk  AB: High Risk  DB: Low risk  CF: High Risk |
|  | Khan 2020  (23) | | | | 66 CCS exposed to radiotherapy | | Mean 9 years (range, 1-20) | 100%  (n=66) | <5 years  >5 or <13 years  >13 years: 1.0 (ref)  <5 years  >5 or <13 years  >13 years: 1.0 (ref)  <5 years  >5 or <13 years  >13 years: 1.0 (ref) | Multivariable logistic regression, crude model OR (95%CI)  3.75 (0.51-27.50) (NS)  2.34 (0.55-9.97) (NS)  Multivariable logistic regression, adjusted for time since treatment  OR (95%CI)  2.22 (0.15-33.44) (NS)  2.06 (0.45-9.51) (NS)  Multivariable logistic regression, adjusted for time since treatment and bleomycin exposure  OR (95%CI)  1.26 (0.06-25.63) (NS)  1.30 (0.19-8.72) (NS) | 1. No  2. Yes  Rosenthal M, Thorax, 1993  3. No  4. Yes: ATS  5. No  6. No | Retrospective cohort  SB: High Risk  AB: Low Risk  DB: Low risk  CF: Low Risk |
| GRADE assessment: | |  |  | | |  | | | | | | |
| Study design: | |  | +4 | 3 retrospective cohort studies | | | | | | | | |
| Study limitations: | |  | -3 | Important limitations: Selection bias high in 3/3; Attrition bias high in 2/3; Detection bias low in 3/3; Confounding high in 2/3 | | | | | | | | |
| Consistency: | |  | 0 | No inconsistency. Two studies show no correlation and non-significant association between restrictive abnormalities and age at radiotherapy. One study shows an increased risk with younger age at radiotherapy, but the associations were not significant and confidence intervals large. | | | | | | | | |
| Directness: | |  | 0 | Results broadly generalizable for CCS treated with radiotherapy exposing lung tissue. PFT quality is good (3/3 report reference values and guidelines used) | | | | | | | | |
| Precision: | |  | -1 | Important imprecision; 1/2 reports correlation coefficient only, 1/2 studies shows effect size but without 95%CI and p-value not significant, 1/3 studies shows 95%CI which are large and not significant | | | | | | | | |
| Publication bias: | |  | 0 | Unlikely | | | | | | | | |
| Effect size: | |  | 0 | No large magnitude of effect | | | | | | | | |
| Dose-response: | |  | 0 | No age-respnce relationship | | | | | | | | |
| Plausible confounding: | |  | 0 | No evidence of possible confounding | | | | | | | | |
| Quality of evidence: | | | ⊕⊖⊖⊖ Very low | | | | | | | | | |
| Conclusion: | | | No significant effect for restrictive abnormalities (TLC, FVC) of older vs. younger age at radiotherapy exposing lung tissue in CAYA cancer survivors.  (3 studies; 145 participants; 145 participants exposed to radiotherapy exposing lung tissue) | | | | | | | | | |
| Comment | | | Important imprecision | | | | | | | | | |

| PICO | Study | | | | No. of participants | | Follow-up (median/mean, range) yr | Radiotherapy exposing lung tissue | Pulmonary function Outcomes | Effect size | PFT quality | Risk of bias |
| --- | --- | --- | --- | --- | --- | --- | --- | --- | --- | --- | --- | --- |
| 8c What is the risk for hyperinflation associated with patient age at time of radiation? | De 2015 (12) | | | | 49  Osteo-sarcoma | | Median 2.91 (range 0.01-8.28) | 100%  (n=49) | Proportion of CCS with abnormal RV/TLC  RV/TLC >123% pred: 21% (12/49) | Univariable logistic regression analysis with age at radiation (cont.)  OR (p-value)  1.05 (NS) | 1. No  2. Yes  Hankinson JL, Am J Respir Crit Care Med, 1999;  Wang X, Pediatr Pulmonol, 1993  3. No  4. Yes: ATS  5. No  6. No | Retrospective cohort  SB: High Risk  AB: High Risk  DB: Low risk  CF: High Risk |
| GRADE assessment: | |  |  | | |  | | | | | | |
| Study design: | |  | +4 | 1 retrospective cohort study | | | | | | | | |
| Study limitations: | |  | -3 | Important limitations: Selection bias high in 1/1; Attrition bias high in 1/1; Detection bias low in 1/1; Confounding high in 1/1 | | | | | | | | |
| Consistency: | |  | 0 | One study only | | | | | | | | |
| Directness: | |  | 0 | Results and outcomes broadly generalizable. PFT quality is good (reference values and guidelines stated). | | | | | | | | |
| Precision: | |  | 0 | Imprecision 1/1 reports OR without 95%CI and p-value is not significant | | | | | | | | |
| Publication bias: | |  | 0 | Unlikely | | | | | | | | |
| Effect size: | |  | 0 | No large magnitude of effect | | | | | | | | |
| Dose-response: | |  | 0 | No clear relation with increase in the outcome with older age at time of radiotherapy | | | | | | | | |
| Plausible confounding: | |  | 0 | No evidence of possible confounding | | | | | | | | |
| Quality of evidence: | | | ⊕⊖⊖⊖ Very low | | | | | | | | | |
| Conclusion: | | | No significant effect on hyperinflation of older vs. younger age at radiotherapy exposing lung tissue in CAYA cancer survivors.  (1 study; 49 participants, 49 participants exposed to radiotherapy exposing the lung tissue) | | | | | | | | | |
| Comment | | | Small sample size and important imprecision | | | | | | | | | |

| PICO | Study | | | | | No. of participants | | Follow-up (median/mean, range) yr | Radiotherapy exposing lung tissue | Pulmonary function Outcomes | Effect size | PFT quality | Risk of bias |
| --- | --- | --- | --- | --- | --- | --- | --- | --- | --- | --- | --- | --- | --- |
| 8c What is the risk for diffusion capacity impairment associated with patient age at time of radiation? | Weiner 2006 (18) | | | | | 30 CSS (Wilms tumor, Hodgkin disease, Sarcoma,  Hepato-blastoma) | | Median 2.79 (range 0-13.7) | 100%  (n=30) | No correlation between severity of abnormal DLCO (z-score) and age at time of radiation:  - DLCO z-score (n=21) | Spearman Correlation:  No correlation with age at time of radiation  r^2^=0.08  (very weak) | 1. No  2. Yes:  Wang X, Pediatr Pulmonol, 1993; Rosenthal M, Thorax 1993  3. No  4. Yes  5. No  6. No | Retrospective cohort  SB: High Risk  AB: High risk  DB: Low risk  CF: High Risk |
|  | De 2015 (12) | | | | | 49  Osteo-sarcoma | | Median 2.91 (range 0.01-8.28) | 100%  (n=49) | Proportion of CCS with abnormal results per parameter  DLCO adj <65% pred: 9% (4/49) | Univariable logistic regression analysis with age at radiation (cont.)  OR (p-value)  1.01 (NS) | 1. No  2. Yes  Hankinson JL, Am J Respir Crit Care Med, 1999;  Wang X, Pediatr Pulmonol, 1993  3. No  4. Yes: ATS  5. No  6. No | Retrospective cohort  SB: High Risk  AB: High Risk  DB: Low risk  CF: High Risk |
|  | Khan 2020  (23) | | | | | 66 CCS exposed to radiotherapy | | Mean 9 years (range, 1-20) | 100%  (n=66) | <5 years  >5 or <13 years  >13 years: 1.0 (ref)  <5 years  >5 or <13 years  >13 years: 1.0 (ref)  <5 years  >5 or <13 years  >13 years: 1.0 (ref) | Multivariable logistic regression, crude model OR (95%CI)  3.75 (0.51-27.5)  3.00 (0.73-12.27)  Multivariable logistic regression, adjusted for time since treatment  OR (95%CI)  4.27 (0.28-64.08)  3.09(0.71-13.45)  Multivariable logistic regression, adjusted for time since treatment and bleomycin exposure  OR (95%CI)  3.64 (0.18-72.86)  2.74 (0.46-16.18) | 1. No  2. Yes  Rosenthal M, Thorax, 1993  3. No  4. Yes: ATS  5. No  6. No | Retrospective cohort  SB: High Risk  AB: Low Risk  DB: Low risk  CF: Low Risk |
| GRADE assessment: | |  | |  | | |  | | | | | | |
| Study design: | | |  | +4 | 3 retrospective cohort studies | | | | | | | | |
| Study limitations: | | |  | -3 | Important limitations: Selection bias high in 3/3; Attrition bias high in 2/3; Detection bias low in 3/3; Confounding high in 2/3 | | | | | | | | |
| Consistency: | | |  | 0 | No inconsistency. Two studies show no correlation and non-significant association between diffusion capacity impairment and age at radiotherapy. One study shows an increased risk with younger age at radiotherapy, but the associations were not significant and confidence intervals very large. | | | | | | | | |
| Directness: | | |  | 0 | Results broadly generalizable for CCS treated with radiotherapy exposing lung tissue. PFT quality is good (2/2 report reference values and guidelines used) | | | | | | | | |
| Precision: | | |  | -1 | Important imprecision; 1/3 reports correlation coefficient only, 1/3 studies shows effect size but without 95%CI and p-value not significant, 1/3 studies shows 95%CI which are very large and not significant | | | | | | | | |
| Publication bias: | | |  | 0 | Unlikely | | | | | | | | |
| Effect size: | | |  | 0 | No large magnitude of effect | | | | | | | | |
| Dose-response: | | |  | 0 | No clear relation with increase in the outcome with older age at time of radiotherapy | | | | | | | | |
| Plausible confounding: | | |  | 0 | No evidence of possible confounding | | | | | | | | |
| Quality of evidence: | | | | ⊕⊖⊖⊖ Very low | | | | | | | | | |
| Conclusion: | | | | No significant effect on diffusion capacity impairment (DLCO) of older vs. younger age at radiotherapy exposing lung tissue in CAYA cancer survivors.  (3 studies; 145 participants; 145 participants exposed to radiotherapy to the thorax) | | | | | | | | | |
| Comment | | | | Important imprecision | | | | | | | | | |

##### 8d Radiosensitizer

| PICO | Study | No. of participants | Follow-up (median/mean, range) yr | Radiotherapy exposing lung tissue | Pulmonary function Outcomes | Effect size | Risk of bias |
| --- | --- | --- | --- | --- | --- | --- | --- |
| 8d What is the risk of pulmonary dysfunction in CAYA treated with radiosensitizing/radiomimetic chemotherapy (doxorubicin, dactinomycin, busulfan, bleomycin, topotecan, irinotecan) combined with radiotherapy involving lung tissue compared to CAYA not treated with radiomimetic chemotherapy combined with radiotherapy involving lung tissue? 🡪 No study | | | | | | | |

#### PICO 9: Thoracic surgery

| PICO | Study | | | | No. of participants | | Follow-up (median/mean, range) yr | Thoracic Surgery | Pulmonary function Outcomes | Effect size | PFT quality | Risk of bias |
| --- | --- | --- | --- | --- | --- | --- | --- | --- | --- | --- | --- | --- |
| 9 What is the risk of obstructive abnormalities in CAYA treated with thoracic surgery compared to CAYA not treated with thoracic surgery? | Record 2016 (1) | | | | 143 CCS | | Mean 14.1 ± 4.8 (SD) | 16.8%  (n=24) | Obstructive  (FVC, FEV1, FEV1/FVC <80% or FEF25–75% <68% predicted)  50.0% (12/24) surgery  21.0% (25/119) no surgery | Univariable comparison Chi2 surgery Yes/No  0.06 | 1. No  2. Yes:  Wang X, Pediatr Pulmonol 2005; Hankinson JL, Am J Respir Crit Care Med 1999  3. No  4. Yes  5. No  6. Yes | Retrospective cohort  SB: High risk  AB: Low risk  DB: Low risk  CF: High risk |
|  | De 2015 (12) | | | | 49  Osteo-sarcoma | | Median 2.91 (range 0.01-8.28) | 18%  (n=9) | Proportion of CCS with abnormal results per parameter  FEV1 <80% pred: 29% (12/49)  FEF25–75% <68% pred: 20% (10/49)  Obstructive  (FEV1/FVC <80%, FEV1<80% or FEF25-75<68% with normal TLC): 24% (12/49) | Logistic regression analysis with surgery yes/no  Odds Ratio (p-value)  8.0 (<0.01)  2.35 (NS)  5.89 (<0.05) | 1. No  2. Yes  Hankinson JL, Am J Respir Crit Care Med, 1999;  Wang X, Pediatr Pulmonol, 1993  3. No  4. Yes: ATS  5. No  6. No | Retrospective cohort  SB: High Risk  AB: High Risk  DB: Low risk  CF: High Risk |
|  | Denbo, 2014 (13) | | | | 21  Osteo-sarcoma | | Mean 20 yr  (SD ±9) | N=15 with 1 Thoracotomy  N=6 with ≥2 Thoracotomy | Proportion of CCS with abnormal results per parameter  FEV1 <80% pred  1 Thoracotomy (6/15) vs ≥2 Thoracotomies (4/6) | Fishers exact test  p-value  0.362 | 1. No  2. Yes  Hankinson JL, Am J Respir Crit Care Med, 1999; Miller A, Am Rev Respir Dis, 1983  3. No  4. Yes: ATS, Morris AH, 1984  5. No  6. No | Prospective cohort  SB: Low risk  AB: Low risk  DB: Unclear  CF: High risk |
|  | Stone 2020  (19) | | | | 62 high-risk neuroblastoma | | Median 5.29 (0.24-15.24) | 23%  (n=14) | **FEV1**  (FEV1 <80% pred)  Surgery yes: 85.7% abnormal  Surgery no: 35.4% abnormal | OR, 95%CI  10.94 (2.19 – 54.71), p=0.001 | 1. No  2. Yes  3. No  4. Yes (ATS)  5. No  6. No | Prospective cohort  SB: high risk  AB: low risk  DB: low risk  CF: high risk |
| GRADE assessment: | |  |  | | |  | | | | | | |
| Study design: | |  | +4 | 2 retrospective cohort studies, 2 prospective cohort studies | | | | | | | | |
| Study limitations: | |  | -2 | Some limitations: Selection bias high in 3/4, low in 1/4; Attrition bias high in 1/4, low in 3/4; Detection bias low in 3/4, unclear in 1/4; Confounding high in 4/4 | | | | | | | | |
| Consistency: | |  | 0 | No important inconsistency. Most studies show generally worse pulmonary outcomes after thoracic surgery, 2 studies significant results but one with large 95%CI | | | | | | | | |
| Directness: | |  | 0 | Population and outcomes are generalizable. PFT quality is good (4/4 state references and guidelines used). | | | | | | | | |
| Precision: | |  | -1 | Important imprecision; in 2/4 studies precision cannot be judged as results are shown as p-values only, 1/4 studies shows results as Odds Ration but without 95%CI, 1/4 studies with Odds Ratio but large 95%CI | | | | | | | | |
| Publication bias: | |  | 0 | Unlikely | | | | | | | | |
| Effect size: | |  | 0 | No large magnitude of effect | | | | | | | | |
| Dose-response: | |  | 0 | One study shows non-significantly higher proportion of obstructive abnormalities after ≥2 thoracotomies compared to one, but small sample size. | | | | | | | | |
| Plausible confounding: | |  | 0 | No evidence of possible confounding | | | | | | | | |
| Quality of evidence: | | | ⊕⊖⊖⊖ Very low | | | | | | | | | |
| Conclusion: | | | Increased risk for obstructive abnormalities (FEV1, FEV1/FVC, FEF25-75, “obstructive”) after thoracic surgery vs. no surgery in CAYA cancer survivors. But 3/4 with selected survivor cohorts (osteosarcoma, neuroblastoma)  (4 studies; 2 studies significant, 2 studies non-significant; 275 participants; 68 exposed to thoracic surgery to the lung or thorax) | | | | | | | | | |
| Comment | | | Only small sample size exposed to thoracic surgery and effect size either without confidence interval or not assessable as results shown as p-value only. | | | | | | | | | |

| PICO | Study | | | No. of participants | | Follow-up (median/mean, range) yr | Thoracic Surgery | Pulmonary function Outcomes | Effect size | PFT quality | Risk of bias |
| --- | --- | --- | --- | --- | --- | --- | --- | --- | --- | --- | --- |
| 9 What is the risk of restrictive abnormalities in CAYA treated with thoracic surgery compared to CAYA not treated with thoracic surgery? | Record 2016 (1) | | | 143 CCS | | Mean 14.1 ± 4.8 (SD) | 16.8%  (n=24) | Restrictive (TLC<80% predicted)  8.3% (2/24) surgery  14.3% (17/119) no surgery | Univariable comparison Chi2 surgery Yes/No  0.01 | 1. No  2. Yes:  Wang X, Pediatr Pulmonol 2005; Hankinson JL, Am J Respir Crit Care Med 1999  3. No  4. Yes  5. No  6. Yes | Retrospective cohort  SB: high risk  AB: low risk  DB: low risk  CF: high risk |
|  | De 2015 (12) | | | 49  Osteo-sarcoma | | Median 2.91 (range 0.01-8.28) | 18%  (n=9) | Proportion of CCS with abnormal results per parameter  FVC <80% pred: 24% (12/49)  TLC <77% pred: 15% 7/49)  Restrictive (TLC <77%): 15% (7/49) | Logistic regression with surgery yes/no  Odds Ratio  (p-value)  3.2 (NS)  1.94 (NS)  1.94 (NS) | 1. No  2. Yes  Hankinson JL, Am J Respir Crit Care Med, 1999;  Wang X, Pediatr Pulmonol, 1993  3. No  4. Yes: ATS  5. No  6. No | Retrospective cohort  SB: High Risk  AB: High Risk  DB: Low risk  CF: High Risk |
|  | Denbo, 2014 (13) | | | 21  Osteo-sarcoma | | Mean 20 yr  (SD ±9) | N=15 with 1 thoracotomy  N=6 with ≥2 thoracotomies | Proportion of CCS with abnormal results per parameter  FVC <80% pred  1 Thoracotomy (5/15) vs  ≥2 Thoracotomies (3/5)  TLC <75% pred  1 Thoracotomy (2/15) vs  ≥2 Thoracotomies (4/6) | Fishers exact test  p-value  0.347  0.031 | 1. No  2. Yes  Hankinson JL, Am J Respir Crit Care Med, 1999; Miller A, Am Rev Respir Dis, 1983  3. No  4. Yes: ATS, Morris AH, 1984  5. No  6. No | Prospective cohort  SB: low risk  AB: low risk  DB: unclear  CF: high risk |
|  | Mulder 2011 (9) | | | 193 CCS | | Median 17.9 (5.6-36.8) | 16.6%  (n=32) | 34/193 Restrictive  (TLC OR FVC <75%)  Of those Exposed: 7.7% | Odds Ratio (95%CI) for surgery yes/no  3.79 (1.25-11.50) | 1. No  2. No  3. No  4. No  5. No  6. No | Retrospective cohort  SB: low risk  AB: low risk  DB: unclear  CF: low risk |
|  | Stone 2020  (19) | | | 62 high-risk neuroblastoma | | Median 5.29 (0.24-15.24) | 23%  (n=14) | **FVC**  (FVC <80% pred)  Surgery yes: 92.9% abnormal  Surgery no: 41.5% abnormal  **TLC**  (TLC <80% pred)  Surgery yes: 64.3% abnormal  Surgery no: 35.4% abnormal | OR, 95%CI  18.20 (2.20 – 150.58), p=0.001  3.28 (0.95 – 11.38), p=0.054 | 1. No  2. Yes  3. No  4. Yes (ATS)  5. No  6. No | Prospective cohort  SB: high risk  AB: low risk  DB: low risk  CF: high risk |
| GRADE assessment: | |  |  | |  | | | | | | |
| Study design: | |  | +4 | 3 retrospective cohort studies, 2 prospective cohort study | | | | | | | |
| Study limitations: | |  | -2 | Some limitations: Selection bias high in 3/5, low in 2/5; Attrition bias high in 1/5, low in 4/5; Detection bias low in 3/5, unclear in 2/5; Confounding high in 4/5, low in 1/5 | | | | | | | |
| Consistency: | |  | 0 | Most studies show similar results; in one study CAYA cancer survivors without thoracic surgery have more restrictive abnormalities, in remaining 3 studies exposed are more at risk. | | | | | | | |
| Directness: | |  | -1 | Results are generalizable. PFT quality is unsure (4/5 state references and guidelines used). | | | | | | | |
| Precision: | |  | -1 | Important imprecision; 1/5 shows effect size with OR and 95%CI, 1/5 shows OR but no CI, 1/5 studies with Odds Ratio but large 95%CI, in 2/5 precision cannot be judged as p-value is shown only | | | | | | | |
| Publication bias: | |  | 0 | Unlikely | | | | | | | |
| Effect size: | |  | 0 | No large magnitude of effect | | | | | | | |
| Dose-response: | |  | 0 | One studies shows higher proportion of obstructive abnormalities after ≥2 thoracotomies compared to one, but very small sample size. | | | | | | | |
| Plausible confounding: | |  | 0 | No evidence of possible confounding | | | | | | | |
| Quality of evidence: | | | ⊕⊖⊖⊖ Very low | | | | | | | | |
| Conclusion: | | | Increased risk for restrictive abnormalities (FVC, TLC, restrictive) in CAYA cancer survivors after thoracic surgery vs. CAYA cancer survivors without thoracic surgery. But 3/5 with selected survivor cohorts (osteosarcoma, neuroblastoma)  (5 studies; 3 significant effect, 2 non-significant effect; 468 participants; 100 participants exposed to thoracic surgery) | | | | | | | | |
| Comment | | | Two studies show effect size, three studies show odds ration or p-value only. One study shows a contradictory result. Different definitions and cutoff values used. | | | | | | | | |

| PICO | Study | | | No. of participants | | Follow-up (median/mean, range) yr | Thoracic Surgery | Pulmonary function Outcomes | Effect size | PFT quality | Risk of bias |
| --- | --- | --- | --- | --- | --- | --- | --- | --- | --- | --- | --- |
| 9 What is the risk of hyperinflation in CAYA treated with thoracic surgery compared to CAYA not treated with thoracic surgery? | Record 2016 (1) | | | 143 CCS | | Mean 14.1 ± 4.8 (SD) | 16.8%  (n=24) | Hyperinflation  (RV >120%predicted or RV/TLC >28% predicted)  58.3% (14/24) surgery  37.8% no (45/119) surgery | Univariable comparison Chi2 surgery Yes/No  0.41 | 1. No  2. Yes:  Wang X, Pediatr Pulmonol 2005; Hankinson JL, Am J Respir Crit Care Med 1999  3. No  4. Yes  5. No  6. Yes | Retrospective cohort  SB: high risk  AB: low risk  DB: low risk  CF: high risk |
|  | De 2015 (12) | | | 49  Osteo-sarcoma | | Median 2.91 (range 0.01-8.28) | 18%  (n=9) | Proportion of CCS with abnormal RV/TLC  RV/TLC ratio >28%: 21% (10/49) | Logistic regression with surgery yes/no  Odds Ratio (p-value)  8.5 (<0.01) | 1. No  2. Yes  Hankinson JL, Am J Respir Crit Care Med, 1999;  Wang X, Pediatr Pulmonol, 1993  3. No  4. Yes: ATS  5. No  6. No | Retrospective cohort  SB: High Risk  AB: High Risk  DB: Low risk  CF: High Risk |
| GRADE assessment: | |  |  | |  | | | | | | |
| Study design: | |  | +4 | 2 retrospective cohort studies | | | | | | | |
| Study limitations: | |  | -3 | Important limitations: Selection bias high in 2/2; Attrition bias high in 1/2, low in 1/2; Detection bias low in 2/2; Confounding high in 2/2 | | | | | | | |
| Consistency: | |  | 0 | Studies show generally more hyperinflation in exposed CAYA cancer survivors, some are significant some not | | | | | | | |
| Directness: | |  | 0 | Results are generalizable. PFT quality is good (2/2 state references and guidelines used). | | | | | | | |
| Precision: | |  | -1 | Important imprecision; 1/2 reports odds ration without confidence interval, in 1/2 precision cannot be judged as the result is shown as p-value only | | | | | | | |
| Publication bias: | |  | 0 | Unlikely | | | | | | | |
| Effect size: | |  | 0 | No large magnitude of effect | | | | | | | |
| Dose-response: | |  | 0 | Not applicable | | | | | | | |
| Plausible confounding: | |  | 0 | No evidence of possible confounding | | | | | | | |
| Quality of evidence: | | | ⊕⊖⊖⊖ Very low | | | | | | | | |
| Conclusion: | | | Inconsistent findings for hyperinflation (RV/TLC, RV) in CAYA cancer survivors after thoracic surgery vs. CAYA cancer survivors without thoracic surgery.  (2 studies; 1 significant effect, 1 non-significant effect; 192 participants; 33 participants exposed to thoracic surgery) | | | | | | | | |
| Comment | | | Significant finding in one of both studies only and very small sample size. | | | | | | | | |

| PICO | Study | | | No. of participants | | Follow-up (median/mean, range) yr | Thoracic Surgery | Pulmonary function Outcomes | Effect size | PFT quality | Risk of bias |
| --- | --- | --- | --- | --- | --- | --- | --- | --- | --- | --- | --- |
| 9 What is the risk of diffusion capacity impairment in CAYA treated with thoracic surgery compared to CAYA not treated with thoracic surgery? | De 2015 (12) | | | 49  Osteo-sarcoma | | Median 2.91 (range 0.01-8.28) | 18%  (n=9) | Proportion of CCS with abnormal DLCO  DLCO_adj_ <65% pred: 9% (4/49)  Diffusion defect  (DLCO <65%): 14% (6/49) | Logistic regression with surgery yes/no  Odds Ratio (p-value)  1.89 (NS)  1.07 (NS) | 1. No  2. Yes  Hankinson JL, Am J Respir Crit Care Med, 1999;  Wang X, Pediatr Pulmonol, 1993  3. No  4. Yes: ATS  5. No  6. No | Retrospective cohort  SB: High Risk  AB: High Risk  DB: Low risk  CF: High Risk |
|  | Denbo, 2014 (13) | | | 21  Osteo-sarcoma | | Mean 20 yr  (SD ±9) | N=15 with 1 thoracotomy  N=6 with ≥2 thoracotomies | Proportion of CCS with abnormal DLCO  DLCO_corr_ <75% pred  1 Thoracotomy (7/15) vs ≥2 Thoracotomies (2/4) | Fishers exact test  p-value  1.00 | 1. No  2. Yes  Hankinson JL, Am J Respir Crit Care Med, 1999; Miller A, Am Rev Respir Dis, 1983  3. No  4. Yes: ATS, Morris AH, 1984  5. No  6. No | Prospective cohort  SB: low risk  AB: low risk  DB: unclear  CF: high risk |
|  | Mulder 2011 (9) | | | 193 CCS | | Median 17.9 (5.6-36.8) | 16.6%  (n=32) | 85 Diffusion capacity impairment (DLCO <75%)  Of those Exposed: 46.2% | Odds Ratio (95%CI) for surgery yes/no  1.98 (0.68-5.75) | 1. No  2. No  3. No  4. No  5. No  6. No | Retrospective cohort  SB: low risk  AB: low risk  DB: unclear  CF: low risk |
|  | Stone 2020  (19) | | | 62 high-risk neuroblastoma | | Median 5.29 (0.24-15.24) | 23%  (n=14) | **DLCO**  (DLCO <80% pred)  Surgery yes: 83.3% abnormal  Surgery no: 68.2% abnormal | OR, 95%CI  2.33 (0.45 – 12.09), p=0.475 | 1. No  2. Yes  3. No  4. Yes (ATS)  5. No  6. No | Prospective cohort  SB: high risk  AB: low risk  DB: low risk  CF: high risk |
| GRADE assessment: | |  |  | |  | | | | | | |
| Study design: | |  | +4 | 2 retrospective cohort studies, 2 prospective cohort study | | | | | | | |
| Study limitations: | |  | -2 | Some limitations: Selection bias high in 2/4, low in 2/4; Attrition bias high in 1/4, low in 3/4; Detection bias low in 2/4, unclear in 2/4; Confounding high in 3/4, low in 1/4 | | | | | | | |
| Consistency: | |  | 0 | Most studies show similar results | | | | | | | |
| Directness: | |  | -1 | Results are generalizable. PFT quality is unsure (3/4 state references and guidelines used). | | | | | | | |
| Precision: | |  | -1 | Important imprecision: 2/4 studies show effect size with CI, 1/4 shows effect size (OR) without CI, and in 1/4 precision cannot be judged as result is shown as p-value only; 3/4 show univariable analysis only | | | | | | | |
| Publication bias: | |  | 0 | Unlikely | | | | | | | |
| Effect size: | |  | 0 | No large magnitude of effect | | | | | | | |
| Dose-response: | |  | 0 | Not applicable | | | | | | | |
| Plausible confounding: | |  | 0 | No evidence of possible confounding | | | | | | | |
| Quality of evidence: | | | ⊕⊖⊖⊖ Very low | | | | | | | | |
| Conclusion: | | | No significant effect on diffusion capacity impairment (DLCO) after thoracic surgery vs. no surgery in CAYA cancer survivors  (4 studies; 325 participants; 76 participants exposed to thoracic surgery) | | | | | | | | |
| Comment | | | Small sample size exposed to thoracic surgery, definition for cutoff values differ between studies, precision unlear. | | | | | | | | |

##### 9a Different resection volumes

| PICO | Study | No. of participants | Follow-up (median/mean, range) yr | Thoracic Surgery | Pulmonary function Outcomes | Effect size | Risk of bias |
| --- | --- | --- | --- | --- | --- | --- | --- |
| 9a What is the risk associated with different resection volumes? | | | | | | | |

**No study**

##### 9b Age at exposure

| PICO | Study | No. of participants | Follow-up (median/mean, range) yr | Thoracic Surgery | Pulmonary function Outcomes | Effect size | Risk of bias |
| --- | --- | --- | --- | --- | --- | --- | --- |
| 9b What is the risk in younger compared to older age at thoracic surgery? | | | | | | | |

**No study**

#### PICO 10: Combinations

##### 10a Thoracic surgery plus chemotherapy

| PICO | Study | No. of participants | Follow-up (median/mean, range) yr | Thoracic Surgery and chemotherapy | Pulmonary function Outcomes | Effect size | PFT quality | Risk of bias |
| --- | --- | --- | --- | --- | --- | --- | --- | --- |
| 10a What is the risk of obstructive abnormalities after thoracic surgery combined with pulmotoxic chemotherapy (bleomycin, CCNU, BCNU, Busulfan, Cyclophosphamide, Methotrexate, Gemcitabine)? | | | | | | | | |

**No study**

| PICO | Study | | | No. of participants | | Follow-up (median/mean, range) yr | Thoracic surgery and chemotherapy | Pulmonary function Outcomes | Effect size | PFT quality | Risk of bias |
| --- | --- | --- | --- | --- | --- | --- | --- | --- | --- | --- | --- |
| 10a What is the risk of restrictive abnormalities after thoracic surgery combined with pulmotoxic chemotherapy (bleomycin, CCNU, BCNU, Busulfan, Cyclophosphamide, Methotrexate, Gemcitabine)? | Mulder 2011 (9) | | | 193 CCS | | Median 17.9 (5.6-36.8) | Bleomycin plus surgery 1.6%  (n=3)  Bleomycin only 50.8%  (n=98) | Restrictive impairment (TLC OR FVC <75%)  Bleomycin only vs Bleomycin + surgery | Odds Ratio (95%CI)  Bleomycin plus surgery vs. Bleomycin only  Not estimable as no cases in Bleomycin + surgery | 1. No  2. No  3. No  4. No  5. No  6. No | Retrospective cohort  SB: low risk  AB: low risk  DB: unclear  CF: low risk |
| GRADE assessment: | |  |  | |  | | | | | | |
| Study design: | |  | +4 | 1 retrospective cohort study | | | | | | | |
| Study limitations: | |  | -1 | Some limitations: Selection bias low in 1/1; Attrition bias low in 1/1; Detection bias unclear in 1/1; Confounding, low in 1/1 | | | | | | | |
| Consistency: | |  | 0 | One study only | | | | | | | |
| Directness: | |  | -1 | Results are broadly generalizable. PFT quality is unsure (no reference values and guidelines mentioned). | | | | | | | |
| Precision: | |  | 0 | OR not estimable, one study only | | | | | | | |
| Publication bias: | |  | 0 | Unlikely | | | | | | | |
| Effect size: | |  | 0 | No large magnitude of effect | | | | | | | |
| Dose-response: | |  | 0 | Not applicable | | | | | | | |
| Plausible confounding: | |  | 0 | No evidence of possible confounding | | | | | | | |
| Quality of evidence: | | | ⊕⊖⊖⊖ Very low | | | | | | | | |
| Conclusion: | | | No statement possible on restrictive abnormalities (TLC, FVC) after thoracic surgery combined with bleomycin vs. bleomycin alone in CAYA cancer survivors.  (1 study; 193 participants; 3 participants exposed to thoracic surgery and bleomycin; 98 participants exposed to bleomycin only) | | | | | | | | |
| Comment | | | Very small sample size exposed to thoracic surgery and bleomycin, no effect size, PFT quality is unsure. | | | | | | | | |

| PICO | Study | No. of participants | Follow-up (median/mean, range) yr | Thoracic surgery and chemotherapy | Pulmonary function Outcomes | Effect size | PFT quality | Risk of bias |
| --- | --- | --- | --- | --- | --- | --- | --- | --- |
| 10a What is the risk of hyperinflation after thoracic surgery combined with pulmotoxic chemotherapy (bleomycin, CCNU, BCNU, Busulfan, Cyclophosphamide, Methotrexate, Gemcitabine)? | | | | | | | | |

**No study**

| PICO | Study | | | | No. of participants | Follow-up (median/mean, range) yr | Thoracic surgery and chemotherapy | Pulmonary function Outcomes | Effect size | PFT quality | Risk of bias |
| --- | --- | --- | --- | --- | --- | --- | --- | --- | --- | --- | --- |
| 10a What is the risk of diffusion capacity impairment after thoracic surgery combined with pulmotoxic chemotherapy (bleomycin, CCNU, BCNU, Busulfan, Cyclophosphamide, Methotrexate, Gemcitabine)? | Mulder 2011 (9) | | | | 193 CCS | Median 17.9 (5.6-36.8) | Bleomycin plus surgery 1.6%  (n=3)  Bleomycin only 50.8%  (n=98) | Diffusion capacity impairment (DLCO <75%)  Bleomycin only vs Bleomycin + surgery | Odds Ratio (95%CI)  Bleomycin plus surgery vs. Bleomycin only  1.38 (0.10-18.66) | 1. No  2. No  3. No  4. No  5. No  6. No | Retrospective cohort  SB: low risk  AB: low risk  DB: unclear  CF: low risk |
| GRADE assessment: | |  |  |  | | | | | | | |
| Study design: | |  | +4 | 1 retrospective cohort study | | | | | | | |
| Study limitations: | |  | -1 | Some limitations: Selection bias low in 1/1; Attrition bias low in 1/1; Detection bias unclear in 1/1; Confounding, low in 1/1 | | | | | | | |
| Consistency: | |  | 0 | One study only | | | | | | | |
| Directness: | |  | -1 | Results are broadly generalizable. PFT quality is unsure (no reference values and guidelines mentioned). | | | | | | | |
| Precision: | |  | -2 | Results shown with effect size but large confidence interval, one study only | | | | | | | |
| Publication bias: | |  | 0 | Unlikely | | | | | | | |
| Effect size: | |  | 0 | No large magnitude of effect | | | | | | | |
| Dose-response: | |  | 0 | Not applicable | | | | | | | |
| Plausible confounding: | |  | 0 | No evidence of possible confounding | | | | | | | |
| Quality of evidence: | | | ⊕⊖⊖⊖ Very low | | | | | | | | |
| Conclusion: | | | No significant effect on diffusion capacity impairment (DLCO) after thoracic surgery combined with bleomycin vs. bleomycin alone in CAYA cancer survivors in one study.  (1 study; 193 participants; 3 participants exposed to thoracic surgery and bleomycin; 98 participants exposed to bleomycin only) | | | | | | | | |
| Comment | | | Very small sample size exposed to thoracic surgery and bleomycin, large confidence interval, PFT quality is unsure. | | | | | | | | |

##### 10b Thoracic surgery plus radiotherapy

| PICO | Study | | | No. of participants | | Follow-up (median/mean, range) yr | Thoracic surgery and radiotherapy | Pulmonary function Outcomes | Effect size | PFT quality | Risk of bias |
| --- | --- | --- | --- | --- | --- | --- | --- | --- | --- | --- | --- |
| 10b What is the risk of obstructive abnormalities after thoracic surgery combined with radiotherapy exposing lung tissue? | Stone 2020  (19) | | | 62 high-risk neuroblastoma | | Median 5.29 (0.24-15.24) | 18%  (n=12) | **FEV1**  (FEV1 <80% pred)  RT + Surgery yes: 91.7% abnormal  RT + Surgery no: 36% abnormal | OR, 95%CI  19.56 (2.33 – 164.05), p=0.001 | 1. No  2. Yes  3. No  4. Yes (ATS)  5. No  6. No | Prospective cohort  SB: high risk  AB: low risk  DB: low risk  CF: high risk |
| *GRADE assessment:* | |  |  | |  | | | | | | |
| Study design: | |  | +4 | 1 prospective cohort study | | | | | | | |
| Study limitations: | |  | -1 | Some limitations: Selection bias high in 1/1; Attrition bias low in 1/1; Detection bias low in 1/1; Confounding high in 1/1 | | | | | | | |
| Consistency: | |  | 0 | One study only | | | | | | | |
| Directness: | |  | 0 | Results are broadly generalizable. PFT quality is good (reference values and guidelines mentioned). | | | | | | | |
| Precision: | |  | -2 | Results shown with effect size but very large confidence interval, one study only | | | | | | | |
| Publication bias: | |  | 0 | Unlikely | | | | | | | |
| Effect size: | |  | +1 | Large magnitude of effect | | | | | | | |
| Dose-response: | |  | 0 | Not applicable | | | | | | | |
| Plausible confounding: | |  | 0 | No evidence of possible confounding | | | | | | | |
| Quality of evidence: | | | ⊕⊖⊖⊖ VERY LOW | | | | | | | | |
| Conclusion: | | | Increased risk for obstructive abnormalities (FEV1) after thoracic surgery combined with radiotherapy exposing lung tissue vs. no exposure in CAYA cancer survivors.  (1 study; 62 participants; 12 participants exposed to radiotherapy exposing lung tissue plus thoracic surgery; 51 participants not exposed) | | | | | | | | |
| Comment | | | One study, very small sample size exposed to thoracic surgery and radiotherapy exposing lung tissue, very large confidence interval. | | | | | | | | |

| PICO | Study | | | No. of participants | | Follow-up (median/mean, range) yr | Thoracic surgery and radiotherapy | Pulmonary function Outcomes | Effect size | PFT quality | Risk of bias |
| --- | --- | --- | --- | --- | --- | --- | --- | --- | --- | --- | --- |
| 10b What is the risk of restrictive abnormalities after thoracic surgery combined with radiotherapy exposing lung tissue? | Mulder 2011 (9) | | | 193 CCS | | Median 17.9 (5.6-36.8) | Radiotherapy plus surgery 8.3%  (n=16)  Bleomycin only 50.8%  (n=98) | Restrictive impairment (TLC OR FVC <75%)  Bleomycin only vs radiotherapy + surgery | Odds Ratio (95%CI)  Radiotherapy plus surgery vs. Bleomycin only  33.44 (7.81-143.09) | 1. No  2. No  3. No  4. No  5. No  6. No | Retrospective cohort  SB: low risk  AB: low risk  DB: unclear  CF: low risk |
|  | Stone 2020  (19) | | | 62 high-risk neuroblastoma | | Median 5.29 (0.24-15.24) | 18%  (n=12) | **FVC**  (FVC <80% pred)  RT + Surgery yes: 91.7% abnormal  RT + Surgery no: 44% abnormal  **TLC**  (TLC <80% pred)  RT + Surgery yes: 75.0% abnormal  RT + Surgery no: 34.0% abnormal | OR, 95%CI  14.00 (1.68 – 116.85),  p=0.003  5.82 (1.39 – 24.38), p=0.010 | 1. No  2. Yes  3. No  4. Yes (ATS)  5. No  6. No | Prospective cohort  SB: high risk  AB: low risk  DB: low risk  CF: high risk |
| *GRADE assessment:* | |  |  | |  | | | | | | |
| Study design: | |  | +4 | 1 retrospective cohort study, 1 prospective cohort study | | | | | | | |
| Study limitations: | |  | -1 | Some limitations: Selection bias low in 1/2, high in 1/2; Attrition bias low in 2/2; Detection bias low in 1/2, unclear in 1/2; Confounding high in 1/2low in 1/2 | | | | | | | |
| Consistency: | |  | 0 | Both studies show an increased risk | | | | | | | |
| Directness: | |  | -1 | Results are broadly generalizable. PFT quality differs (1/2 reference values and guidelines mentioned). | | | | | | | |
| Precision: | |  | -1 | Results shown with effect size but very large confidence intervals | | | | | | | |
| Publication bias: | |  | 0 | Unlikely | | | | | | | |
| Effect size: | |  | +1 | Large magnitude of effect | | | | | | | |
| Dose-response: | |  | 0 | Not applicable | | | | | | | |
| Plausible confounding: | |  | 0 | No evidence of possible confounding | | | | | | | |
| Quality of evidence: | | | ⊕⊖⊖⊖ VERY LOW | | | | | | | | |
| Conclusion: | | | Increased risk for restrictive abnormalities (TLC, FVC) after thoracic surgery combined with radiotherapy exposing lung tissue vs. bleomycin alone or no exposure to thoracic surgery and radiotherapy exposing lung tissue in CAYA cancer survivors.  (2 studies; 255 participants; 28 participants exposed to radiotherapy exposing lung tissue plus thoracic surgery; 98 participants exposed to bleomycin only or 50 not exposed) | | | | | | | | |
| Comment | | | Two study, very small sample size exposed to thoracic surgery and radiotherapy exposing lung tissue, very large confidence interval. Different comparators used. | | | | | | | | |

| PICO | Study | No. of participants | Follow-up (median/mean, range) yr | Thoracic surgery and radiotherapy | Pulmonary function Outcomes | Effect size | PFT quality | Risk of bias |
| --- | --- | --- | --- | --- | --- | --- | --- | --- |
| 10b What is the risk of hyperinflation after thoracic surgery combined with radiotherapy exposing lung tissue? | | | | | | | | |

**No study**

| PICO | Study | | | | No. of participants | | Follow-up (median/mean, range) yr | Thoracic surgery and radiotherapy | Pulmonary function Outcomes | Effect size | PFT quality | Risk of bias |
| --- | --- | --- | --- | --- | --- | --- | --- | --- | --- | --- | --- | --- |
| 10b What is the risk of diffusion capacity impairment after thoracic surgery combined with radiotherapy exposing lung tissue? | Mulder 2011 (9) | | | | 193 CCS | | Median 17.9 (5.6-36.8) | Radiotherapy plus surgery 8.3%  (n=16)  Bleomycin only 50.8%  (n=98) | Impaired diffusion  (DLCO <75%)  Bleomycin only vs radiotherapy + surgery | Odds Ratio (95%CI)  Radiotherapy plus surgery vs. Bleomycin only  5.98 (1.64-21.81) | 1. No  2. No  3. No  4. No  5. No  6. No | Retrospective cohort  SB: low risk  AB: low risk  DB: unclear  CF: low risk |
|  | Stone 2020  (19) | | | | 62 high-risk neuroblastoma | | Median 5.29 (0.24-15.24) | 18%  (n=12) | **DLCO**  (DLCO <80% pred)  RT + Surgery yes: 80.0% abnormal  RT + Surgery no: 69.6% abnormal | OR, 95%CI  1.75 (0.33 – 9.31), p=0.70 | 1. No  2. Yes  3. No  4. Yes (ATS)  5. No  6. No | Prospective cohort  SB: high risk  AB: low risk  DB: low risk  CF: high risk |
| *GRADE assessment:* | |  |  | | |  | | | | | | |
| Study design: | |  | +4 | 1 retrospective cohort study, 1 prospective cohort study | | | | | | | | |
| Study limitations: | |  | -1 | Some limitations: Selection bias low in 1/2, high in 1/2; Attrition bias low in 2/2; Detection bias low in 1/2, unclear in 1/2; Confounding, low in 1/2, high in 1/2 | | | | | | | | |
| Consistency: | |  | 0 | Both studies show an increased risk, one with a significant effect | | | | | | | | |
| Directness: | |  | -1 | Results are broadly generalizable. PFT quality differs (1/2 reference values and guidelines mentioned). | | | | | | | | |
| Precision: | |  | -1 | Results shown with effect size but large confidence interval | | | | | | | | |
| Publication bias: | |  | 0 | Unlikely | | | | | | | | |
| Effect size: | |  | +1 | Large magnitude of effect | | | | | | | | |
| Dose-response: | |  | 0 | Not applicable | | | | | | | | |
| Plausible confounding: | |  | 0 | No evidence of possible confounding | | | | | | | | |
| Quality of evidence: | | | ⊕⊖⊖⊖ VERY LOW | | | | | | | | | |
| *Conclusion:* | | | Increased risk for diffusion capacity impairment (DLCO) after thoracic surgery combined with radiotherapy exposing lung tissue vs. bleomycin alone or no exposure to thoracic surgery and radiotherapy exposing lung tissue in CAYA cancer survivors  (2 studies; 255 participants; 28 participants exposed to radiotherapy exposing lung tissue and thoracic surgery; 98 participants exposed to bleomycin only, 50 not exposed to thoracic surgery and radiotherapy exposing lung tissue) | | | | | | | | | |
| *Comment* | | | Very small sample size exposed to thoracic surgery and radiotherapy exposing lung tissue, very large confidence intervals. | | | | | | | | | |

##### 10c Chemo plus radiotherapy

| PICO | Study | | | No. of participants | | Follow-up (median/mean, range) yr | Chemotherapy and radiotherapy | Pulmonary function Outcomes | Effect size | PFT quality | Risk of bias |
| --- | --- | --- | --- | --- | --- | --- | --- | --- | --- | --- | --- |
| 10c What is the risk of obstructive abnormalities after pulmotoxic chemotherapy combined with radiotherapy exposing lung tissue? | Nysom 1998 (24) | | | 41 Lymphoma survivors | | Median 10.5 (range 2.3- 23.7) | 51% (n=21) chemo plus thoracic radiation  49% (n=20) chemo only | Number of CCS with abnormal FEV1 (z-score <-1.645 or >1.645)  11/41 total abnormal FEV1 | Estimated difference in z-score for FEV1 (p-value)  Chemo plus radiotherapy vs. chemo only  0.8 (0.004) | 1. No  2. Yes  Quanjer 1983 and 1995, Rosenthal 1993  3. Yes  4. Yes  5. No  6. No | Retrospective cohort  SB: Low risk  AB: Low risk  DB: High risk  CF: High risk |
| GRADE assessment: | |  |  | |  | | | | | | |
| Study design: | |  | +4 | 1 retrospective cohort study | | | | | | | |
| Study limitations: | |  | -2 | Some limitations: Selection bias low in 1/1; Attrition bias low in 1/1; Detection bias high in 1/1; Confounding high in 1/3 | | | | | | | |
| Consistency: | |  | 0 | One study only | | | | | | | |
| Directness: | |  | 0 | Results generalizable. PFT quality is good (reference values and guidelines mentioned). | | | | | | | |
| Precision: | |  | -2 | Important imprecision; only one study and small sample size | | | | | | | |
| Publication bias: | |  | 0 | Unlikely | | | | | | | |
| Effect size: | |  | 0 | No large magnitude of effect | | | | | | | |
| Dose-response: | |  | 0 | Not applicable | | | | | | | |
| Plausible confounding: | |  | 0 | No evidence of possible confounding | | | | | | | |
| Quality of evidence: | | | ⊕⊖⊖⊖ Very low | | | | | | | | |
| Conclusion: | | | Decreased risk for obstructive abnormalities (FEV1) after radiotherapy exposing lung tissue combined with chemotherapy vs. chemotherapy alone in CAYA cancer survivors  (1 study; 41 participants; 21 participants exposed to chemotherapy and radiotherapy exposing lung tissue ; 20 participants exposed to chemotherapy only) | | | | | | | | |
| Comment | | | One study only, small sample size, univariable analysis only | | | | | | | | |

| PICO | Study | | | | No. of participants | | Follow-up (median/mean, range) yr | Chemotherapy and  radiotherapy exposing lung tissue | Pulmonary function Outcomes | Effect size | PFT quality | Risk of bias |
| --- | --- | --- | --- | --- | --- | --- | --- | --- | --- | --- | --- | --- |
| 10c What is the risk of restrictive abnormalities after pulmotoxic chemotherapy combined with radiotherapy exposing lung tissue? | Nysom 1998 (20) | | | | 41 Lymphoma survivors | | Median 10.5 (range 2.3- 23.7) | 51% (n=21) chemo plus thoracic radiation  49% (n=20) chemo only | Number of CCS with abnormal lung function parameter  Total 16/41 restrictive flow volume curve:  chemo+RT vs chemo | p-value  0.4 | 1. No  2. Yes  Quanjer 1983 and 1995, Rosenthal 1993  3. Yes  4. Yes  5. No  6. No | Retrospective cohort  SB: Low risk  AB: Low risk  DB: High risk  CF: High risk |
|  | Mulder 2011 (9) | | | | 193 CCS | | Median 17.9 (5.6-36.8) | Bleomycin only 50.8% (n=98)  Bleomycin plus radiotherapy 4.7% (n=9) | Restrictive impairment (TLC OR FVC <75%) | Odds Ratio (95%CI)  Radiotherapy plus bleomycin vs. Bleomycin only  9.41 (1.71-51.86) | 1. No  2. No  3. No  4. No  5. No  6. No | Retrospective cohort  SB: low risk  AB: low risk  DB: unclear  CF: low risk |
| GRADE assessment: | |  |  | | |  | | | | | | |
| Study design: | |  | +4 | 2 retrospective cohort studies | | | | | | | | |
| Study limitations: | |  | -2 | Some limitations: Selection bias low in 2/2; Attrition bias low in 2/2; Detection bias high in 1/2, unclear in 1/2; Confounding high in 1/2, low in 1/2 | | | | | | | | |
| Consistency: | |  | 0 | Both studies show tendency to restrictive abnormalities | | | | | | | | |
| Directness: | |  | -1 | Results broadly generalizable but unsure PFT quality (1/2 stated reference values, ½ stated lung function procedure used) | | | | | | | | |
| Precision: | |  | -1 | Important imprecision, 1/2 with very wide confidence interval, in 1/2 precision cannot be judged as result shown as p-value only | | | | | | | | |
| Publication bias: | |  | 0 | Unlikely | | | | | | | | |
| Effect size: | |  | +1 | Large magnitude of effect | | | | | | | | |
| Dose-response: | |  | 0 | Not applicable | | | | | | | | |
| Plausible confounding: | |  | 0 | No evidence of possible confounding | | | | | | | | |
| Quality of evidence: | | | ⊕⊖⊖⊖ Very low | | | | | | | | | |
| Conclusion: | | | Increased risk for restrictive abnormalities (TLC or FVC) after radiotherapy exposing lung tissue combined with bleomycin vs. bleomycin alone in CAYA cancer survivors  (2 studies; 1 study significant, 1 study non-significant; 234 participants; 30 participants exposed to chemotherapy and radiotherapy exposing lung tissue ) | | | | | | | | | |
| Comment | | | Important imprecision, PFT quality unsure, small sample size exposed to chemotherapy and radiotherapy exposing lung tissue; one study focusses on bleoymcin-containing chemotherapy the second does not differentiate between type of chemotherapy | | | | | | | | | |

| PICO | Study | No. of participants | Follow-up (median/mean, range) yr | Chemotherapy and radiotherapy | Pulmonary function Outcomes | Effect size | PFT quality | Risk of bias |
| --- | --- | --- | --- | --- | --- | --- | --- | --- |
| 10c What is the risk of hyperinflation after pulmotoxic chemotherapy combined with radiotherapy to the chest? | | | | | | | | |

**No stud y**

| PICO | Study | | | No. of participants | | Follow-up (median/mean, range) yr | Chemotherapy and radiotherapy | Pulmonary function Outcomes | Effect size | PFT quality | Risk of bias |
| --- | --- | --- | --- | --- | --- | --- | --- | --- | --- | --- | --- |
| 10c What is the risk of diffusion capacity impairment after pulmotoxic chemotherapy combined with radiotherapy to the chest? | Nysom 1998 (20) | | | 41 Lymphoma survivors | | Median 10.5 (range 2.3- 23.7) | 51% (n=21) chemo plus thoracic radiation  49% (n=20) chemo only | Number of CCS with abnormal transfer factor  Total 16/41 abnormal transfer factor: | Estimated difference in standardized residuals (p-value)  Chemo+RT vs chemo alone  0.1 (0.7) | 1. No  2. Yes  Quanjer 1983 and 1995, Rosenthal 1993  3. Yes  4. Yes  5. No  6. No | Retrospective cohort  SB: Low risk  AB: Low risk  DB: High risk  CF: High risk |
|  | Mulder 2011 (9) | | | 193 CCS | | Median 17.9 (5.6-36.8) | Bleomycin only 50.8% (n=98)  Bleomycin plus radiotherapy 4.7% (n=9) | Diffusion impairment (DLCO <75%) | Odds Ratio (95%CI)  Bleomycin only vs radiotherapy + bleomycin  6.17 (1.37-27.84) | 1. No  2. No  3. No  4. No  5. No  6. No | Retrospective cohort  SB: low risk  AB: low risk  DB: unclear  CF: low risk |
| GRADE assessment: | |  |  | |  | | | | | | |
| Study design: | |  | +4 | 2 retrospective cohort studies | | | | | | | |
| Study limitations: | |  | -2 | Some limitations: Selection bias low in 2/2; Attrition bias low in 2/2; Detection bias high in 1/2, unclear in 1/2; Confounding high in 1/2, low in ½ | | | | | | | |
| Consistency: | |  | 0 | Both studies show tendency to diffusion capacity impairment | | | | | | | |
| Directness: | |  | -1 | Results broadly generalizable but unsure PFT quality (1/2 stated reference values, ½ stated lung function procedure used) | | | | | | | |
| Precision: | |  | -1 | Important imprecision, 1/2 with very wide confidence interval, in 1/2 precision cannot be judged as result shown as p-value only | | | | | | | |
| Publication bias: | |  | 0 | Unlikely | | | | | | | |
| Effect size: | |  | +1 | Large magnitude of effect | | | | | | | |
| Dose-response: | |  | 0 | Not applicable | | | | | | | |
| Plausible confounding: | |  | 0 | No evidence of possible confounding | | | | | | | |
| Quality of evidence: | | | ⊕⊖⊖⊖ Very low | | | | | | | | |
| Conclusion: | | | Increased risk for diffusion capacity impairment abnormalities after radiotherapy exposing lung tissue combined with bleomycin vs. bleomycin alone in CAYA cancer survivors.  (2 studies; 1 study significant, 1 study non-significant; 234 participants; 30 participants exposed to chemotherapy and radiotherapy) | | | | | | | | |
| Comment | | | Important imprecision, PFT quality unsure, small sample size exposed to chemotherapy and radiotherapy; one study focusses on bleoymcin-containing chemotherapy the second does not differentiate between type of chemotherapy | | | | | | | | |

#### PICO 11: Smoking

| PICO | Study | | | No. of participants | | Follow-up (median/mean, range) yr | Tobacco exposure | Pulmonary function Outcomes | Effect size | PFT quality | Risk of bias |
| --- | --- | --- | --- | --- | --- | --- | --- | --- | --- | --- | --- |
| 11 What is the risk of obstructive abnormalities in CAYA who have a history of tobacco exposure compared to CAYA with no history of tobacco exposure | Stone 2020  (19) | | | 62 high-risk neuroblastoma | | Median 5.29 (0.24-15.24) | 18%  (n=11) | **FEV1**  (FEV1 <80% pred)  Smoke yes: 36.4% abnormal  Smoke no: 49% abnormal | OR, 95%CI  0.59 (0.16 – 2.28), p=0.446 | 1. No  2. Yes  3. No  4. Yes (ATS)  5. No  6. No | Prospective cohort  SB: high risk  AB: low risk  DB: low risk  CF: high risk |
| GRADE assessment: | |  |  | |  | | | | | | |
| Study design: | |  | +4 | 1 prospective cohort study | | | | | | | |
| Study limitations: | |  | -2 | Some limitations: Selection bias high in 1/1; Attrition bias low in 1/1; Detection bias low in 1/1; Confounding high in 1/1 | | | | | | | |
| Consistency: | |  | 0 | One study only | | | | | | | |
| Directness: | |  | 0 | Results broadly generalizable. PFT good (references stated, lung function procedure mentioned) | | | | | | | |
| Precision: | |  | -1 | No important imprecision (effect size with 95%CI), only one study | | | | | | | |
| Publication bias: | |  | 0 | Unlikely | | | | | | | |
| Effect size: | |  | 0 | No large magnitude of effect | | | | | | | |
| Dose-response: | |  | 0 | No dose-response relationship | | | | | | | |
| Plausible confounding: | |  | 0 | No evidence of possible confounding | | | | | | | |
| Quality of evidence: | | | ⊕⊖⊖⊖ Very low | | | | | | | | |
| Conclusion: | | | No significant effect on reduced risk for obstructive abnormalities (FEV1) CAYA cancer survivors with a smoking history compared to those without.  (1 study; 62 participants; 11 former or current smoker) | | | | | | | | |
| Comment | | | One study only, small sample size, only neuroblastoma survivors | | | | | | | | |

| PICO | Study | | | No. of participants | | Follow-up (median/mean, range) yr | Tobacco exposure | Pulmonary function Outcomes | Effect size | PFT quality | Risk of bias |
| --- | --- | --- | --- | --- | --- | --- | --- | --- | --- | --- | --- |
| 11 What is the risk of restrictive abnormalities in CAYA who have a history of tobacco exposure compared to CAYA with no history of tobacco exposure | Stone 2020  (19) | | | 62 high-risk neuroblastoma | | Median 5.29 (0.24-15.24) | 18%  (n=11) | **FVC**  (FVC <80% pred)  Smoke yes: 45.5% abnormal  Smoke no: 54.9% abnormal  **TLC**  (TLC <80% pred)  Smoke yes: 36.4% abnormal  Smoke no: 43.1% abnormal | OR, 95%CI  0.69 (0.19 – 2.53), p=0.569  0.75 (0.20 – 2.90), p=0.748 | 1. No  2. Yes  3. No  4. Yes (ATS)  5. No  6. No | Prospective cohort  SB: high risk  AB: low risk  DB: low risk  CF: high risk |
| GRADE assessment: | |  |  | |  | | | | | | |
| Study design: | |  | +4 | 1 prospective cohort study | | | | | | | |
| Study limitations: | |  | -2 | Some limitations: Selection bias high in 1/1; Attrition bias low in 1/1; Detection bias low in 1/1; Confounding high in 1/1 | | | | | | | |
| Consistency: | |  | 0 | One study only | | | | | | | |
| Directness: | |  | 0 | Results broadly generalizable. PFT good (references stated, lung function procedure mentioned) | | | | | | | |
| Precision: | |  | -1 | No important imprecision (effect size with 95%CI), only one study | | | | | | | |
| Publication bias: | |  | 0 | Unlikely | | | | | | | |
| Effect size: | |  | 0 | No large magnitude of effect | | | | | | | |
| Dose-response: | |  | 0 | No dose-response relationship | | | | | | | |
| Plausible confounding: | |  | 0 | No evidence of possible confounding | | | | | | | |
| Quality of evidence: | | | ⊕⊖⊖⊖ Very low | | | | | | | | |
| Conclusion: | | | No significant effect on reduced risk for obstructive abnormalities (FVC, TLC) CAYA cancer survivors with a smoking history compared to those without.  (1 study; 62 participants; 11 former or current smoker) | | | | | | | | |
| Comment | | | One study only, small sample size, only neuroblastoma survivors | | | | | | | | |

| PICO | Study | No. of participants | Follow-up (median/mean, range) yr | Tobacco exposure | Pulmonary function Outcomes | Effect size | PFT quality | Risk of bias |
| --- | --- | --- | --- | --- | --- | --- | --- | --- |
| 11 What is the risk of hyperinflation in CAYA who have a history of tobacco exposure compared to CAYA with no history of tobacco exposure | | | | | | | | |

**No study**

| PICO | Study | | | No. of participants | | Follow-up (median/mean, range) yr | Tobacco exposure | Pulmonary function Outcomes | Effect size | PFT quality | Risk of bias |
| --- | --- | --- | --- | --- | --- | --- | --- | --- | --- | --- | --- |
| 11 What is the risk of diffusion capacity impairment in CAYA who have a history of tobacco exposure compared to CAYA with no history of tobacco exposure | Stone 2020  (19) | | | 62 high-risk neuroblastoma | | Median 5.29 (0.24-15.24) | 18%  (n=11) | **DLCO**  (FVC <80% pred)  Smoke yes: 54.6% abnormal  Smoke no: 75.6% abnormal | OR, 95%CI  0.39 (0.10 – 1.52), p=0.263 | 1. No  2. Yes  3. No  4. Yes (ATS)  5. No  6. No | Prospective cohort  SB: high risk  AB: low risk  DB: low risk  CF: high risk |
| GRADE assessment: | |  |  | |  | | | | | | |
| Study design: | |  | +4 | 1 prospective cohort study | | | | | | | |
| Study limitations: | |  | -2 | Some limitations: Selection bias high in 1/1; Attrition bias low in 1/1; Detection bias low in 1/1; Confounding high in 1/1 | | | | | | | |
| Consistency: | |  | 0 | One study only | | | | | | | |
| Directness: | |  | 0 | Results broadly generalizable. PFT good (references stated, lung function procedure mentioned) | | | | | | | |
| Precision: | |  | -1 | No important imprecision (effect size with 95%CI), only one study | | | | | | | |
| Publication bias: | |  | 0 | Unlikely | | | | | | | |
| Effect size: | |  | 0 | No large magnitude of effect | | | | | | | |
| Dose-response: | |  | 0 | No dose-response relationship | | | | | | | |
| Plausible confounding: | |  | 0 | No evidence of possible confounding | | | | | | | |
| Quality of evidence: | | | ⊕⊖⊖⊖ Very low | | | | | | | | |
| Conclusion: | | | No significant effect on reduced risk for obstructive abnormalities (DLCO) CAYA cancer survivors with a smoking history compared to those without.  (1 study; 62 participants; 11 former or current smoker) | | | | | | | | |
| Comment | | | One study only, small sample size, only neuroblastoma survivors | | | | | | | | |

##### 11a Smoker vs ex-smoker

| PICO | Study | | | No. of participants | | Follow-up (median/mean, range) yr | Tobacco exposure | Pulmonary function Outcomes | Effect size | PFT quality | Risk of bias |
| --- | --- | --- | --- | --- | --- | --- | --- | --- | --- | --- | --- |
| 11a What is the risk of obstructive abnormalities in smokers/ex-smokers compared to non-smokers? | Oancea 2014 (25) | | | 433 CCS | | >10 yrs from diagnosis | a. Never Smoker:  62% (n=269)  b. Former: 18%  (n=80)  c. Current: 19%  (n=84)  d. Ever smoker  <6PY (n=69)  e. Ever smoker ≥6PY  (n=80) | % predicted median (IQR)  **FEV1/FVC**  a. 1.02 (0.96-1.06)  b. 0.98 (0.93-1.04)  c. 1.00 (0.94-1.04)  d. 1.00 (0.94-1.04)  e. 0.99 (0.92-1.03)  **FEV1**  a. 79.0 (69.0-92.0)  b. 76.5 (65.5-86.0)  c. 79.5 (67.0-89.0)  d. 79.0 (69.0-88.0)  e. 78.0 (66.0-87.0) | Comparison with never smoker as ref., using the DSCF procedure  p=0.01  p=0.03  p=0.38  p=0.005  p=0.23  p=0.83  p=0.66  p=0.38 | 1. No  2. No  3. No  4. Yes: ATS  5. No  6. No | Retrospective cohort  SB: high risk  AB: low risk  DB: low risk  CF: unclear |
| GRADE assessment: | |  |  | |  | | | | | | |
| Study design: | |  | +4 | 1 retrospective cohort study | | | | | | | |
| Study limitations: | |  | -2 | Some limitations: Selection bias high in 1/1; Attrition bias high in 1/1; Detection bias low in 1/1; Confounding unclear in 1/1 | | | | | | | |
| Consistency: | |  | 0 | One study only | | | | | | | |
| Directness: | |  | -1 | Results broadly generalizable but unsure PFT quality (no references stated, lung function procedure mentioned) | | | | | | | |
| Precision: | |  | -2 | Important imprecision, precision cannot be judged because results shown as p-value only, only one study | | | | | | | |
| Publication bias: | |  | 0 | Unlikely | | | | | | | |
| Effect size: | |  | 0 | No large magnitude of effect | | | | | | | |
| Dose-response: | |  | 0 | No dose-response relationship | | | | | | | |
| Plausible confounding: | |  | 0 | No evidence of possible confounding | | | | | | | |
| Quality of evidence: | | | ⊕⊖⊖⊖ Very low | | | | | | | | |
| Conclusion: | | | Increased risk for obstructive abnormalities (FEV1/FVC) in current and former smoker and those who smoked ≥6 PY vs. never smokers in CAYA cancer survivors.  (1 study; 433 participants; 164 former or current smoker) | | | | | | | | |
| Comment | | | Important imprecision, PFT quality unsure | | | | | | | | |

| PICO | Study | | | No. of participants | | Follow-up (median/mean, range) yr | Tobacco exposure | Pulmonary function Outcomes | Effect size | PFT quality | Risk of bias |
| --- | --- | --- | --- | --- | --- | --- | --- | --- | --- | --- | --- |
| 11a What is the risk of restrictive abnormalities in smokers/ex-smokers compared to non-smokers? | Oancea 2014 (21) | | | 433 CCS | | >10 yrs from diagnosis | a. Never Smoker:  62% (n=269)  b. Former: 18%  (n=80)  c. Current: 19%  (n=84)  d. Ever smoker  <6PY (n=69)  e. Ever smoker ≥6PY  (n=80) | % predicted median (IQR)  **TLC**  a. 80.0% (69-91)  b. 82.0% (73-93)  c. 87.0% (74-94)  d. 81.0% (70-90)  e. 86.5% (74-94)  **FVC**  a. 79.0 (67.0-91.0)  b. 77.0 (67.5-88.0)  c. 83.0 (70.0-90.0)  d. 80.0 (68.0-87.0)  e. 81.5 (68.0-88.5) | Comparison with never smoker as ref., using the DSCF procedure  p=0.54  p=0.12  p=0.98  p=0.08  p=0.80  p=0.88  p=0.85  p=0.99 | 1. No  2. No  3. No  4. Yes: ATS  5. No  6. No | Retrospective cohort  SB: high risk  AB: low risk  DB: low risk  CF: unclear |
|  | Nysom 1998 (11) | | | 94 leukemia survivors | | Median 10.6 (range 3.4-23.4) | 19% smoker (n=18)  4% former smoker (n=4) | 15 TLC reduced/raised | Regression coeff. (95%CI, p-value):  0.31  (-0.18 - 0.80, 0.2) | 1. No  2. Yes  Reference form own laboratory by adjusting published  reference values (Quanjer PH, Pediatr Pulmonol. 1995; Rosenthal M, Thorax, 1993; Quanjer PH, Bull Eur Physiopathol Respir, 1983: Stam H, Pediatr Pulmonl, 1996)  3. Yes  4. Yes  5. No  6. No | Prospective cohort  SB: High risk  AB: Low risk  DB: Unclear  CF: High risk |
|  | Armenian, 2015 (14) | | | 121 CCS | | Median 17.1 (6.3-40.1) | 5.0%  (n=6) | Total 29 restrictive  (TLC <75% and FEV1 ≥80% predicted) | Logistic regression  Odds Ratio (95%CI)  0.9 (0.7-1.9) | 1. Yes  2. No  3. No  4. Yes: ATS  5. No  6. Yes | Prospective cohort  SB: Low risk  AB: low risk  DB: low risk  CF: high risk |
| GRADE assessment: | |  |  | |  | | | | | | |
| Study design: | |  | +4 | 1 retrospective cohort studies, 2 prospective cohort studies | | | | | | | |
| Study limitations: | |  | -2 | Some limitations: Selection bias high in 2/3, low in 1/3; Attrition bias low in 3/3; Detection bias low in 2/3, unclear in 1/3; Confounding high in 2/3, unclear 1/3 | | | | | | | |
| Consistency: | |  | 0 | All studies show similar results | | | | | | | |
| Directness: | |  | -1 | Results broadly generalizable but unsure PFT quality (1/3 stated reference values, 3/3 mention lung function procedure) | | | | | | | |
| Precision: | |  | -1 | Important imprecision, 2/3 with small confidence intervals, in 1/3 precision cannot be judged as results are shown as p-values only | | | | | | | |
| Publication bias: | |  | 0 | Unlikely | | | | | | | |
| Effect size: | |  | 0 | No large magnitude of effect | | | | | | | |
| Dose-response: | | NA | 0 | No dose-response relationship | | | | | | | |
| Plausible confounding: | |  | 0 | No evidence of possible confounding | | | | | | | |
| Quality of evidence: | | | ⊕⊖⊖⊖ Very low | | | | | | | | |
| Conclusion: | | | No significant effect on restrictive abnormalities (TLC, FVC, “restrictive”) in CAYA cancer survivors who smoke/smoked compared to non-smoker.  (3 studies; 648 participants; 182 participants current or former smoker) | | | | | | | | |
| Comment | | | Two studies with very small number of CCS who smoke/smoked. PFT quality is unsure. Different definitions for “restrictive”. | | | | | | | | |

| PICO | Study | No. of participants | Follow-up (median/mean, range) yr | Tobacco exposure | Pulmonary function Outcomes | Effect size | PFT quality | Risk of bias |
| --- | --- | --- | --- | --- | --- | --- | --- | --- |
| 11a What is the risk of hyperinflation in smokers/ex-smokers compared to non-smokers? | | | | | | | | |

**No study**

| PICO | Study | | | No. of participants | | Follow-up (median/mean, range) yr | Tobacco exposure | Pulmonary function Outcomes | Effect size | PFT quality | Risk of bias |
| --- | --- | --- | --- | --- | --- | --- | --- | --- | --- | --- | --- |
| 11a What is the risk of diffusion capacity impairment in smokers/ex-smokers compared to non-smokers? | Myrdal 2018 (26) | | | 116 ALL | | Median 23.2 (range 7.4 – 40.0) | 19%  (n=22) | DLCO %predicted in CCS smoking vs. non-smoker  Total 22% (n=25) DLCO below %pred | Multivariable analysis, Correlation coeff. β (95% CI, p-value  -9.8 (-16.0 - -3.6, 0.002) | 1. No  2. Yes:  Wanger J, Eur Respir J, 2005; Pellegrino R, Eur Respir J, 2005  3. No  4. Yes: ERS  5. No  6. No | Prospective cross-sectional  SB: unclear  AB: low risk  DB: unclear  CF: low risk |
|  | Oancea 2014 (21) | | | 433 CCS | | >10 yrs from diagnosis | a. Never Smoker:  62% (n=269)  b. Former: 18%  (n=80)  c. Current: 19%  (n=84)  d. Ever smoker  <6PY (n=69)  e. Ever smoker ≥6PY  (n=80) | % predicted median (IQR)  **DLCOcorr**  a. 77.5% (66.0-89.0)  b. 77.0% (68.6-86.5)  c. 74.0% (60.0-82.0)  d. 77.5 (64.5-85.0)  e. 71.5% (62.0-81.0) | Comparison with never smoker as ref., using the DSCF procedure  p=0.99  p=0.02  p=0.96  p=0.03 | 1. No  2. No  3. No  4. Yes: ATS  5. No  6. No | Retrospective cohort  SB: high risk  AB: low risk  DB: low risk  CF: unclear |
|  | Armenian, 2015 (14) | | | 121 CCS | | Median 17.1 (6.3-40.1) | 5.0%  (n=6) | Total 42 diffusion abnormality | Univariable regression  Odds Ratio (95%CI)  0.9 (0.2-5.3) | 1. Yes  2. No  3. No  4. Yes: ATS  5. No  6. Yes | Prospective cohort  SB: Low risk  AB: low risk  DB: low risk  CF: high risk |
|  | Zorzi 2015 (16) | | | 143 CCS (Hodgkin, extracranial germ cell tumor) | | Median 4.4  (2 – 7.4) | 2% (n=3) | Total 27 abnormal DLCO | p=0.04 | 1. No  2. Yes  Stanojevic S, Am J Respir Crit Care Med, 2008; Wanger J, Eur Respir J, 2005; Weng TR, Am Rev Respir Dis, 1969; Pellegrino R, Eur Respir J, 2005; reference equations from Sick Children  3. No  4. No  5. No  6. No | Retrospective cross-sectional  SB: high risk  AB: low risk  DB: low risk  CF: unclear |
| GRADE assessment: | |  |  | |  | | | | | | |
| Study design: | |  | +4 | 1 retrospective cohort study, 1 prospective cohort study, 1 prospective cross-sectional study, 1 retrospective cross-sectional study | | | | | | | |
| Study limitations: | |  | -2 | Some limitations: Selection bias high in 2/4, low in 1/4, unclear in 1/4; Attrition bias low in 4/4; Detection bias low in 3/4, unclear in 1/4; Confounding high in 1/4, low in 1/4, unclear 2/4 | | | | | | | |
| Consistency: | |  | 0 | Most studies show similar results | | | | | | | |
| Directness: | |  | -1 | Results broadly generalizable but unsure PFT quality (2/4 stated reference values, 3/4 mention lung function procedure) | | | | | | | |
| Precision: | |  | -1 | Important imprecision, 1/4 with small confidence interval, 1/4 with large confidence interval, in 2/4 precision cannot be judged as results are shown as p-value only | | | | | | | |
| Publication bias: | |  | 0 | Unlikely | | | | | | | |
| Effect size: | |  | 0 | No large magnitude of effect | | | | | | | |
| Dose-response: | |  | 0 | No dose-response relationship | | | | | | | |
| Plausible confounding: | |  | 0 | No evidence of possible confounding | | | | | | | |
| Quality of evidence: | | | ⊕⊖⊖⊖ Very low | | | | | | | | |
| Conclusion: | | | Inconsistent findings for diffusion capacity impairment in CAYA cancer survivors for current smoker and those who ever smoked ≥6py vs. ….?? .  (4 studies; 813 participants; 195 exposed to smoking) | | | | | | | | |
| Comment | | | Two studies with very small sample size exposed to smoking, important imprecision, and PFT quality is unsure. | | | | | | | | |

##### 11b Different doses

| PICO | Study | No. of participants | Follow-up (median/mean, range) yr | Tobacco exposure | Pulmonary function Outcomes | Effect size | Risk of bias |
| --- | --- | --- | --- | --- | --- | --- | --- |
| 11b What is the risk associated with different doses (pack-years)? | | | | | | | |

**No study**

##### 11c Environmental tobacco smoke

| PICO | Study | No. of participants | Follow-up (median/mean, range) yr | Tobacco exposure | Pulmonary function Outcomes | Effect size | Risk of bias |
| --- | --- | --- | --- | --- | --- | --- | --- |
| 11c What is the risk in patients exposed to environmental tobacco smoke compared to not exposed? | | | | | | | |

**No study**

##### 11d Marijuana

| PICO | Study | No. of participants | Follow-up (median/mean, range) yr | Tobacco exposure | Pulmonary function Outcomes | Effect size | Risk of bias |
| --- | --- | --- | --- | --- | --- | --- | --- |
| 11d.What is the risk in marijuana smokers compared to non-smokers? | | | | | | | |

**No study**

### L) Evidence to decision Framework

**General remarks for evidence to decision framework:**

- All types of pulmonary dysfunction (restrictive, obstructive, diffusion capacity) combined, specified if needed
- All types of pulmonary function tests (spirometry, body plethysmography, and DLCO measurement) combined, specified if needed
- All suspected lung-toxic cancer treatments (chemotherapeutic agents [bleomycin, busulfan, nitrosureas], thoracic surgery, and radiotherapy) combined, but specified if needed
- Cyclophosphamide, methotrexate, gemcitabine and combination of different treatment modalities not included as not enough evidence / no studies available

|  | **Criteria** | **Judgements** | **Research evidence** | **Additional considerations, incl. expert opinion** |
| --- | --- | --- | --- | --- |
| PROBLEM | Is the problem a priority? | - No - Probably no - Uncertain - Probably yes   ■ Yes | Child, adolescent, and young adult cancer survivors (CAYA-CS) who have been treated with chest radiation, chemotherapy (bleomycin, busulfan, nitrosureas), thoracic surgery or haematopoietic stem cell transplantation are at risk for pulmonary dysfunction.  Evidence from systematic literature search (some reported prevalence only)   - Prevalence of any pulmonary dysfunction in CAYA-CS at risk* is ~45% (9, 14) - CAYA-CS at risk* are more likely to have restrictive defects (24% vs 5%) and diffusion abnormalities (35% vs 10%) than controls and are more likely to be symptomatic (22% vs. 5%) (14). - Prevalence of any pulmonary dysfunction in CAYA-CS at risk* is ~45% (9, 14) - Proportion of pulmonary dysfunction is higher in studies including specific sub-groups of CAYA-CS (e.g. allogeneic HSCT, whole lung irradiation) (21, 27, 28). - Higher risk in CAYA-CS exposed to thoracic radiotherapy and allogeneic HSCT. - Evidence for other risk factors limited by quality of the studies.   *at risk = exposure to bleomycin, busulfan, or nitrosoureas, and/or chest radiation, and/or allogeneic HSCT, and/or thoracic surgery | - Effect of single chemotherapeutics vs. effect of multiagent treatment strategies on pulmonary dysfunction in CAYA-CS is unclear. The combination of different cancer treatments may be more important than single agents *(expert opinion)*. - Impact of cancer diagnosis (severe disease) and overall cancer therapy on lung growth in general is unknown, might be comparable to pulmonary damage after malnutrition or severe infections *(expert opinion)*. |
| BENEFITS AND HARMS | What is the overall certainty of this evidence? | - No included studies   ■ Very low  ■ Low  ■ Moderate   - High | Evidence from systematic literature search   - For most examined risk factors (see PICO questions), we found either no study, no significant effect or an increased risk for pulmonary dysfunction with very low to moderate quality of evidence: - Bleomycin, busulfan, nitrosureas: no study, no significant effect or decreased risk with very low quality (bleomycin) - Thoracic radiotherapy: increased risk with very low quality of evidence - Thoracic surgery: increased risk with very low quality of evidence - Allogeneic HSCT, including total body irradiation: increased risk with very low to moderate quality of evidence   Evidence from additional studies (not from systematic literature search) supporting the lung toxic effect of:   - chemotherapeutics in CAYA-CS: Jenney et al(29); Huang et al(30), Abid et al(31), Lohani et al(32), Sleijfer et al(33)¸ Matijasic et al(34), O’Driscoll et al (35, 36), Lohani et al (32) - radiotherapy in CAYA-CS: Benoist et al (37), Bolling et al(38), Venkatramani et al(39), Motosue et al(27) - Thoracic surgery in CAYA-CS: Interiano et al(40), Gawade et al(41) - allogeneic HSCT in CAYA-CS: Srinivasan et al(42), Diab et al(43)   Pre-clinical studies on lung toxic effects of:   - Bleomycin: Hay et al(44); Della Latta et al(45) | Additional considerations about “Who needs surveillance” (WG1)   - No significant effect with low quality of evidence and no available study does not exclude the relevance of the studied factors for CAYA-CS. *(expert opinion)* - International LTFU care guidelines recommend screening for CAYA-CS exposed to bleomycin, busulfan, nitrosureas, thoracic radiotherapy and surgery, and HSCT *(Guidelines)* - Early detection of pulmonary dysfunction by PFT as intermediate outcome might reduce mortality, increase quality of life (QoL) and life expectancy in CAYA-CS *(expert opinion)*   Additional considerations about selection of testing modalities   - Spirometry, body plethysmography, and DLCO measurement are standard tests to assess pulmonary function, used for many decades, and are part of routine clinical practice *(expert opinion)*. |
|  | Is there important uncertainty about how much people value the main outcomes? | - Important uncertainty or variability - Possibly important uncertainty or variability   ■ Probably no important uncertainty or variability   - No important uncertainty or variability | Evidence from systematic literature search   - Question cannot be answered by the literature search performed   Evidence from other studies   - Severe pulmonary dysfunction is linked with higher morbidity and mortality even in healthy individuals (46, 47) | The expert panel believes that all key stakeholders (clinicians, CAYA-CS, policy makers, health insurance) would consider the main outcomes (pulmonary dysfunction and morbidity and mortality due to pulmonary dysfunction) as relevant and important outcomes.  Additional considerations   - Moderate and severe pulmonary dysfunction might be linked with CAYA-CS ´QoL *(expert opinion)* - Pulmonary dysfunction becomes more important for CAYA-CS when it affects daily life (e.g. reduced exercise capacity, travelling restrictions, frequent hospitalisations) *(expert opinion)* |
|  | Are the desirable anticipated effects large? | - No - Probably no   ■ Uncertain (lung-toxic chemotherapy)  ■ Probably yes  (thoracic surgery)  ■ Yes (allogeneic HSCT and radiotherapy)   - Varies | Evidence from systematic literature search   - CAYA-CS exposed to allogeneic HSCT benefit the most from surveillance (very low to moderate quality of evidence, dependent on the type of dysfunction), *followed by:* - CAYA-CS exposed to radiotherapy to lung tissue (very low quality of evidence), - CAYA-CS exposed to thoracic surgery to chest or lung tissue (very low quality of evidence), and - CAYA-CS exposed to lung toxic chemotherapy (no study, no significant effect or decreased risk).   Additional evidence   - PFT are non-invasive and the burden of pulmonary dysfunction can be high. Compared to the general population, CAYA-CS experience a four-fold risk of hospitalization (48) due to respiratory problems and a 8 to 14-fold higher risk of respiratory mortality (49, 50). - Yield of screening for subclinical pulmonary dysfunction is high (51, 52) | **Overall, the desirable anticipated effects of surveillance vary depending on the risk to develop pulmonary dysfunction (radiotherapy > allogeneic HSCT > thoracic surgery > bleomycin/ busulfan/ nitrosureas)**  Additional considerations   - Surveillance helps to detect secondary (potentially treatable) complications of pulmonary dysfunction earlier (e.g. pulmonary arterial hypertension), which might be treatable and result in better outcomes (improved life expectancy, quality of life, decreased morbidity) (*expert opinion*). - Surveillance gives the treating physicians an overview of CAYA-CS with pulmonary dysfunction, which may benefit from specific treatment options, when they become newly available (*expert opinion*). - Detection of abnormal pulmonary function parameters may help to strengthen counseling of CAYA-CS, such as advice against smoking, no work with exposure to lung toxic aerosols (e.g. baker, hairdresser, construction worker, car painter), and may have an impact on sports or rehabilitation (*expert opinion*). - Normal surveillance may provide satisfaction and reassurance for CAYA-CS (*expert opinion*).   - Surveillance may provide a baseline against which PFT results can be compared if problems develop in the future (because of the significant interindividual variability of PFT) (*expert opinion*).   - Surveillance may prevent overdiagnosis of pulmonary diseases (e.g. asthma) based on knowledge on preexisting slightly decreased pulmonary function *(expert opinion)* |
|  | Are the undesirable anticipated effects small? | - No - Probably no - Uncertain   ■ Probably yes (for all  test methods)   - Yes - Varies | Evidence from systematic literature search   - Question cannot be answered by the literature search performed | **Overall, the undesirable anticipated effects of surveillance are judged as being small for all PFT methods**  Additional considerations   - Surveillance may result in false-positive findings, e.g. if the CAYA-CS has an intercurrent upper respiratory infection or in case of poor cooperation. However, this can usually be avoided if tests are performed correctly. - Surveillance may result in detection of slightly abnormal PFT parameters, which are clinically not relevant and will not have an impact on survival and quality of life throughout the CAYA-CSs’ life. *(expert opinion)* - Body plethysmography may cause stress, anxiety in people with claustrophobia, but these reactions are not common. *(expert opinion)* - Surveillance might affect self-perception of being a CAYA-CS versus healthy however the recommendations do not require frequent visits to a specialist/hospital. *(expert opinion)*   Additional considerations about selection of testing modalities   - DLCO is more sensitive than spirometry and body plethysmography and might result in more frequent detection of abnormal findings with no impact on survival and QoL of CAYA-CSs *(expert opinion)* |
|  | Are the desirable effects large relative to undesirable effects? | - No - Probably no - Uncertain   ■ Probably yes  (thoracic surgery, lung-toxic chemotherapy)  ■ Yes (allogeneic HSCT and radiotherapy) | Evidence from systematic literature search   - Radiotherapy and allogeneic HSCT: In CAYA-CS treated with thoracic radiotherapy or allogeneic HSCT, the benefits of surveillance with PFT, including DLCO outweight the harms - IGHG literature search: increased risk for both exposures - Thoracic surgery: In CAYA-CS treated with thoracic surgery, the benefits of surveillance with PFT (without DLCO) outweigh the harms - IGHG literature search: increased risk with the exception of DLCO (explained by the pathomechanism – reduction in volume and no change in diffusion capacity itself) - Chemotherapy: In CAYA-CS treated with specific chemotherapeutics (busulfan, belomycin, nitrosureas), the benefits of surveillance with PFT outweighs the harms. Screening should include DLCO due to the pathophysiological mechanism of free radical formation, which may also cause damage to the alveolar-capillary membrane. - IGHG literature search: no studies, no significant effect or decreased risk   Additional evidence   - No evidence available on long-term benefit of screening, such as difference in morbidity or mortality in CAYA-CS with and without screening. However, yield of screening is high (51, 52). - Radiotherapy and allogeneic HSCT: Included in current LTFU guidelines - Thoracic surgery: Included in current LTFU guidelines - Chemotherapy: Included in current LTFU guidelines | Overall, the balance of desirable and undesirable anticipated effects of surveillance vary depending on the risk for pulmonary dysfunction. |
| RESOURCE USE | Are the resources required small? | - No - Probably no - Uncertain - Probably yes   ■ Yes  (Varies to some degree on the PFT performed) | Evidence from systematic literature search   - Question cannot be answered by the literature search performed | Overall, the resources required depend on the test performed and are smallest for spirometry.  Additional considerations   - Availability: Spirometry is widely available in all hospitals and medical practices. Body plethysmography and DLCO are available in larger hospitals and doctors’ offices *(expert opinion)* - Costs: Of the three test modalities, spirometry is cheaper than body plethysmography and DLCO; variabilities may exist based on country-specific health care systems *(expert opinion)* - Time: The test duration is ~10minutes for spirometry and DLCO each, and ~15 minutes for body plethysmography in specialist practices of pulmonologists. However, time can be longer for some patients, for instance young children. Effective time for technicians is longer, as it needs time to instruct CAYA-CS and to enter data *(expert opinion)* - Trained personnel are essential to perform PFT in order to obtain test results of best quality *(expert opinion)* - Interpretation of test results should be made by pulmonologists that are aware of the issues of CAYA-CS *(expert opinion)* |
|  | Is the incremental cost small relative to the net benefits? | - No - Probably no - Uncertain - Probably yes - Yes   ■ Varies depending of the presence and severity of pulmonary dysfunction | Evidence from systematic literature search   - No evidence available about cost-effectiveness of pulmonary function screening in CAYA-CS. | The expert panel is of the opinion that the benefits overall outweigh the costs. |
| EQUITY | What would be the impact on health inequities? | - Increased   ■ Probably increased   - Uncertain - Probably reduced - Reduced   ■ Varies, depending on the test method | Evidence from systematic literature search   - No evidence available. | Additional considerations   - Spirometry is available in most countries, including low and middle income countries, but not all CAYA-CS may have access (e.g. missing health insurance, not affordable, minorities) *(expert opinion)* - Body plethysmography and DLCO might not be available in all places *(expert opinion)* - The panel estimates that the equity of access of tests might differ based on the healthcare system. |
| ACCEPTABILITY | Is the option acceptable to key stakeholders? | - No - Probably no - Uncertain   ■ Probably yes  Yes | Evidence from systematic literature search   - No evidence available. | The panel estimates the option of routine spirometry to be acceptable for all key stakeholders (HCP, CAYA-CS, insurance, other cost bearers).  The panel estimates the option of routine body plethysmography and DLCO to be acceptable for all key stakeholders, if the test is available. |
| FEASIBILITY | Is the option feasible to implement? | - No - Probably no - Uncertain - Probably yes   ■ Yes  ■ Varies, depending on the test method | Evidence from systematic literature search   - No evidence available. | The panel estimates that implementing spirometry should be feasible everywhere. As body plethysmography and DLCO are more specialised and expensive test methods, their implementation might be more challenging in some countries. The same applies to trained personnel. |

**Overall conclusions**

| **Balance of consequences – CAYA-CS exposed to radiotherapy** | | | | | |
| --- | --- | --- | --- | --- | --- |
| Spirometry | Undesirable consequences *clearly outweigh* desirable consequences in most settings  ☐ | Undesirable consequences *probably outweigh* desirable consequences  in most settings  ☐ | The balance between desirable and undesirable consequences  *is closely balanced or uncertain*  ☐ | Desirable consequences *probably outweigh* undesirable consequences  in most settings  ☐ | Desirable consequences *clearly outweigh* undesirable consequences in most settings  ■ |
| Body pleth. |  |  |  |  | ■ |
| DLCO |  |  |  |  | ■ |

| **Balance of consequences – CAYA-CS exposed to allogeneic HSCT** | | | | | |
| --- | --- | --- | --- | --- | --- |
| Spirometry | Undesirable consequences *clearly outweigh* desirable consequences in most settings  ☐ | Undesirable consequences *probably outweigh* desirable consequences  in most settings  ☐ | The balance between desirable and undesirable consequences  *is closely balanced or uncertain*  ☐ | Desirable consequences *probably outweigh* undesirable consequences  in most settings  ☐ | Desirable consequences *clearly outweigh* undesirable consequences in most settings  ■ |
| Body pleth. |  |  |  |  | ■ |
| DLCO |  |  |  |  | ■ |

|  | **Balance of consequences – CAYA-CS exposed to thoracic surgery** | | | | |
| --- | --- | --- | --- | --- | --- |
| Spirometry | Undesirable consequences *clearly outweigh* desirable consequences in most settings  ☐ | Undesirable consequences *probably outweigh* desirable consequences  in most settings  ☐ | The balance between desirable and undesirable consequences  *is closely balanced or uncertain*  ☐ | Desirable consequences *probably outweigh* undesirable consequences  in most settings  ■ | Desirable consequences *clearly outweigh* undesirable consequences in most settings  ☐ |
| Body pleth. |  |  |  | ■ |  |
| DLCO |  |  |  | ■ |  |

|  | **Balance of consequences – CAYA-CS exposed to chemotherapy** | | | | |
| --- | --- | --- | --- | --- | --- |
| Spirometry | Undesirable consequences *clearly outweigh* desirable consequences in most settings  ☐ | Undesirable consequences *probably outweigh* desirable consequences  in most settings  ☐ | The balance between desirable and undesirable consequences  *is closely balanced or uncertain*  ☐ | Desirable consequences *probably outweigh* undesirable consequences  in most settings  ■ | Desirable consequences *clearly outweigh* undesirable consequences in most settings  ☐ |
| Body pleth. |  |  |  | ■ |  |
| DLCO |  |  |  | ■ |  |
